# Characterizing Long COVID: Deep Phenotype of a Complex Condition

**DOI:** 10.1101/2021.06.23.21259416

**Authors:** Rachel R Deer, Madeline A Rock, Nicole Vasilevsky, Leigh Carmody, Halie Rando, Alfred J Anzalone, Tiffany J Callahan, Carolyn T Bramante, Christopher G Chute, Casey S Greene, Joel Gagnier, Haitao Chu, Farrukh M Koraishy, Chen Liang, Feifan Liu, Charisse R Madlock-Brown, Diego R Mazzotti, Douglas S McNair, Ann M Parker, Ben D Coleman, Hannah E Davis, Mallory A Perry, Justin T Reese, Joel Saltz, Anthony E Solomonides, Anupam A Sule, Gary S Stein, Sebastian Köhler, Teshamae S Monteith, Vithal Madhira, Wesley D Kimble, Ramakanth Kavuluru, William B Hillegass, Lauren E Chan, James Brian Byrd, Eilis A Boudreau, Hongfang Liu, Julie A McMurry, Emily Pfaff, Nicolas Matentzoglu, Rose Relevo, Richard A Moffitt, Robert A Schuff, Julian Solway, Heidi Spratt, Timothy Bergquist, Tellen D Bennett, Marc D Basson, Umit Topaloglu, Liwei Wang, Melissa A Haendel, Peter N Robinson

**Author notes:** correspondence: Rachel Deer; Melissa Haendel; Peter N Robinson.

## Abstract

**Importance:** Since late 2019, the novel coronavirus SARS-CoV-2 has given rise to a global pandemic and introduced many health challenges with economic, social, and political consequences. In addition to a complex acute presentation that can affect multiple organ systems, there is mounting evidence of various persistent long-term sequelae. The worldwide scientific community is characterizing a diverse range of seemingly common long-term outcomes associated with SARS-CoV-2 infection, but the underlying assumptions in these studies vary widely making comparisons difficult. Numerous publications describe the clinical manifestations of post-acute sequelae of SARS-CoV-2 infection (PASC or “long COVID”), but they are difficult to integrate because of heterogeneous methods and the lack of a standard for denoting the many phenotypic manifestations of long COVID.

**Observations:** We identified 303 articles published before April 29, 2021, curated 59 relevant manuscripts that described clinical manifestations in 81 cohorts of individuals three weeks or more following acute COVID-19, and mapped 287 unique clinical findings to Human Phenotype Ontology (HPO) terms.

**Conclusions and Relevance:** Patients and clinicians often use different terms to describe the same symptom or condition. Addressing the heterogeneous and inconsistent language used to describe the clinical manifestations of long COVID combined with the lack of standardized terminologies for long COVID will provide a necessary foundation for comparison and meta-analysis of different studies. Translating long COVID manifestations into computable HPO terms will improve the analysis, data capture, and classification of long COVID patients. If researchers, clinicians, and patients share a common language, then studies can be compared or pooled more effectively. Furthermore, mapping lay terminology to HPO for long COVID manifestations will help patients assist clinicians and researchers in creating phenotypic characterizations that are computationally accessible, which may improve the stratification and thereby diagnosis and treatment of long COVID.

## Introduction

Coronavirus SARS-CoV-2 emerged in late 2019 as the third human coronavirus identified in the 21st century. Coronavirus disease 2019 (COVID-19) affects diverse organ systems, including the lungs, digestive tract, kidneys, heart, and brain.^1,2^ As of mid-2021, the full spectrum of the clinical consequences of COVID-19 is not completely understood. Individual symptoms and disease severity vary widely among patients during the acute infection, with some patients developing only mild symptoms or even remaining asymptomatic. In contrast, others experience acute respiratory distress syndrome (ARDS), sepsis, and other life-threatening conditions.^3,4^ As more information about patient recovery has been collected, it has become clear that a wide range of outcomes can also emerge following the acute phase of the illness, with some patients experiencing residual symptoms or developing new symptoms long after the initial infection. This post-acute infection, often referred to as long COVID, post-acute sequelae of COVID (PASC), or post-acute COVID-19 syndrome (PACS), represents a significant challenge for patients, physicians, and society because the causes, patient profile, and even symptom patterns remain difficult to characterize.^5^ These substantial challenges in describing long COVID have led patients to self-organize and perform research to try to expedite the characterization of this disease and therefore how to best ameliorate the substantial impact that long COVID has had on their lives.^5,6^

Long COVID, a multisystem disease, can occur following either severe, mild, or even asymptomatic SARS-CoV-2 infection.^7,8^ There is currently no accepted definition of long COVID; however, it can be broadly defined as delayed recovery from infection with SARS-CoV-2. Long COVID can occur following cases of COVID-19 that were managed in either inpatient or outpatient settings. It is characterized by lasting effects of the infection, unexplained persistence of symptoms, or onset of new chronic diseases, for far longer than would be expected.^9^

Given long COVID’s very recent emergence, no standard framework has yet been established for identifying and assessing associated symptoms or other clinical indicators. Furthermore, symptoms frequently reported by long COVID patients are not assessed consistently across studies. A systematic review available as a preprint^33^ evaluated all research on long COVID released prior to January 1, 2021, that included at least 100 patients; based on the 15 studies that met the inclusion criteria, the authors identified 55 symptoms of long COVID. None of the most common symptoms were assessed by all 15 studies. The authors concluded that the symptoms of long COVID are extremely heterogeneous, and that the assessment of these symptoms varies widely among studies. A recent systematic review concluded that 73% of individuals who had acute COVID-19 experience at least one persistent symptom. However, the authors of the study concluded that the wide variation in design and quality of the studies limited the direct comparability and combinability of the data.^10^ The wide range of symptoms attributed to long COVID are highlighted by an extensive patient-led survey (Patient-Led Research Collaborative). This study conducted deep longitudinal characterization of the long COVID symptoms and trajectories in suspected and confirmed COVID-19 patients who reported illness lasting more than 28 days.^6^ Evaluating data from 3,762 respondents to 257 survey questions, this analysis documented 205 phenotypic features associated with long COVID. The fact that this patient-led study characterized 205 phenotypic features, while the studies cited in the aforementioned systematic review reported only 84 signs or symptoms and 19 laboratory or imaging measurements^10^ suggests that the research community has not yet characterized the full spectrum of clinical manifestations of long COVID. This significant disparity in patient versus clinical characterization motivates the proposed ontological approach to specifying manifestations, which will improve capture and integration of future long COVID studies.

Deep phenotyping is the precise and comprehensive analysis of individual phenotypic abnormalities, with a focus on computational accessibility. In the field of rare disease, the Human Phenotype Ontology (HPO) has become an international standard for deep phenotyping that enables integrated computational analysis of genotype and phenotype for diagnostics, novel disease gene discovery, and translational research.^11,12^ Existing publications on the clinical aspects of long COVID have not used a standard vocabulary to report phenotypic abnormalities, impeding the search, analysis, and integration of information relevant to long COVID in databases such as PubMed. Ontologies like HPO are systematic representations of knowledge that define terminology in a human-readable format and define relationships between concepts in a way that allows computational logical reasoning supporting the integration and analysis of large amounts of data.^13^ The HPO currently has over 16,000 terms that define specific phenotypic abnormalities and includes 37,072 synonyms to enhance search and retrieval.^11,12^

We present a detailed analysis of 287 phenotypic abnormalities, including symptoms, signs, laboratory and imaging abnormalities, that have been reported in previous cohort studies on long COVID. Additionally, we present layperson synonyms and definitions that can be used to link patient self-report questionnaires to standard medical terminology.

## Methods

We searched for publications on long COVID using CoronaCentral, which uses machine learning to process the literature on SARS-CoV-2.^14^ We retrieved 303 articles predicted to be relevant to long COVID on April 29th, 2021. From these, we selected 59 articles that described the clinical manifestations in clinical cohorts of individuals three weeks or more following acute COVID-19. We restricted our analysis to articles describing manifestations three weeks or more after the initial symptoms for outpatients and three weeks or more following discharge for hospitalized patients. Descriptions of long COVID manifestations were mapped to Human Phenotype Ontology (https://hpo.jax.org/app/) terms. For this study, the HPO release 2021-06-08 was used. Four curators, one with experience in long COVID and three with extensive experience in HPO curation, manually reviewed the articles and identified HPO terms that corresponded to the description of clinical abnormalities (symptoms, signs, laboratory abnormalities, abnormal imaging findings) in the articles (**Figure 1**) and mapped them in a spreadsheet. Each mapping was reviewed by four curators and discrepancies were resolved with discussion until consensus was reached. Some publications described multiple time points (e.g., early and late), or varying severities of acute illness (e.g., critical/severe, moderate, mild), which were treated as separate cohorts for the purposes of the current descriptive analysis (**Supplemental Table S1**). We tabulated the relationships between publications and the symptoms they reported, the mapped HPO terms, and body systems (**Supplemental Table S2**).

**Figure 1.**
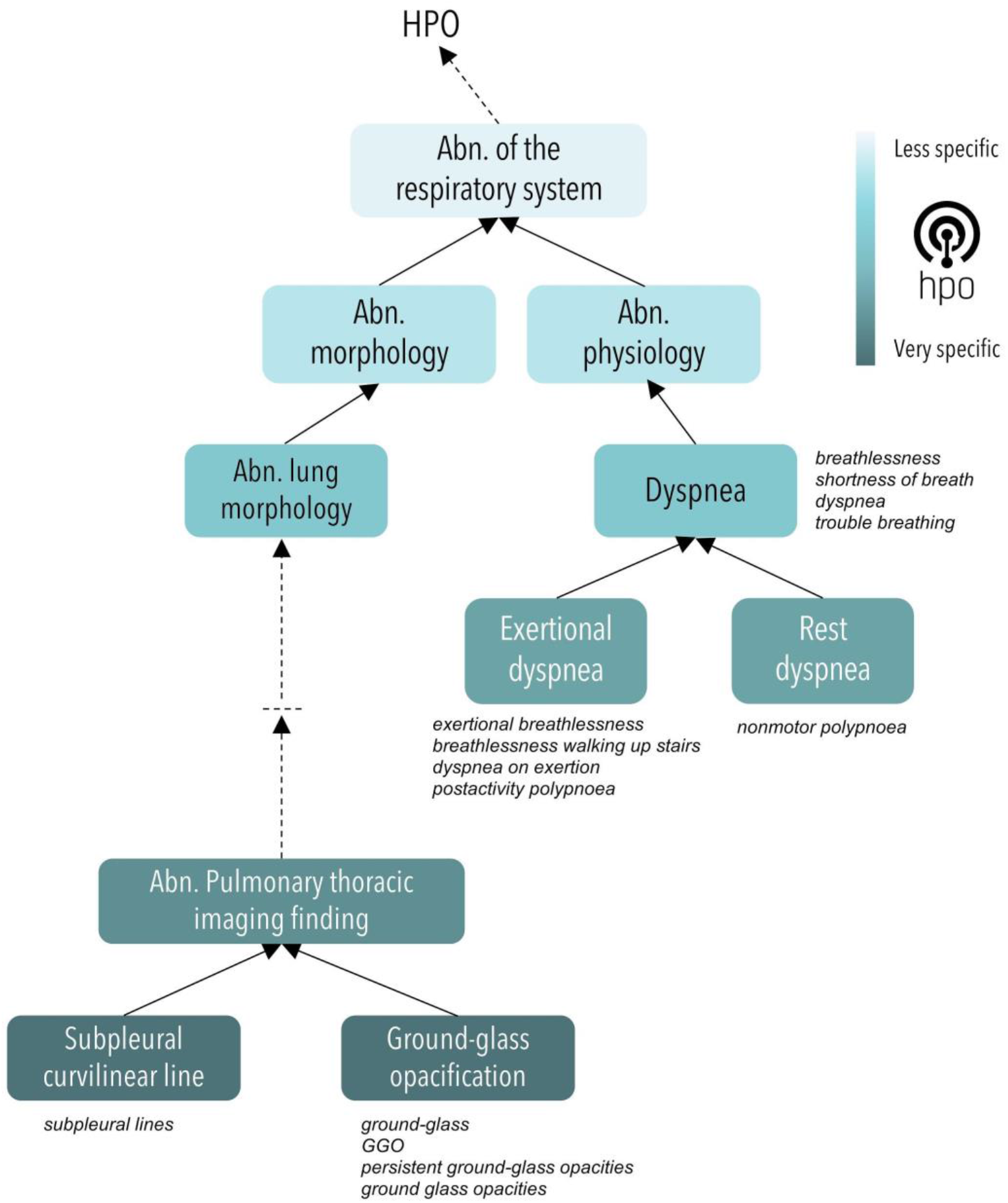
The HPO is arranged in a hierarchy from general to more specific. This graph shows a representative hierarchy of a portion of the HPO ‘abnormality of the respiratory system’ branch. In this study, observations from 59 publications were mapped to the corresponding HPO terms (nodes). A selection of the original terminology used in the manuscripts (in italics) is shown adjacent to the HPO term to which it was mapped. A detailed list of all mapped terms is provided in **Supplemental File 2**.

## Discussion/Observations

We reviewed 303 articles that were predicted to be relevant to long COVID. We excluded articles that were reviews, related only to acute-COVID timepoints, or did not provide sufficient details to extract percentages for the symptoms (i.e. only provided averages but not the number of patients affected in a cohort). Analysis of the remaining 59 articles revealed a variety of criteria were used to identify and evaluate patients with post-acute COVID-19 sequelae. The studies included 11 cohorts of patients who had been treated in the intensive care unit (ICU) during acute COVID-19, 36 cohorts of patients who were hospitalized but not admitted to an ICU during the acute phase, 16 cohorts of patients who were not hospitalized during the acute phase, and 19 mixed cohorts. Some articles pulled data from electronic health records (EHRs) while others strictly relied on patient-reported symptoms from surveys. Studies also varied in the method of collection and instruments used. Methods of collecting data came from phone or electronic surveys, in person review, or pull from electronic medical records. For 26 cohorts, information was collected by in-person, telephone, email, or other online questionnaire. For 51 cohorts, information was collected by clinical examination, and for 5 cohorts, information was collected by questionnaire and clinical examination (**Supplemental Table 1**). The time frame for data collection and followup also differed across studies. Some used a relatively precise window for patient assessment (e.g. 21 days after symptom onset) while others included participants at various distances from acute SARS-COV-2 infection. Some studies aimed to collect information only on patients suffering from long COVID while others collected follow-up information on all patients that had previously had COVID-19 regardless of if they currently or ever had long COVID. Studies differed in how they referred to the phenomenon studied. Some referred to it as long COVID or using a similar term such as post-acute COVID-19 syndrome, whereas others discussed the clinical course or patient recovery without mentioning long COVID specifically. Finally, studies varied widely in the terminology used to describe patient-reported symptoms.

In the 59 publications and 81 cohorts curated for this study **(Supplemental Figure S1)**, a total of 287 phenotypic abnormalities were identified and represented as HPO terms. Of these, 132 terms were used in only one cohort (24.2%), 51 in two cohorts (9.4%), and 62 terms in at least 5 cohorts (21.6%). In most cases, multiple synonyms were mapped to the same HPO term; for instance, *Hepatic steatosis* (HP:0001397) corresponded to four descriptions used in the literature (Steatosis, Liver steatosis, Fatty infiltration of liver, Fatty liver). Among terms reported in at least 10 cohorts (n=31 terms), the most commonly reported feature was *Fatigue* (median 45.1%), and the least commonly reported was *Nausea* (median 3.9%), but the reported percentages varied widely between studies (**Figure 2**).

**Figure 2.**
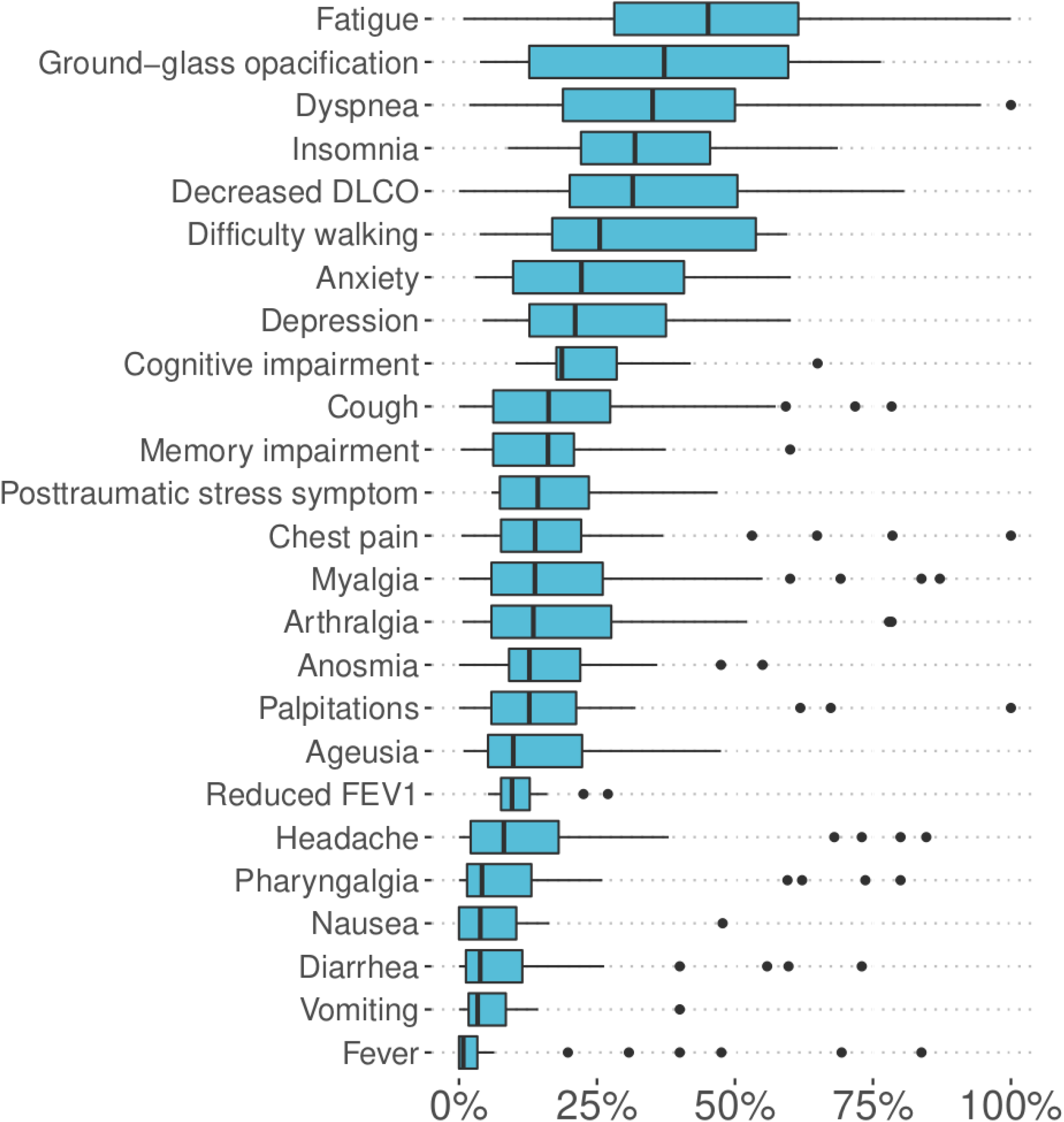
Reported frequencies for the 25 phenotypic features identified in 12 or more cohorts. Box plots are shown for each item, displaying the minimum (1.5 times the interquartile range below the lower quartile), first quartile, median, third quartile, and maximum (1.5 times the interquartile range above the upper quartile). Outliers are shown as dots. DLCO: diffusing capacity of the lungs for carbon monoxide, FEV1: forced expiratory volume in one second; TLC: total lung capacity.

### Clinical Presentation

A wide range of outcomes following acute COVID-19 have emerged as more information about patient recovery has been collected and pathophysiologic mechanisms are revealed. Some patients experience residual symptoms and others develop new symptoms long after the initial infection. These symptoms appear to arise from pathophysiologic changes that span many organ systems and tissues, potentially explained by SARS-CoV-2’s interaction with the endothelium.^15^ Given the timeline of SARS-CoV-2’s emergence, studies to date have tracked patients’ clinical course up to six months post-infection,^16–18^ but anecdotal reports are available describing patients with ongoing symptoms as long as one year post-infection.^19^ Symptoms experienced after the acute illness represent a significant challenge for patients, physicians, and society as a whole. The causes, patient profile, and even symptom patterns associated with long COVID remain difficult to isolate, and the natural history of this condition remains uncharacterized.

Few studies of long COVID to date have conducted analyses elucidating the presence or extent of organ damage. However, preliminary investigations of a number of organ systems have identified organ damage in long COVID patients. These findings are important because they highlight the possibility of asymptomatic long COVID patients who could sustain organ damage due to the SARS-CoV-2 virus that does not immediately present with symptoms. Therefore, an improved understanding of organ damage as an outcome of acute COVID-19 or as a long-term sequelae of the SARS-CoV-2 virus may present new options for patients experiencing persistent symptoms or elucidate new information about how the SARS-CoV-2 virus interacts with a range of organ systems. Here **(Table 1)** we provide details on the vast heterogeneity of symptoms, organized by the organ systems that are likely involved. Full curation details are available in **Supplemental File 2. Supplemental Figures S2-S26** provide an overview of the 287 phenotypic features arranged by category, and **Supplemental Note 1** provides additional commentaries.

**Table 1.** Overview of abnormal phenotypic findings by category. The most commonly reported findings are shown for each category. The total number of features reported in each organ system is shown in the second column. Categories with at least 7 features are shown.

| Category | Total reported features | Most commonly reported feature | Median Percent (number of cohorts) |
| --- | --- | --- | --- |
| abnormalities of smell and taste | 7 | *Anosmia* (HP:0000458) | 12.8% (n=44) |
| behavioral abnormalities | 17 | *Anxiety* (HP:0000739) | 22.2% (n=24) |
| cardiovascular findings | 16 | *Hypertension* (HP:0000822) | 20.4% (n=4) |
| cognitive dysfunction | 8 | *Cognitive impairment* (HP:0100543) | 18.6% (n=13) |
| dermatological findings | 10 | *Alopecia* (HP:0001596) | 18.8% (n=9) |
| emotion/mood abnormalities | 9 | *Depression* (HP:0000716) | 21.1% (n=25) |
| gastrointestinal findings | 9 | *Hepatic steatosis* (HP:0001397) | 26.5% (n=2) |
| gastrointestinal symptoms | 10 | *Diarrhea* (HP:0002014) | 3.8% (n=29) |
| general symptoms | 23 | *Fatigue* (HP:0012378) | 45.1% (n=48) |
| laboratory abnormalities | 23 | *Elevated circulating D-dimer concentration* (HP:0033106) | 26.0% (n=6) |
| neuropsychiatric findings | 30 | *Dysphagia* (HP:0002015) | 1.0% (n=7) |
| ocular abnormalities | 13 | *Blurred vision* (HP:0000622) | 9.7% (n=7) |
| pain | 9 | *Myalgia* (HP:0003326) | 13.8% (n=36) |
| pulmonary findings | 16 | *Decreased DLCO* (HP:0045051) | 31.5% (n=18) |
| pulmonary imaging findings | 15 | *Parenchymal consolidation* (HP:0032177) | 16.8% (n=8) |
| reproductive, genitourinary, endocrine, or metabolism findings | 18 | *Fever* (HP:0001945) | 0.8% (n=29) |
| respiratory symptoms | 14 | *Dyspnea* (HP:0002094) | 35.1% (n=56) |
| sleep impairment | 7 | *Insomnia* (HP:0100785) | 31.9% (n=12) |

In our study, we curated 287 HPO terms representing clinical abnormalities observed in individuals following COVID-19. More research will be needed to determine which of the terms, and potentially which additional terms, are specifically and causally related to SARS-CoV-2 infection. For instance, 10 phenotypic abnormalities in our study have also been reported to occur in Post ICU syndrome (PICS). While most of the manifestations were reported to occur at similar frequencies, Dyspnea was more commonly reported in patients following COVID-19 (**Supplemental Figure S26**). Additional comments are provided in Supplemental Note 2. It is conceivable that in some cases, the occurrence of these ten manifestations in COVID-19 patients is related to care in the ICU. However, all ten symptoms have also been reported in cohorts of individuals treated for COVID-19 as outpatients (**Supplemental File 2**).

Additionally, in many cases it is difficult to know if a clinical abnormality was present prior to acute COVID-19 and was merely diagnosed by investigations and additional clinical tests that were performed following the diagnosis of COVID-19. For instance, Hepatic steatosis was reported to be more common in individuals following acute COVID-19 infection in a single study.20 However, this is a common finding in the general population and additional research will be required to characterize its precise relation to long COVID.

### Linking layperson and health-professional research

One of the challenges in characterizing long COVID is the fact that patients report symptoms that may not be captured by clinical evaluation or in surveys. To truly characterize long COVID and therefore stratify patients into subtypes for care decisions, it is necessary to engage patients directly in the description of their long COVID features. Toward these ends, we have developed layperson synonyms and definitions for each of the 287 HPO terms, expanding on earlier efforts to improve patient accessibility to the HPO by adding layperson synonyms.^21^ A full list is available in the supplemental material in human and computer readable form. This common set of definitions can promote integration of research by translating between patient and clinician descriptions of symptoms. We anticipate that these terms, synonyms and definitions will be a critical resource for use in survey instruments and patient apps for standardizing patient-reporting in the future study of long COVID.

The HPO includes a standardized vocabulary of phenotypic abnormalities associated with over 7,000 diseases. The HPO enables non-exact matching of sets of phenotypic features (phenotype profile) against known diseases, other patients, and model organisms. The algorithms have been implemented for computational comparison of abnormalities and for use in genetic disease diagnostics, and they are the *de facto* standard for deep phenotyping in the field of rare disease.

However, medical terminology is often perplexing to patients, making it difficult for patient-researchers to use resources like the HPO. Patients themselves are an eager and untapped source of accurate information about symptoms and phenotypes, some of which may go unnoticed by the clinician.^6^ Here, we systematically abstracted 287 long COVID manifestations including signs, symptoms, and laboratory as well as imaging abnormalities, added layperson synonyms where missing, and mapped layperson to HPO terminology. We wrote plain-language definitions for these terms to supplement the existing definitions that are aimed at healthcare professionals and researchers.

Patients and clinicians use different terms to describe the same symptoms or conditions. In many cases, the clinical term is an exact match to the layperson synonym; however other times the layperson terminology is less precise. HPO allows layperson synonyms to be mapped to an ontology, with more specific terms being defined as subtypes of more general terms. Mapping lay terminology to HPO for long COVID symptoms will help patients assist clinicians and researchers in creating robust computational phenotype profiles which may improve diagnosis and treatment of long COVID.

In addition, the HPO layperson synonyms mapping from this study will facilitate constructing common data elements related to long COVID research. A strategic plan of the National Library of Medicine (NLM) 2017-2017 and a recent NIH strategic plan for data science identify common data elements as a means by which to improve data interoperability across studies.^22^ Developing a standardized vocabulary with HPO mapped to layperson terms to characterize long COVID symptoms and findings will provide valuable guidance in building common data elements for textual and imaging annotation schema as well as creating patient-centered measurement instruments and clinical surveys.

As long COVID related data increasingly accumulates in EHRs, it is promising to generate practice-based evidence through the secondary use of EHR. However, it is challenging to conduct such research without using natural language processing (NLP) since much information is only stored in unstructured clinical narratives; additionally, because many providers are unfamiliar with the vast array of long COVID symptoms, many are not recorded in these narratives. The layperson definitions and associated synonym mapping would greatly accelerate the development and evaluation of NLP algorithms for extracting long COVID signs and symptoms from EHRs. Baseline NLP algorithms based on HPO can be implemented. Specifically, a fast trie based string matching approach can spot long COVID terms on the fly in massive clinical corpora for near real-time interactive analyses of long COVID phenomena from EHR data. A many-to-one mapping from synonyms to HPO terms identified in this effort will then facilitate more rapid long COVID analytics. Additionally, this effort would enable the rapid development of an annotation guideline for generating benchmarking data, a critical component in developing and evaluating NLP algorithms. If this annotation includes notes from several sites, even spelling mistakes (that nevertheless refer to long COVID terms) can be spotted by first building a named entity recognition (NER) tool and then mapping mentions to long COVID HPO terms through approximate matching via neural word embeddings constructed from character-based neural language models. Another important affordance of this effort is to be able to mine social media posts for long COVID disclosures from patients and healthcare consumers to complement EHR-derived surveillance.^23^

As the National COVID Cohort Collaborative (N3C) established a collaboration among multiple organizations through pandemic data sharing, i.e., Common Data Model (CDM), the long COVID concept standardization enabled by this study will definitely play an indispensable role in achieving the semantic interoperability for the secondary use of EHR among multiple sites.

## Discussion

The fact that some COVID-19 patients experience symptoms following recovery from acute infection is not unexpected. Other infectious diseases, including Epstein-Barr Virus,^24^ Giardia lamblia, Coxiella burnetii, Borrelia burgdorferi (Lyme disease) and Ross River virus,^25^ are also associated with an increased risk for post-infectious sequelae. These sequelae include symptoms such as disabling fatigue, musculoskeletal pain, neurocognitive difficulties, and mood disturbance.^24–26^ Chronic fatigue syndrome (CFS) is frequently preceded by a viral infection.^27^ However, although these sequelae are well documented, they are still not well understood, and the molecular mechanisms underlying these post-acute presentations have yet to be elucidated.

Post-infectious sequelae have also been documented following infection by other coronaviruses. A subset of patients with severe acute respiratory syndrome (SARS), caused by the coronavirus SARS-CoV, and Middle-Eastern Respiratory Syndrome (MERS), caused by the coronavirus MERS-CoV, were observed to experience persistent or new-onset symptoms, including fatigue,^28^ following recovery from the acute infection.^28–30^ For SARS, follow-ups have been conducted up to 15 years post-infection. In addition to fatigue, studies reported effects on lung health and capacity,^31–34^ psychological health,^28^ bone health,^34^ and lipid metabolism,^35^ with the latter two attributed to treatments involving large doses of steroids.^34,35^ Most of the improvements among SARS patients occurred within the first one to two years following infection,^34,36,37^ although some patients continued to experience decreased quality of life for more than a decade following the acute illness.^35^ Though follow-up studies in MERS patients are more sparse, effects on pulmonary function were observed at one year post-infection, with patients who experienced more severe cases at greater risk for long-term effects.^38^

Long-term consequences of COVID-19 comprise an unprecedented range of clinical abnormalities that we are just beginning to understand. Goals of research on long COVID include understanding the natural history of the disease including the prognosis of the many individual manifestations of disease, whether there are well delineated subtypes, whether specific characteristics of the acute phase of COVID-19 predispose to long COVID, and what treatments may best accelerate recovery. Here, we have reported 287 HPO terms representing clinical anomalies reported as PASC in persons following acute COVID-19. For some of the terms, such as those reported only once to date, further research will be required to determine if the abnormalities are specifically related to COVID-19 and their frequency. We have presented plain-language ‘translations’ of all terms that can be used to create patient questionnaires.

### Improving future research on the natural history of long COVID

All published studies analyzed in this work present their results in aggregate rather than providing row level data for each participant. This prevents most data reuse to analyze correlations between comorbidities and risk for long COVID or for specific manifestations of long COVID, and this makes it impossible to investigate potential correlations between long COVID manifestations. Therefore, future studies should present non-identifiable information about individual patients. This will allow correlations between variables. Also, studies need to use controlled vocabulary to classify patients and need to agree on a minimal set of information. Presenting (non-identifiable) data in the form of a table with one row per patient would be a great improvement over the current status. More sophisticated strategies for recording individual clinical histories are available, such as the Global Alliance for Genomics and Health phenopacket schema.^39^

The majority of studies included in this analysis did not apply inclusion criteria to correspond to any specific definition of long COVID, but instead studied groups of patients who had previously undergone infection by SARS-CoV2 with a range of manifestations in the acute phase. Long COVID can be broadly defined as delayed recovery from an episode of COVID-19 and is characterized by lasting effects of the infection, e.g., persistence of symptoms or onset of new chronic diseases, for longer than would be expected.^9^ Although no firm criteria have been established to define the post-acute period or sub-categories within long COVID, several sets of guidelines have been proposed for the classification of COVID-19-related disease phenotypes, and these criteria were compared to the definitions used in the literature. For example, a recently proposed public health framework classifies SARS-CoV-2-related disease into three categories.^40^ The first is acute COVID-19, or the disease most commonly associated with acute SARS-CoV-2 infection. The second category includes Multisystem Inflammatory Syndrome in Children (MIS-C) and in adults (MIS-A), a less common presentation of SARS-CoV-2 infection characterized by hyperinflammation that can appear 4-6 weeks after viral infection.^41^ The third category describes late sequelae.^40^ In terms of defining study cohorts, adherence with this definition would require a clinical diagnosis, rather than a SARS-CoV-2 test alone, to distinguish MIS-C/A and COVID-19. While it appears too early to propose a set of computable definitions for the various types of disease associated with SARS-CoV2 (because we are still learning about the natural history), it would be advantageous for studies to apply a currently accepted definition of long COVID and to describe details in the methods. Studies should denote comorbidities using a standard ontology of diseases such as Mondo.^42^

The studies analyzed in this work varied widely in the terminology used to describe patient-reported symptoms as well as clinical signs, laboratory abnormalities, and imaging findings. For example, the studies analyzed included a mixture of reports of ageusia,^43–45^ anosmia,^43–46^ anosmia/ageusia,^47^ loss of smell,^48,49^ loss of taste,^49^ loss of smell and taste,^50^ loss of smell or taste,^51^ and loss of smell and/or taste.^52^ While in many cases there are parallels among studies (e.g., studies reporting anosmia and loss of smell are likely to be asking the same or similar questions of patients), the lack of a strict definition prevents straight-forward symptom matching across multiple published analyses. Different studies measure clinical manifestations in different ways. For instance, the presence of fatigue can be measured by a yes/no question in an online questionnaire or can be inferred from the results of a multidimensional study instrument such as the Short Form-36 Vitality scale.^53^ This is where standard use of a full terminology such as HPO would be useful to create expressive and consistent meaning across studies. Future studies should make data available using HPO terms provided here or others of the total of over 16,000 HPO terms.

## Conclusions

Working toward computable long COVID phenotypes in this way will improve our ability to understand the natural history of long COVID. Such phenotypes will also allow observational analyses of factors that may reduce long COVID symptoms. The standardized phenotypic features and synonyms bundled in the HPO terms presented here are a foundation for natural language processing of EHR data, clinical decision support tools, and analytic approaches such as machine learning.

## Data Availability

All data is available in supplemental files attached to this submission.

## Supplemental Files

**1. Supplemental File 1**

Contains Figure S1 (Number of HPO terms per cohort); Figures S2-S25 (Reported frequencies of 287 HPO terms arranged according to categories). Table S1. Summary of papers reviewed for inclusion in this work. Table S2 (postacute COVID-19 studies curated in this work, including cohort characteristics and PubMed identifiers). Table S3 (HPO terms used to annotated PICU cohorts).

**2. Supplemental File 2**

Excel file with detailed curations (HPO label, id, original description, PubMed identifier, first author, year, as well as the counts and percentages in the original study.

**3. Supplemental File 3**

Word file with tables that contain HPO ids, labels, definitions, synonyms, and plain-language labels and definitions for the 287 HPO terms used in this work.

