## Supplemental File 1 for "Characterizing Long COVID: Deep Phenotype of a Complex Condition"

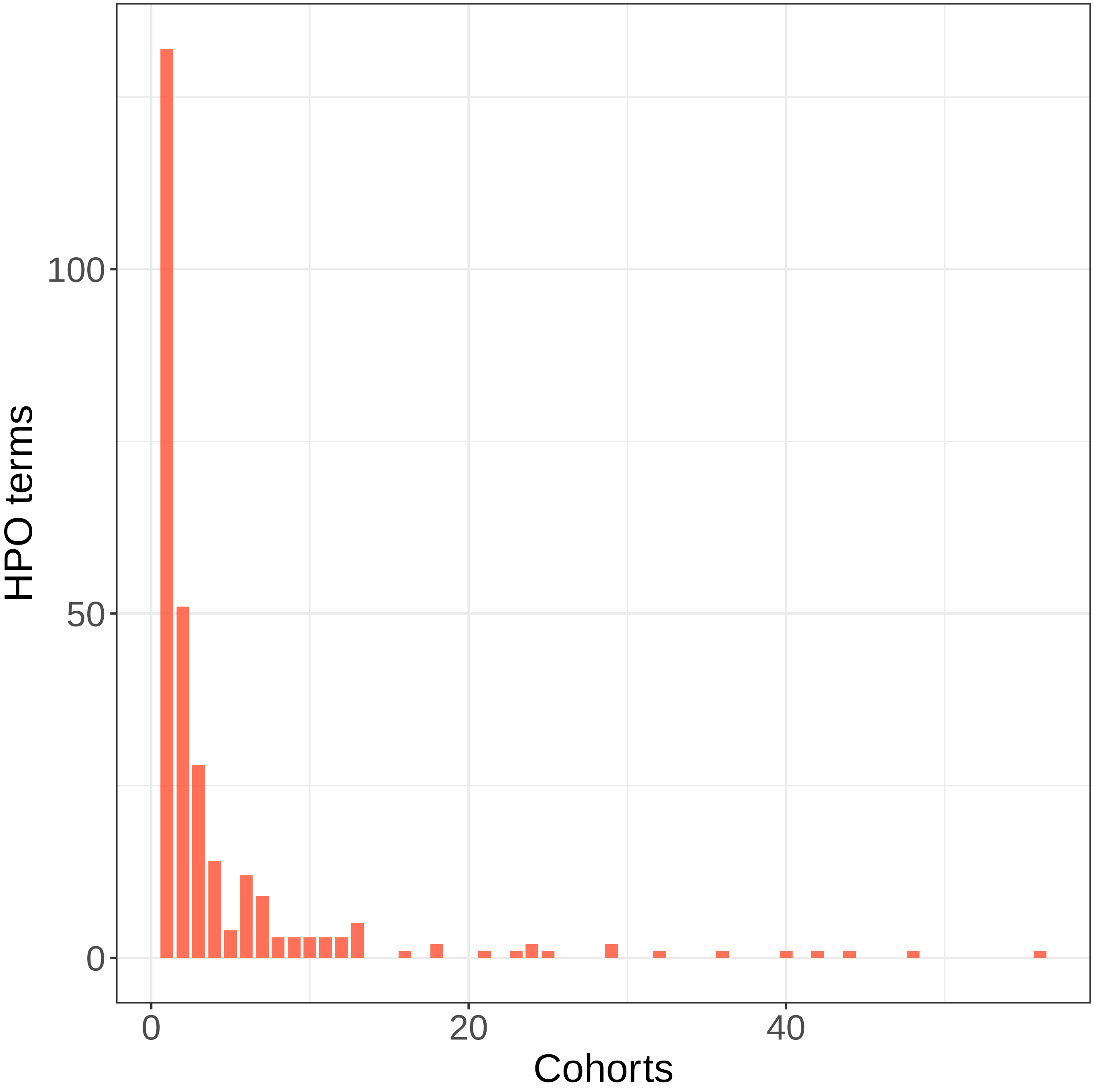


**Figure S1:** 81 cohorts were analyzed for this analysis, being annotated to a total of 287 HPO terms. 132 terms were used for only one cohort, 51 were used for two cohorts, and so on up to one term that was used for 56 cohorts (the maximum). 62 terms (21.6%) were used to annotate at least 5 cohorts.

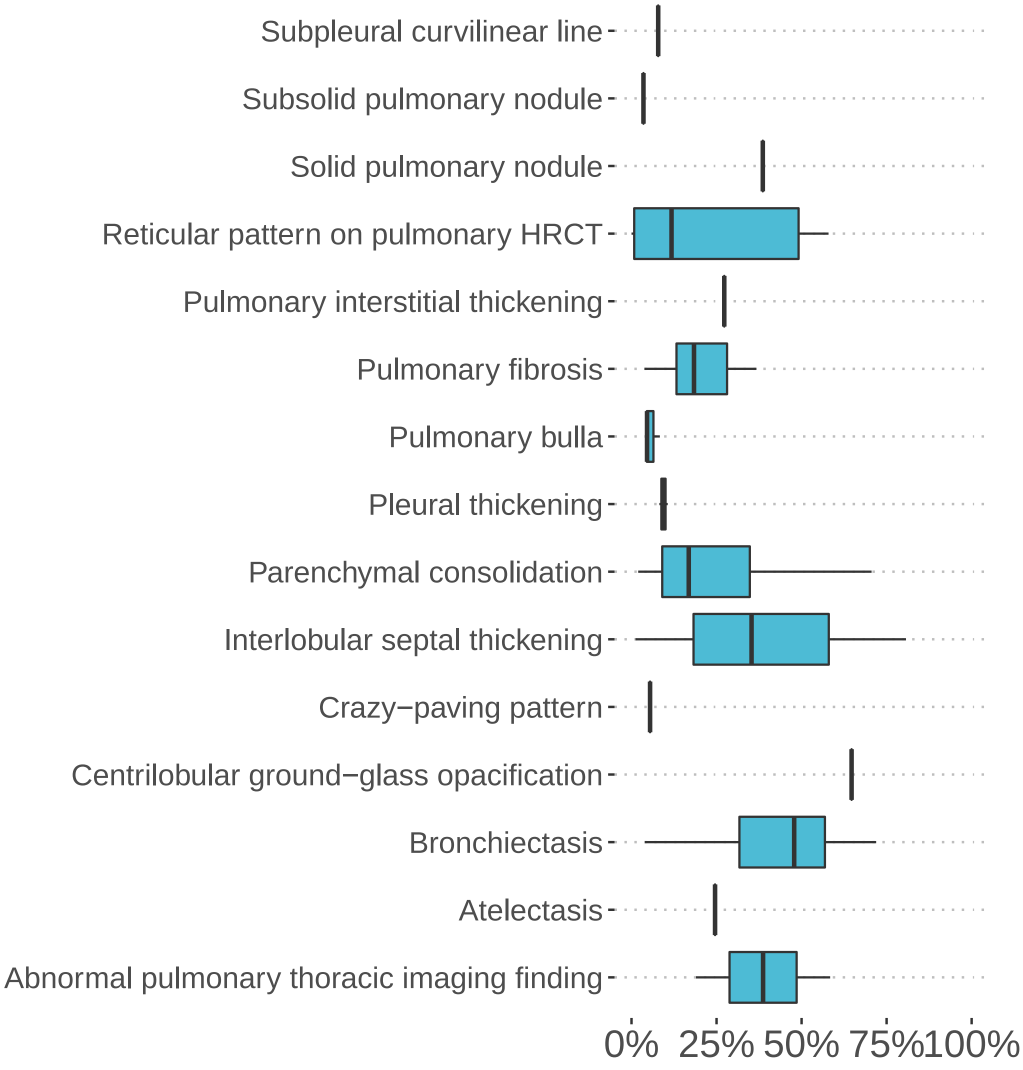


**Figure S2: Reported frequencies of pulmonary imaging findings in persons with long COVID**. Abnormal pulmonary thoracic imaging finding (HP:0031983, 2 studies, median percentage: 38.7%), Atelectasis (HP:0100750, 1 study, median percentage: 24.6%), Bronchiectasis (HP:0002110, 5 studies, median percentage: 47.8%), Centrilobular ground-glass opacification on pulmonary HRCT (HP:0025180, 1 study, median percentage: 64.7%), Crazy-paving pattern (HP:0033659, 1 study, median percentage: 5.5%), Interlobular septal thickening (HP:0030879, 3 studies, median percentage: 35.3%), Parenchymal consolidation (HP:0032177, 8 studies, median percentage: 16.8%), Pleural thickening (HP:0031944, 3 studies, median percentage: 9.4%), Pulmonary bulla (HP:0032446, 3 studies, median percentage: 4.6%), Pulmonary fibrosis (HP:0002206, 6 studies, median percentage: 18.4%), Pulmonary interstitial thickening (HP:0033711, 1 study, median percentage: 27.3%), Reticular pattern on pulmonary HRCT (HP:0025390, 5 studies, median percentage: 11.8%), Solid pulmonary nodule (HP:0033609, 1 study, median percentage: 38.6%), Subsolid pulmonary nodule (HP:0033610, 1 study, median percentage: 3.5%), Subpleural curvilinear line (HP:0033702, 1 study, median percentage: 7.8%)

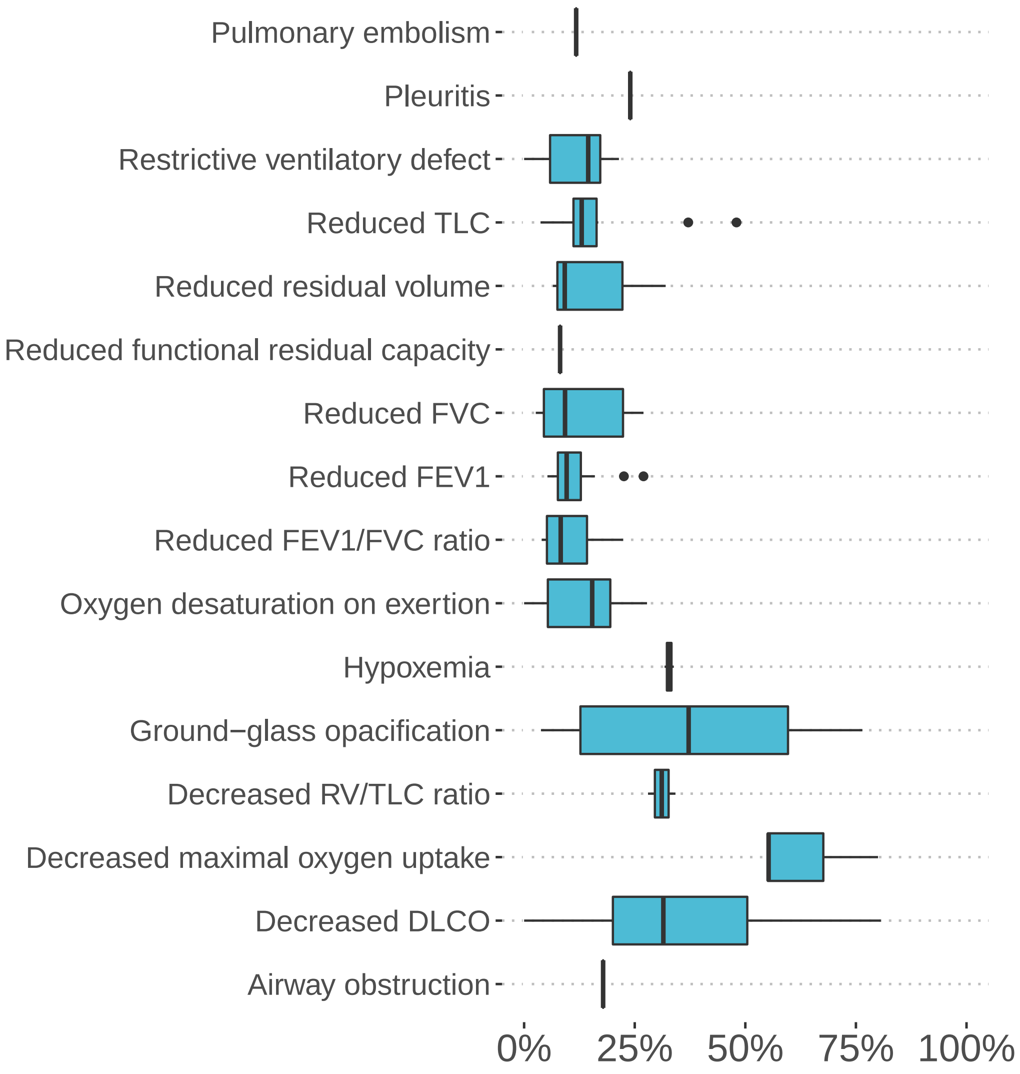


**Figure S3: Reported frequencies of pulmonary findings in persons with long COVID**. Airway obstruction (HP:0006536, 1 study, median percentage: 17.9%), Decreased DLCO (HP:0045051, 18 studies, median percentage: 31.5%), Decreased maximal oxygen uptake (HP:0033760, 3 studies, median percentage: 55.3%), Decreased RV/TLC ratio (HP:0033773, 2 studies, median percentage: 31.1%), Ground-glass opacification (HP:0025179, 13 studies, median percentage: 37.2%), Hypoxemia (HP:0012418, 2 studies, median percentage: 32.8%), Oxygen desaturation on exertion (HP:0030874, 7 studies, median percentage: 15.4%), Reduced FEV1/FVC ratio (HP:0030877, 7 studies, median percentage: 8.3%), Reduced forced expiratory volume in one second (HP:0032342, 12 studies, median percentage: 9.6%), Reduced forced vital capacity (HP:0032341, 8 studies, median percentage: 9.3%), Reduced functional residual capacity (HP:0033750, 1 study, median percentage: 8.1%), Reduced residual volume (HP:0033753, 6 studies, median percentage: 9.2%), Reduced total lung capacity (HP:0033169, 10 studies, median percentage: 13.0%), Restrictive ventilatory defect (HP:0002091, 6 studies, median percentage: 14.5%), Pleuritis (HP:0002102, 1 study, median percentage: 24.0%), Pulmonary embolism (HP:0002204, 1 study, median percentage: 11.8%)

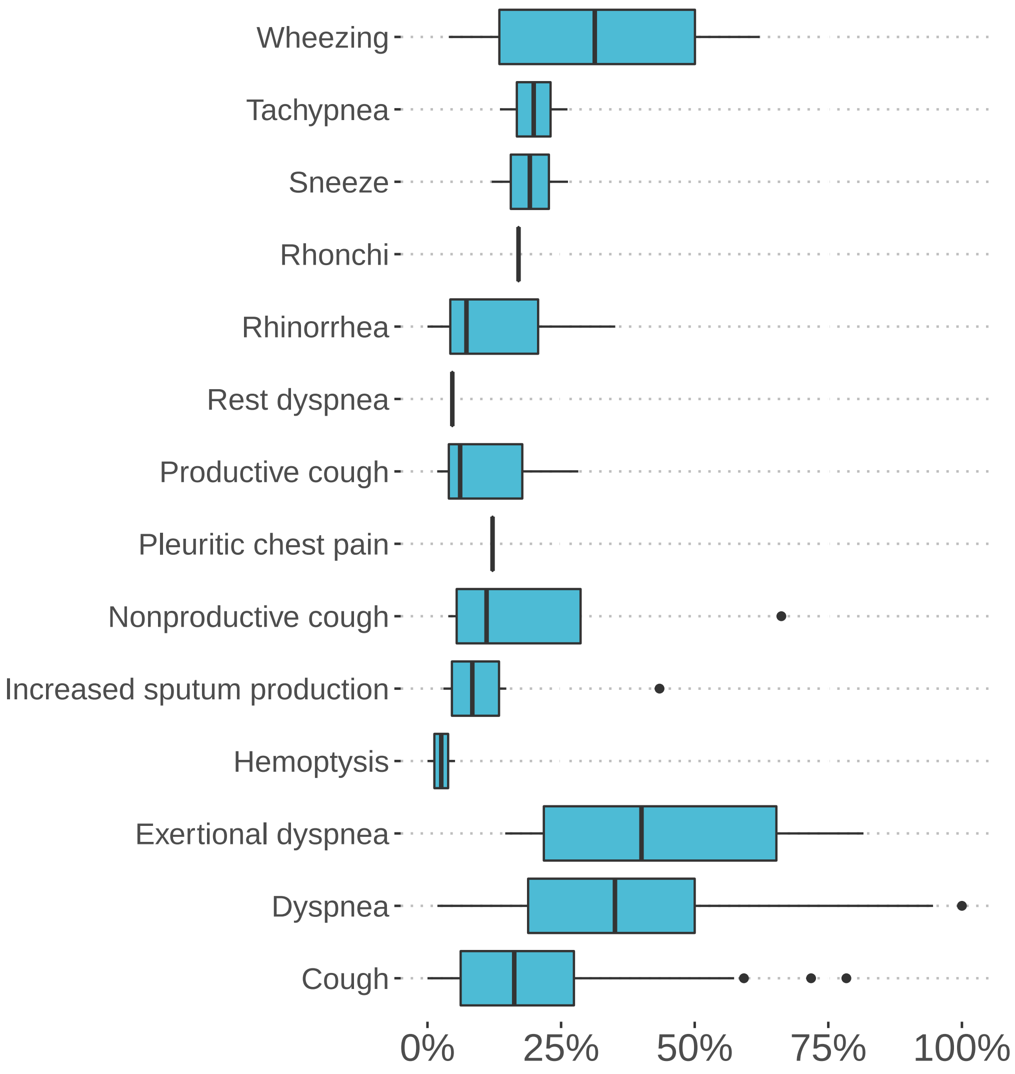


**Figure S4: Reported frequencies of respiratory symptoms in persons with long COVID**. Cough (HP:0012735, 40 studies, median percentage: 16.2%), Dyspnea (HP:0002094, 56 studies, median percentage: 35.1%), Exertional dyspnea (HP:0002875, 6 studies, median percentage: 40.0%), Hemoptysis (HP:0002105, 2 studies, median percentage: 2.6%), Increased sputum production (HP:0033709, 7 studies, median percentage: 8.4%), Nonproductive cough (HP:0031246, 4 studies, median percentage: 11.1%), Pleuritic chest pain (HP:0033771, 1 study, median percentage: 12.2%), Productive cough (HP:0031245, 5 studies, median percentage: 6.1%), Rest dyspnea (HP:0033710, 1 study, median percentage: 4.6%), Rhinorrhea (HP:0031417, 11 studies, median percentage: 7.3%), Rhonchi (HP:0030831, 1 study, median percentage: 17.0%), Sneeze (HP:0025095, 2 studies, median percentage: 19.2%), Tachypnea (HP:0002789, 2 studies, median percentage: 19.9%), Wheezing (HP:0030828, 4 studies, median percentage: 31.3%)

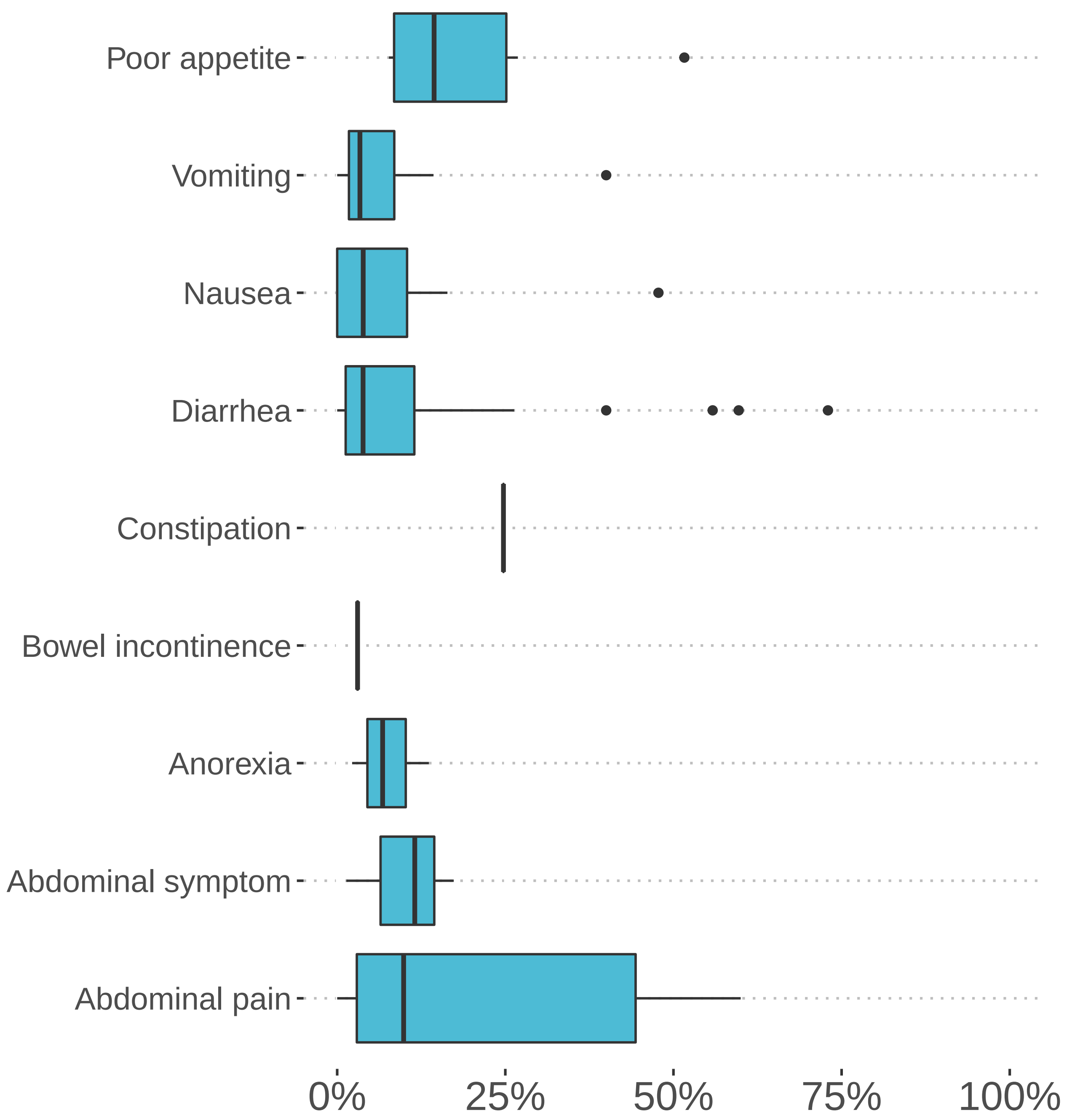


**Figure S5: Reported frequencies of gastrointestinal symptoms in persons with long COVID**. Abdominal pain (HP:0002027, 10 studies, median percentage: 9.9%), Abdominal symptom (HP:0011458, 3 studies, median percentage: 11.5%), Anorexia (HP:0002039, 3 studies, median percentage: 6.8%), Bowel incontinence (HP:0002607, 2 studies, median percentage: 3.0%), Constipation (HP:0002019, 1 study, median percentage: 24.7%), Diarrhea (HP:0002014, 29 studies, median percentage: 3.8%), Early satiety (HP:0033842, 1 study, median percentage: 30.8%), Nausea (HP:0002018, 16 studies, median percentage: 3.9%), Vomiting (HP:0002013, 12 studies, median percentage: 3.4%), Poor appetite (HP:0004396, 6 studies, median percentage: 14.4%)

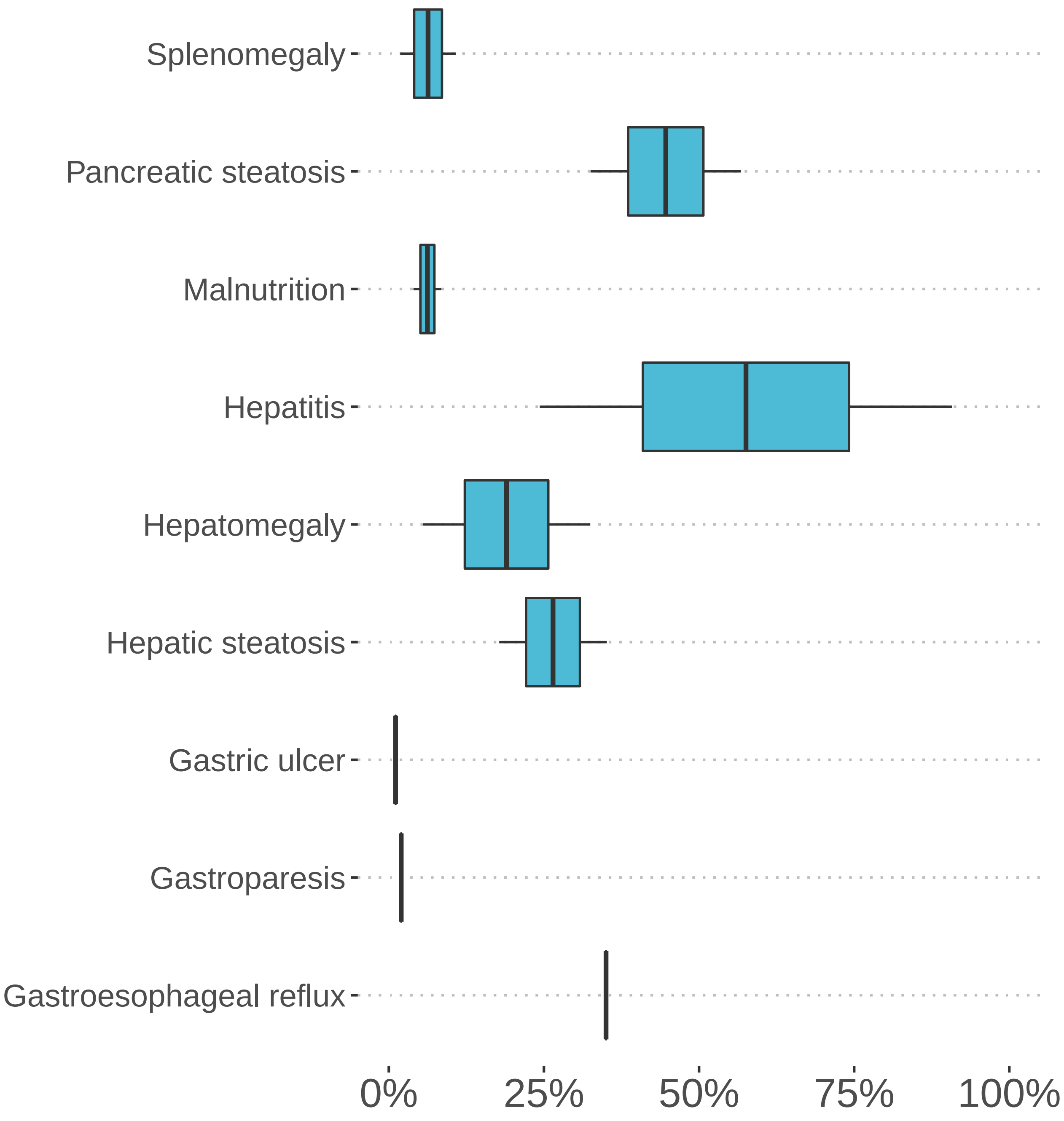


**Figure S6: Reported frequencies of gastrointestinal findings in persons with long COVID**. Gastroesophageal reflux (HP:0002020, 1 study, median percentage: 35.0%), Gastroparesis (HP:0002578, 1 study, median percentage: 2.0%), Gastric ulcer (HP:0002592, 1 study, median percentage: 1.1%), Hepatic steatosis (HP:0001397, 2 studies, median percentage: 26.5%), Hepatomegaly (HP:0002240, 2 studies, median percentage: 19.0%), Hepatitis (HP:0012115, 2 studies, median percentage: 57.6%), Malnutrition (HP:0004395, 2 studies, median percentage: 6.2%), Pancreatic steatosis (HP:0033757, 2 studies, median percentage: 44.6%), Splenomegaly (HP:0001744, 2 studies, median percentage: 6.3%)

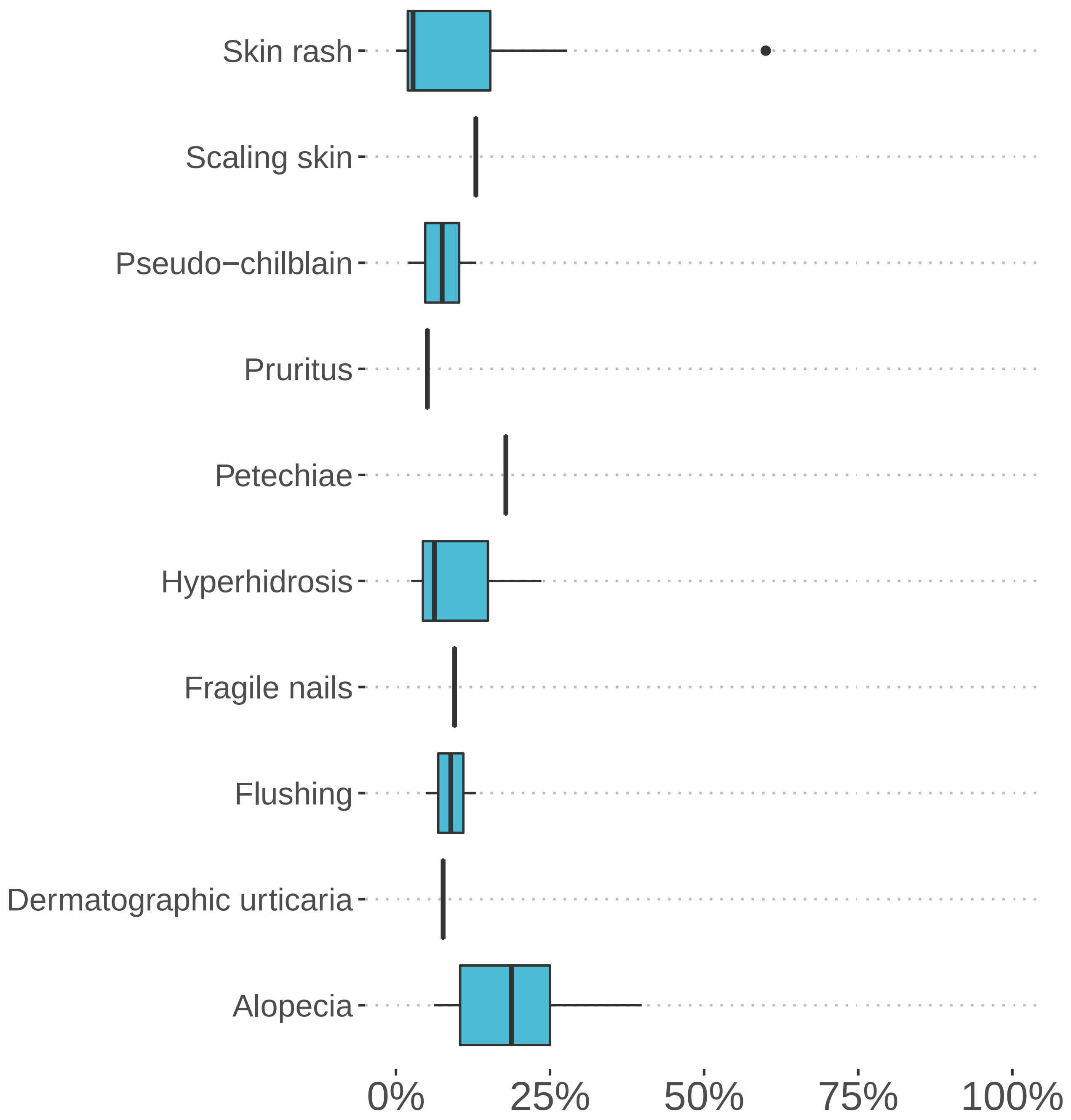


**Figure S7: Reported frequencies of dermatological findings in persons with long COVID**. Alopecia (HP:0001596, 9 studies, median percentage: 18.8%), Dermatographic urticaria (HP:0011971, 1 study, median percentage: 7.7%), Flushing (HP:0031284, 2 studies, median percentage: 8.9%), Fragile nails (HP:0001808, 1 study, median percentage: 9.5%), Hyperhidrosis (HP:0000975, 3 studies, median percentage: 6.3%), Petechiae (HP:0000967, 1 study, median percentage: 17.8%), Pruritus (HP:0000989, 1 study, median percentage: 5.1%), Pseudo-chilblain (HP:0033696, 2 studies, median percentage: 7.5%), Scaling skin (HP:0040189, 1 study, median percentage: 13.0%), Skin rash (HP:0000988, 7 studies, median percentage: 2.8%)

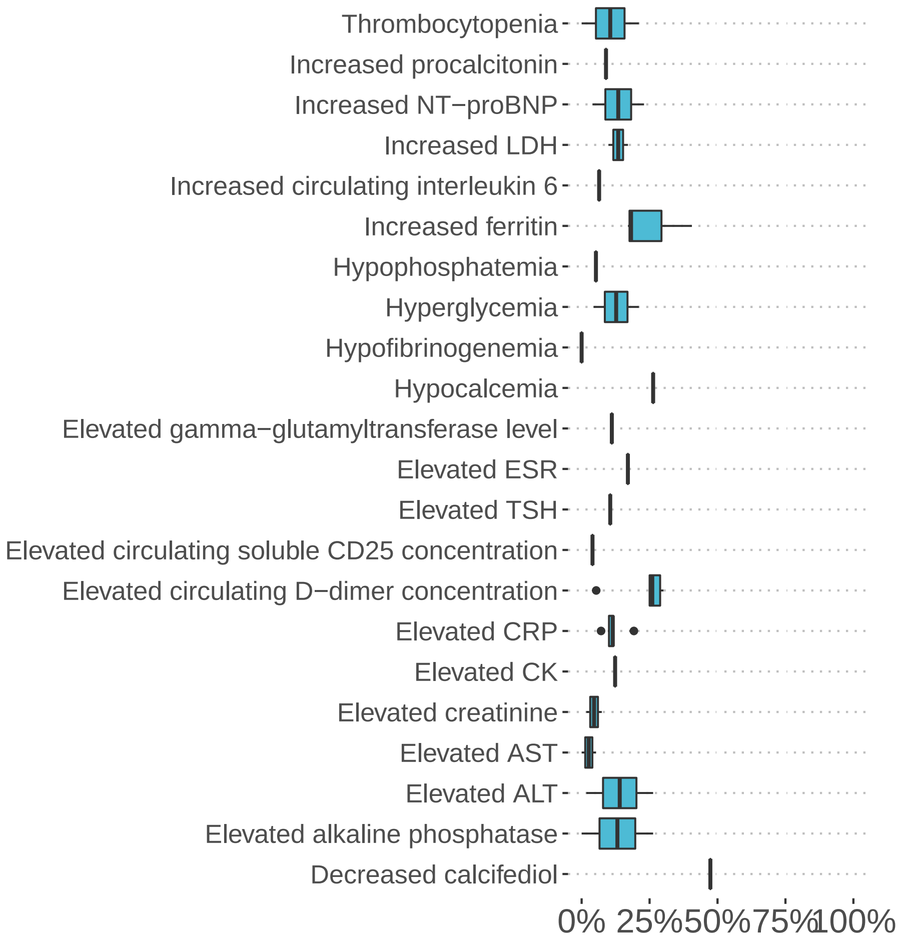


**Figure S8: Reported frequencies of laboratory abnormalities in persons with long COVID**. Decreased circulating calcifediol concentration (HP:0012053, 1 study, median percentage: 47.4%), Elevated circulating alkaline phosphatase concentration (HP:0003155, 2 studies, median percentage: 13.2%), Elevated circulating alanine aminotransferase concentration (HP:0031964, 2 studies, median percentage: 14.0%), Elevated circulating aspartate aminotransferase concentration (HP:0031956, 2 studies, median percentage: 2.6%), Elevated circulating creatinine concentration (HP:0003259, 2 studies, median percentage: 4.6%), Elevated circulating creatine kinase concentration (HP:0003236, 1 study, median percentage: 12.3%), Elevated circulating C-reactive protein concentration (HP:0011227, 6 studies, median percentage: 11.5%), Elevated circulating D-dimer concentration (HP:0033106, 6 studies, median percentage: 26.0%), Elevated circulating soluble CD25 concentration (HP:0033833, 1 study, median percentage: 4.0%), Elevated circulating thyroid-stimulating hormone concentration (HP:0002925, 1 study, median percentage: 10.5%), Elevated erythrocyte sedimentation rate (HP:0003565, 1 study, median percentage: 17.0%), Elevated gamma-glutamyltransferase level (HP:0030948, 1 study, median percentage: 11.1%), Hypocalcemia (HP:0002901, 1 study, median percentage: 26.3%), Hypofibrinogenemia (HP:0011900, 1 study, median percentage: 0.0%), Hyperglycemia (HP:0003074, 2 studies, median percentage: 12.7%), Hypoglycemia (HP:0001943, 1 study, median percentage: 1.7%), Hypophosphatemia (HP:0002148, 1 study, median percentage: 5.3%), Increased circulating ferritin concentration (HP:0003281, 3 studies, median percentage: 18.2%), Increased circulating interleukin 6 (HP:0030783, 2 studies, median percentage: 6.4%), Increased circulating lactate dehydrogenase concentration (HP:0025435, 2 studies, median percentage: 13.5%), Increased circulating NT-proBNP concentration (HP:0031185, 2 studies, median percentage: 13.5%), Increased circulating procalcitonin concentration (HP:0032308, 1 study, median percentage: 9.0%), Thrombocytopenia (HP:0001873, 2 studies, median percentage: 10.5%)

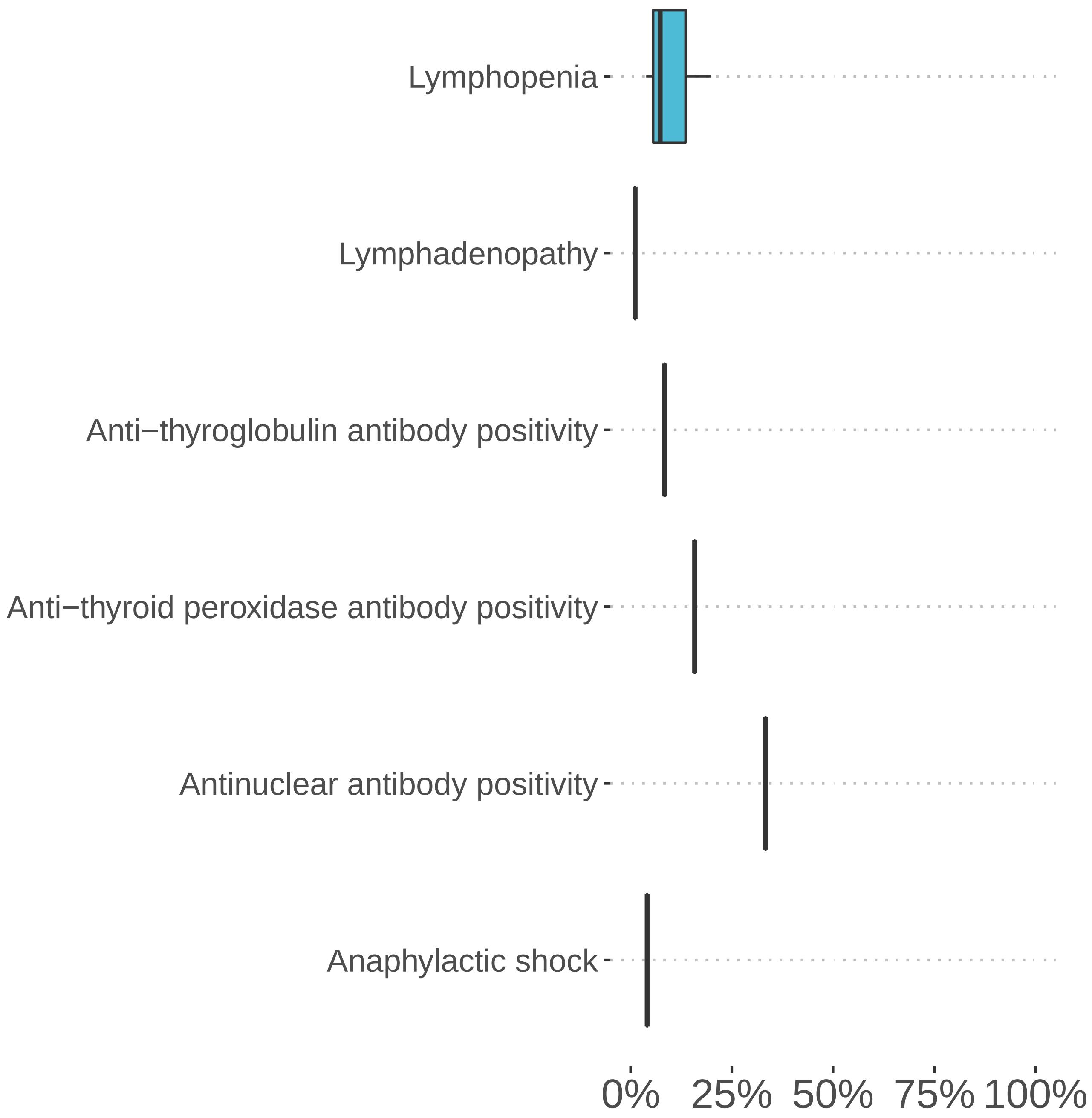


**Figure S9: Reported frequencies of immunology/autoimmunity findings in persons with long COVID**. Anaphylactic shock (HP:0100845, 1 study, median percentage: 4.1%), Antinuclear antibody positivity (HP:0003493, 1 study, median percentage: 33.3%), Anti-thyroid peroxidase antibody positivity (HP:0025379, 1 study, median percentage: 15.8%), Anti-thyroglobulin antibody positivity (HP:0032069, 1 study, median percentage: 8.4%), Lymphadenopathy (HP:0002716, 1 study, median percentage: 1.1%), Lymphopenia (HP:0001888, 3 studies, median percentage: 7.3%)

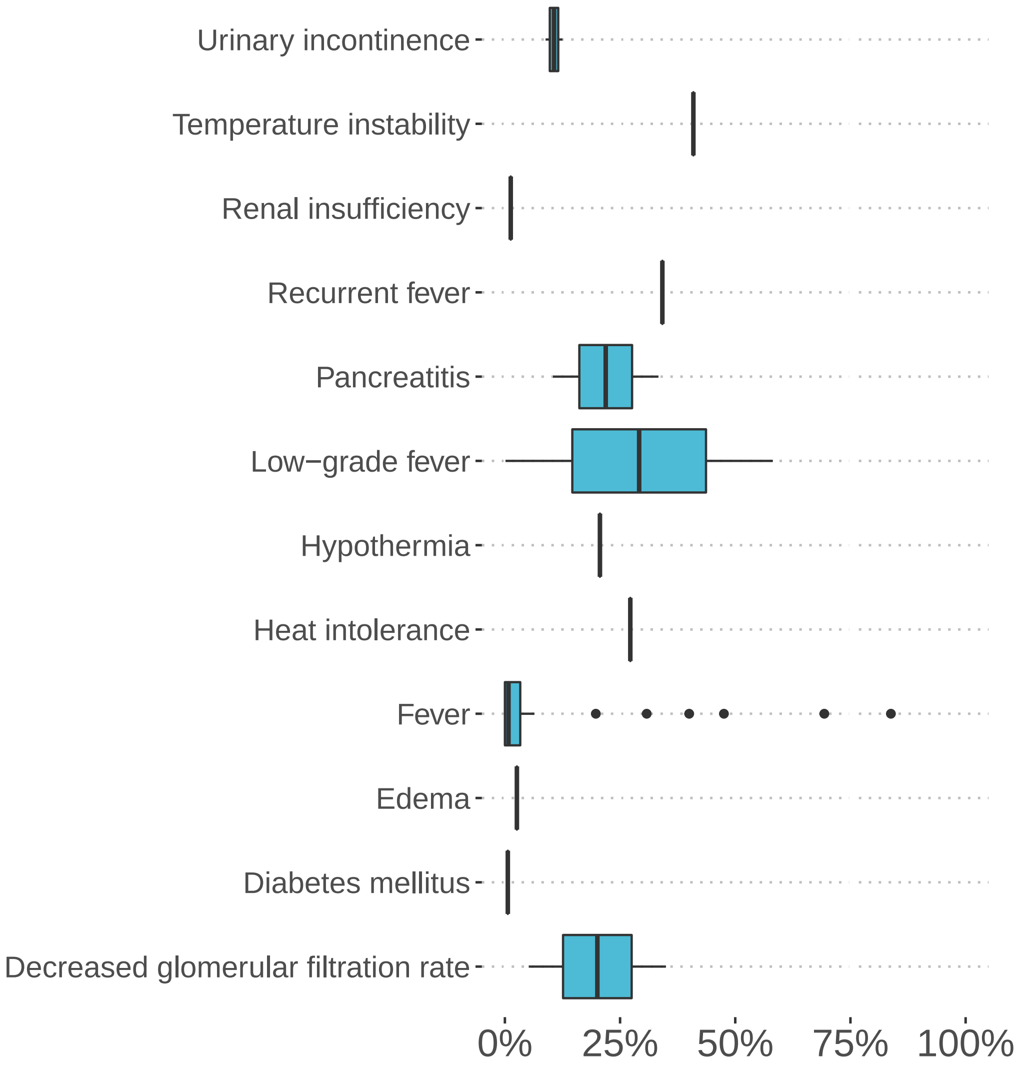


**Figure S10: Reported frequencies of reproductive, genitourinary, endocrine, or metabolism findings in persons with long COVID**. Decreased glomerular filtration rate (HP:0012213, 2 studies, median percentage: 20.1%), Diabetes mellitus (HP:0000819, 1 study, median percentage: 0.6%), Edema (HP:0000969, 1 study, median percentage: 2.6%), Female sexual dysfunction (HP:0030014, 1 study, median percentage: 8.0%), Fever (HP:0001945, 29 studies, median percentage: 0.8%), Heat intolerance (HP:0002046, 1 study, median percentage: 27.2%), Hypothermia (HP:0002045, 1 study, median percentage: 20.6%), Irregular menstruation (HP:0000858, 1 study, median percentage: 26.5%), Low-grade fever (HP:0011134, 2 studies, median percentage: 29.1%), Male sexual dysfunction (HP:0040307, 1 study, median percentage: 14.6%), Menorrhagia (HP:0000132, 1 study, median percentage: 20.1%), Pancreatitis (HP:0001733, 2 studies, median percentage: 21.9%), Postmenopausal bleeding (HP:0033840, 1 study, median percentage: 3.1%), Recurrent fever (HP:0001954, 1 study, median percentage: 34.2%), Renal insufficiency (HP:0000083, 1 study, median percentage: 1.3%), Temperature instability (HP:0005968, 1 study, median percentage: 40.9%), Testicular pain (HP:0033839, 1 study, median percentage: 10.3%), Urinary incontinence (HP:0000020, 3 studies, median percentage: 12.5%)

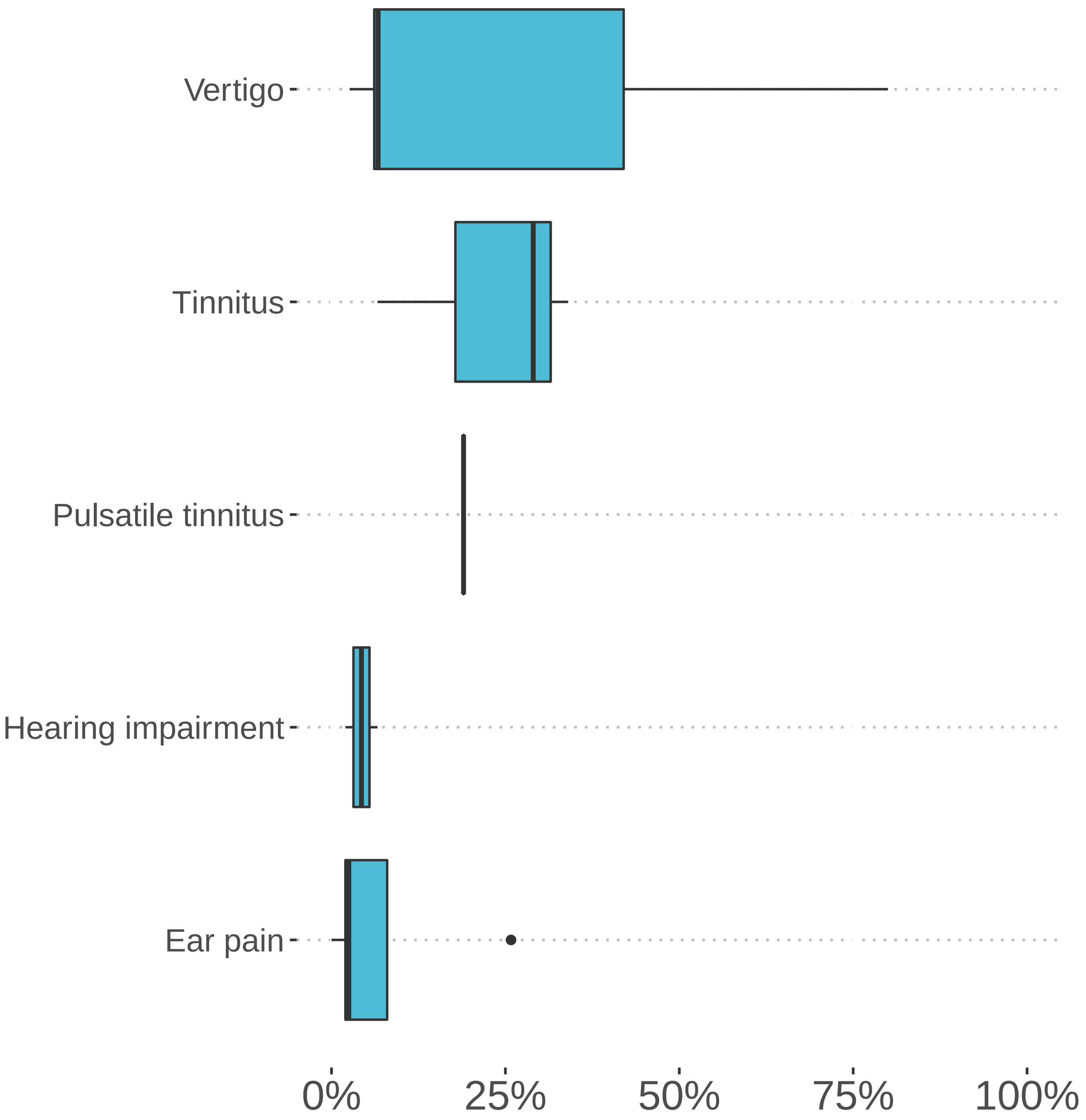


**Figure S11: Reported frequencies of ear disorders in persons with long COVID**. Ear pain (HP:0030766, 5 studies, median percentage: 2.5%), Hearing impairment (HP:0000365, 3 studies, median percentage: 6.6%), Hyperacusis (HP:0010780, 1 study, median percentage: 34.7%), Pulsatile tinnitus (HP:0008629, 1 study, median percentage: 19.0%), Tinnitus (HP:0000360, 3 studies, median percentage: 29.0%), Vertigo (HP:0002321, 10 studies, median percentage: 6.7%)

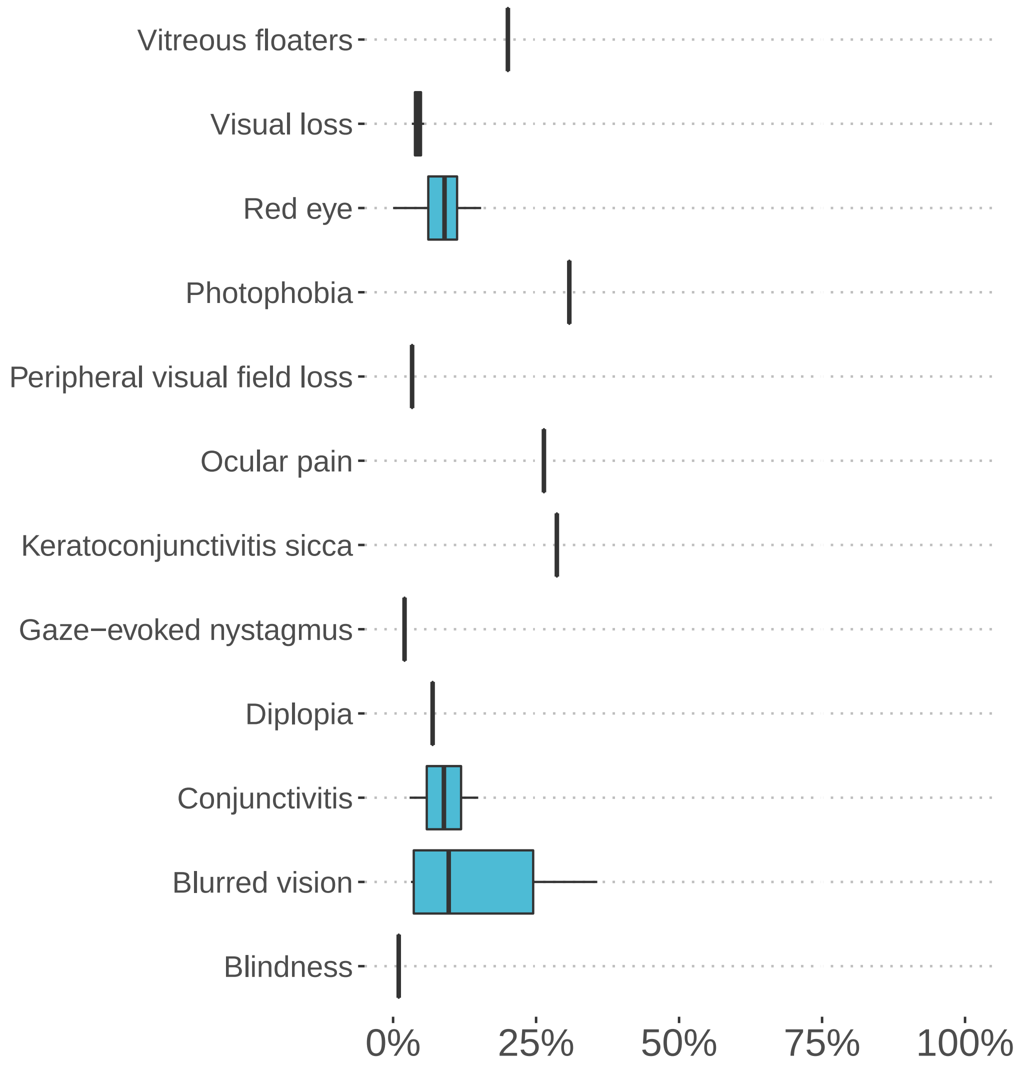


**Figure S12: Reported frequencies of ocular abnormalities in persons with long COVID**. Blindness (HP:0000618, 1 study, median percentage: 1.0%), Blurred vision (HP:0000622, 7 studies, median percentage: 9.7%), Conjunctivitis (HP:0000509, 2 studies, median percentage: 8.9%), Diplopia (HP:0000651, 1 study, median percentage: 6.9%), Gaze-evoked nystagmus (HP:0000640, 1 study, median percentage: 2.0%), Keratoconjunctivitis sicca (HP:0001097, 1 study, median percentage: 28.6%), Ocular pain (HP:0200026, 1 study, median percentage: 26.4%), Ocular pruritus (HP:0033841, 1 study, median percentage: 24.2%), Peripheral visual field loss (HP:0007994, 1 study, median percentage: 3.3%), Photophobia (HP:0000613, 1 study, median percentage: 30.8%), Red eye (HP:0025337, 4 studies, median percentage: 9.0%), Visual loss (HP:0000572, 2 studies, median percentage: 4.3%), Vitreous floaters (HP:0100832, 1 study, median percentage: 20.1%)

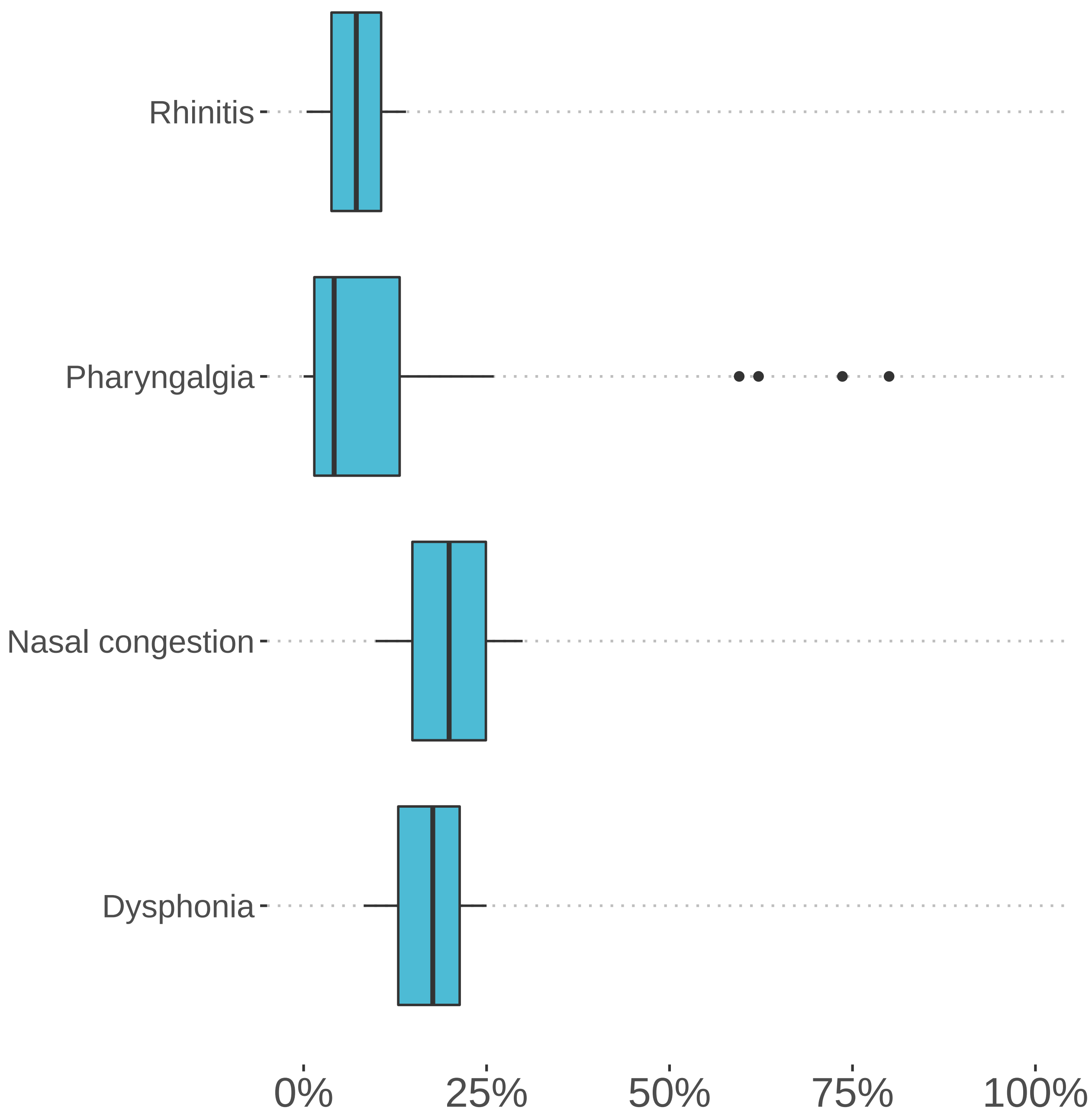


**Figure S13: Reported frequencies of nose and throat symptoms in persons with long COVID**. Dysphonia (HP:0001618, 4 studies, median percentage: 21.3%), Nasal congestion (HP:0001742, 2 studies, median percentage: 19.9%), Pharyngalgia (HP:0033050, 23 studies, median percentage: 4.2%), Rhinitis (HP:0012384, 2 studies, median percentage: 7.2%)

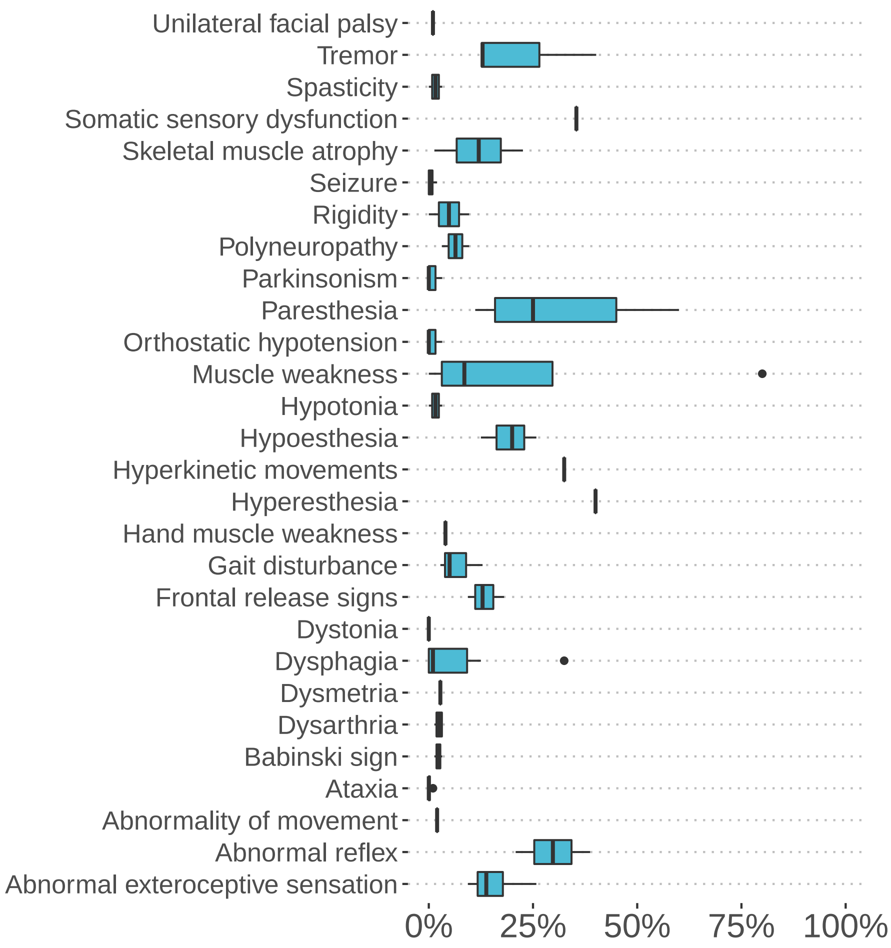


**Figure S14: Reported frequencies of neuropsychiatric findings in persons with long COVID**. Abnormal exteroceptive sensation (HP:0033747, 4 studies, median percentage: 13.8%), Abnormal reflex (HP:0031826, 2 studies, median percentage: 29.8%), Abnormality of movement (HP:0100022, 1 study, median percentage: 2.0%), Ataxia (HP:0001251, 4 studies, median percentage: 0.0%), Babinski sign (HP:0003487, 2 studies, median percentage: 2.3%), Dysarthria (HP:0001260, 4 studies, median percentage: 2.6%), Dysmetria (HP:0001310, 1 study, median percentage: 2.8%), Dysphagia (HP:0002015, 7 studies, median percentage: 1.0%), Dystonia (HP:0001332, 2 studies, median percentage: 0.0%), Facial paralysis (HP:0007209, 1 study, median percentage: 3.4%), Frontal release signs (HP:0000743, 3 studies, median percentage: 12.9%), Gait disturbance (HP:0001288, 3 studies, median percentage: 5.0%), Hand muscle weakness (HP:0030237, 1 study, median percentage: 4.0%), Hyperesthesia (HP:0100963, 1 study, median percentage: 40.0%), Hyperkinetic movements (HP:0002487, 1 study, median percentage: 32.5%), Hypoesthesia (HP:0033748, 4 studies, median percentage: 22.9%), Hypotonia (HP:0001252, 2 studies, median percentage: 1.6%), Muscle spasm (HP:0003394, 1 study, median percentage: 32.5%), Muscle weakness (HP:0001324, 4 studies, median percentage: 8.5%), Orthostatic hypotension (HP:0001278, 3 studies, median percentage: 0.0%), Paresthesia (HP:0003401, 6 studies, median percentage: 25.0%), Parkinsonism (HP:0001300, 3 studies, median percentage: 0.0%), Polyneuropathy (HP:0001271, 2 studies, median percentage: 6.4%), Rigidity (HP:0002063, 2 studies, median percentage: 4.8%), Seizure (HP:0001250, 6 studies, median percentage: 0.3%), Skeletal muscle atrophy (HP:0003202, 2 studies, median percentage: 12.0%), Somatic sensory dysfunction (HP:0003474, 1 study, median percentage: 35.4%), Spasticity (HP:0001257, 2 studies, median percentage: 1.6%), Tremor (HP:0001337, 3 studies, median percentage: 12.9%), Unilateral facial palsy (HP:0012799, 1 study, median percentage: 1.0%)

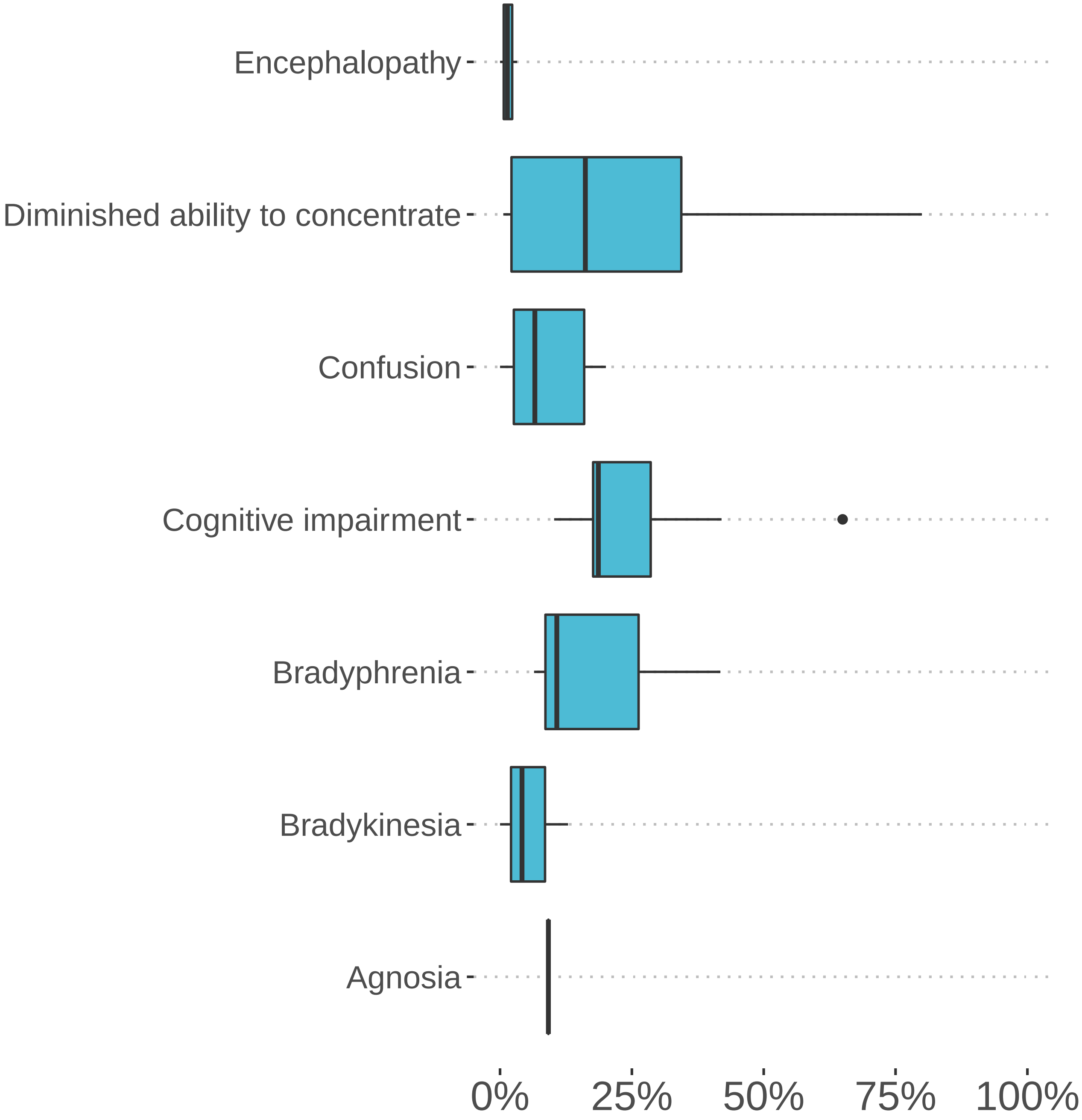


**Figure S15: Reported frequencies of cognitive dysfunction in persons with long COVID**. Agnosia (HP:0010524, 1 study, median percentage: 9.2%), Bradykinesia (HP:0002067, 3 studies, median percentage: 4.2%), Bradyphrenia (HP:0031843, 3 studies, median percentage: 10.8%), Cognitive impairment (HP:0100543, 13 studies, median percentage: 18.6%), Confusion (HP:0001289, 6 studies, median percentage: 6.6%), Diminished ability to concentrate (HP:0031987, 9 studies, median percentage: 16.2%), Encephalopathy (HP:0001298, 3 studies, median percentage: 1.4%), Tachyphrenia (HP:0033844, 1 study, median percentage: 15.2%)

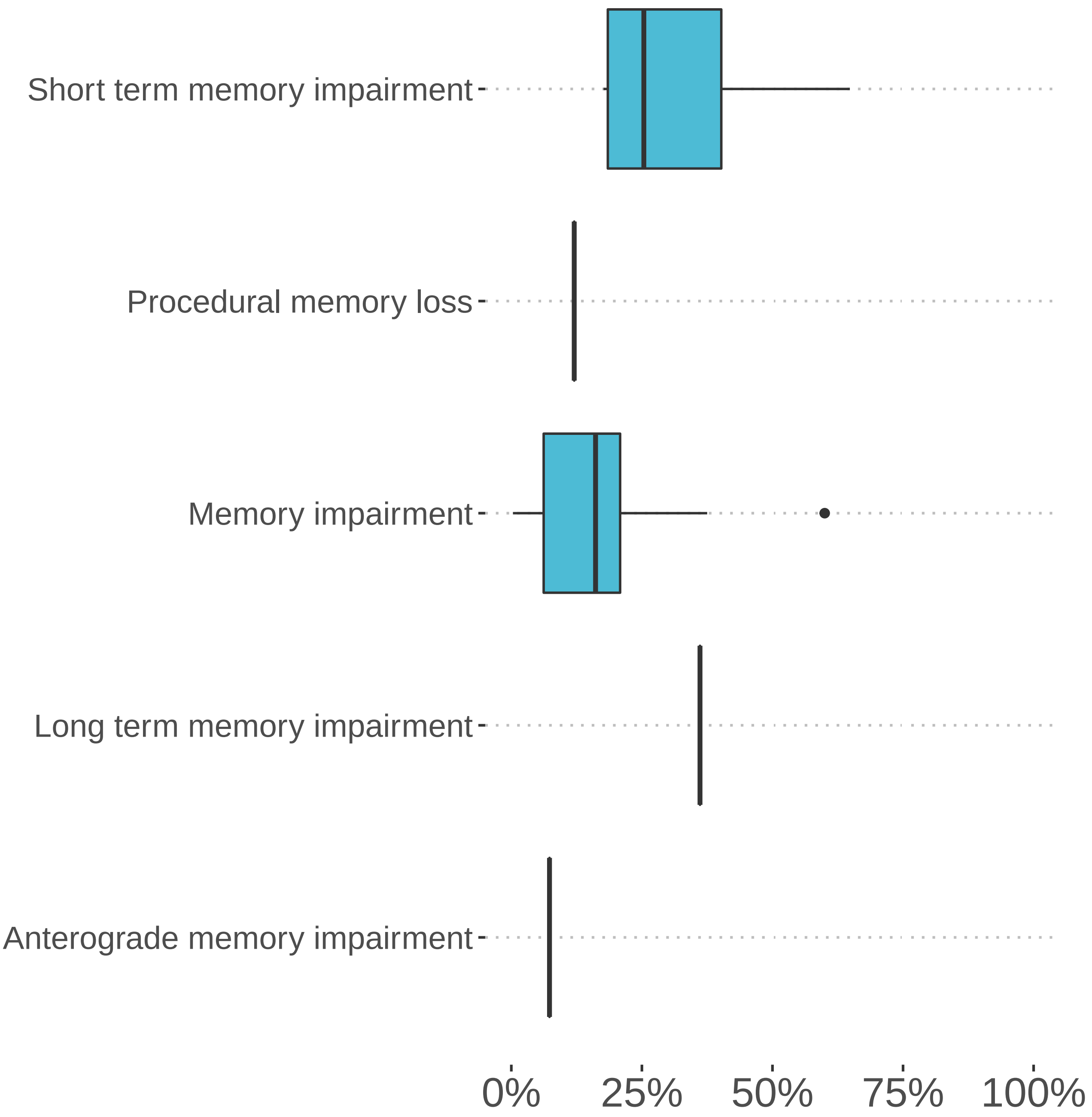


**Figure S16: Reported frequencies of memory impairment in persons with long COVID**. Anterograde memory impairment (HP:0033689, 1 study, median percentage: 7.3%), Long term memory impairment (HP:0033688, 1 study, median percentage: 36.1%), Memory impairment (HP:0002354, 13 studies, median percentage: 16.1%), Procedural memory loss (HP:0033691, 1 study, median percentage: 12.0%), Short term memory impairment (HP:0033687, 4 studies, median percentage: 25.4%)

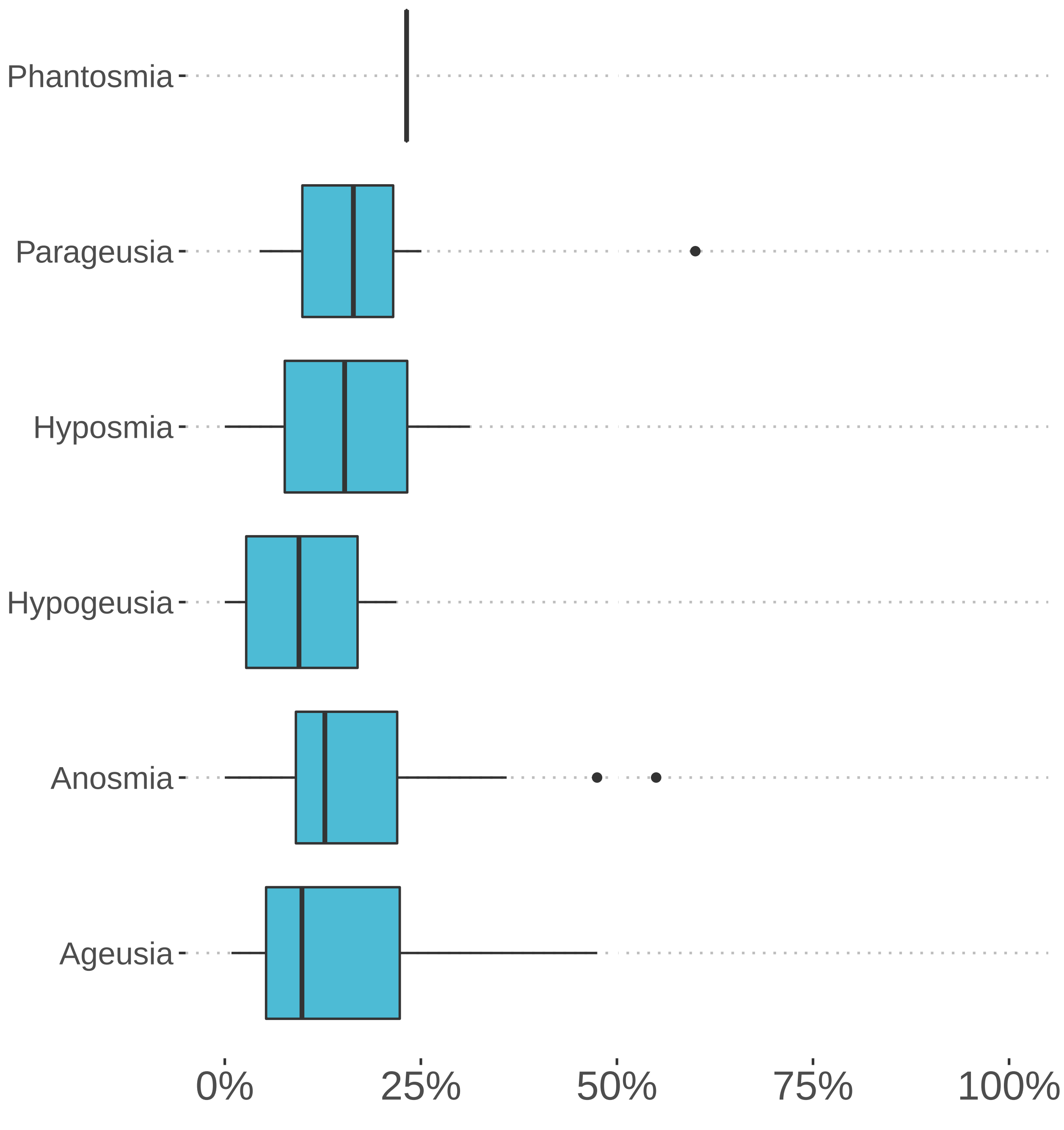


**Figure S17: Reported frequencies of abnormalities of smell and taste in persons with long COVID**. Ageusia (HP:0041051, 21 studies, median percentage: 9.8%), Anosmia (HP:0000458, 44 studies, median percentage: 12.8%), Hypogeusia (HP:0000224, 4 studies, median percentage: 9.5%), Hyposmia (HP:0004409, 3 studies, median percentage: 15.3%), Parageusia (HP:0031249, 11 studies, median percentage: 16.4%), Phantageusia (HP:0033847, 1 study, median percentage: 9.0%), Phantosmia (HP:0033693, 1 study, median percentage: 23.2%)

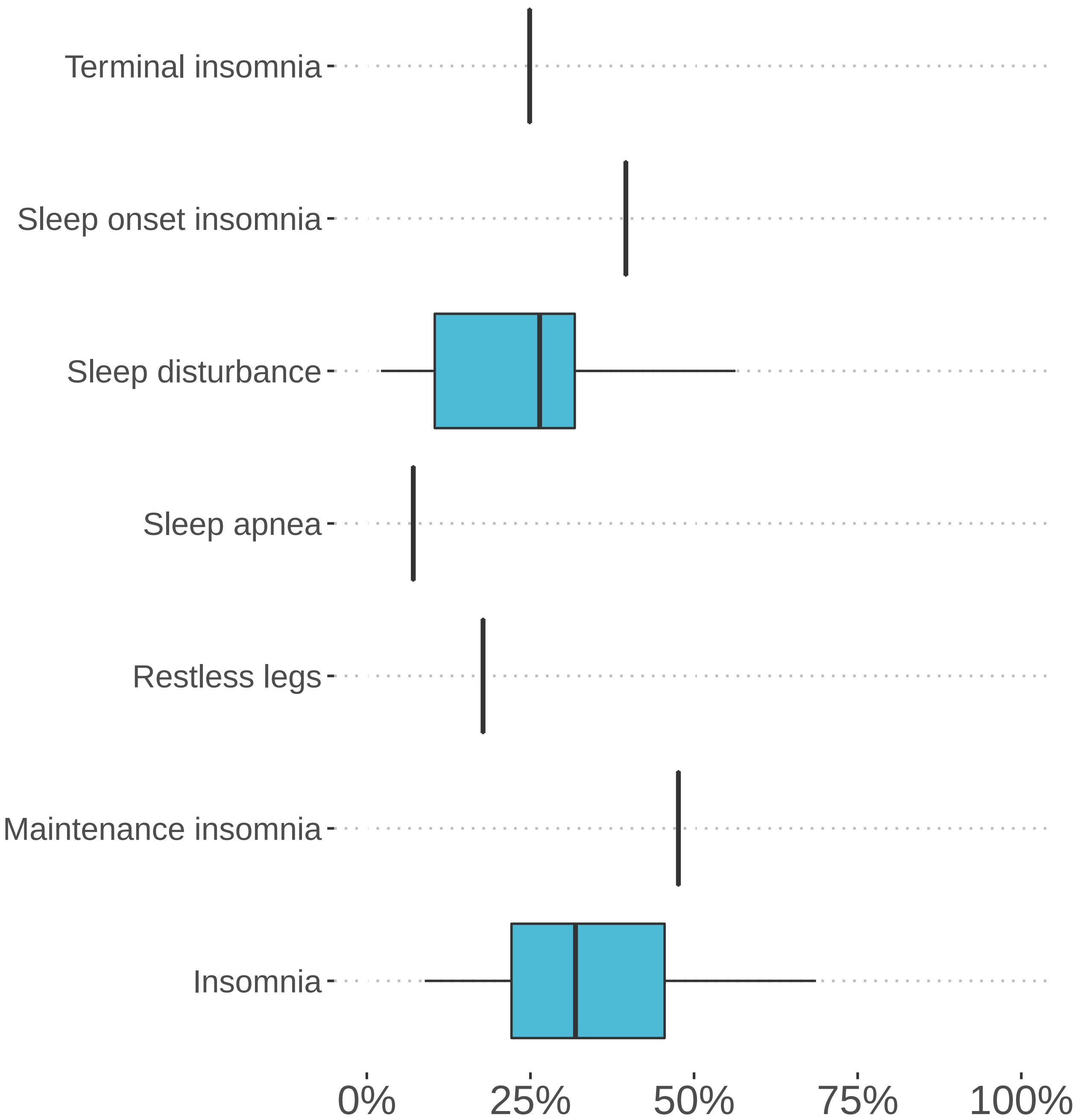


**Figure S18: Reported frequencies of sleep impairment in persons with long COVID**. Insomnia (HP:0100785, 12 studies, median percentage: 31.9%), Maintenance insomnia (HP:0031355, 1 study, median percentage: 47.6%), Restless legs (HP:0012452, 1 study, median percentage: 17.8%), Sleep apnea (HP:0010535, 1 study, median percentage: 7.1%), Sleep disturbance (HP:0002360, 11 studies, median percentage: 26.4%), Sleep onset insomnia (HP:0031354, 1 study, median percentage: 39.6%), Terminal insomnia (HP:0031356, 1 study, median percentage: 24.9%)

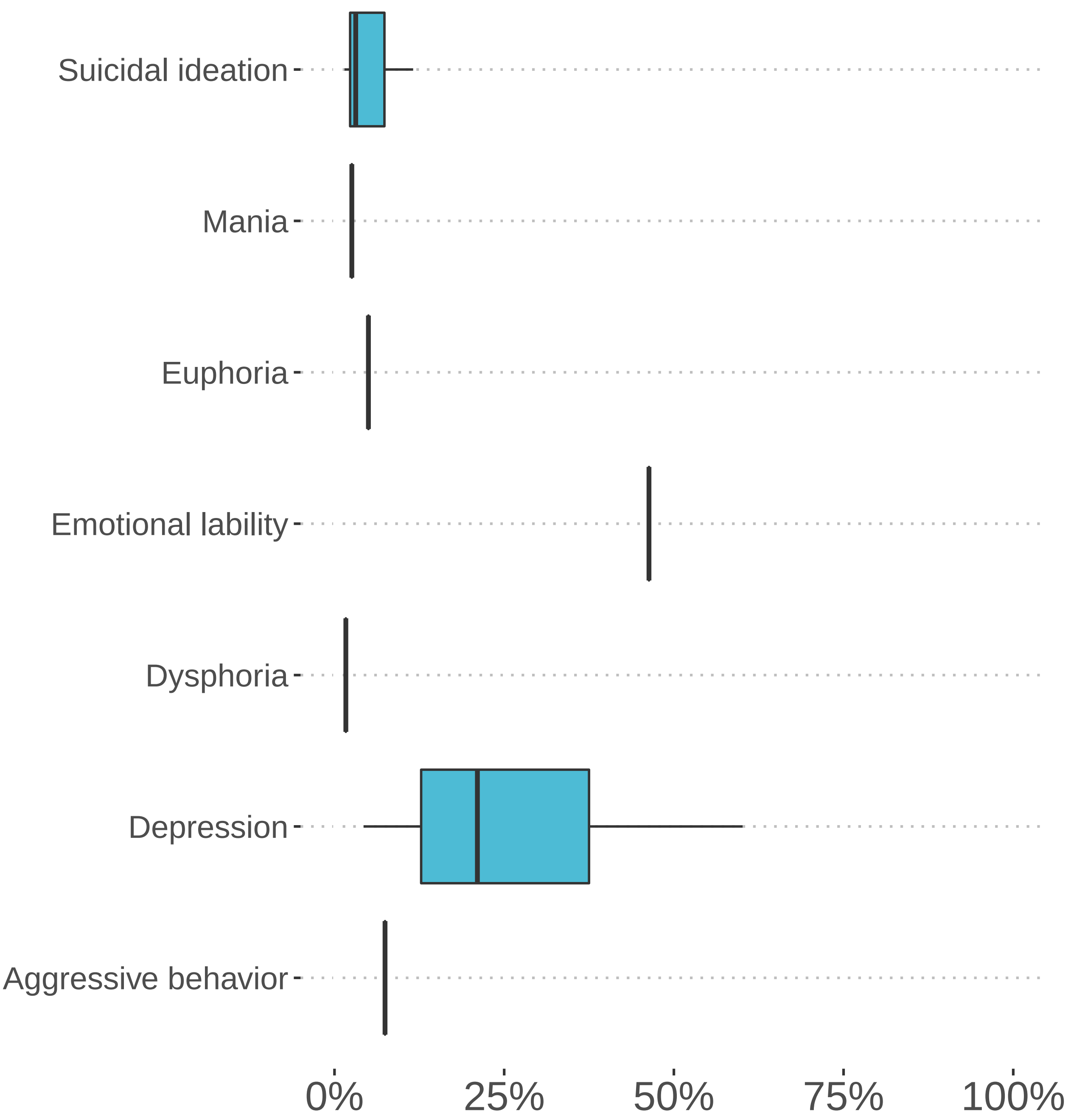


**Figure S19: Reported frequencies of emotion/mood abnormalities in persons with long COVID**. Aggressive behavior (HP:0000718, 1 study, median percentage: 7.4%), Depression (HP:0000716, 25 studies, median percentage: 21.1%), Dysphoria (HP:0033838, 1 study, median percentage: 1.7%), Emotional lability (HP:0000712, 1 study, median percentage: 46.3%), Euphoria (HP:0031844, 1 study, median percentage: 5.0%), Mania (HP:0100754, 1 study, median percentage: 2.6%), Sense of impending doom (HP:0033845, 1 study, median percentage: 33.7%), Suicidal ideation (HP:0031589, 3 studies, median percentage: 3.1%), Tearfulness (HP:0033705, 1 study, median percentage: 42.5%)

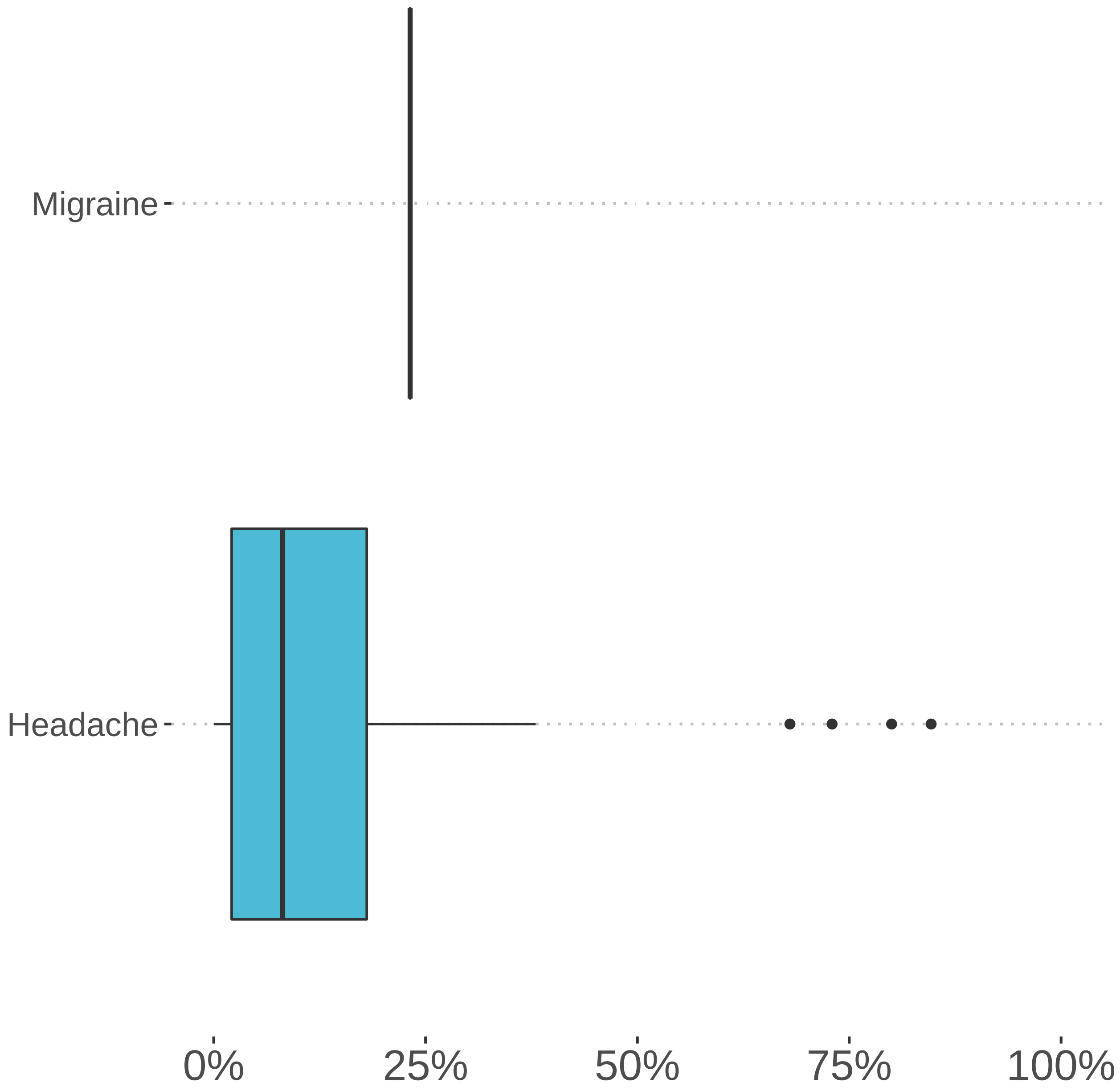


**Figure S20: Reported frequencies of headache in persons with long COVID**. Headache (HP:0002315, 42 studies, median percentage: 8.1%), Migraine (HP:0002076, 1 study, median percentage: 23.2%)

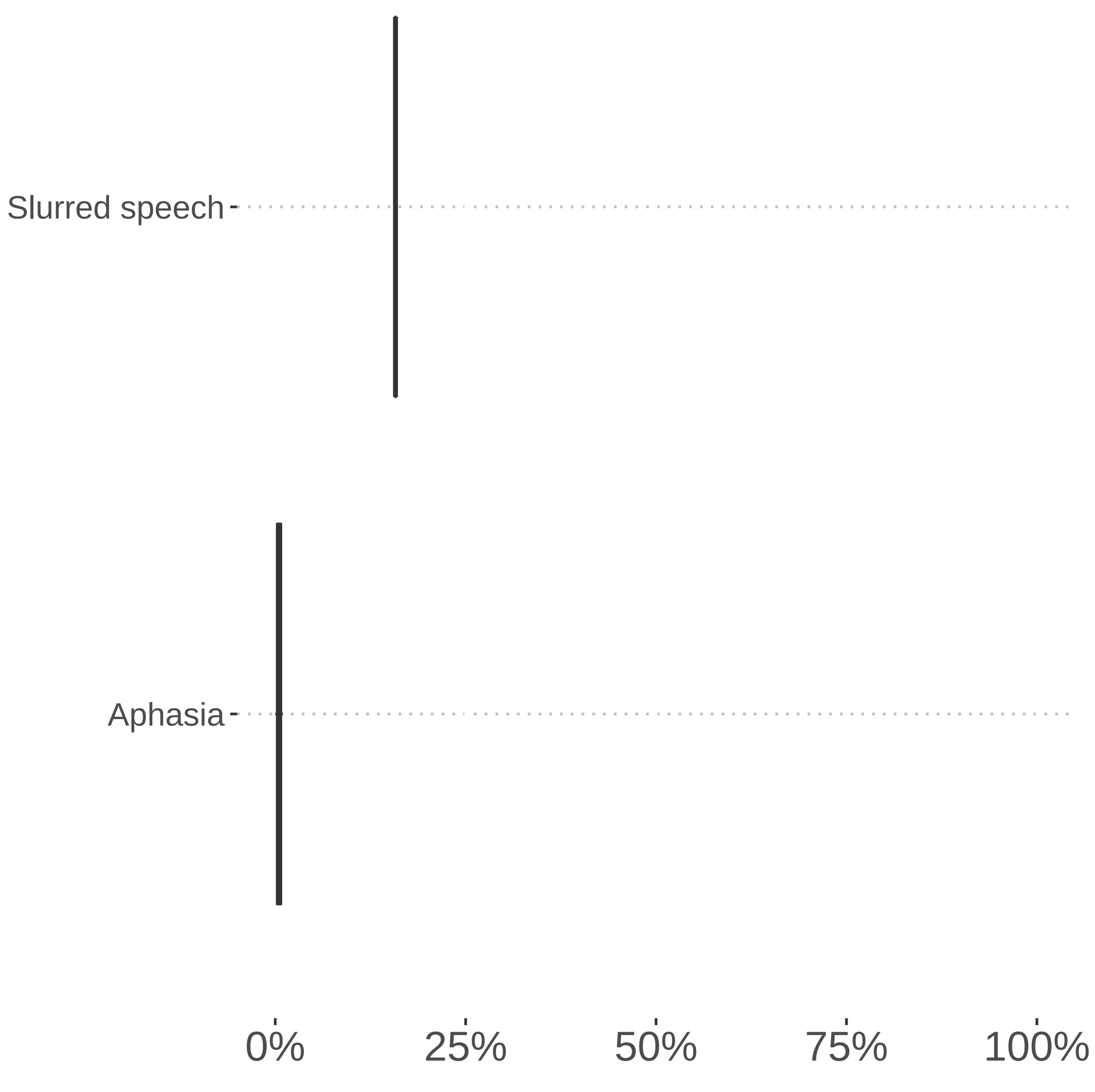


**Figure S21: Reported frequencies of speech and language abnormalities in persons with long COVID**. Anomic aphasia (HP:0030784, 1 study, median percentage: 46.3%), Aphasia (HP:0002381, 2 studies, median percentage: 0.5%), Bilingual aphasia (HP:0033849, 1 study, median percentage: 28.9%), Expressive aphasia (HP:0002427, 1 study, median percentage: 22.2%), Receptive aphasia (HP:0033848, 1 study, median percentage: 23.8%), Slurred speech (HP:0001350, 1 study, median percentage: 15.8%)

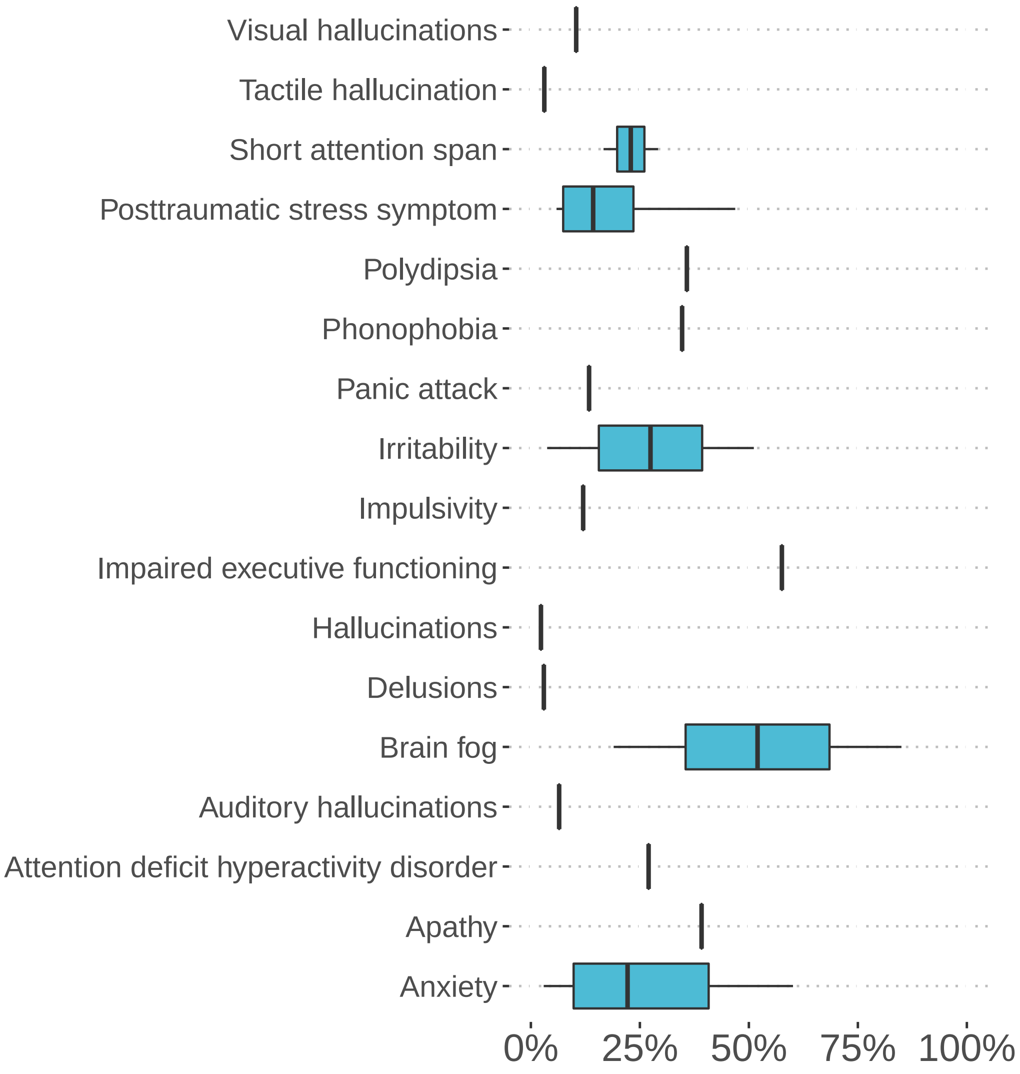


**Figure S22: Reported frequencies of behavioral abnormalities in persons with long COVID**. Anxiety (HP:0000739, 24 studies, median percentage: 22.2%), Apathy (HP:0000741, 1 study, median percentage: 39.2%), Attention deficit hyperactivity disorder (HP:0007018, 1 study, median percentage: 27.0%), Auditory hallucinations (HP:0008765, 1 study, median percentage: 6.5%), Brain fog (HP:0033630, 3 studies, median percentage: 85.0%), Delusions (HP:0000746, 1 study, median percentage: 3.0%), Hallucinations (HP:0000738, 1 study, median percentage: 2.3%), Impaired executive functioning (HP:0033051, 1 study, median percentage: 57.6%), Impulsivity (HP:0100710, 1 study, median percentage: 12.0%), Irritability (HP:0000737, 2 studies, median percentage: 27.4%), Panic attack (HP:0025269, 1 study, median percentage: 13.4%), Phonophobia (HP:0002183, 1 study, median percentage: 34.7%), Polydipsia (HP:0001959, 1 study, median percentage: 35.8%), Posttraumatic stress symptom (HP:0033676, 13 studies, median percentage: 14.3%), Short attention span (HP:0000736, 2 studies, median percentage: 22.9%), Tactile hallucination (HP:0033694, 1 study, median percentage: 3.1%), Visual hallucinations (HP:0002367, 1 study, median percentage: 10.4%)

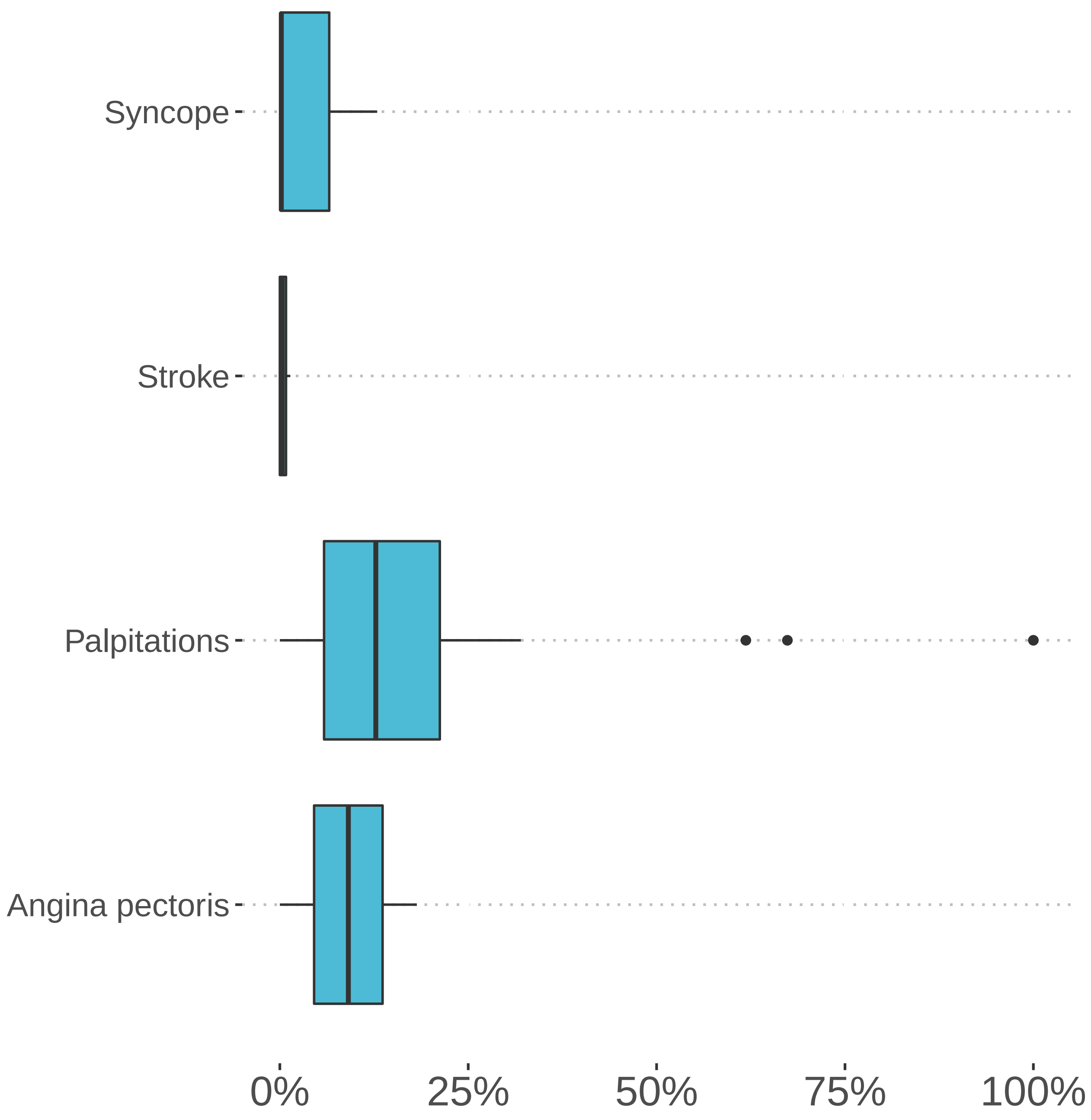


**Figure S23: Reported frequencies of cardiovascular symptoms in persons with long COVID**. Angina pectoris (HP:0001681, 2 studies, median percentage: 9.1%), Palpitations (HP:0001962, 18 studies, median percentage: 12.7%), Stroke (HP:0001297, 4 studies, median percentage: 0.3%), Syncope (HP:0001279, 3 studies, median percentage: 0.2%)

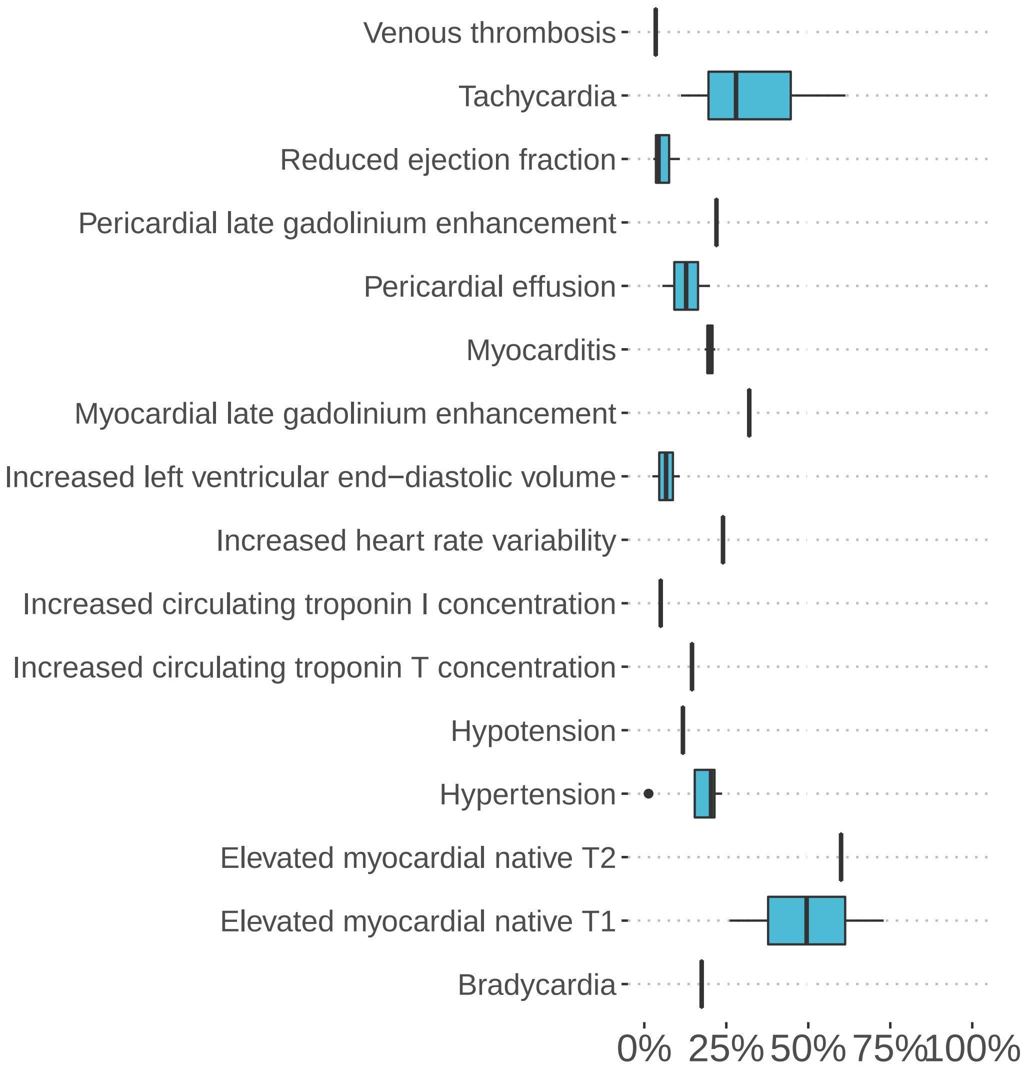


**Figure S24: Reported frequencies of cardiovascular findings in persons with long COVID**. Bradycardia (HP:0001662, 1 study, median percentage: 17.5%), Elevated myocardial native T1 (HP:4000006, 2 studies, median percentage: 49.5%), Elevated myocardial native T2 (HP:4000003, 1 study, median percentage: 60.0%), Hypertension (HP:0000822, 4 studies, median percentage: 20.4%), Hypotension (HP:0002615, 1 study, median percentage: 11.8%), Increased circulating troponin T concentration (HP:0410174, 1 study, median percentage: 14.5%), Increased circulating troponin I concentration (HP:0410173, 1 study, median percentage: 5.0%), Increased heart rate variability (HP:0031862, 1 study, median percentage: 24.0%), Increased left ventricular end-diastolic volume (HP:0033755, 2 studies, median percentage: 6.6%), Myocardial late gadolinium enhancement (HP:4000004, 1 study, median percentage: 32.0%), Myocarditis (HP:0012819, 2 studies, median percentage: 20.0%), Pericardial effusion (HP:0001698, 2 studies, median percentage: 12.8%), Pericardial late gadolinium enhancement (HP:4000005, 1 study, median percentage: 22.0%), Reduced ejection fraction (HP:0012664, 3 studies, median percentage: 4.3%), Tachycardia (HP:0001649, 3 studies, median percentage: 28.0%), Venous thrombosis (HP:0004936, 1 study, median percentage: 3.5%)

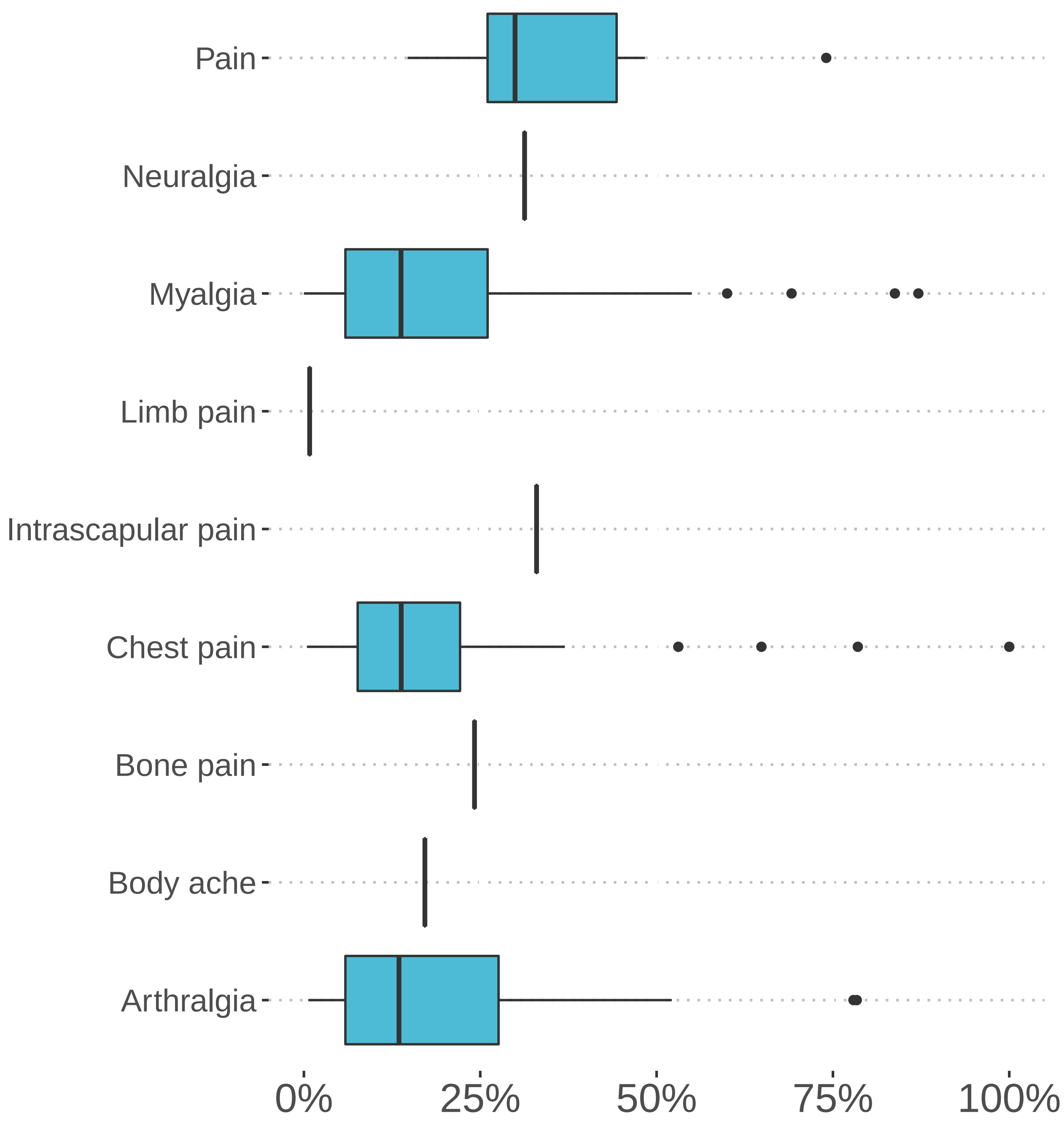


**Figure S25: Reported frequencies of pain in persons with long COVID**. Arthralgia (HP:0002829, 24 studies, median percentage: 13.5%), Body ache (HP:0033047, 1 study, median percentage: 17.2%), Bone pain (HP:0002653, 1 study, median percentage: 24.2%), Chest pain (HP:0100749, 32 studies, median percentage: 13.8%), Intrascapular pain (HP:0033746, 1 study, median percentage: 33.0%), Limb pain (HP:0009763, 1 study, median percentage: 0.8%), Myalgia (HP:0003326, 36 studies, median percentage: 13.8%), Neuralgia (HP:0033345, 1 study, median percentage: 31.3%), Pain (HP:0012531, 8 studies, median percentage: 29.9%)

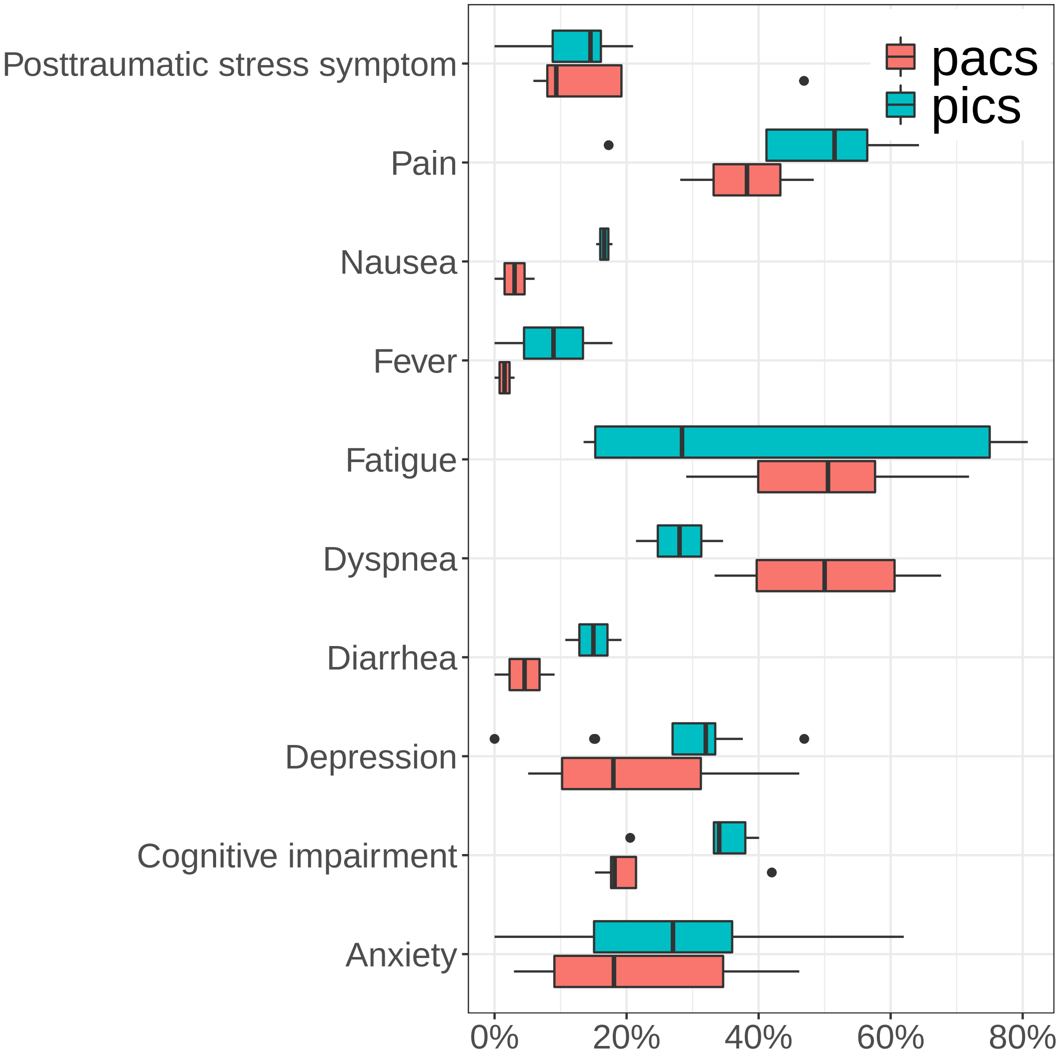


**Figure S26:** Frequencies of features reported in PASC following ICU admission in the acute phase compared to PICS. There is an overlap of the overall experienced symptoms between PASC and PICS, such as Post-traumatic stress and Anxiety, whereas unique to PASC, individuals reported Dyspnea (50%) nearly twice as frequently as the general PICS populations (28%). While reported in both groups, those in the PICS population report more substantial Pain (55%), Nausea (18%), Fever (9%), Diarrhea (18%), Depression (32%) and Cognitive impairment (35%). Fatigue was overwhelmingly reported in those with PICS, though of note, median reports of fatigue in PASC cohorts is over 50% of those who suffer. Dyspnea and fatigue are the most commonly reported symptoms in the PASC cohort. In light of the heterogeneity of study designs, we did not perform formal statistical testing for difference in mean frequencies for the two groups.

| Category | Count |
|---|---|
| longcovid cohort counts | 48 |
| long covid cohort other | 91 |
| acute COVID-19 | 9 |
| PICS | 1 |
| SARS1 | 10 |
| research | 3 |
| review | 67 |
| no pmid | 74 |

**Table S1:** Papers considered for inclusion for curation. We searched for publications on long COVID using CoronaCentral, which uses machine learning to process the literature on SARS-CoV-2.TODO We retrieved 303 articles predicted to be relevant to long COVID on April 29th, 2021. We included papers about long COVID that reported counts of individual features in groups of individuals following acute COVID-19 infection (`longcovid cohort counts`). We eexcluded papers about long COVID that reported other clinical aspects (`long covid cohort other`), papers that primarily studies acute COVID-19 (`acute`), post ICU syndrome (`PICS`), sequellae following infection with SARS (`SARS1`), papers with molecualr research about acute COVID-19 or long COVID (`research`), review papers (`review`), and manuscripts that were not listed in PubMed (`no pmid`). In addition, ten publications with PumMed identifiers were identified by manual searches [1, 2, 3, 4, 5, 6, 7, 8, 9, 10] and one preprint was chosen from medRxiv [11].

**Table S2:** Post acute COVID studies included in this analysis.

| Author, Year | Cohort type | Age | m/N | Study Type | Days |
|---|---|---|---|---|---|
| Carvalho-Schneider (2021) [4] | mixed in/out-patient (non-ICU) | 49.0y | 66/150 | Telephone questionnaire | 60.0 d |
| Arnold (2020) [12] | ICU | 62.0y | 11/18 | Clinical exam | 84.0 d |
| Tenforde (2020) [8] | out-patient | 42.5y | 120/264 | Telephone questionnaire | 17.5 d |
| Carvalho-Schneider (2021) [4] | mixed in/out-patient (non-ICU) | 49.0y | 66/150 | Telephone questionnaire | 30.0 d |
| Petersen (2020) [13] | out-patient | 39.9y | 82/180 | Telephone questionnaire | 99.0 d |
| Garg (2021) [14] | out-patient | 37.0y | 10/19 | Clinical exam | 47.0 d |
| Zhao (2020) [9] | in-patient | 47.7y | 32/55 | Clinical exam | 78.5 d (h) |
| Taboada (2021) [15] | ICU | 65.5y | 59/91 | Clinical exam | 180.0 d (h) |
| Garrigues (2020) [16] | in-patient (non-ICU) | 64.1y | 56/96 | Telephone questionnaire | 110.9 d |
| Mandal (2020) [17] | in-patient | 59.9y | 238/384 | Telephone clinical mixed | 54.0 d (h) |
| Weerahandi (2021) [18] | in-patient | 62.0y | 95/152 | Telephone online questionnaire | 37.0 d (h) |
| Goertz (2020) [19] | mixed in/out-patient (non-ICU) | 47.0y | 310/2113 | Written questionnaire | 79.0 d |
| Venturelli (2021) [20] | mixed in/out-patient | 63.0y | 515/767 | Clinical exam | 81.0 d (h) |
| Halpin (2021) [21] | ICU | 58.5y | 19/32 | Telephone questionnaire | 48.0 d (h) |
| Trinkmann (2021) [22] | mixed in/out-patient | 48.0y | 108/246 | Clinical exam | 68.0 d |

*Continued on next page*

Table **S2** – *Continued from previous page*

| **Author, Year** | **Cohort type** | **Age** | **m/N** | **Study Type** | **Days** |
|---|---|---|---|---|---|
| Halpin (2021) [21] | in-patient (non-ICU) | 70.5y | 35/68 | Telephone questionnaire | 48.0 d (h) |
| Ramani (2021) [23] | in-patient | 55.0y | 34/58 | Clinical exam | 48.0 d (h) |
| Carfi (2020) [24] | in-patient | 56.5y | 90/153 | Clinical exam | 60.3 d |
| Rosales-Castillo (2021) [25] | in-patient | 60.2y | 66/118 | Clinical exam | 50.8 d (h) |
| Townsend (2020) [26] | mixed in/out-patient | 49.5y | 59/128 | Clinical exam | 72.0 d |
| de Graaf (2021) [27] | in-patient (non-ICU) | 60.8y | 29/47 | Clinical exam | 42.0 d (h) |
| Ludvigsson (2021) [1] | out-patient | 12.0y | 1/5 | Telephone questionnaire | 210.0 d |
| Moreno-Perez (2021) [28] | mixed in/out-patient | 56.0y | 146/277 | Clinical exam | 77.0 d |
| Chopra (2020) [10] | in-patient | 62.0y | 253/488 | Telephone questionnaire | 60.0 d (h) |
| Huang (2021) [29] | in-patient | 57.0y | 897/1733 | Clinical exam | 186.0 d |
| van den Borst (2020) [30] | ICU | 60.0y | 35/46 | Clinical exam | 91.0 d (h) |
| Havervall (2021) [5] | out-patient | 43.0y | 55/323 | Written questionnaire | 240.0 d |
| Havervall (2021) [5] | out-patient | 43.0y | 55/323 | Written questionnaire | 120.0 d |
| Havervall (2021) [5] | out-patient | 43.0y | 55/323 | Written questionnaire | 60.0 d |
| van den Borst (2020) [30] | out-patient | 52.0y | 8/27 | Clinical exam | 91.0 d (h) |
| van den Borst (2020) [30] | in-patient (non-ICU) | 61.0y | 31/51 | Clinical exam | 91.0 d (h) |
| Arnold (2020) [12] | in-patient (non-ICU) | 57.0y | 44/65 | Clinical exam | 84.0 d |
| Arnold (2020) [12] | in-patient (non-ICU) | 47.0y | 13/27 | Clinical exam | 84.0 d |

*Continued on next page*

Table **S2** – *Continued from previous page*

| **Author, Year** | **Cohort type** | **Age** | **m/N** | **Study Type** | **Days** |
|---|---|---|---|---|---|
| Zhang (2021) [31] | in-patient | 42.5y | 294/527 | Clinical exam | 180.0 d (h) |
| Zhang (2021) [31] | in-patient | 42.5y | 294/527 | Clinical exam | 90.0 d (h) |
| Rass (2021) [32] | out-patient | 45.0y | 10/32 | Clinical exam | 90.0 d (h) |
| Munro (2020) [33] | in-patient | 64.0y | 120/138 | Telephone questionnaire | 56.0 d (h) |
| Ramani (2021) [34] | ICU | 56.0y | 17/28 | Clinical exam | 39.5 d (h) |
| Wong (2020) [35] | in-patient | 62.0y | 50/78 | Clinical exam | 91.0 d |
| Akter (2020) [36] | in-patient | 40.0y | 558/734 | Telephone questionnaire | 28.0 d |
| Daher (2020) [37] | ICU | 64.0y | 22/33 | Clinical exam | 56.0 d (h) |
| Stavem (2020) [38] | out-patient | 49.7y | 198/451 | Written questionnaire | 117.0 d |
| Jacobs (2020) [39] | in-patient (non-ICU) | 57.0y | 112/183 | Telephone email questionnaire | 35.0 d (h) |
| Writing Committee (2021) [40] | in-patient (non-ICU) | 60.9y | 76/126 | Telephone clinical mixed | 121.0 d (h) |
| Writing Committee (2021) [40] | ICU | 60.9y | 43/73 | Telephone clinical mixed | 93.0 d (h) |
| González (2021) [41] | ICU | 60.0y | 46/62 | Clinical exam | 90.0 d (h) |
| Iqbal (2021) [42] | mixed in/out-patient | 32.1y | 71/158 | Telephone questionnaire | 38.1 d (h) |
| Prieto (2021) [43] | mixed in/out-patient | 43.0y | 38/85 | Clinical exam | 53.0 d |
| Cheng (2021) [2] | mixed in/out-patient | 73.0y | 76/139 | Telephone clinical mixed | 63.0 d |
| Graham (2021) [44] | out-patient | 43.2y | 17/50 | Telephone clinical mixed | 141.0 d |

*Continued on next page*

Table **S2** – *Continued from previous page*

| **Author, Year** | **Cohort type** | **Age** | **m/N** | **Study Type** | **Days** |
|---|---|---|---|---|---|
| Dennis (2021) [45] | mixed in/out-patient (non-ICU) | 44.0y | 49/201 | Clinical exam | 141.0 d |
| Ordinola Navarro (2021) [46] | mixed in/out-patient | 58.0y | 50/115 | Clinical exam | 40.0 d |
| Bellan (2021) [6] | in-patient | 61.0y | 142/238 | Telephone questionnaire | 105.0 d (h) |
| Puntmann (2020) [47] | mixed in/out-patient | 49.0y | 53/100 | Clinical exam | 71.0 d |
| Dennis (2021) [45] | out-patient | 43.0y | 45/163 | Clinical exam | 141.0 d |
| Sonnweber (2020) [48] | mixed in/out-patient | 57.0y | 82/145 | Clinical exam | 103.0 d |
| Sonnweber (2020) [48] | mixed in/out-patient | 57.0y | 82/145 | Clinical exam | 63.0 d |
| Dennis (2021) [45] | in-patient | 50.0y | 14/37 | Clinical exam | 138.0 d |
| Jacobson (2021) [49] | in-patient | 50.6y | 14/22 | Clinical exam | 119.3 d |
| Zhang (2021) [31] | in-patient | 42.5y | 294/527 | Clinical exam | 28.0 d (h) |
| Rass (2021) [32] | in-patient (non-ICU) | 61.0y | 48/72 | Clinical exam | 90.0 d (h) |
| Rass (2021) [32] | ICU | 58.0y | 24/31 | Clinical exam | 90.0 d (h) |
| Jacobson (2021) [49] | out-patient | 41.6y | 49/96 | Clinical exam | 119.3 d |
| Chun (2021) [50] | mixed in/out-patient | 53.0y | 34/61 | Clinical exam | 63.0 d |
| Baricich (2021) [51] | in-patient | 57.9y | 123/204 | Clinical exam | 124.7 d (h) |
| Townsend (2021) [52] | mixed in/out-patient | 47.3y | 65/150 | Clinical exam | 80.5 d |
| Garrigues (2020) [16] | ICU | 59.6y | 19/24 | Telephone questionnaire | 110.9 d |
| Liu (2020) [3] | in-patient | 46.6y | 21/51 | Clinical exam | 28.0 d (h) |
| Suárez-Robles (2020) [53] | in-patient | 58.5y | 62/134 | Telephone questionnaire | 90.0 d (h) |

*Continued on next page*

Table **S2** – *Continued from previous page*

| Author, Year | Cohort type | Age | m/N | Study Type | Days |
|---|---|---|---|---|---|
| Bellan (2021) [6] | in-patient | 61.0y | 142/238 | Clinical exam | 105.0 d (h) |
| Darley (2021) [54] | mixed in/out-patient | 47.0y | 51/78 | Clinical exam | 69.0 d |
| Liang (2020) [55] | in-patient (non-ICU) | 41.3y | 21/76 | Clinical exam | 90.0 d (h) |
| Xiong (2021) [7] | in-patient | 52.0y | 245/538 | Telephone questionnaire | 97.0 d (h) |
| Blanco (2021) [56] | in-patient | 55.0y | 64/100 | Clinical exam | 104.0 d |
| Logue (2021) [57] | in-patient | 54.0y | 8/16 | Written questionnaire | 169.0 d |
| Logue (2021) [57] | out-patient | 46.3y | 68/161 | Written questionnaire | 169.0 d |
| Davis2020 | mixed in/out-patient | 45.0y | 793/3762 | Online questionnaire | 114.5 d |
| Abdallah (2021) [58] | in-patient | 59.1y | 16/25 | Clinical exam | 102.3 d (h) |
| de Graaf (2021) [27] | ICU | 60.8y | 22/34 | Clinical exam | 42.0 d (h) |
| De Lorenzo (2020) [59] | in-patient | 61.0y | 92/126 | Clinical exam | 21.5 d (h) |
| De Lorenzo (2020) [59] | out-patient | 50.0y | 31/59 | Clinical exam | 26.0 d |
| Abdallah (2021) [58] | out-patient | 42.4y | 20/38 | Clinical exam | 129.8 d |

**Table S3:** Post-ICU syndrome (PICS) phenotypic features curated from studies analyzed in this work. **days**: Number of days following discharge from ICU

| HPO label | HPO id | PMID | author (year) | days | counts | perc. |
|---|---|---|---|---|---|---|
| Diarrhea | HP:0002014 | 23856099 | Choi (2013) | 120 | 5/26 | 19.2% |
| Diarrhea | HP:0002014 | 23856099 | Choi (2013) | 60 | 3/28 | 10.7% |
| Memory impairment | HP:0002354 | 33593406 | Kawakami (2021) | 180 | 36/61 | 59.0% |
| Sleep disturbance | HP:0002360 | 29243344 | Langerud (2018) | 360 | 40/89 | 44.9% |
| Sleep disturbance | HP:0002360 | 29243344 | Langerud (2018) | 90 | 58/118 | 49.2% |
| Sleep disturbance | HP:0002360 | 23856099 | Choi (2013) | 120 | 12/26 | 46.2% |
| Sleep disturbance | HP:0002360 | 23856099 | Choi (2013) | 60 | 18/28 | 64.3% |
| Poor appetite | HP:0004396 | 23856099 | Choi (2013) | 120 | 6/26 | 23.1% |
| Poor appetite | HP:0004396 | 33625631 | Miyamoto (2021) | 90 | 6/81 | 7.4% |
| Poor appetite | HP:0004396 | 23856099 | Choi (2013) | 60 | 8/28 | 28.6% |
| Dyspnea | HP:0002094 | 23856099 | Choi (2013) | 120 | 9/26 | 34.6% |
| Dyspnea | HP:0002094 | 23856099 | Choi (2013) | 60 | 6/28 | 21.4% |
| Diminished physical functioning | HP:0033666 | 24815803 | Jackson (2014) | 360 | 102/374 | 27.3% |
| Diminished physical functioning | HP:0033666 | 32995017 | Shima (2020) | 90 | 37/117 | 31.6% |
| Diminished physical functioning | HP:0033666 | 29787415 | Marra (2018) | 90 | 100/383 | 26.1% |
| Diminished physical functioning | HP:0033666 | 33593406 | Kawakami (2021) | 180 | 31/61 | 50.8% |
| Diminished physical functioning | HP:0033666 | 29787415 | Marra (2018) | 360 | 69/332 | 20.8% |
| Diminished physical functioning | HP:0033666 | 24815803 | Jackson (2014) | 90 | 139/428 | 32.5% |
| Diminished physical functioning | HP:0033666 | 32995017 | Shima (2020) | 360 | 15/74 | 20.3% |
| Posttraumatic stress symptom | HP:0033676 | 24815803 | Jackson (2014) | 360 | 24/361 | 6.6% |
| Posttraumatic stress symptom | HP:0033676 | 32398018 | Tripathy (2020) | 180 | 0/322 | 0.0% |
| Posttraumatic stress symptom | HP:0033676 | 30466485 | Hatch (2018) | 90 | 504/3151 | 16.0% |
| Posttraumatic stress symptom | HP:0033676 | 32995017 | Shima (2020) | 90 | 17/117 | 14.5% |
| Posttraumatic stress symptom | HP:0033676 | 29243344 | Langerud (2018) | 360 | 13/89 | 14.6% |
| Posttraumatic stress symptom | HP:0033676 | 29243344 | Langerud (2018) | 90 | 15/118 | 12.7% |
| Posttraumatic stress symptom | HP:0033676 | 24815803 | Jackson (2014) | 90 | 27/415 | 6.5% |

*Continued on next page*

Table **S3** – *Continued from previous page*

| **HPO label** | **HPO id** | **PMID** | **author (year)** | **days** | **counts** | **perc.** |
|---|---|---|---|---|---|---|
| Posttraumatic stress symptom | HP:0033676 | 32643362 | Anon (2020) | 90 | 8/73 | 11.0% |
| Posttraumatic stress symptom | HP:0033676 | 33625631 | Miyamoto (2021) | 90 | 17/81 | 21.0% |
| Posttraumatic stress symptom | HP:0033676 | 30466485 | Hatch (2018) | 360 | 567/3151 | 18.0% |
| Posttraumatic stress symptom | HP:0033676 | 32995017 | Shima (2020) | 360 | 12/74 | 16.2% |
| Fatigue | HP:0012378 | 29243344 | Langerud (2018) | 360 | 12/89 | 13.5% |
| Fatigue | HP:0012378 | 29243344 | Langerud (2018) | 90 | 18/118 | 15.3% |
| Fatigue | HP:0012378 | 23856099 | Choi (2013) | 120 | 21/26 | 80.8% |
| Fatigue | HP:0012378 | 33625631 | Miyamoto (2021) | 90 | 23/81 | 28.4% |
| Fatigue | HP:0012378 | 23856099 | Choi (2013) | 60 | 21/28 | 75.0% |
| Diminished mental health | HP:0033667 | 33593406 | Kawakami (2021) | 180 | 14/61 | 23.0% |
| Restrictive ventilatory defect | HP:0002091 | 32643362 | Anon (2020) | 90 | 18/73 | 24.7% |
| Cognitive impairment | HP:0100543 | 29787415 | Marra (2018) | 90 | 128/337 | 38.0% |
| Cognitive impairment | HP:0100543 | 24088092 | Pandharipande (2013) | 90 | 228/569 | 40.1% |
| Cognitive impairment | HP:0100543 | 29787415 | Marra (2018) | 360 | 97/292 | 33.2% |
| Cognitive impairment | HP:0100543 | 32643362 | Anon (2020) | 90 | 15/73 | 20.5% |
| Cognitive impairment | HP:0100543 | 24088092 | Pandharipande (2013) | 360 | 130/382 | 34.0% |
| Insomnia | HP:0100785 | 33625631 | Miyamoto (2021) | 90 | 9/81 | 11.1% |
| Pain | HP:0012531 | 29243344 | Langerud (2018) | 90 | 58/118 | 49.2% |
| Pain | HP:0012531 | 23856099 | Choi (2013) | 120 | 14/26 | 53.8% |
| Pain | HP:0012531 | 33625631 | Miyamoto (2021) | 90 | 14/81 | 17.3% |
| Pain | HP:0012531 | 23856099 | Choi (2013) | 60 | 18/28 | 64.3% |
| Depression | HP:0000716 | 24815803 | Jackson (2014) | 360 | 116/347 | 33.4% |
| Depression | HP:0000716 | 32398018 | Tripathy (2020) | 180 | 0/180 | 0.0% |
| Depression | HP:0000716 | 30466485 | Hatch (2018) | 90 | 1029/3319 | 31.0% |
| Depression | HP:0000716 | 32995017 | Shima (2020) | 90 | 44/117 | 37.6% |
| Depression | HP:0000716 | 29243344 | Langerud (2018) | 360 | 24/89 | 27.0% |
| Depression | HP:0000716 | 29243344 | Langerud (2018) | 90 | 18/118 | 15.3% |
| Depression | HP:0000716 | 29787415 | Marra (2018) | 90 | 121/363 | 33.3% |
| Depression | HP:0000716 | 29787415 | Marra (2018) | 360 | 97/313 | 31.0% |
| Depression | HP:0000716 | 24815803 | Jackson (2014) | 90 | 149/407 | 36.6% |
| Depression | HP:0000716 | 32643362 | Anon (2020) | 90 | 11/73 | 15.1% |
| Depression | HP:0000716 | 33625631 | Miyamoto (2021) | 90 | 38/81 | 46.9% |
| Depression | HP:0000716 | 30466485 | Hatch (2018) | 360 | 1062/3319 | 32.0% |

*Continued on next page*

Table **S3** – *Continued from previous page*

| **HPO label** | **HPO id** | **PMID** | **author (year)** | **days** | **counts** | **perc.** |
|---|---|---|---|---|---|---|
| Depression | HP:0000716 | 32995017 | Shima (2020) | 360 | 24/74 | 32.4% |
| Anxiety | HP:0000739 | 32398018 | Tripathy (2020) | 180 | 0/180 | 0.0% |
| Anxiety | HP:0000739 | 30466485 | Hatch (2018) | 90 | 1192/3312 | 36.0% |
| Anxiety | HP:0000739 | 32995017 | Shima (2020) | 90 | 37/117 | 31.6% |
| Anxiety | HP:0000739 | 29243344 | Langerud (2018) | 360 | 11/89 | 12.4% |
| Anxiety | HP:0000739 | 29243344 | Langerud (2018) | 90 | 28/118 | 23.7% |
| Anxiety | HP:0000739 | 32643362 | Anon (2020) | 90 | 11/73 | 15.1% |
| Anxiety | HP:0000739 | 33625631 | Miyamoto (2021) | 90 | 33/81 | 40.7% |
| Anxiety | HP:0000739 | 30466485 | Hatch (2018) | 360 | 2053/3312 | 62.0% |
| Anxiety | HP:0000739 | 32995017 | Shima (2020) | 360 | 20/74 | 27.0% |
| Fever | HP:0001945 | 23856099 | Choi (2013) | 120 | 0/26 | 0.0% |
| Fever | HP:0001945 | 23856099 | Choi (2013) | 60 | 5/28 | 17.9% |
| Muscle weakness | HP:0001324 | 23856099 | Choi (2013) | 120 | 16/26 | 61.5% |
| Muscle weakness | HP:0001324 | 32643362 | Anon (2020) | 90 | 28/73 | 38.4% |
| Muscle weakness | HP:0001324 | 33625631 | Miyamoto (2021) | 90 | 31/81 | 38.3% |
| Muscle weakness | HP:0001324 | 23856099 | Choi (2013) | 60 | 18/28 | 64.3% |
| Nausea | HP:0002018 | 23856099 | Choi (2013) | 120 | 4/26 | 15.4% |
| Nausea | HP:0002018 | 23856099 | Choi (2013) | 60 | 5/28 | 17.9% |
| Constipation | HP:0002019 | 23856099 | Choi (2013) | 120 | 4/26 | 15.4% |
| Constipation | HP:0002019 | 23856099 | Choi (2013) | 60 | 2/28 | 7.1% |

### Supplemental Note 1

This note provides background information and comments about selected phenotypic features mentioned in the main manuscript.

General (constitutional) symptoms were commonly reported. The most commonly reported symptom was *Fatigue* (HP:0012378). Other symptoms reported in 10 or more cohorts included *Abdominal pain* (HP:0002027), *Arthralgia* (HP:0002829), *Chest pain* (HP:0100749), *Difficulty walking* (HP:0002355), *Myalgia* (HP:0003326), and *Pharyngalgia* (HP:0033050) (See **Figure 2** in the main manuscript and **Figure S25**).

Abdominal complaints such as *Nausea* (HP:0002018), *Vomiting* (HP:0002013), *Diarrhea* (HP:0002014), and *Poor appetite* (HP:0004396) are commonly reported in persons with long COVID (**Figure S5**). *Hepatic steatosis* (HP:0001397) and *Pancreatic steatosis* (HP:0033757), *Hepatomegaly* (HP:0002240), *Splenomegaly* (HP:0001744), and *Hepatitis* (HP:0012115) were reported in one publication that described two cohorts, whereby the frequency of these features was significantly higher than in healthy controls [45]. However, given that these features are relatively common in the general population [60], additional research will be required to assess the extent of increased risk exists for persons with long COVID (**Figure S6**).

Persistent lung abnormalities were the most common long-term sequelae of Severe Acute Respiratory syndrome (SARS) and Middle East Respiratory Syndrome (MERS), epidemic viral illness related to betacoronavirus infection [61]. Respiratory abnormalities are also very common in long COVID. Fourteen imaging abnormalities ascertained by computed tomography or plain-film radiography were reported (**Figure S2**). Thirteen abnormalities ascertained by pulmonary function testing (PFT) were reported, often related to restrictive ventilatory defects and reduced diffusion capacity (**Figure S3**). The most commonly reported symptom was *Dyspnea* (HP:0002094), whose occurrence and severity correlate with defects in PFT [55].

Cardiovascular symptoms were relatively common, including *Angina pectoris* (HP:0001681), *Palpitations* (HP:0001962), and *Syncope* (HP:0001279) (**Figure S23**). A small number of cohorts documented abnormal echocardiographic and cardiac magnetic resonance imaging findings as well as heart rhythm abnormalities (**Figure S24**).

*Acute kidney injury* (HP:0001919) is a common complication of COVID-19. AKI is a risk factor for the development of chronic kidney disease (CKD) [62]. In a recently published study from Sweden, among a group of 60 ICU patients admitted for COVID-19 infection, inpatient AKI severity was associated with higher CKD stages at 3- to 6-month follow up [63]. Another study from the USA recently reported that AKI in the setting of COVID-19, was associated with a greater rate of eGFR decrease after discharge compared with AKI without COVID-19.26 Another study reported decreased glomerular filtration rate in 35% of patients six months following discharge for acute COVID-19 [29]. However, a recent study suggested that most patients with AKI in the setting of COVID-19 will experience long-term renal recovery [64]. More studies with longer follow-up will help our understanding on the long-term renal consequences of COVID-19 (**Figure S10**).

*Blurred vision* (HP:0000622) was the most commonly reported ocular finding, affecting 15% of long COVID patients. Other findings, such as *Photophobia* (HP:0000613), *Ocular pain* (HP:0200026), and *Vitreous floaters* (HP:0100832), were reported in one or two cohorts each (**Figure S12**), but further work will be required to define the natural history of ocular complaints in long COVID. *Vertigo* (HP:0002321; dizziness) was reported in 25.5% of participants of 10 cohorts. *Ear pain* (HP:0030766) and *Tinnitus* (HP:0000360) were also common (**Figure S11**).

The largest number of distinct symptoms and findings were reported for the nervous system, which could be associated with both peripheral or central nervous system dysfunction. Symptoms related to *Cognitive impairment* (HP:0100543) and *Tremor* (HP:0001337), as well as *Paresthesia* (HP:0003401) and related findings, were all commonly reported (**Figures S15** and **S14**) . Symptoms included reduced or abolished sense of taste and smell (**Figure S17**), *Diminished ability to concentrate* (HP:0031987), *Bradyphrenia* (HP:0031843), and *Headache* (HP:0002315) (**Figures S15** and **S20**). .

Many cases of COVID-19 are marked by short- and long-term psychiatric symptoms. While it has been suggested that these symptoms occur through viral invasion of the brain, without definitive evidence, it seems more likely that the psychiatric effects of COVID-19 infection are due to a combination of neuroinflammation and microvascular injury [65]. Evidence exists for an increased incidence of new-onset mental illness in patients with prior COVID-19 infection compared to controls with a comparable health event [66]. Apart from the direct effects of COVID-19 infection, mental health is sensitive to societal events such as the restrictions imposed in many countries in response to the COVID-19 pandemic, and it has been reported that the overall prevalence of depression symptoms in the US increased more than 3-fold during the COVID-19 pandemic [67]. Additionally, some anxiety and mood symptoms may be due to testing issues resulting in inadequate care: one study found that anxiety and mood symptoms were more common in those without lab confirmation of COVID, and that those without lab confirmation experienced an 8-week delay in care [68]. Another showed that those without lab confirmation were less likely to have adequate rest, less likely to have had time off, more likely to experience loss of income, and more likely to be unable to live alone at 6 weeks [69]. Given the myriad of societal changes, psychiatric symptoms of long COVID must be considered in light of both biological and social factors affecting disease. In the cohorts analyzed in our study, anxiety and depression were reported in 22 and 23 cohorts respectively (**Figure 2** in the main manuscript), but a wide range of other complaints including Depression (HP:0000716), *Anxiety* (HP:0000739), *Posttraumatic stress symptom* (HP:0033676), and *Irritability* (HP:0000737) were also reported (**Figures S19** and **S22**).

Sleep-related symptoms were commonly reported in multiple studies. The term *Sleep disturbance* (HP:0002360) was identified in ten cohorts. Sleep disturbances were reported by 24.0% of patients in these cohorts. More specific sleep related features (i.e., child terms of Sleep Disturbance) were also reported. *Insomnia* (HP:0100785) was the most prevalent feature, followed by *Restless legs* (HP:0012452) and *Sleep apnea* (HP:0010535). Specific types of insomnia, as represented by child terms *Sleep onset insomnia* (HP:0031354), *Maintenance insomnia* (HP:0031355), and *Terminal insomnia* (HP:0031356) were also reported in one cohort by 39.6%, 47.6% and 24.9% of patients, respectively (**Figure S18**). Because sleep and circadian disruptions may affect multiple organ systems, the impact of these disruptions should be considered when investigating other domains such as respiratory, neurological, and behavioral function. *Fatigue* (HP:0012378), an inexact term characterized by extreme tiredness and impaired daytime function, was reported by 70.2% of patients across 34 studies. Patients reporting fatigue represented a very heterogeneous group including those with mild to severe disease, and seen in both those hospitalized and not hospitalized. While fatigue is commonly associated with sleep disturbance [70], this relationship is not fully characterized but is believed to be mediated by inflammatory molecules such interleukins 1 and -6, and neuropeptides such as hypocretin-1. Acutely, fatigue may be a compensatory mechanism to drive increased sleep responses after immune challenges, but long-term may represent a maladaptive response [71].

*Brain fog* (HP:0033630), another term commonly used by patients with sleep problems to describe decreased cognitive ability [72], was reported in 44.6% of patients across two studies. The relationship between brain fog, sleep, and fatigue is poorly understood but the high number of patients reporting on this in these two studies suggests it should be included in future investigations of sleep fatigue, and cognitive dysfunction association with long COVID. Finally, several other neurological and psychiatric features observed in long COVID patients and highlighted by this review could be attributed to changes in sleep and circadian rhythms observed in these patients during and after COVID infection, including *Diminished ability to concentrate* (HP:0031987), *Short term memory impairment* (HP:0033687), *Irri-*

*tability* (HP:0000737), *Emotional lability* (HP:0000712), *Memory impairment* (HP:0002354), *Headache* (HP:0002315), *Depression* (HP:0000716), *Anxiety* (HP:0000739), *Cognitive impairment* (HP:0100543), *Pain* (HP:0012531), and *Apathy* (HP:0000741). Dissecting the directionality of sleep disturbances and symptoms related to other systems is fundamental to develop approaches that might positively impact the quality of life in long COVID patients.

### Supplemental Note 2: Post ICU Syndrome (PICS) vs. long COVID

In addition to identifying studies for long COVID, we also curated 18 other studies (Supplemental Table **S3**), which directly discussed post-Intensive Care Unit (ICU) care and recovery with 13,358 patients, annotated to 19 HPO terms. While long COVID has been identified as a novel consequence of COVID-19, there is a direct need to understand long COVID as it pertains specifically to critically ill individuals that were hospitalized in the ICU - a unique population. In addition to understanding long COVID, one must consider the phenomenon of Post-Intensive Care Syndrome (PICS), which is an all-encompassing term for the myriad of symptoms experienced by those who survive critical illness and their families, lasting months to years in duration [**?**]. As the focus shifts from survival to survivorship, healthcare providers and researchers must consider the lasting effects of the ICU environment and its associated therapies on individuals who concurrently are diagnosed with COVID-19. While life-preserving, many of these therapies come with consequences seen in PICS – physical, cognitive, and emotional. Examples may include, but are not limited to, ICU-acquired weakness propagated by prolonged immobilization, neuromuscular blockade, sedation, and/or corticosteroids [**?**]. Many of the symptoms experienced by ICU survivors are shared with those who are critically ill COVID-19 survivors who required ICU admission, but also those who did not require ICU [**?**]. This further elucidates the point that both long COVID and PICS are unique and distinct phenomena, which have shared symptomatology in the presence of critical illness. Early recognition and intervention are necessary to offset these untoward long-term side effects.

[30] B. van den Borst, J. B. Peters, M. Brink, Y. Schoon, C. P. Bleeker-Rovers, H. Schers, H. W. H. van Hees, H. van Helvoort, M. van den Boogaard, H. van der Hoeven, M. H. Reijers, M. Prokop, J. Vercoulen, and M. van den Heuvel. Comprehensive health assessment three months after recovery from acute COVID-19. *Clin Infect Dis*, Nov 2020.

[31] J. Zhang, J. Xu, S. Zhou, C. Wang, X. Wang, W. Zhang, K. Ning, Y. Pan, T. Liu, J. Zhao, and L. Dong. The characteristics of 527 discharged COVID-19 patients undergoing long-term follow-up in China. *Int J Infect Dis*, 104:685–692, Mar 2021.

[32] V. Rass, R. Beer, A. J. Schiefecker, M. Kofler, A. Lindner, P. Mahlknecht, B. Heim, V. Limmert, S. Sahanic, A. Pizzini, T. Sonnweber, I. Tancevski, C. Scherfler, L. Zamarian, R. Bellmann-Weiler, G. Weiss, A. Djamshidian, S. Kiechl, K. Seppi, J. Loeffler-Ragg, B. Pfausler, and R. Helbok. Neurological outcome and quality of life 3 months after COVID-19: A prospective observational cohort study. *Eur J Neurol*, Mar 2021.

[41] J. González, I. D. Benítez, P. Carmona, S. Santisteve, A. Monge, A. Moncusí-Moix, C. Gort-Paniello, L. Pinilla, A. Carratalá, M. Zuil, R. Ferrer, A. Ceccato, L. Fernández, A. Motos, J. Riera, R. Menéndez, D. Garcia-Gasulla, O. Peñuelas, J. F. Bermejo-Martin, G. Labarca, J. Caballero, G. Torres, D. de Gonzalo-Calvo, A. Torres, and F. Barbé. Pulmonary Function and Radiologic Features in Survivors of Critical COVID-19: A 3-Month Prospective Cohort. *Chest*, Mar 2021.

[48] T. Sonnweber, S. Sahanic, A. Pizzini, A. Luger, C. Schwabl, B. Sonnweber, K. Kurz, S. Koppelstätter, D. Haschka, V. Petzer, A. Boehm, M. Aichner, P. Tymoszuk, D. Lener, M. Theurl, A. Lorsbach-Köhler, A. Tancevski, A. Schapfl, M. Schaber, R. Hilbe, M. Nairz, B. Puchner, D. Hüttenberger, C. Tschurtschenthaler, M. Aßhoff, A. Peer, F. Hartig, R. Bellmann, M. Joannidis, C. Gollmann-Tepeköylü, J. Holfeld, G. Feuchtner, A. Egger, G. Hoermann, A. Schroll, G. Fritsche, S. Wildner, R. Bellmann-Weiler, R. Kirchmair, R. Helbok, H. Prosch, D. Rieder, Z. Trajanoski, F. Kronenberg, E. Wöll, G. Weiss, G. Widmann, J. Löffler-Ragg, and I. Tancevski. Cardiopulmonary recovery after COVID-19: an observational prospective multicentre trial. *Eur Respir J*, 57(4), 04 2021.

[49] K. B. Jacobson, M. Rao, H. Bonilla, A. Subramanian, I. Hack, M. Madrigal, U. Singh, P. Jagannathan, and P. Grant. Patients with uncomplicated COVID-19 have long-term persistent symptoms and functional impairment similar to patients with severe COVID-19: a cautionary tale during a global pandemic. *Clin Infect Dis*, Feb 2021.

[50] H. J. Chun, E. Coutavas, A. Pine, A. I. Lee, V. Yu, M. Shallow, C. X. Giovacchini, A. Mathews, B. Stephenson, L. G. Que, P. J. Lee, and B. D. Kraft. Immuno-fibrotic drivers of impaired lung function in post-COVID-19 syndrome. *medRxiv*, Feb 2021.

[51] A. Baricich, M. B. Borg, D. Cuneo, E. Cadario, D. Azzolina, P. E. Balbo, M. Bellan, P. Zeppegno, M. Pirisi, C. Cisari, G. Aimaretti, G. C. Avanzi, G. Baldon, G. Bartolomei, M. Battaglia, S. Battistini, V. Binda, V. Cantaluppi, L. M. Castello, E. Clivati, M. Costanzo, A. Croce, C. DE Benedittis, S. DE Vecchi, A. Feggi, M. Gai, E. Gambaro, E. Gattoni, C. Gramaglia, L. Grisafi, C. Guerriero,

E. Hayden, A. Jona, M. Invernizzi, L. Lorenzini, L. Loreti, M. Martelli, E. Matino, A. Panero, E. Paracchini, F. Patrucco, G. Patti, A. Pirovano, P. Prosperini, R. Quaglino, C. Rigamonti, P. P. Sainaghi, D. Soddu, C. Vecchi, and E. Zecca. Midterm functional sequelae and implications in rehabilitation after COVID-19: a cross-sectional study. *Eur J Phys Rehabil Med*, 57(2):199–207, Apr 2021.

[58] S. J. Abdallah, N. Voduc, V. F. Corrales-Medina, M. McGuinty, A. Pratt, A. Chopra, A. Law, H. A. Garuba, K. Thavorn, R. E. R. Reid, K. L. Lavoie, A. Crawley, J. A. Chirinos, and J. Cowan. Symptoms, Pulmonary Function and Functional Capacity Four Months after COVID-19. *Ann Am Thorac Soc*, Apr 2021.

[68] O. O'Sullivan, R. M. Barker-Davies, K. Thompson, S. Bahadur, M. Gough, S. Lewis, M. Martin, A. Segalini, G. Wallace, R. Phillip, and M. Cranley. Rehabilitation post-COVID-19: cross-sectional observations using the Stanford Hall remote assessment tool. *BMJ Mil Health*, May 2021.

[69] Nida Ziauddeen, Deepti Gurdasani, Margaret E O'Hara, Claire Hastie, Paul Roderick, Guiqing Yao, and Nisreen A Alwan. Characteristics of long covid: findings from a social media survey. *medRxiv*, 2021.

[70] R. Omdal. The biological basis of chronic fatigue: neuroinflammation and innate immunity. *Curr Opin Neurol*, 33(3):391–396, 06 2020.

[71] N. Rohleder, M. Aringer, and M. Boentert. Role of interleukin-6 in stress, sleep, and fatigue. *Ann N Y Acad Sci*, 1261:88–96, Jul 2012.

[72] L. M. Trotti, J. C. Ong, D. T. Plante, C. Friederich Murray, R. King, and D. L. Bliwise. Disease symptomatology and response to treatment in people with idiopathic hypersomnia: initial data from the Hypersomnia Foundation registry. *Sleep Med*, 75:343–349, 11 2020.
