## Supplemental File 2 for "Characterizing Long COVID: Deep Phenotype of a Complex Condition"

| HPO.label | HPO.id | description.original | PMID | first.author | year | cohort | counts | percentage |
|---|---|---|---|---|---|---|---|---|
| Dyspnea | HP:0002094 | Breathlessness | PMID:33273026 | Arnold | 2020 | Inpatient (ICU); mean of 84.0 days post discharge | 10/18/21 | 55.60% |
| Dyspnea | HP:0002094 | Breathlessness | PMID:33273026 | Arnold | 2020 | Inpatient (non-ICU); mean of 84.0 days post discharge | 26/65 | 40.00% |
| Dyspnea | HP:0002094 | Breathlessness | PMID:33273026 | Arnold | 2020 | Inpatient (non-ICU); mean of 84.0 days post discharge | 07/27/21 | 25.90% |
| Dyspnea | HP:0002094 | Dyspnea at follow up | PMID:33502487 | Bellan | 2021 | Inpatient; mean of 105.0 days post discharge | 13/238 | 5.50% |
| Dyspnea | HP:0002094 | Dyspnea | PMID:33502487 | Bellan | 2021 | Inpatient; mean of 105.0 days post discharge | 13/238 | 5.50% |
| Dyspnea | HP:0002094 | Dyspnea | PMID:32644129 | Carfi | 2020 | Inpatient; mean of 60.3 days post discharge | 62/143 | 43.40% |
| Dyspnea | HP:0002094 | Dyspnoea/shortness of breath | PMID:33031948 | Carvalho-Schneider | 2021 | Mixed; mean of 60.0 days post diagnosis | 10/130 | 7.70% |
| Dyspnea | HP:0002094 | Dyspnoea/shortness of breath | PMID:33031948 | Carvalho-Schneider | 2021 | Mixed; mean of 30.0 days post diagnosis | 16/150 | 10.70% |
| Dyspnea | HP:0002094 | Forty-one patients (36%) had an MRC breathlessness score above their pre-admission baseline at follow-up | PMID:33731329 | Cheng | 2021 | Mixed; mean of 63.0 days post diagnosis | 41/113 | 36.30% |
| Dyspnea | HP:0002094 | Shortness of breath/chest tightness/wheezing | PMID:33175566 | Chopra | 2020 | Inpatient; mean of 60.0 days post discharge | 81/488 | 16.60% |
| Dyspnea | HP:0002094 | dyspnea (69%) | PMID:33564789 | Chun | 2021 | Mixed; mean of 63.0 days post diagnosis | 42/61 | 68.90% |
| Dyspnea | HP:0002094 | Dyspnea | PMID:33120193 | Daher | 2020 | Inpatient (ICU); mean of 56.0 days post discharge | 11/33 | 33.30% |
| Dyspnea | HP:0002094 | shortness of breath (15) | PMID:33657671 | Darley | 2021 | Mixed; mean of 69.0 days post diagnosis | 15/78 | 19.20% |
| Dyspnea | HP:0002094 | Shortness of Breath | PMID:-1 | Davis | 2020 | Mixed; mean of 114.5 days post diagnosis | 2913/3762 | 77.40% |
| Dyspnea | HP:0002094 | Mild dyspnoea - very severe dyspnoea | PMID:33052920 | De Lorenzo | 2020 | Inpatient; mean of 21.5 days post discharge | 40/126 | 31.70% |
| Dyspnea | HP:0002094 | Mild dyspnoea - Very severe dyspnoea | PMID:33052920 | De Lorenzo | 2020 | Outpatient; mean of 26.0 days post diagnosis | 18/59 | 30.50% |
| Dyspnea | HP:0002094 | Shortness of breath | PMID:33785495 | Dennis | 2021 | Inpatient; mean of 138.0 days post discharge | 35/37 | 94.60% |
| Dyspnea | HP:0002094 | Shortness of breath | PMID:33785495 | Dennis | 2021 | Outpatient; mean of 141.0 days post diagnosis | 141/163 | 86.50% |
| Dyspnea | HP:0002094 | Dyspnoea | PMID:32853602 | Garrigues | 2020 | Inpatient (non-ICU); mean of 110.9 days post discharge | 38/96 | 39.60% |
| Dyspnea | HP:0002094 | Dyspnoea | PMID:32853602 | Garrigues | 2020 | Inpatient (ICU); mean of 110.9 days post discharge | 12/24/21 | 50.00% |
| Dyspnea | HP:0002094 | Dyspnoea (71%) | PMID:33257910 | Goertz | 2020 | Mixed; mean of 79.0 days post diagnosis | 1500/2113 | 71.00% |
| Dyspnea | HP:0002094 | Dyspnea (1,2,3) | PMID:33676998 | González | 2021 | Inpatient (ICU); mean of 90.0 days post discharge | 28/62 | 45.20% |
| Dyspnea | HP:0002094 | Shortness of breath | PMID:33755344 | Graham | 2021 | Outpatient; mean of 141.0 days post diagnosis | 46/100 | 46.00% |
| Dyspnea | HP:0002094 | Any new or worsened breathlessness | PMID:32729939 | Halpin | 2021 | Inpatient (ICU); mean of 48.0 days post discharge | 21/32 | 65.60% |
| Dyspnea | HP:0002094 | Any new or worsened breathlessness | PMID:32729939 | Halpin | 2021 | Inpatient (non-ICU); mean of 48.0 days post discharge | 29/68 | 42.60% |
| Dyspnea | HP:0002094 | Dyspnea | PMID:33825846 | Havervall | 2021 | Outpatient; mean of 120.0 days post diagnosis | 11/323 | 3.40% |
| Dyspnea | HP:0002094 | Dyspnea | PMID:33825846 | Havervall | 2021 | Outpatient; mean of 60.0 days post diagnosis | 14/323 | 4.30% |
| Dyspnea | HP:0002094 | Dyspnea | PMID:33825846 | Havervall | 2021 | Outpatient; mean of 240.0 days post diagnosis | 6/323 | 1.90% |
| Dyspnea | HP:0002094 | Dyspnea | PMID:33680620 | Iqbal | 2021 | Mixed; mean of 38.1 days post diagnosis | 79/158 | 50.00% |
| Dyspnea | HP:0002094 | Shortness of breath | PMID:33306721 | Jacobs | 2020 | Inpatient (non-ICU); mean of 35.0 days post discharge | 58/183 | 31.70% |
| Dyspnea | HP:0002094 | Dyspnea | PMID:33624010 | Jacobson | 2021 | Inpatient; mean of 119.3 days post discharge | 11/22/21 | 50.00% |
| Dyspnea | HP:0002094 | Dyspnea | PMID:33624010 | Jacobson | 2021 | Outpatient; mean of 119.3 days post diagnosis | 20/96 | 20.80% |
| Dyspnea | HP:0002094 | Dyspnea | PMID:33289374 | Liang | 2020 | Inpatient (non-ICU); mean of 90.0 days post discharge | 46/76 | 60.50% |
| Dyspnea | HP:0002094 | Trouble breathing | PMID:33606031 | Logue | 2021 | Inpatient; mean of 169.0 days post discharge | 01/16/21 | 6.30% |
| Dyspnea | HP:0002094 | Trouble breathing | PMID:33606031 | Logue | 2021 | Outpatient; mean of 169.0 days post diagnosis | 10/161 | 6.20% |
| Dyspnea | HP:0002094 | dyspnoea | PMID:33205450 | Ludvigsson | 2021 | Outpatient; mean of 210.0 days post diagnosis | 05/05/21 | 100.00% |
| Dyspnea | HP:0002094 | 53% reported persistent breathlessness | PMID:33172844 | Mandal | 2020 | Inpatient; mean of 54.0 days post discharge | 204/384 | 53.10% |
| Dyspnea | HP:0002094 | Dyspnea | PMID:33450302 | Moreno-Perez | 2021 | Mixed; mean of 77.0 days post diagnosis | 95/277 | 34.30% |
| Dyspnea | HP:0002094 | Dyspnea | PMID:33865161 | Ordinola Navarro | 2021 | Mixed; mean of 40.0 days post diagnosis | 36/115 | 31.30% |
| Dyspnea | HP:0002094 | Dyspnea (Figure 1) | PMID:33252665 | Petersen | 2020 | Outpatient; mean of 99.0 days post diagnosis | 14/180 | 7.80% |
| Dyspnea | HP:0002094 | disnea 18% | PMID:33787016 | Prieto | 2021 | Mixed; mean of 53.0 days post diagnosis | 15/85 | 17.60% |
| Dyspnea | HP:0002094 | ongoing shortness of breath | PMID:32730619 | Puntmann | 2020 | Mixed; mean of 71.0 days post diagnosis | 36/100 | 36.00% |
| Dyspnea | HP:0002094 | Dyspnoea | PMID:33490928 | Ramani | 2021 | Inpatient; mean of 48.0 days post discharge | 36/56 | 64.30% |
| Dyspnea | HP:0002094 | dyspnoea (31.4%) | PMID:33521308 | Rosales-Castillo | 2021 | Inpatient; mean of 50.8 days post discharge | 37/118 | 31.40% |
| Dyspnea | HP:0002094 | Dyspnea | PMID:33303539 | Sonnweber | 2020 | Mixed; mean of 103.0 days post diagnosis | 52/145 | 35.90% |
| Dyspnea | HP:0002094 | Dyspnoea | PMID:33273028 | Stavem | 2020 | Outpatient; mean of 117.0 days post diagnosis | 72/451 | 16.00% |
| Dyspnea | HP:0002094 | Dyspnea | PMID:33654513 | Suárez-Robles | 2020 | Inpatient; mean of 90.0 days post discharge | 54/134 | 40.30% |
| Dyspnea | HP:0002094 | Dyspnea | PMID:32730238 | Tenforde | 2020 | Outpatient; mean of 17.5 days post diagnosis | 79/274 | 28.80% |
| Dyspnea | HP:0002094 | dyspnoea | PMID:33479105 | Trinkmann | 2021 | Mixed; mean of 68.0 days post diagnosis | 79/246 | 32.10% |
| Dyspnea | HP:0002094 | Dyspnoea | PMID:33461632 | Venturelli | 2021 | Mixed; mean of 81.0 days post diagnosis | 167/515 | 32.40% |
| Dyspnea | HP:0002094 | 113 (74.3%) participants reported some shortness of breath | PMID:33443703 | Weerahandi | 2021 | Inpatient; mean of 37.0 days post discharge | 113/152 | 74.30% |
| Dyspnea | HP:0002094 | dyspnoea (50%) | PMID:33008936 | Wong | 2020 | Inpatient; mean of 91.0 days post discharge | 39/78 | 50.00% |
| Dyspnea | HP:0002094 | Dyspnea | PMID:33729425 | Writing Committee | 2021 | Inpatient (ICU); mean of 93.0 days post discharge | 25/73 | 34.20% |
| Dyspnea | HP:0002094 | Dyspnea | PMID:33729425 | Writing Committee | 2021 | Inpatient (non-ICU); mean of 121.0 days post discharge | 53/405 | 13.10% |
| Dyspnea | HP:0002094 | Dyspnea | PMID:33532720 | de Graaf | 2021 | Inpatient (ICU); mean of 42.0 days post discharge | 23/34 | 67.60% |
| Dyspnea | HP:0002094 | Dyspnea | PMID:33532720 | de Graaf | 2021 | Inpatient (non-ICU); mean of 42.0 days post discharge | 23/47 | 48.90% |
| Fatigue | HP:0012378 | Fatigue | PMID:33872135 | Abdallah | 2021 | Inpatient; mean of 102.3 days post discharge | 18/25 | 72.00% |
| Fatigue | HP:0012378 | Fatigue | PMID:33872135 | Abdallah | 2021 | Outpatient; mean of 129.8 days post diagnosis | 37/38 | 97.40% |
| Fatigue | HP:0012378 | Excessive Fatigue | PMID:33273026 | Arnold | 2020 | Inpatient (ICU); mean of 84.0 days post discharge | 10/18/21 | 55.60% |
| Fatigue | HP:0012378 | Excessive Fatigue | PMID:33273026 | Arnold | 2020 | Inpatient (non-ICU); mean of 84.0 days post discharge | 26/65 | 40.00% |

| | | | | | | | | |
|---|---|---|---|---|---|---|---|---|
| Fatigue | HP:0012378 | Excessive Fatigue | PMID:33273026 | Arnold | 2020 | Inpatient (non-ICU); mean of 84.0 days post discharge | 07/27/21 | 25.90% |
| Fatigue | HP:0012378 | Fatigue | PMID:32644129 | Carfi | 2020 | Inpatient; mean of 60.3 days post discharge | 76/143 | 53.10% |
| Fatigue | HP:0012378 | fatigue | PMID:33731329 | Cheng | 2021 | Mixed; mean of 63.0 days post diagnosis | 69/113 | 61.10% |
| Fatigue | HP:0012378 | fatigue (29%) | PMID:33564789 | Chun | 2021 | Mixed; mean of 63.0 days post diagnosis | 18/61 | 29.50% |
| Fatigue | HP:0012378 | Fatigue | PMID:33120193 | Daher | 2020 | Inpatient (ICU); mean of 56.0 days post discharge | 15/33 | 45.50% |
| Fatigue | HP:0012378 | fatigue (17 patients) | PMID:33657671 | Darley | 2021 | Mixed; mean of 69.0 days post diagnosis | 17/78 | 21.80% |
| Fatigue | HP:0012378 | Fatigue | PMID:-1 | Davis | 2020 | Mixed; mean of 114.5 days post diagnosis | 3699/3762 | 98.30% |
| Fatigue | HP:0012378 | Fatigue | PMID:33785495 | Dennis | 2021 | Inpatient; mean of 138.0 days post discharge | 37/37 | 100.00% |
| Fatigue | HP:0012378 | Fatigue | PMID:33785495 | Dennis | 2021 | Outpatient; mean of 141.0 days post diagnosis | 159/163 | 97.50% |
| Fatigue | HP:0012378 | Fatigue | PMID:32853602 | Garrigues | 2020 | Inpatient (non-ICU); mean of 110.9 days post discharge | 52/96 | 54.20% |
| Fatigue | HP:0012378 | Fatigue | PMID:32853602 | Garrigues | 2020 | Inpatient (ICU); mean of 110.9 days post discharge | 14/24 | 58.30% |
| Fatigue | HP:0012378 | Fatigue (87%) | PMID:33257910 | Goertz | 2020 | Mixed; mean of 79.0 days post diagnosis | 1838/2113 | 87.00% |
| Fatigue | HP:0012378 | Muscular fatigue | PMID:33676998 | González | 2021 | Inpatient (ICU); mean of 90.0 days post discharge | 18/62 | 29.00% |
| Fatigue | HP:0012378 | Fatigue | PMID:33755344 | Graham | 2021 | Outpatient; mean of 141.0 days post diagnosis | 85/100 | 85.00% |
| Fatigue | HP:0012378 | Any new fatigue | PMID:32729939 | Halpin | 2021 | Inpatient (ICU); mean of 48.0 days post discharge | 23/32 | 71.90% |
| Fatigue | HP:0012378 | Any new fatigue | PMID:32729939 | Halpin | 2021 | Inpatient (non-ICU); mean of 48.0 days post discharge | 41/68 | 60.30% |
| Fatigue | HP:0012378 | Fatigue | PMID:33825846 | Havervall | 2021 | Outpatient; mean of 120.0 days post diagnosis | 22/323 | 6.80% |
| Fatigue | HP:0012378 | Fatigue | PMID:33825846 | Havervall | 2021 | Outpatient; mean of 60.0 days post diagnosis | 27/323 | 8.40% |
| Fatigue | HP:0012378 | Fatigue | PMID:33825846 | Havervall | 2021 | Outpatient; mean of 240.0 days post diagnosis | 13/323 | 4.00% |
| Fatigue | HP:0012378 | Fatigue or muscle weakness | PMID:33428867 | Huang | 2021 | Inpatient; mean of 186.0 days post discharge | 1038/1655 | 62.70% |
| Fatigue | HP:0012378 | Fatigue | PMID:33680620 | Iqbal | 2021 | Mixed; mean of 38.1 days post diagnosis | 131/158 | 82.90% |
| Fatigue | HP:0012378 | Fatigue | PMID:33306721 | Jacobs | 2020 | Inpatient (non-ICU); mean of 35.0 days post discharge | 82/183 | 44.80% |
| Fatigue | HP:0012378 | Fatigue | PMID:33624010 | Jacobson | 2021 | Inpatient; mean of 119.3 days post discharge | 08/22/21 | 36.40% |
| Fatigue | HP:0012378 | Fatigue | PMID:33624010 | Jacobson | 2021 | Outpatient; mean of 119.3 days post diagnosis | 28/96 | 29.20% |
| Fatigue | HP:0012378 | Fatigue | PMID:33289374 | Liang | 2020 | Inpatient (non-ICU); mean of 90.0 days post discharge | 45/76 | 59.20% |
| Fatigue | HP:0012378 | Fatigue | PMID:33606031 | Logue | 2021 | Inpatient; mean of 169.0 days post discharge | 04/16/21 | 25.00% |
| Fatigue | HP:0012378 | Fatigue | PMID:33606031 | Logue | 2021 | Outpatient; mean of 169.0 days post diagnosis | 20/161 | 12.40% |
| Fatigue | HP:0012378 | fatigue | PMID:33205450 | Ludvigsson | 2021 | Outpatient; mean of 210.0 days post diagnosis | 05/05/21 | 100.00% |
| Fatigue | HP:0012378 | at early follow-up, 69% had persistent fatigue | PMID:33172844 | Mandal | 2020 | Inpatient; mean of 54.0 days post discharge | 265/384 | 69.00% |
| Fatigue | HP:0012378 | Fatigue | PMID:33450302 | Moreno-Perez | 2021 | Mixed; mean of 77.0 days post diagnosis | 96/277 | 34.70% |
| Fatigue | HP:0012378 | Fatigue | PMID:33865161 | Ordinola Navarro | 2021 | Mixed; mean of 40.0 days post diagnosis | 18/115 | 15.70% |
| Fatigue | HP:0012378 | Fatigue (Figure 1) | PMID:33252665 | Petersen | 2020 | Outpatient; mean of 99.0 days post diagnosis | 50/180 | 27.80% |
| Fatigue | HP:0012378 | la fatiga (49%) | PMID:33787016 | Prieto | 2021 | Mixed; mean of 53.0 days post diagnosis | 42/85 | 49.40% |
| Fatigue | HP:0012378 | general exhaustion | PMID:32730619 | Puntmann | 2020 | Mixed; mean of 71.0 days post diagnosis | 36/100 | 36.00% |
| Fatigue | HP:0012378 | Fatigue | PMID:33490928 | Ramani | 2021 | Inpatient; mean of 48.0 days post discharge | 30/55 | 54.50% |
| Fatigue | HP:0012378 | General malaise | PMID:33654513 | Suárez-Robles | 2020 | Inpatient; mean of 90.0 days post discharge | 25/134 | 18.70% |
| Fatigue | HP:0012378 | Fatigue | PMID:32730238 | Tenforde | 2020 | Outpatient; mean of 17.5 days post diagnosis | 101/274 | 36.90% |
| Fatigue | HP:0012378 | Based on the CFQ-11 case definition, 52.3% (67/128) met the criteria for fatigue | PMID:33166287 | Townsend | 2020 | Mixed; mean of 72.0 days post diagnosis | 67/128 | 52.30% |
| Fatigue | HP:0012378 | Seventy--seven patients (51%) met the case definition for fatigue | PMID:33587810 | Townsend | 2021 | Mixed; mean of 80.5 days post diagnosis | 77/150 | 51.30% |
| Fatigue | HP:0012378 | fatigue | PMID:33479105 | Trinkmann | 2021 | Mixed; mean of 68.0 days post diagnosis | 2/246 | 0.80% |
| Fatigue | HP:0012378 | Fatigue | PMID:33729425 | Writing Committee | 2021 | Inpatient (ICU); mean of 93.0 days post discharge | 24/63 | 38.10% |
| Fatigue | HP:0012378 | Fatigue | PMID:33729425 | Writing Committee | 2021 | Inpatient (non-ICU); mean of 121.0 days post discharge | 110/368 | 29.90% |
| Fatigue | HP:0012378 | Physical decline/fatigue | PMID:32979574 | Xiong | 2021 | Inpatient; mean of 97.0 days post discharge | 152/538 | 28.30% |
| Fatigue | HP:0012378 | fatigue (16.36%) | PMID:32838236 | Zhao | 2020 | Inpatient; mean of 78.5 days post discharge | 9/55 | 16.40% |
| Anosmia | HP:0000458 | Anosmia | PMID:33273026 | Arnold | 2020 | Inpatient (ICU); mean of 84.0 days post discharge | 04/18/21 | 22.20% |
| Anosmia | HP:0000458 | Anosmia | PMID:33273026 | Arnold | 2020 | Inpatient (non-ICU); mean of 84.0 days post discharge | 6/65 | 9.20% |
| Anosmia | HP:0000458 | Anosmia | PMID:33273026 | Arnold | 2020 | Inpatient (non-ICU); mean of 84.0 days post discharge | 03/27/21 | 11.10% |
| Anosmia | HP:0000458 | Anosmia At follow-up | PMID:33502487 | Bellan | 2021 | Inpatient; mean of 105.0 days post discharge | 11/238 | 4.60% |
| Anosmia | HP:0000458 | Anosmia | PMID:33502487 | Bellan | 2021 | Inpatient; mean of 105.0 days post discharge | 11/238 | 4.60% |
| Anosmia | HP:0000458 | Anosmia | PMID:32644129 | Carfi | 2020 | Inpatient; mean of 60.3 days post discharge | 23/143 | 16.10% |
| Anosmia | HP:0000458 | Anosmia/ageusia | PMID:33031948 | Carvalho-Schneider | 2021 | Mixed; mean of 60.0 days post diagnosis | 29/130 | 22.30% |
| Anosmia | HP:0000458 | Anosmia/ageusia | PMID:33031948 | Carvalho-Schneider | 2021 | Mixed; mean of 30.0 days post diagnosis | 40/150 | 26.70% |
| Anosmia | HP:0000458 | Continued loss of taste and/or smell | PMID:33175566 | Chopra | 2020 | Inpatient; mean of 60.0 days post discharge | 64/488 | 13.10% |
| Anosmia | HP:0000458 | Anosmia | PMID:33564789 | Chun | 2021 | Mixed; mean of 63.0 days post diagnosis | 10/61 | 16.40% |
| Anosmia | HP:0000458 | Loss of Smell | PMID:33120193 | Daher | 2020 | Inpatient (ICU); mean of 56.0 days post discharge | 4/33 | 12.10% |
| Anosmia | HP:0000458 | Impaired olfaction was evident in 18 patients, including four with severe anosmia | PMID:33657671 | Darley | 2021 | Mixed; mean of 69.0 days post diagnosis | 18/78 | 23.10% |
| Anosmia | HP:0000458 | Loss of smell | PMID:-1 | Davis | 2020 | Mixed; mean of 114.5 days post diagnosis | 1352/3762 | 35.90% |
| Anosmia | HP:0000458 | Anosmia | PMID:32853602 | Garrigues | 2020 | Inpatient (non-ICU); mean of 110.9 days post discharge | 14/96 | 14.60% |
| Anosmia | HP:0000458 | Anosmia | PMID:32853602 | Garrigues | 2020 | Inpatient (ICU); mean of 110.9 days post discharge | 02/24/21 | 8.30% |
| Anosmia | HP:0000458 | Anosmia (13%) | PMID:33257910 | Goertz | 2020 | Mixed; mean of 79.0 days post diagnosis | 275/2113 | 13.00% |

| | | | | | | | | |
|---|---|---|---|---|---|---|---|---|
| Anosmia | HP:0000458 | Anosmia | PMID:33755344 | Graham | 2021 | Outpatient; mean of 141.0 days post diagnosis | 55/100 | 55.00% |
| Anosmia | HP:0000458 | Anosmia | PMID:33825846 | Havervall | 2021 | Outpatient; mean of 120.0 days post diagnosis | 35/323 | 10.80% |
| Anosmia | HP:0000458 | Anosmia | PMID:33825846 | Havervall | 2021 | Outpatient; mean of 60.0 days post diagnosis | 47/323 | 14.60% |
| Anosmia | HP:0000458 | Anosmia | PMID:33825846 | Havervall | 2021 | Outpatient; mean of 240.0 days post diagnosis | 29/323 | 9.00% |
| Anosmia | HP:0000458 | Smell disorder | PMID:33428867 | Huang | 2021 | Inpatient; mean of 186.0 days post discharge | 176/1655 | 10.60% |
| Anosmia | HP:0000458 | Loss of smell and taste | PMID:33680620 | Iqbal | 2021 | Mixed; mean of 38.1 days post diagnosis | 75/158 | 47.50% |
| Anosmia | HP:0000458 | Lack of smell | PMID:33306721 | Jacobs | 2020 | Inpatient (non-ICU); mean of 35.0 days post discharge | 17/183 | 9.30% |
| Anosmia | HP:0000458 | Loss of taste/smell | PMID:33624010 | Jacobson | 2021 | Inpatient; mean of 119.3 days post discharge | 02/22/21 | 9.10% |
| Anosmia | HP:0000458 | Loss of taste/smell | PMID:33624010 | Jacobson | 2021 | Outpatient; mean of 119.3 days post diagnosis | 23/96 | 24.00% |
| Anosmia | HP:0000458 | Loss of sense of taste or smell | PMID:33606031 | Logue | 2021 | Inpatient; mean of 169.0 days post discharge | 02/16/21 | 12.50% |
| Anosmia | HP:0000458 | Loss of sense of taste or smell | PMID:33606031 | Logue | 2021 | Outpatient; mean of 169.0 days post diagnosis | 21/161 | 13.00% |
| Anosmia | HP:0000458 | deranged smell and taste | PMID:33205450 | Ludvigsson | 2021 | Outpatient; mean of 210.0 days post diagnosis | 01/05/21 | 20.00% |
| Anosmia | HP:0000458 | Anosmia-dysgeusia | PMID:33450302 | Moreno-Perez | 2021 | Mixed; mean of 77.0 days post diagnosis | 59/277 | 21.30% |
| Anosmia | HP:0000458 | Loss of smell (Figure 1) | PMID:33252665 | Petersen | 2020 | Outpatient; mean of 99.0 days post diagnosis | 41/180 | 22.80% |
| Anosmia | HP:0000458 | Anosmia 22% | PMID:33787016 | Prieto | 2021 | Mixed; mean of 53.0 days post diagnosis | 19/85 | 22.40% |
| Anosmia | HP:0000458 | Anosmia, self-reported | PMID:33682276 | Rass | 2021 | Outpatient; mean of 90.0 days post diagnosis | 0/32 | 0.00% |
| Anosmia | HP:0000458 | Anosmia, self-reported | PMID:33682276 | Rass | 2021 | Inpatient (non-ICU); mean of 90.0 days post discharge | 2/72 | 2.80% |
| Anosmia | HP:0000458 | Anosmia, self-reported | PMID:33682276 | Rass | 2021 | Inpatient (ICU); mean of 90.0 days post discharge | 0/31 | 0.00% |
| Anosmia | HP:0000458 | anosmia (1.7%) | PMID:33521308 | Rosales-Castillo | 2021 | Inpatient; mean of 50.8 days post discharge | 2/118 | 1.70% |
| Anosmia | HP:0000458 | Hyposmia/.anosmia | PMID:33303539 | Sonnweber | 2020 | Mixed; mean of 103.0 days post diagnosis | 28/145 | 19.30% |
| Anosmia | HP:0000458 | Loss/disturbance of smell | PMID:33273028 | Stavem | 2020 | Outpatient; mean of 117.0 days post diagnosis | 54/451 | 12.00% |
| Anosmia | HP:0000458 | Anosmia | PMID:33654513 | Suárez-Robles | 2020 | Inpatient; mean of 90.0 days post discharge | 35/134 | 26.10% |
| Anosmia | HP:0000458 | Anosmia | PMID:33413976 | Taboada | 2021 | Inpatient (ICU); mean of 180.0 days post discharge | 10/91 | 11.00% |
| Anosmia | HP:0000458 | Loss of smell | PMID:32730238 | Tenforde | 2020 | Outpatient; mean of 17.5 days post diagnosis | 60/274 | 21.90% |
| Anosmia | HP:0000458 | Olfactory loss | PMID:33479105 | Trinkmann | 2021 | Mixed; mean of 68.0 days post diagnosis | 10/246 | 4.10% |
| Anosmia | HP:0000458 | Anosmia/Dysgeusia | PMID:33461632 | Venturelli | 2021 | Mixed; mean of 81.0 days post diagnosis | 23/515 | 4.50% |
| Anosmia | HP:0000458 | Anosmia | PMID:33729425 | Writing Committee | 2021 | Inpatient (ICU); mean of 93.0 days post discharge | 6/62 | 9.70% |
| Anosmia | HP:0000458 | Anosmia | PMID:33729425 | Writing Committee | 2021 | Inpatient (non-ICU); mean of 121.0 days post discharge | 19/357 | 5.30% |
| Headache | HP:0002315 | Headache | PMID:33273026 | Arnold | 2020 | Inpatient (ICU); mean of 84.0 days post discharge | 0/18 | 0.00% |
| Headache | HP:0002315 | Headache | PMID:33273026 | Arnold | 2020 | Inpatient (non-ICU); mean of 84.0 days post discharge | 1/65 | 1.50% |
| Headache | HP:0002315 | Headache | PMID:33273026 | Arnold | 2020 | Inpatient (non-ICU); mean of 84.0 days post discharge | 01/27/21 | 3.70% |
| Headache | HP:0002315 | Headache At follow-up | PMID:33502487 | Bellan | 2021 | Inpatient; mean of 105.0 days post discharge | 0/238 | 0.00% |
| Headache | HP:0002315 | Headache | PMID:33502487 | Bellan | 2021 | Inpatient; mean of 105.0 days post discharge | 0/238 | 0.00% |
| Headache | HP:0002315 | Headache | PMID:32644129 | Carfi | 2020 | Inpatient; mean of 60.3 days post discharge | 13/143 | 9.10% |
| Headache | HP:0002315 | Flulike symptoms (Myalgia, headache and/or asthenia) | PMID:33031948 | Carvalho-Schneider | 2021 | Mixed; mean of 60.0 days post diagnosis | 28/130 | 21.50% |
| Headache | HP:0002315 | Flulike symptoms (Myalgia, headache and/or asthenia) | PMID:33031948 | Carvalho-Schneider | 2021 | Mixed; mean of 30.0 days post diagnosis | 54/150 | 36.00% |
| Headache | HP:0002315 | headache (23%) | PMID:33564789 | Chun | 2021 | Mixed; mean of 63.0 days post diagnosis | 14/61 | 23.00% |
| Headache | HP:0002315 | Headache | PMID:33120193 | Daher | 2020 | Inpatient (ICU); mean of 56.0 days post discharge | 5/33 | 15.20% |
| Headache | HP:0002315 | Headache | PMID:33657671 | Darley | 2021 | Mixed; mean of 69.0 days post diagnosis | 7/78 | 9.00% |
| Headache | HP:0002315 | Headaches, behind the eyes | PMID:-1 | Davis | 2020 | Mixed; mean of 114.5 days post diagnosis | 1509/3762 | 40.10% |
| Headache | HP:0002315 | Headache | PMID:33785495 | Dennis | 2021 | Inpatient; mean of 138.0 days post discharge | 27/37 | 73.00% |
| Headache | HP:0002315 | Headache | PMID:33785495 | Dennis | 2021 | Outpatient; mean of 141.0 days post diagnosis | 138/163 | 84.70% |
| Headache | HP:0002315 | Headache (38%) | PMID:33257910 | Goertz | 2020 | Mixed; mean of 79.0 days post diagnosis | 803/2113 | 38.00% |
| Headache | HP:0002315 | Headache | PMID:33755344 | Graham | 2021 | Outpatient; mean of 141.0 days post diagnosis | 68/100 | 68.00% |
| Headache | HP:0002315 | Headache | PMID:33825846 | Havervall | 2021 | Outpatient; mean of 120.0 days post diagnosis | 8/323 | 2.50% |
| Headache | HP:0002315 | Headache | PMID:33825846 | Havervall | 2021 | Outpatient; mean of 60.0 days post diagnosis | 9/323 | 2.80% |
| Headache | HP:0002315 | Headache | PMID:33825846 | Havervall | 2021 | Outpatient; mean of 240.0 days post diagnosis | 5/323 | 1.50% |
| Headache | HP:0002315 | Headache | PMID:33428867 | Huang | 2021 | Inpatient; mean of 186.0 days post discharge | 33/1655 | 2.00% |
| Headache | HP:0002315 | Continuous headache | PMID:33680620 | Iqbal | 2021 | Mixed; mean of 38.1 days post diagnosis | 57/158 | 36.10% |
| Headache | HP:0002315 | Headache | PMID:33306721 | Jacobs | 2020 | Inpatient (non-ICU); mean of 35.0 days post discharge | 23/183 | 12.60% |
| Headache | HP:0002315 | Headache | PMID:33624010 | Jacobson | 2021 | Inpatient; mean of 119.3 days post discharge | 0/22 | 0.00% |
| Headache | HP:0002315 | Headache | PMID:33624010 | Jacobson | 2021 | Outpatient; mean of 119.3 days post diagnosis | 7/96 | 7.30% |
| Headache | HP:0002315 | Headache | PMID:33606031 | Logue | 2021 | Inpatient; mean of 169.0 days post discharge | 0/16 | 0.00% |
| Headache | HP:0002315 | Headache | PMID:33606031 | Logue | 2021 | Outpatient; mean of 169.0 days post diagnosis | 8/161 | 5.00% |
| Headache | HP:0002315 | headaches | PMID:33205450 | Ludvigsson | 2021 | Outpatient; mean of 210.0 days post diagnosis | 04/05/21 | 80.00% |
| Headache | HP:0002315 | Headache | PMID:33450302 | Moreno-Perez | 2021 | Mixed; mean of 77.0 days post diagnosis | 49/277 | 17.70% |
| Headache | HP:0002315 | Headache | PMID:33865161 | Ordinola Navarro | 2021 | Mixed; mean of 40.0 days post diagnosis | 11/115 | 9.60% |
| Headache | HP:0002315 | Headache (Figure 1) | PMID:33252665 | Petersen | 2020 | Outpatient; mean of 99.0 days post diagnosis | 13/180 | 7.20% |
| Headache | HP:0002315 | Cefalea 11% | PMID:33787016 | Prieto | 2021 | Mixed; mean of 53.0 days post diagnosis | 9/85 | 10.60% |
| Headache | HP:0002315 | New cephalea | PMID:33682276 | Rass | 2021 | Outpatient; mean of 90.0 days post diagnosis | 1/32 | 3.10% |
| Headache | HP:0002315 | New cephalea | PMID:33682276 | Rass | 2021 | Inpatient (non-ICU); mean of 90.0 days post discharge | 2/72 | 2.80% |
| Headache | HP:0002315 | New cephalea | PMID:33682276 | Rass | 2021 | Inpatient (ICU); mean of 90.0 days post discharge | 03/31/21 | 9.70% |

| | | | | | | | | |
|---|---|---|---|---|---|---|---|---|
| Headache | HP:0002315 | Headache | PMID:33273028 | Stavem | 2020 | Outpatient; mean of 117.0 days post diagnosis | 27/451 | 6.00% |
| Headache | HP:0002315 | Headaches | PMID:33654513 | Suárez-Robles | 2020 | Inpatient; mean of 90.0 days post discharge | 33/134 | 24.60% |
| Headache | HP:0002315 | Headache | PMID:32730238 | Tenforde | 2020 | Outpatient; mean of 17.5 days post diagnosis | 44/274 | 16.10% |
| Headache | HP:0002315 | cephalgia | PMID:33479105 | Trinkmann | 2021 | Mixed; mean of 68.0 days post diagnosis | 1/246 | 0.40% |
| Headache | HP:0002315 | Headache | PMID:33461632 | Venturelli | 2021 | Mixed; mean of 81.0 days post diagnosis | 4/515 | 0.80% |
| Headache | HP:0002315 | Headaches | PMID:33729425 | Writing Committee | 2021 | Inpatient (ICU); mean of 93.0 days post discharge | 1/62 | 1.60% |
| Headache | HP:0002315 | Headaches | PMID:33729425 | Writing Committee | 2021 | Inpatient (non-ICU); mean of 121.0 days post discharge | 22/358 | 6.10% |
| Headache | HP:0002315 | headache (18.18%) | PMID:32838236 | Zhao | 2020 | Inpatient; mean of 78.5 days post discharge | 10/55 | 18.20% |
| Cough | HP:0012735 | Cough | PMID:33273026 | Arnold | 2020 | Inpatient (ICU); mean of 84.0 days post discharge | 01/18/21 | 5.60% |
| Cough | HP:0012735 | Cough | PMID:33273026 | Arnold | 2020 | Inpatient (non-ICU); mean of 84.0 days post discharge | 10/65 | 15.40% |
| Cough | HP:0012735 | Cough | PMID:33273026 | Arnold | 2020 | Inpatient (non-ICU); mean of 84.0 days post discharge | 02/27/21 | 7.40% |
| Cough | HP:0012735 | Cough at follow up | PMID:33502487 | Bellan | 2021 | Inpatient; mean of 105.0 days post discharge | 6/238 | 2.50% |
| Cough | HP:0012735 | Cough | PMID:33502487 | Bellan | 2021 | Inpatient; mean of 105.0 days post discharge | 6/238 | 2.50% |
| Cough | HP:0012735 | Cough | PMID:32644129 | Carfi | 2020 | Inpatient; mean of 60.3 days post discharge | 24/143 | 16.80% |
| Cough | HP:0012735 | cough (17%) | PMID:33731329 | Cheng | 2021 | Mixed; mean of 63.0 days post diagnosis | 19/113 | 16.80% |
| Cough | HP:0012735 | Cough | PMID:33175566 | Chopra | 2020 | Inpatient; mean of 60.0 days post discharge | 75/488 | 15.40% |
| Cough | HP:0012735 | cough (58%) | PMID:33564789 | Chun | 2021 | Mixed; mean of 63.0 days post diagnosis | 35/61 | 57.40% |
| Cough | HP:0012735 | Cough | PMID:33120193 | Daher | 2020 | Inpatient (ICU); mean of 56.0 days post discharge | 11/33 | 33.30% |
| Cough | HP:0012735 | Cough | PMID:33657671 | Darley | 2021 | Mixed; mean of 69.0 days post diagnosis | 14/78 | 17.90% |
| Cough | HP:0012735 | Cough | PMID:33785495 | Dennis | 2021 | Inpatient; mean of 138.0 days post discharge | 29/37 | 78.40% |
| Cough | HP:0012735 | Cough | PMID:33785495 | Dennis | 2021 | Outpatient; mean of 141.0 days post diagnosis | 117/163 | 71.80% |
| Cough | HP:0012735 | Cough | PMID:32853602 | Garrigues | 2020 | Inpatient (non-ICU); mean of 110.9 days post discharge | 14/96 | 14.60% |
| Cough | HP:0012735 | Cough | PMID:32853602 | Garrigues | 2020 | Inpatient (ICU); mean of 110.9 days post discharge | 06/24/21 | 25.00% |
| Cough | HP:0012735 | Cough (29%) | PMID:33257910 | Goertz | 2020 | Mixed; mean of 79.0 days post diagnosis | 613/2113 | 29.00% |
| Cough | HP:0012735 | Persistent cough | PMID:33680620 | Iqbal | 2021 | Mixed; mean of 38.1 days post diagnosis | 70/158 | 44.30% |
| Cough | HP:0012735 | Cough | PMID:33306721 | Jacobs | 2020 | Inpatient (non-ICU); mean of 35.0 days post discharge | 46/183 | 25.10% |
| Cough | HP:0012735 | Cough | PMID:33624010 | Jacobson | 2021 | Inpatient; mean of 119.3 days post discharge | 0/22 | 0.00% |
| Cough | HP:0012735 | Cough | PMID:33624010 | Jacobson | 2021 | Outpatient; mean of 119.3 days post diagnosis | 1/96 | 1.00% |
| Cough | HP:0012735 | Cough | PMID:33289374 | Liang | 2020 | Inpatient (non-ICU); mean of 90.0 days post discharge | 45/76 | 59.20% |
| Cough | HP:0012735 | Cough | PMID:32692945 | Liu | 2020 | Inpatient; mean of 28.0 days post discharge | 8/51 | 15.70% |
| Cough | HP:0012735 | Cough | PMID:33606031 | Logue | 2021 | Inpatient; mean of 169.0 days post discharge | 01/16/21 | 6.30% |
| Cough | HP:0012735 | Cough | PMID:33606031 | Logue | 2021 | Outpatient; mean of 169.0 days post diagnosis | 5/161 | 3.10% |
| Cough | HP:0012735 | chronic cough | PMID:33205450 | Ludvigsson | 2021 | Outpatient; mean of 210.0 days post diagnosis | 01/05/21 | 20.00% |
| Cough | HP:0012735 | at early follow-up, 34% had persistent cough | PMID:33172844 | Mandal | 2020 | Inpatient; mean of 54.0 days post discharge | 131/384 | 34.10% |
| Cough | HP:0012735 | Cough | PMID:33450302 | Moreno-Perez | 2021 | Mixed; mean of 77.0 days post diagnosis | 59/277 | 21.30% |
| Cough | HP:0012735 | Cough | PMID:33865161 | Ordinola Navarro | 2021 | Mixed; mean of 40.0 days post diagnosis | 7/115 | 6.10% |
| Cough | HP:0012735 | la tos (33%) | PMID:33787016 | Prieto | 2021 | Mixed; mean of 53.0 days post diagnosis | 28/85 | 32.90% |
| Cough | HP:0012735 | cough (5%) | PMID:33521308 | Rosales-Castillo | 2021 | Inpatient; mean of 50.8 days post discharge | 6/118 | 5.10% |
| Cough | HP:0012735 | Cough | PMID:33303539 | Sonnweber | 2020 | Mixed; mean of 103.0 days post diagnosis | 25/145 | 17.20% |
| Cough | HP:0012735 | Cough | PMID:33654513 | Suárez-Robles | 2020 | Inpatient; mean of 90.0 days post discharge | 36/134 | 26.90% |
| Cough | HP:0012735 | Cough | PMID:33413976 | Taboada | 2021 | Inpatient (ICU); mean of 180.0 days post discharge | 13/91 | 14.30% |
| Cough | HP:0012735 | Cough | PMID:32730238 | Tenforde | 2020 | Outpatient; mean of 17.5 days post diagnosis | 123/274 | 44.90% |
| Cough | HP:0012735 | cough | PMID:33479105 | Trinkmann | 2021 | Mixed; mean of 68.0 days post diagnosis | 35/246 | 14.20% |
| Cough | HP:0012735 | Cough | PMID:33461632 | Venturelli | 2021 | Mixed; mean of 81.0 days post diagnosis | 23/515 | 4.50% |
| Cough | HP:0012735 | 23% of patients reported the presence of cough | PMID:33008936 | Wong | 2020 | Inpatient; mean of 91.0 days post discharge | 18/78 | 23.10% |
| Cough | HP:0012735 | Cough | PMID:33729425 | Writing Committee | 2021 | Inpatient (ICU); mean of 93.0 days post discharge | 5/62 | 8.10% |
| Cough | HP:0012735 | Cough | PMID:33729425 | Writing Committee | 2021 | Inpatient (non-ICU); mean of 121.0 days post discharge | 21/420 | 5.00% |
| Cough | HP:0012735 | Cough | PMID:32979574 | Xiong | 2021 | Inpatient; mean of 97.0 days post discharge | 38/538 | 7.10% |
| Myalgia | HP:0003326 | Myalgia | PMID:33273026 | Arnold | 2020 | Inpatient (ICU); mean of 84.0 days post discharge | 07/18/21 | 38.90% |
| Myalgia | HP:0003326 | Myalgia | PMID:33273026 | Arnold | 2020 | Inpatient (non-ICU); mean of 84.0 days post discharge | 14/65 | 21.50% |
| Myalgia | HP:0003326 | Myalgia | PMID:33273026 | Arnold | 2020 | Inpatient (non-ICU); mean of 84.0 days post discharge | 04/27/21 | 14.80% |
| Myalgia | HP:0003326 | Myalgia At follow-up | PMID:33502487 | Bellan | 2021 | Inpatient; mean of 105.0 days post discharge | 14/238 | 5.90% |
| Myalgia | HP:0003326 | Myalgia | PMID:33502487 | Bellan | 2021 | Inpatient; mean of 105.0 days post discharge | 14/238 | 5.90% |
| Myalgia | HP:0003326 | Myalgia | PMID:32644129 | Carfi | 2020 | Inpatient; mean of 60.3 days post discharge | 7/143 | 4.90% |
| Myalgia | HP:0003326 | Flulike symptoms (Myalgia, headache and/or asthenia) | PMID:33031948 | Carvalho-Schneider | 2021 | Mixed; mean of 60.0 days post diagnosis | 28/130 | 21.50% |
| Myalgia | HP:0003326 | Flulike symptoms (Myalgia, headache and/or asthenia) | PMID:33031948 | Carvalho-Schneider | 2021 | Mixed; mean of 30.0 days post diagnosis | 54/150 | 36.00% |
| Myalgia | HP:0003326 | Myalgia | PMID:33120193 | Daher | 2020 | Inpatient (ICU); mean of 56.0 days post discharge | 5/33 | 15.20% |
| Myalgia | HP:0003326 | Muscle aches | PMID:33657671 | Darley | 2021 | Mixed; mean of 69.0 days post diagnosis | 5/78 | 6.40% |
| Myalgia | HP:0003326 | Muscle aches | PMID:-1 | Davis | 2020 | Mixed; mean of 114.5 days post diagnosis | 2601/3762 | 69.10% |
| Myalgia | HP:0003326 | Muscle ache | PMID:33785495 | Dennis | 2021 | Inpatient; mean of 138.0 days post discharge | 31/37 | 83.80% |
| Myalgia | HP:0003326 | Muscle ache | PMID:33785495 | Dennis | 2021 | Outpatient; mean of 141.0 days post diagnosis | 142/163 | 87.10% |
| Myalgia | HP:0003326 | Muscle pain (36%) | PMID:33257910 | Goertz | 2020 | Mixed; mean of 79.0 days post diagnosis | 761/2113 | 36.00% |

| | | | | | | | | |
|---|---|---|---|---|---|---|---|---|
| Myalgia | HP:0003326 | Myalgia | PMID:33755344 | Graham | 2021 | Outpatient; mean of 141.0 days post diagnosis | 55/100 | 55.00% |
| Myalgia | HP:0003326 | Muscle/joint pain | PMID:33825846 | Havervall | 2021 | Outpatient; mean of 120.0 days post diagnosis | 5/323 | 1.50% |
| Myalgia | HP:0003326 | Muscle/joint pain | PMID:33825846 | Havervall | 2021 | Outpatient; mean of 60.0 days post diagnosis | 5/323 | 1.50% |
| Myalgia | HP:0003326 | Muscle/joint pain | PMID:33825846 | Havervall | 2021 | Outpatient; mean of 240.0 days post diagnosis | 2/323 | 0.60% |
| Myalgia | HP:0003326 | Myalgia | PMID:33428867 | Huang | 2021 | Inpatient; mean of 186.0 days post discharge | 39/1655 | 2.40% |
| Myalgia | HP:0003326 | Muscular pain | PMID:33306721 | Jacobs | 2020 | Inpatient (non-ICU); mean of 35.0 days post discharge | 39/183 | 21.30% |
| Myalgia | HP:0003326 | Myalgias | PMID:33624010 | Jacobson | 2021 | Inpatient; mean of 119.3 days post discharge | 05/22/21 | 22.70% |
| Myalgia | HP:0003326 | Myalgias | PMID:33624010 | Jacobson | 2021 | Outpatient; mean of 119.3 days post diagnosis | 16/96 | 16.70% |
| Myalgia | HP:0003326 | Muscle or body aches | PMID:33606031 | Logue | 2021 | Inpatient; mean of 169.0 days post discharge | 01/16/21 | 6.30% |
| Myalgia | HP:0003326 | Muscle or body aches | PMID:33606031 | Logue | 2021 | Outpatient; mean of 169.0 days post diagnosis | 10/161 | 6.20% |
| Myalgia | HP:0003326 | muscle pain | PMID:33205450 | Ludvigsson | 2021 | Outpatient; mean of 210.0 days post diagnosis | 03/05/21 | 60.00% |
| Myalgia | HP:0003326 | Myalgias-arthralgias | PMID:33450302 | Moreno-Perez | 2021 | Mixed; mean of 77.0 days post diagnosis | 54/277 | 19.50% |
| Myalgia | HP:0003326 | Myalgia (Figure 1) | PMID:33252665 | Petersen | 2020 | Outpatient; mean of 99.0 days post diagnosis | 13/180 | 7.20% |
| Myalgia | HP:0003326 | Mialgias | PMID:33787016 | Prieto | 2021 | Mixed; mean of 53.0 days post diagnosis | 0/85 | 0.00% |
| Myalgia | HP:0003326 | Myalgia/persistent muscle pain | PMID:33682276 | Rass | 2021 | Outpatient; mean of 90.0 days post diagnosis | 2/32 | 6.30% |
| Myalgia | HP:0003326 | Myalgia/persistent musclepain | PMID:33682276 | Rass | 2021 | Inpatient (non-ICU); mean of 90.0 days post discharge | 6/72 | 8.30% |
| Myalgia | HP:0003326 | Myalgia/persistent musclepain | PMID:33682276 | Rass | 2021 | Inpatient (ICU); mean of 90.0 days post discharge | 6/31 | 19.40% |
| Myalgia | HP:0003326 | myalgia (13%) | PMID:33521308 | Rosales-Castillo | 2021 | Inpatient; mean of 50.8 days post discharge | 15/118 | 12.70% |
| Myalgia | HP:0003326 | Myalgia | PMID:33273028 | Stavem | 2020 | Outpatient; mean of 117.0 days post diagnosis | 36/451 | 8.00% |
| Myalgia | HP:0003326 | Myalgia | PMID:33413976 | Taboada | 2021 | Inpatient (ICU); mean of 180.0 days post discharge | 34/91 | 37.40% |
| Myalgia | HP:0003326 | Myalgia | PMID:33461632 | Venturelli | 2021 | Mixed; mean of 81.0 days post diagnosis | 29/515 | 5.60% |
| Myalgia | HP:0003326 | Myalgia | PMID:32979574 | Xiong | 2021 | Inpatient; mean of 97.0 days post discharge | 24/538 | 4.50% |
| Chest pain | HP:0100749 | Chest pain | PMID:33273026 | Arnold | 2020 | Inpatient (ICU); mean of 84.0 days post discharge | 02/18/21 | 11.10% |
| Chest pain | HP:0100749 | Chest pain | PMID:33273026 | Arnold | 2020 | Inpatient (non-ICU); mean of 84.0 days post discharge | 10/65 | 15.40% |
| Chest pain | HP:0100749 | Chest pain | PMID:33273026 | Arnold | 2020 | Inpatient (non-ICU); mean of 84.0 days post discharge | 02/27/21 | 7.40% |
| Chest pain | HP:0100749 | Chest pain At follow-up | PMID:33502487 | Bellan | 2021 | Inpatient; mean of 105.0 days post discharge | 1/238 | 0.40% |
| Chest pain | HP:0100749 | Chest pain | PMID:33502487 | Bellan | 2021 | Inpatient; mean of 105.0 days post discharge | 1/238 | 0.40% |
| Chest pain | HP:0100749 | Chest pain | PMID:32644129 | Carfi | 2020 | Inpatient; mean of 60.3 days post discharge | 31/143 | 21.70% |
| Chest pain | HP:0100749 | Chest pain | PMID:33031948 | Carvalho-Schneider | 2021 | Mixed; mean of 60.0 days post diagnosis | 17/130 | 13.10% |
| Chest pain | HP:0100749 | Chest pain | PMID:33031948 | Carvalho-Schneider | 2021 | Mixed; mean of 30.0 days post diagnosis | 27/150 | 18.00% |
| Chest pain | HP:0100749 | chest pain (26%) | PMID:33564789 | Chun | 2021 | Mixed; mean of 63.0 days post diagnosis | 16/61 | 26.20% |
| Chest pain | HP:0100749 | Chest pain | PMID:33657671 | Darley | 2021 | Mixed; mean of 69.0 days post diagnosis | 6/78 | 7.70% |
| Chest pain | HP:0100749 | Pain/burning in chest | PMID:-1 | Davis | 2020 | Mixed; mean of 114.5 days post diagnosis | 1997/3762 | 53.10% |
| Chest pain | HP:0100749 | Chest pain | PMID:33785495 | Dennis | 2021 | Inpatient; mean of 138.0 days post discharge | 24/37 | 64.90% |
| Chest pain | HP:0100749 | Chest pain | PMID:33785495 | Dennis | 2021 | Outpatient; mean of 141.0 days post diagnosis | 128/163 | 78.50% |
| Chest pain | HP:0100749 | Chest pain | PMID:32853602 | Garrigues | 2020 | Inpatient (non-ICU); mean of 110.9 days post discharge | 11/96 | 11.50% |
| Chest pain | HP:0100749 | Chest pain | PMID:32853602 | Garrigues | 2020 | Inpatient (ICU); mean of 110.9 days post discharge | 02/24/21 | 8.30% |
| Chest pain | HP:0100749 | Chest pain | PMID:33755344 | Graham | 2021 | Outpatient; mean of 141.0 days post diagnosis | 37/100 | 37.00% |
| Chest pain | HP:0100749 | Chest pain | PMID:33428867 | Huang | 2021 | Inpatient; mean of 186.0 days post discharge | 75/1655 | 4.50% |
| Chest pain | HP:0100749 | Chest pain | PMID:33680620 | Iqbal | 2021 | Mixed; mean of 38.1 days post diagnosis | 56/158 | 35.40% |
| Chest pain | HP:0100749 | Chest pain | PMID:33624010 | Jacobson | 2021 | Inpatient; mean of 119.3 days post discharge | 02/22/21 | 9.10% |
| Chest pain | HP:0100749 | Chest pain | PMID:33624010 | Jacobson | 2021 | Outpatient; mean of 119.3 days post diagnosis | 14/96 | 14.60% |
| Chest pain | HP:0100749 | chest pain | PMID:33205450 | Ludvigsson | 2021 | Outpatient; mean of 210.0 days post diagnosis | 05/05/21 | 100.00% |
| Chest pain | HP:0100749 | Dolor toracico 6% | PMID:33787016 | Prieto | 2021 | Mixed; mean of 53.0 days post diagnosis | 5/85 | 5.90% |
| Chest pain | HP:0100749 | atypical chest pain | PMID:32730619 | Puntmann | 2020 | Mixed; mean of 71.0 days post diagnosis | 17/100 | 17.00% |
| Chest pain | HP:0100749 | Chest pain | PMID:33413976 | Taboada | 2021 | Inpatient (ICU); mean of 180.0 days post discharge | 8/91 | 8.80% |
| Chest pain | HP:0100749 | Chest pain | PMID:32730238 | Tenforde | 2020 | Outpatient; mean of 17.5 days post diagnosis | 55/274 | 20.10% |
| Chest pain | HP:0100749 | thoracic pain | PMID:33479105 | Trinkmann | 2021 | Mixed; mean of 68.0 days post diagnosis | 15/246 | 6.10% |
| Chest pain | HP:0100749 | Chest pain | PMID:33461632 | Venturelli | 2021 | Mixed; mean of 81.0 days post diagnosis | 24/515 | 4.70% |
| Chest pain | HP:0100749 | Chest discomfort/pain | PMID:33729425 | Writing Committee | 2021 | Inpatient (ICU); mean of 93.0 days post discharge | 9/62 | 14.50% |
| Chest pain | HP:0100749 | Chest discomfort/pain | PMID:33729425 | Writing Committee | 2021 | Inpatient (non-ICU); mean of 121.0 days post discharge | 25/356 | 7.00% |
| Chest pain | HP:0100749 | Chest pain | PMID:32979574 | Xiong | 2021 | Inpatient; mean of 97.0 days post discharge | 66/538 | 12.30% |
| Chest pain | HP:0100749 | Chest pain | PMID:33532720 | de Graaf | 2021 | Inpatient (ICU); mean of 42.0 days post discharge | 8/34 | 23.50% |
| Chest pain | HP:0100749 | chest pain (all types) | PMID:33532720 | de Graaf | 2021 | Inpatient (non-ICU); mean of 42.0 days post discharge | 7/47 | 14.90% |
| Fever | HP:0001945 | Fever | PMID:33273026 | Arnold | 2020 | Inpatient (ICU); mean of 84.0 days post discharge | 0/18 | 0.00% |
| Fever | HP:0001945 | Fever | PMID:33273026 | Arnold | 2020 | Inpatient (non-ICU); mean of 84.0 days post discharge | 1/65 | 1.50% |
| Fever | HP:0001945 | Fever | PMID:33273026 | Arnold | 2020 | Inpatient (non-ICU); mean of 84.0 days post discharge | 0/27 | 0.00% |
| Fever | HP:0001945 | Fever at follow up | PMID:33502487 | Bellan | 2021 | Inpatient; mean of 105.0 days post discharge | 0/238 | 0.00% |
| Fever | HP:0001945 | Fever | PMID:33502487 | Bellan | 2021 | Inpatient; mean of 105.0 days post discharge | 0/238 | 0.00% |
| Fever | HP:0001945 | Fever (temperature >38°C) | PMID:33031948 | Carvalho-Schneider | 2021 | Mixed; mean of 60.0 days post diagnosis | 0/130 | 0.00% |
| Fever | HP:0001945 | Fever | PMID:33031948 | Carvalho-Schneider | 2021 | Mixed; mean of 30.0 days post diagnosis | 5/150 | 3.30% |
| Fever | HP:0001945 | fever (47%) | PMID:33564789 | Chun | 2021 | Mixed; mean of 63.0 days post diagnosis | 29/61 | 47.50% |

| | | | | | | | | |
|---|---|---|---|---|---|---|---|---|
| Fever | HP:0001945 | Fever | PMID:33120193 | Daher | 2020 | Inpatient (ICU); mean of 56.0 days post discharge | 1/33 | 3.00% |
| Fever | HP:0001945 | Fever/Chills | PMID:33657671 | Darley | 2021 | Mixed; mean of 69.0 days post diagnosis | 5/78 | 6.40% |
| Fever | HP:0001945 | Fever (>100.4 F) | PMID:-1 | Davis | 2020 | Mixed; mean of 114.5 days post diagnosis | 1158/3762 | 30.80% |
| Fever | HP:0001945 | Fever | PMID:33785495 | Dennis | 2021 | Inpatient; mean of 138.0 days post discharge | 31/37 | 83.80% |
| Fever | HP:0001945 | Fever | PMID:33785495 | Dennis | 2021 | Outpatient; mean of 141.0 days post diagnosis | 113/163 | 69.30% |
| Fever | HP:0001945 | Fever (body temperature above 38 degrees) (2%) | PMID:33257910 | Goertz | 2020 | Mixed; mean of 79.0 days post diagnosis | 42/21133 | 0.20% |
| Fever | HP:0001945 | Fever | PMID:33306721 | Jacobs | 2020 | Inpatient (non-ICU); mean of 35.0 days post discharge | 2/183 | 1.10% |
| Fever | HP:0001945 | Fever/Chills | PMID:33624010 | Jacobson | 2021 | Inpatient; mean of 119.3 days post discharge | 0/22 | 0.00% |
| Fever | HP:0001945 | Fever/chills | PMID:33624010 | Jacobson | 2021 | Outpatient; mean of 119.3 days post diagnosis | 1/96 | 1.00% |
| Fever | HP:0001945 | Fever | PMID:33289374 | Liang | 2020 | Inpatient (non-ICU); mean of 90.0 days post discharge | 15/76 | 19.70% |
| Fever | HP:0001945 | Feeling feverish | PMID:33606031 | Logue | 2021 | Inpatient; mean of 169.0 days post discharge | 0/16 | 0.00% |
| Fever | HP:0001945 | Feeling feverish | PMID:33606031 | Logue | 2021 | Outpatient; mean of 169.0 days post diagnosis | 4/161 | 2.50% |
| Fever | HP:0001945 | remitting fever | PMID:33205450 | Ludvigsson | 2021 | Outpatient; mean of 210.0 days post diagnosis | 02/05/21 | 40.00% |
| Fever | HP:0001945 | Fever | PMID:33450302 | Moreno-Perez | 2021 | Mixed; mean of 77.0 days post diagnosis | 0/277 | 0.00% |
| Fever | HP:0001945 | Fever (Figure 1) | PMID:33252665 | Petersen | 2020 | Outpatient; mean of 99.0 days post diagnosis | 0/180 | 0.00% |
| Fever | HP:0001945 | Fiebre | PMID:33787016 | Prieto | 2021 | Mixed; mean of 53.0 days post diagnosis | 0/85 | 0.00% |
| Fever | HP:0001945 | Fever | PMID:33303539 | Sonnweber | 2020 | Mixed; mean of 103.0 days post diagnosis | 0/145 | 0.00% |
| Fever | HP:0001945 | Fever | PMID:33273028 | Stavem | 2020 | Outpatient; mean of 117.0 days post diagnosis | 0/451 | 0.00% |
| Fever | HP:0001945 | Fever | PMID:32730238 | Tenforde | 2020 | Outpatient; mean of 17.5 days post diagnosis | 8/274 | 2.90% |
| Fever | HP:0001945 | pyrexia | PMID:33479105 | Trinkmann | 2021 | Mixed; mean of 68.0 days post diagnosis | 1/246 | 0.40% |
| Fever | HP:0001945 | Fever | PMID:33461632 | Venturelli | 2021 | Mixed; mean of 81.0 days post diagnosis | 4/515 | 0.80% |
| Diarrhea | HP:0002014 | Diarrhoea | PMID:33273026 | Arnold | 2020 | Inpatient (ICU); mean of 84.0 days post discharge | 0/18 | 0.00% |
| Diarrhea | HP:0002014 | Diarrhoea | PMID:33273026 | Arnold | 2020 | Inpatient (non-ICU); mean of 84.0 days post discharge | 1/65 | 1.50% |
| Diarrhea | HP:0002014 | Diarrhoea | PMID:33273026 | Arnold | 2020 | Inpatient (non-ICU); mean of 84.0 days post discharge | 0/27 | 0.00% |
| Diarrhea | HP:0002014 | Diarrhea At follow-up | PMID:33502487 | Bellan | 2021 | Inpatient; mean of 105.0 days post discharge | 3/238 | 1.30% |
| Diarrhea | HP:0002014 | Diarrhea | PMID:33502487 | Bellan | 2021 | Inpatient; mean of 105.0 days post discharge | 3/238 | 1.30% |
| Diarrhea | HP:0002014 | Diarrhea | PMID:32644129 | Carfi | 2020 | Inpatient; mean of 60.3 days post discharge | 4/143 | 2.80% |
| Diarrhea | HP:0002014 | Diarrhea | PMID:33564789 | Chun | 2021 | Mixed; mean of 63.0 days post diagnosis | 7/61 | 11.50% |
| Diarrhea | HP:0002014 | Diarrhea | PMID:33120193 | Daher | 2020 | Inpatient (ICU); mean of 56.0 days post discharge | 3/33 | 9.10% |
| Diarrhea | HP:0002014 | Diarrhoea | PMID:33657671 | Darley | 2021 | Mixed; mean of 69.0 days post diagnosis | 3/78 | 3.80% |
| Diarrhea | HP:0002014 | Diarrhea | PMID:-1 | Davis | 2020 | Mixed; mean of 114.5 days post diagnosis | 2246/3762 | 59.70% |
| Diarrhea | HP:0002014 | Diarrhoea | PMID:33785495 | Dennis | 2021 | Inpatient; mean of 138.0 days post discharge | 27/37 | 73.00% |
| Diarrhea | HP:0002014 | Diarrhoea | PMID:33785495 | Dennis | 2021 | Outpatient; mean of 141.0 days post diagnosis | 91/163 | 55.80% |
| Diarrhea | HP:0002014 | Diarrhoea (12%) | PMID:33257910 | Goertz | 2020 | Mixed; mean of 79.0 days post diagnosis | 211/2113 | 10.00% |
| Diarrhea | HP:0002014 | Diarrhea | PMID:33755344 | Graham | 2021 | Outpatient; mean of 141.0 days post diagnosis | 19/100 | 19.00% |
| Diarrhea | HP:0002014 | Diarrhoea or vomiting | PMID:33428867 | Huang | 2021 | Inpatient; mean of 186.0 days post discharge | 80/1655 | 4.80% |
| Diarrhea | HP:0002014 | Diarrhea | PMID:33306721 | Jacobs | 2020 | Inpatient (non-ICU); mean of 35.0 days post discharge | 7/183 | 3.80% |
| Diarrhea | HP:0002014 | Nausea/vomiting/diarrhea | PMID:33624010 | Jacobson | 2021 | Inpatient; mean of 119.3 days post discharge | 0/22 | 0.00% |
| Diarrhea | HP:0002014 | Nausea/vomiting/Diarrhea | PMID:33624010 | Jacobson | 2021 | Outpatient; mean of 119.3 days post diagnosis | 8/96 | 8.30% |
| Diarrhea | HP:0002014 | Diarrhea | PMID:33289374 | Liang | 2020 | Inpatient (non-ICU); mean of 90.0 days post discharge | 20/76 | 26.30% |
| Diarrhea | HP:0002014 | Diarrhea | PMID:33606031 | Logue | 2021 | Inpatient; mean of 169.0 days post discharge | 0/16 | 0.00% |
| Diarrhea | HP:0002014 | Diarrhea | PMID:33606031 | Logue | 2021 | Outpatient; mean of 169.0 days post diagnosis | 5/161 | 3.10% |
| Diarrhea | HP:0002014 | diarrhoea | PMID:33205450 | Ludvigsson | 2021 | Outpatient; mean of 210.0 days post diagnosis | 02/05/21 | 40.00% |
| Diarrhea | HP:0002014 | Diarrhoea | PMID:33450302 | Moreno-Perez | 2021 | Mixed; mean of 77.0 days post diagnosis | 29/277 | 10.50% |
| Diarrhea | HP:0002014 | Diarrhea (Figure 1) | PMID:33252665 | Petersen | 2020 | Outpatient; mean of 99.0 days post diagnosis | 5/180 | 2.80% |
| Diarrhea | HP:0002014 | Diarrea | PMID:33787016 | Prieto | 2021 | Mixed; mean of 53.0 days post diagnosis | 0/85 | 0.00% |
| Diarrhea | HP:0002014 | Diarrhea or vomiting | PMID:33303539 | Sonnweber | 2020 | Mixed; mean of 103.0 days post diagnosis | 13/145 | 9.00% |
| Diarrhea | HP:0002014 | Diarrhoea | PMID:33273028 | Stavem | 2020 | Outpatient; mean of 117.0 days post diagnosis | 9/451 | 2.00% |
| Diarrhea | HP:0002014 | Diarrhea | PMID:32730238 | Tenforde | 2020 | Outpatient; mean of 17.5 days post diagnosis | 33/274 | 12.00% |
| Diarrhea | HP:0002014 | diarrhoea | PMID:33479105 | Trinkmann | 2021 | Mixed; mean of 68.0 days post diagnosis | 0/246 | 0.00% |
| Depression | HP:0000716 | Anxiety/depression | PMID:33113469 | Akter | 2020 | Inpatient; mean of 28.0 days post discharge | 159/734 | 21.70% |
| Depression | HP:0000716 | 6% of patients had GAD-2 scores of >3 (6%), representing a positive screen for depression and anxiety respectively | PMID:33731329 | Cheng | 2021 | Mixed; mean of 63.0 days post diagnosis | 17/113 | 15.00% |
| Depression | HP:0000716 | The Depression in the Medically Ill questionnaire (DMI-10) 6 was administered to 76 patients; 16 (21%) reported symptoms consistent with depression. | PMID:33657671 | Darley | 2021 | Mixed; mean of 69.0 days post diagnosis | 16/76 | 21.10% |
| Depression | HP:0000716 | Depression | PMID:-1 | Davis | 2020 | Mixed; mean of 114.5 days post diagnosis | 1779/3762 | 47.30% |
| Depression | HP:0000716 | HADS Depression score abnormal | PMID:33676998 | González | 2021 | Inpatient (ICU); mean of 90.0 days post discharge | 3/59 | 5.10% |
| Depression | HP:0000716 | Depression/Anxiety | PMID:33755344 | Graham | 2021 | Outpatient; mean of 141.0 days post diagnosis | 47/100 | 47.00% |
| Depression | HP:0000716 | Worsened anxiety/depression | PMID:32729939 | Halpin | 2021 | Inpatient (ICU); mean of 48.0 days post discharge | 12/32 | 37.50% |
| Depression | HP:0000716 | Worsened anxiety/depression | PMID:32729939 | Halpin | 2021 | Inpatient (non-ICU); mean of 48.0 days post discharge | 11/68 | 16.20% |
| Depression | HP:0000716 | Anxiety or depression | PMID:33428867 | Huang | 2021 | Inpatient; mean of 186.0 days post discharge | 367/1617 | 22.70% |

| | | | | | | | | |
|---|---|---|---|---|---|---|---|---|
| Depression | HP:0000716 | anxious or depressed | PMID:33680620 | Iqbal | 2021 | Mixed; mean of 38.1 days post diagnosis | 95/158 | 60.10% |
| Depression | HP:0000716 | depression | PMID:33205450 | Ludvigsson | 2021 | Outpatient; mean of 210.0 days post diagnosis | 03/05/21 | 60.00% |
| Depression | HP:0000716 | Fifteen per cent were depressed | PMID:33172844 | Mandal | 2020 | Inpatient; mean of 54.0 days post discharge | 58/383 | 15.10% |
| Depression | HP:0000716 | Anxiety/Depression | PMID:33865161 | Ordinola Navarro | 2021 | Mixed; mean of 40.0 days post diagnosis | 60/115 | 52.20% |
| Depression | HP:0000716 | At follow-up, seven patients had mild to moderate depression. | PMID:32835708 | Ramani | 2021 | Inpatient (ICU); mean of 39.5 days post discharge | 07/28/21 | 25.00% |
| Depression | HP:0000716 | PHQ-9, moderate-severe | PMID:33490928 | Ramani | 2021 | Inpatient; mean of 48.0 days post discharge | 11/57 | 19.30% |
| Depression | HP:0000716 | Moderately anxious or depressed/Extremely anxious or depressed | PMID:33413976 | Taboada | 2021 | Inpatient (ICU); mean of 180.0 days post discharge | 42/91 | 46.20% |
| Depression | HP:0000716 | HADS-Depression | PMID:33461632 | Venturelli | 2021 | Mixed; mean of 81.0 days post diagnosis | 33/727 | 4.50% |
| Depression | HP:0000716 | Symptoms of depression (BDI test) | PMID:33729425 | Writing Committee | 2021 | Inpatient (ICU); mean of 93.0 days post discharge | 9/50 | 18.00% |
| Depression | HP:0000716 | Symptoms of depression (BDI test) | PMID:33729425 | Writing Committee | 2021 | Inpatient (non-ICU); mean of 121.0 days post discharge | 26/120 | 21.70% |
| Depression | HP:0000716 | Depression | PMID:32979574 | Xiong | 2021 | Inpatient; mean of 97.0 days post discharge | 23/538 | 4.30% |
| Depression | HP:0000716 | Depression (PHQ-9 ≥ 10) | PMID:33532720 | de Graaf | 2021 | Inpatient (ICU); mean of 42.0 days post discharge | 4/34 | 11.80% |
| Depression | HP:0000716 | Depression (PHQ-9 ≥ 10) | PMID:33532720 | de Graaf | 2021 | Inpatient (non-ICU); mean of 42.0 days post discharge | 6/47 | 12.80% |
| Depression | HP:0000716 | HADS-depression >10,No. (%) | PMID:33220049 | van den Borst | 2020 | Inpatient (ICU); mean of 91.0 days post discharge | 4/46 | 8.70% |
| Depression | HP:0000716 | HADS-depression >10,No. (%) | PMID:33220049 | van den Borst | 2020 | Outpatient; mean of 91.0 days post diagnosis | 06/27/21 | 22.20% |
| Depression | HP:0000716 | HADS-depression >10,No. (%) | PMID:33220049 | van den Borst | 2020 | Inpatient (non-ICU); mean of 91.0 days post discharge | 4/51 | 7.80% |
| Arthralgia | HP:0002829 | Arthralgia | PMID:33273026 | Arnold | 2020 | Inpatient (ICU); mean of 84.0 days post discharge | 03/18/21 | 16.70% |
| Arthralgia | HP:0002829 | Arthralgia | PMID:33273026 | Arnold | 2020 | Inpatient (non-ICU); mean of 84.0 days post discharge | 1/65 | 1.50% |
| Arthralgia | HP:0002829 | Arthralgia | PMID:33273026 | Arnold | 2020 | Inpatient (non-ICU); mean of 84.0 days post discharge | 01/27/21 | 3.70% |
| Arthralgia | HP:0002829 | Arthralgia At follow-up | PMID:33502487 | Bellan | 2021 | Inpatient; mean of 105.0 days post discharge | 14/238 | 5.90% |
| Arthralgia | HP:0002829 | Arthralgia | PMID:33502487 | Bellan | 2021 | Inpatient; mean of 105.0 days post discharge | 14/238 | 5.90% |
| Arthralgia | HP:0002829 | Joint pain | PMID:32644129 | Carfi | 2020 | Inpatient; mean of 60.3 days post discharge | 39/143 | 27.30% |
| Arthralgia | HP:0002829 | Arthralgia | PMID:33031948 | Carvalho-Schneider | 2021 | Mixed; mean of 60.0 days post diagnosis | 21/130 | 16.20% |
| Arthralgia | HP:0002829 | Arthralgia | PMID:33031948 | Carvalho-Schneider | 2021 | Mixed; mean of 30.0 days post diagnosis | 13/150 | 8.70% |
| Arthralgia | HP:0002829 | Joint pain | PMID:-1 | Davis | 2020 | Mixed; mean of 114.5 days post diagnosis | 1962/3762 | 52.20% |
| Arthralgia | HP:0002829 | Joint pain | PMID:33785495 | Dennis | 2021 | Inpatient; mean of 138.0 days post discharge | 29/37 | 78.40% |
| Arthralgia | HP:0002829 | Joint pain | PMID:33785495 | Dennis | 2021 | Outpatient; mean of 141.0 days post diagnosis | 127/163 | 77.90% |
| Arthralgia | HP:0002829 | Joint pain (27%) | PMID:33257910 | Goertz | 2020 | Mixed; mean of 79.0 days post diagnosis | 465/2113 | 22.00% |
| Arthralgia | HP:0002829 | Muscle/joint pain | PMID:33825846 | Havervall | 2021 | Outpatient; mean of 120.0 days post diagnosis | 5/323 | 1.50% |
| Arthralgia | HP:0002829 | Muscle/joint pain | PMID:33825846 | Havervall | 2021 | Outpatient; mean of 60.0 days post diagnosis | 5/323 | 1.50% |
| Arthralgia | HP:0002829 | Muscle/joint pain | PMID:33825846 | Havervall | 2021 | Outpatient; mean of 240.0 days post diagnosis | 2/323 | 0.60% |
| Arthralgia | HP:0002829 | Joint pain | PMID:33428867 | Huang | 2021 | Inpatient; mean of 186.0 days post discharge | 154/1655 | 9.30% |
| Arthralgia | HP:0002829 | Joint pain | PMID:33680620 | Iqbal | 2021 | Mixed; mean of 38.1 days post diagnosis | 75/158 | 47.50% |
| Arthralgia | HP:0002829 | Joint pain | PMID:33306721 | Jacobs | 2020 | Inpatient (non-ICU); mean of 35.0 days post discharge | 29/183 | 15.80% |
| Arthralgia | HP:0002829 | joint pain | PMID:33205450 | Ludvigsson | 2021 | Outpatient; mean of 210.0 days post diagnosis | 02/05/21 | 40.00% |
| Arthralgia | HP:0002829 | Myalgias-arthralgias | PMID:33450302 | Moreno-Perez | 2021 | Mixed; mean of 77.0 days post diagnosis | 54/277 | 19.50% |
| Arthralgia | HP:0002829 | Arthralgia (Figure 1) | PMID:33252665 | Petersen | 2020 | Outpatient; mean of 99.0 days post diagnosis | 20/180 | 11.10% |
| Arthralgia | HP:0002829 | Arthralgia | PMID:33273028 | Stavem | 2020 | Outpatient; mean of 117.0 days post diagnosis | 41/451 | 9.10% |
| Arthralgia | HP:0002829 | Arthralgia | PMID:33413976 | Taboada | 2021 | Inpatient (ICU); mean of 180.0 days post discharge | 26/91 | 28.60% |
| Arthralgia | HP:0002829 | Arthralgia | PMID:32979574 | Xiong | 2021 | Inpatient; mean of 97.0 days post discharge | 41/538 | 7.60% |
| Anxiety | HP:0000739 | Anxiety/depression | PMID:33113469 | Akter | 2020 | Inpatient; mean of 28.0 days post discharge | 159/734 | 21.70% |
| Anxiety | HP:0000739 | 6% of patients had GAD-2 scores of >3 (6%), representing a positive screen for depression and anxiety respectively | PMID:33731329 | Cheng | 2021 | Mixed; mean of 63.0 days post diagnosis | 7/113 | 6.20% |
| Anxiety | HP:0000739 | Anxiety | PMID:-1 | Davis | 2020 | Mixed; mean of 114.5 days post diagnosis | 2179/3762 | 57.90% |
| Anxiety | HP:0000739 | Anxiety | PMID:33052920 | De Lorenzo | 2020 | Inpatient; mean of 21.5 days post discharge | 32/126 | 25.40% |
| Anxiety | HP:0000739 | Anxiety | PMID:33052920 | De Lorenzo | 2020 | Outpatient; mean of 26.0 days post diagnosis | 23/59 | 39.00% |
| Anxiety | HP:0000739 | HADS Anxiety score abnormal | PMID:33676998 | González | 2021 | Inpatient (ICU); mean of 90.0 days post discharge | 6/59 | 10.20% |
| Anxiety | HP:0000739 | Depression/Anxiety | PMID:33755344 | Graham | 2021 | Outpatient; mean of 141.0 days post diagnosis | 47/100 | 47.00% |
| Anxiety | HP:0000739 | Worsened anxiety/depression | PMID:32729939 | Halpin | 2021 | Inpatient (ICU); mean of 48.0 days post discharge | 12/32 | 37.50% |
| Anxiety | HP:0000739 | Worsened anxiety/depression | PMID:32729939 | Halpin | 2021 | Inpatient (non-ICU); mean of 48.0 days post discharge | 11/68 | 16.20% |
| Anxiety | HP:0000739 | Anxiety or depression | PMID:33428867 | Huang | 2021 | Inpatient; mean of 186.0 days post discharge | 367/1617 | 22.70% |
| Anxiety | HP:0000739 | anxious or depressed | PMID:33680620 | Iqbal | 2021 | Mixed; mean of 38.1 days post diagnosis | 95/158 | 60.10% |
| Anxiety | HP:0000739 | Anxiety/Depression | PMID:33865161 | Ordinola Navarro | 2021 | Mixed; mean of 40.0 days post diagnosis | 60/115 | 52.20% |
| Anxiety | HP:0000739 | GAD-7, moderate - severe | PMID:33490928 | Ramani | 2021 | Inpatient; mean of 48.0 days post discharge | 8/57 | 14.00% |
| Anxiety | HP:0000739 | Anxiety | PMID:33654513 | Suárez-Robles | 2020 | Inpatient; mean of 90.0 days post discharge | 76/134 | 56.70% |
| Anxiety | HP:0000739 | Moderately anxious or depressed/Extremely anxious or depressed | PMID:33413976 | Taboada | 2021 | Inpatient (ICU); mean of 180.0 days post discharge | 42/91 | 46.20% |
| Anxiety | HP:0000739 | HADS-Anxiety | PMID:33461632 | Venturelli | 2021 | Mixed; mean of 81.0 days post diagnosis | 82/727 | 11.30% |
| Anxiety | HP:0000739 | Symptoms of anxiety (HADS-Anxiety) | PMID:33729425 | Writing Committee | 2021 | Inpatient (ICU); mean of 93.0 days post discharge | 13/50 | 26.00% |
| Anxiety | HP:0000739 | Symptoms of anxiety (HADS-Anxiety) | PMID:33729425 | Writing Committee | 2021 | Inpatient (non-ICU); mean of 121.0 days post discharge | 40/119 | 33.60% |
| Anxiety | HP:0000739 | Anxiety | PMID:32979574 | Xiong | 2021 | Inpatient; mean of 97.0 days post discharge | 35/538 | 6.50% |

| | | | | | | | | |
|---|---|---|---|---|---|---|---|---|
| Anxiety | HP:0000739 | Anxiety (GAD-7 ≥ 10) | PMID:33532720 | de Graaf | 2021 | Inpatient (ICU); mean of 42.0 days post discharge | 1/34 | 2.90% |
| Anxiety | HP:0000739 | Anxiety (GAD-7 ≥ 10), | PMID:33532720 | de Graaf | 2021 | Inpatient (non-ICU); mean of 42.0 days post discharge | 2/47 | 4.30% |
| Anxiety | HP:0000739 | HADS-anxiety >10, No.(%) | PMID:33220049 | van den Borst | 2020 | Inpatient (ICU); mean of 91.0 days post discharge | 4/46 | 8.70% |
| Anxiety | HP:0000739 | HADS-anxiety >10, No.(%) | PMID:33220049 | van den Borst | 2020 | Outpatient; mean of 91.0 days post diagnosis | 02/27/21 | 7.40% |
| Anxiety | HP:0000739 | HADS-anxiety >10, No.(%) | PMID:33220049 | van den Borst | 2020 | Inpatient (non-ICU); mean of 91.0 days post discharge | 6/51 | 11.80% |
| Pharyngalgia | HP:0033050 | Sore throat At follow-up | PMID:33502487 | Bellan | 2021 | Inpatient; mean of 105.0 days post discharge | 0/238 | 0.00% |
| Pharyngalgia | HP:0033050 | Sore throat | PMID:33502487 | Bellan | 2021 | Inpatient; mean of 105.0 days post discharge | 0/238 | 0.00% |
| Pharyngalgia | HP:0033050 | Sore throat | PMID:32644129 | Carfi | 2020 | Inpatient; mean of 60.3 days post discharge | 10/143 | 7.00% |
| Pharyngalgia | HP:0033050 | Sore throat | PMID:33564789 | Chun | 2021 | Mixed; mean of 63.0 days post diagnosis | 1/61 | 1.60% |
| Pharyngalgia | HP:0033050 | Sore throat 3 (note authors state Pharyngalgia 0, we will curate 3). | PMID:33120193 | Daher | 2020 | Inpatient (ICU); mean of 56.0 days post discharge | 3/33 | 9.10% |
| Pharyngalgia | HP:0033050 | Sore throat | PMID:33657671 | Darley | 2021 | Mixed; mean of 69.0 days post diagnosis | 1/78 | 1.30% |
| Pharyngalgia | HP:0033050 | Sore Throat | PMID:-1 | Davis | 2020 | Mixed; mean of 114.5 days post diagnosis | 2239/3762 | 59.50% |
| Pharyngalgia | HP:0033050 | Sore throat | PMID:33785495 | Dennis | 2021 | Inpatient; mean of 138.0 days post discharge | 23/37 | 62.20% |
| Pharyngalgia | HP:0033050 | Sore throat | PMID:33785495 | Dennis | 2021 | Outpatient; mean of 141.0 days post diagnosis | 120/163 | 73.60% |
| Pharyngalgia | HP:0033050 | Sore throat (26%) | PMID:33257910 | Goertz | 2020 | Mixed; mean of 79.0 days post diagnosis | 549/2113 | 26.00% |
| Pharyngalgia | HP:0033050 | Sore throat or difficult to swallow | PMID:33428867 | Huang | 2021 | Inpatient; mean of 186.0 days post discharge | 69/1655 | 4.20% |
| Pharyngalgia | HP:0033050 | Sore throat | PMID:33624010 | Jacobson | 2021 | Inpatient; mean of 119.3 days post discharge | 0/22 | 0.00% |
| Pharyngalgia | HP:0033050 | Sore throat | PMID:33624010 | Jacobson | 2021 | Outpatient; mean of 119.3 days post diagnosis | 3/96 | 3.10% |
| Pharyngalgia | HP:0033050 | Throat discomfort | PMID:32692945 | Liu | 2020 | Inpatient; mean of 28.0 days post discharge | 3/51 | 5.90% |
| Pharyngalgia | HP:0033050 | Sore throat | PMID:33606031 | Logue | 2021 | Inpatient; mean of 169.0 days post discharge | 01/16/21 | 6.30% |
| Pharyngalgia | HP:0033050 | Sore throat | PMID:33606031 | Logue | 2021 | Outpatient; mean of 169.0 days post diagnosis | 5/161 | 3.10% |
| Pharyngalgia | HP:0033050 | sore throat | PMID:33205450 | Ludvigsson | 2021 | Outpatient; mean of 210.0 days post diagnosis | 04/05/21 | 80.00% |
| Pharyngalgia | HP:0033050 | Sore throat (Figure 1) | PMID:33252665 | Petersen | 2020 | Outpatient; mean of 99.0 days post diagnosis | 4/180 | 2.20% |
| Pharyngalgia | HP:0033050 | odinofagia | PMID:33787016 | Prieto | 2021 | Mixed; mean of 53.0 days post diagnosis | 0/85 | 0.00% |
| Pharyngalgia | HP:0033050 | Sore throat | PMID:33273028 | Stavem | 2020 | Outpatient; mean of 117.0 days post diagnosis | 23/451 | 5.10% |
| Pharyngalgia | HP:0033050 | Sore throat | PMID:32730238 | Tenforde | 2020 | Outpatient; mean of 17.5 days post diagnosis | 47/274 | 17.20% |
| Pharyngalgia | HP:0033050 | sore throat | PMID:33479105 | Trinkmann | 2021 | Mixed; mean of 68.0 days post diagnosis | 1/246 | 0.40% |
| Pharyngalgia | HP:0033050 | Throat pain | PMID:32979574 | Xiong | 2021 | Inpatient; mean of 97.0 days post discharge | 17/538 | 3.20% |
| Ageusia | HP:0041051 | Ageusia At follow-up | PMID:33502487 | Bellan | 2021 | Inpatient; mean of 105.0 days post discharge | 12/238 | 5.00% |
| Ageusia | HP:0041051 | Ageusia | PMID:33502487 | Bellan | 2021 | Inpatient; mean of 105.0 days post discharge | 12/238 | 5.00% |
| Ageusia | HP:0041051 | Anosmia/ageusia | PMID:33031948 | Carvalho-Schneider | 2021 | Mixed; mean of 60.0 days post diagnosis | 29/130 | 22.30% |
| Ageusia | HP:0041051 | Anosmia/ageusia | PMID:33031948 | Carvalho-Schneider | 2021 | Mixed; mean of 30.0 days post diagnosis | 40/150 | 26.70% |
| Ageusia | HP:0041051 | Continued loss of taste and/or smell | PMID:33175566 | Chopra | 2020 | Inpatient; mean of 60.0 days post discharge | 64/488 | 13.10% |
| Ageusia | HP:0041051 | Loss of Taste | PMID:33120193 | Daher | 2020 | Inpatient (ICU); mean of 56.0 days post discharge | 3/33 | 9.10% |
| Ageusia | HP:0041051 | Ageusia | PMID:33657671 | Darley | 2021 | Mixed; mean of 69.0 days post diagnosis | 1/78 | 1.30% |
| Ageusia | HP:0041051 | Loss of taste | PMID:-1 | Davis | 2020 | Mixed; mean of 114.5 days post diagnosis | 1267/3762 | 33.70% |
| Ageusia | HP:0041051 | Ageusia | PMID:32853602 | Garrigues | 2020 | Inpatient (non-ICU); mean of 110.9 days post discharge | 9/96 | 9.40% |
| Ageusia | HP:0041051 | Ageusia | PMID:32853602 | Garrigues | 2020 | Inpatient (ICU); mean of 110.9 days post discharge | 04/24/21 | 16.70% |
| Ageusia | HP:0041051 | Ageusia (11%) | PMID:33257910 | Goertz | 2020 | Mixed; mean of 79.0 days post diagnosis | 232/2113 | 11.00% |
| Ageusia | HP:0041051 | Ageusia | PMID:33825846 | Havervall | 2021 | Outpatient; mean of 120.0 days post diagnosis | 17/323 | 5.30% |
| Ageusia | HP:0041051 | Ageusia | PMID:33825846 | Havervall | 2021 | Outpatient; mean of 60.0 days post diagnosis | 25/323 | 7.70% |
| Ageusia | HP:0041051 | Ageusia | PMID:33825846 | Havervall | 2021 | Outpatient; mean of 240.0 days post diagnosis | 12/323 | 3.70% |
| Ageusia | HP:0041051 | Loss of smell and taste | PMID:33680620 | Iqbal | 2021 | Mixed; mean of 38.1 days post diagnosis | 75/158 | 47.50% |
| Ageusia | HP:0041051 | Lack of taste | PMID:33306721 | Jacobs | 2020 | Inpatient (non-ICU); mean of 35.0 days post discharge | 18/183 | 9.80% |
| Ageusia | HP:0041051 | Loss of taste/smell | PMID:33624010 | Jacobson | 2021 | Inpatient; mean of 119.3 days post discharge | 02/22/21 | 9.10% |
| Ageusia | HP:0041051 | Loss of taste/smell | PMID:33624010 | Jacobson | 2021 | Outpatient; mean of 119.3 days post diagnosis | 23/96 | 24.00% |
| Ageusia | HP:0041051 | Loss of taste (Figure 1) | PMID:33252665 | Petersen | 2020 | Outpatient; mean of 99.0 days post diagnosis | 27/180 | 15.00% |
| Ageusia | HP:0041051 | ageusia (1%) | PMID:33521308 | Rosales-Castillo | 2021 | Inpatient; mean of 50.8 days post discharge | 1/118 | 0.80% |
| Ageusia | HP:0041051 | Loss of taste | PMID:32730238 | Tenforde | 2020 | Outpatient; mean of 17.5 days post diagnosis | 63/274 | 23.00% |
| Palpitations | HP:0001962 | Palpitations | PMID:33031948 | Carvalho-Schneider | 2021 | Mixed; mean of 60.0 days post diagnosis | 14/130 | 10.80% |
| Palpitations | HP:0001962 | Palpitations | PMID:33031948 | Carvalho-Schneider | 2021 | Mixed; mean of 30.0 days post diagnosis | 9/150 | 6.00% |
| Palpitations | HP:0001962 | Heart palpitations | PMID:-1 | Davis | 2020 | Mixed; mean of 114.5 days post diagnosis | 2534/3762 | 67.40% |
| Palpitations | HP:0001962 | Heart palpitations (32%) | PMID:33257910 | Goertz | 2020 | Mixed; mean of 79.0 days post diagnosis | 676/2113 | 32.00% |
| Palpitations | HP:0001962 | Palpitations | PMID:33825846 | Havervall | 2021 | Outpatient; mean of 120.0 days post diagnosis | 7/323 | 2.20% |
| Palpitations | HP:0001962 | Palpitations | PMID:33825846 | Havervall | 2021 | Outpatient; mean of 60.0 days post diagnosis | 8/323 | 2.50% |
| Palpitations | HP:0001962 | Palpitations | PMID:33825846 | Havervall | 2021 | Outpatient; mean of 240.0 days post diagnosis | 2/323 | 0.60% |
| Palpitations | HP:0001962 | Palpitations | PMID:33428867 | Huang | 2021 | Inpatient; mean of 186.0 days post discharge | 154/1655 | 9.30% |
| Palpitations | HP:0001962 | Palpitations | PMID:33624010 | Jacobson | 2021 | Inpatient; mean of 119.3 days post discharge | 0/22 | 0.00% |
| Palpitations | HP:0001962 | Palpitations | PMID:33624010 | Jacobson | 2021 | Outpatient; mean of 119.3 days post diagnosis | 7/96 | 7.30% |
| Palpitations | HP:0001962 | Palpitations on exertion | PMID:33289374 | Liang | 2020 | Inpatient (non-ICU); mean of 90.0 days post discharge | 47/76 | 61.80% |
| Palpitations | HP:0001962 | heart palpitations | PMID:33205450 | Ludvigsson | 2021 | Outpatient; mean of 210.0 days post diagnosis | 05/05/21 | 100.00% |

| | | | | | | | | |
|---|---|---|---|---|---|---|---|---|
| Palpitations | HP:0001962 | Palpitationes 20% | PMID:33787016 | Prieto | 2021 | Mixed; mean of 53.0 days post diagnosis | 17/85 | 20.00% |
| Palpitations | HP:0001962 | Palpitations | PMID:32730619 | Puntmann | 2020 | Mixed; mean of 71.0 days post diagnosis | 20/100 | 20.00% |
| Palpitations | HP:0001962 | Palpitations | PMID:33654513 | Suárez-Robles | 2020 | Inpatient; mean of 90.0 days post discharge | 29/134 | 21.60% |
| Palpitations | HP:0001962 | Palpitations | PMID:33461632 | Venturelli | 2021 | Mixed; mean of 81.0 days post diagnosis | 30/515 | 5.80% |
| Palpitations | HP:0001962 | Palpitations | PMID:33532720 | de Graaf | 2021 | Inpatient (ICU); mean of 42.0 days post discharge | 5/34 | 14.70% |
| Palpitations | HP:0001962 | palpitations | PMID:33532720 | de Graaf | 2021 | Inpatient (non-ICU); mean of 42.0 days post discharge | 7/47 | 14.90% |
| Decreased DLCO | HP:0045051 | DLCO < LLN, n (%) | PMID:33872135 | Abdallah | 2021 | Inpatient; mean of 102.3 days post discharge | 17/25 | 68.00% |
| Decreased DLCO | HP:0045051 | DLCO < LLN, n (%) | PMID:33872135 | Abdallah | 2021 | Outpatient; mean of 129.8 days post diagnosis | 12/38 | 31.60% |
| Decreased DLCO | HP:0045051 | Dlco was less than 80% of expected in 113 patients | PMID:33502487 | Bellan | 2021 | Inpatient; mean of 105.0 days post discharge | 113/224 | 50.40% |
| Decreased DLCO | HP:0045051 | Dlco was less than 80% of expected in 113 patients | PMID:33502487 | Bellan | 2021 | Inpatient; mean of 105.0 days post discharge | 113/224 | 50.40% |
| Decreased DLCO | HP:0045051 | DLCO <80 | PMID:33662544 | Blanco | 2021 | Inpatient; mean of 104.0 days post discharge | 52/100 | 52.00% |
| Decreased DLCO | HP:0045051 | Diffusion capacity for carbon monoxide was below the lower limit of normal in 11 patients (14%) | PMID:33657671 | Darley | 2021 | Mixed; mean of 69.0 days post diagnosis | 11/65 | 16.90% |
| Decreased DLCO | HP:0045051 | DLCO, mL/min/mm Hg < 80% | PMID:33676998 | González | 2021 | Inpatient (ICU); mean of 90.0 days post discharge | 50/62 | 80.60% |
| Decreased DLCO | HP:0045051 | DLCO <80%, % of predicted | PMID:33428867 | Huang | 2021 | Inpatient; mean of 186.0 days post discharge | 114/334 | 34.10% |
| Decreased DLCO | HP:0045051 | 15 (47%) had decreased DLCO levels | PMID:33289374 | Liang | 2020 | Inpatient (non-ICU); mean of 90.0 days post discharge | 15/76 | 19.70% |
| Decreased DLCO | HP:0045051 | seven (26.92%) had reduced diffusion capacity | PMID:32835708 | Ramani | 2021 | Inpatient (ICU); mean of 39.5 days post discharge | 07/28/21 | 25.00% |
| Decreased DLCO | HP:0045051 | DLCO <80% of predicted normal – no. (%) | PMID:33303539 | Sonnweber | 2020 | Mixed; mean of 63.0 days post diagnosis | 39/126 | 31.00% |
| Decreased DLCO | HP:0045051 | DLCO <80% of predicted normal – no. (%) | PMID:33303539 | Sonnweber | 2020 | Mixed; mean of 103.0 days post diagnosis | 28/133 | 21.10% |
| Decreased DLCO | HP:0045051 | DLCO <70% | PMID:33729425 | Writing Committee | 2021 | Inpatient (ICU); mean of 93.0 days post discharge | 17/47 | 36.20% |
| Decreased DLCO | HP:0045051 | DLCO <70% | PMID:33729425 | Writing Committee | 2021 | Inpatient (non-ICU); mean of 121.0 days post discharge | 16/105 | 15.20% |
| Decreased DLCO | HP:0045051 | DLCO impaired group (Supplementary Table 5) | PMID:32838236 | Zhao | 2020 | Inpatient; mean of 78.5 days post discharge | 9/55 | 16.40% |
| Decreased DLCO | HP:0045051 | DLCO<LLN, No. (%) | PMID:33220049 | van den Borst | 2020 | Inpatient (ICU); mean of 91.0 days post discharge | 25/46 | 54.30% |
| Decreased DLCO | HP:0045051 | DLCO<LLN, No. (%) | PMID:33220049 | van den Borst | 2020 | Outpatient; mean of 91.0 days post diagnosis | 0/27 | 0.00% |
| Decreased DLCO | HP:0045051 | DLCO<LLN, No. (%) | PMID:33220049 | van den Borst | 2020 | Inpatient (non-ICU); mean of 91.0 days post discharge | 16/51 | 31.40% |
| Nausea | HP:0002018 | Nausea | PMID:33273026 | Arnold | 2020 | Inpatient (ICU); mean of 84.0 days post discharge | 0/18 | 0.00% |
| Nausea | HP:0002018 | Nausea | PMID:33273026 | Arnold | 2020 | Inpatient (non-ICU); mean of 84.0 days post discharge | 0/65 | 0.00% |
| Nausea | HP:0002018 | Nausea | PMID:33273026 | Arnold | 2020 | Inpatient (non-ICU); mean of 84.0 days post discharge | 0/27 | 0.00% |
| Nausea | HP:0002018 | Nausea | PMID:33564789 | Chun | 2021 | Mixed; mean of 63.0 days post diagnosis | 10/61 | 16.40% |
| Nausea | HP:0002018 | Nausea | PMID:33120193 | Daher | 2020 | Inpatient (ICU); mean of 56.0 days post discharge | 2/33 | 6.10% |
| Nausea | HP:0002018 | Nausea/vomiting | PMID:33657671 | Darley | 2021 | Mixed; mean of 69.0 days post diagnosis | 3/78 | 3.80% |
| Nausea | HP:0002018 | Nausea | PMID:-1 | Davis | 2020 | Mixed; mean of 114.5 days post diagnosis | 1797/3762 | 47.80% |
| Nausea | HP:0002018 | Nausea (12%) | PMID:33257910 | Goertz | 2020 | Mixed; mean of 79.0 days post diagnosis | 254/2113 | 12.00% |
| Nausea | HP:0002018 | nausea | PMID:33755344 | Graham | 2021 | Outpatient; mean of 141.0 days post diagnosis | 12/100 | 12.00% |
| Nausea | HP:0002018 | Nausea/vomiting/diarrhea | PMID:33624010 | Jacobson | 2021 | Inpatient; mean of 119.3 days post discharge | 0/22 | 0.00% |
| Nausea | HP:0002018 | Nausea/vomiting/Diarrhea | PMID:33624010 | Jacobson | 2021 | Outpatient; mean of 119.3 days post diagnosis | 8/96 | 8.30% |
| Nausea | HP:0002018 | Nausea | PMID:33606031 | Logue | 2021 | Inpatient; mean of 169.0 days post discharge | 0/16 | 0.00% |
| Nausea | HP:0002018 | Nausea | PMID:33606031 | Logue | 2021 | Outpatient; mean of 169.0 days post diagnosis | 4/161 | 2.50% |
| Nausea | HP:0002018 | Nausea (Figure 1) | PMID:33252665 | Petersen | 2020 | Outpatient; mean of 99.0 days post diagnosis | 7/180 | 3.90% |
| Nausea | HP:0002018 | Vomiting/nausea | PMID:33273028 | Stavem | 2020 | Outpatient; mean of 117.0 days post diagnosis | 9/451 | 2.00% |
| Nausea | HP:0002018 | Nausea | PMID:32730238 | Tenforde | 2020 | Outpatient; mean of 17.5 days post diagnosis | 27/274 | 9.90% |
| Difficulty walking | HP:0002355 | Some problems in walking plus Confined to bed | PMID:33113469 | Akter | 2020 | Inpatient; mean of 28.0 days post discharge | 124/734 | 16.90% |
| Difficulty walking | HP:0002355 | 53 patients (22.3%) were found to have limited mobility based on SPPB test results. | PMID:33502487 | Bellan | 2021 | Inpatient; mean of 105.0 days post discharge | 128/238 | 53.80% |
| Difficulty walking | HP:0002355 | 53 patients (22.3%) were found to have limited mobility based on SPPB test results. All other patients underwent a 2-minute walk test, which revealed a subtler impairment in 75 patients (31.5%). By this method, we identified 128 patients (53.8%) with some degree of functional impairment. | PMID:33502487 | Bellan | 2021 | Inpatient; mean of 105.0 days post discharge | 128/238 | 53.80% |
| Difficulty walking | HP:0002355 | inability to walk | PMID:33785495 | Dennis | 2021 | Inpatient; mean of 138.0 days post discharge | 22/37 | 59.50% |
| Difficulty walking | HP:0002355 | inability to walk | PMID:33785495 | Dennis | 2021 | Outpatient; mean of 141.0 days post diagnosis | 58/163 | 35.60% |
| Difficulty walking | HP:0002355 | Distance walked in 6 min, m, Less than lower limit of thenormal range | PMID:33428867 | Huang | 2021 | Inpatient; mean of 186.0 days post discharge | 392/1692 | 23.20% |
| Difficulty walking | HP:0002355 | After recovery, were you able to walk about? (Not normal) | PMID:33680620 | Iqbal | 2021 | Mixed; mean of 38.1 days post diagnosis | 79/158 | 50.00% |
| Difficulty walking | HP:0002355 | Some/much difficulty walking | PMID:33306721 | Jacobs | 2020 | Inpatient (non-ICU); mean of 35.0 days post discharge | 27/183 | 14.80% |
| Difficulty walking | HP:0002355 | Walking disturbances | PMID:33654513 | Suárez-Robles | 2020 | Inpatient; mean of 90.0 days post discharge | 5/134 | 3.70% |
| Difficulty walking | HP:0002355 | Mobility, some problems | PMID:33413976 | Taboada | 2021 | Inpatient (ICU); mean of 180.0 days post discharge | 51/91 | 56.00% |
| Difficulty walking | HP:0002355 | 6MWD <80%predicted,No. (%) | PMID:33220049 | van den Borst | 2020 | Inpatient (ICU); mean of 91.0 days post discharge | 9/46 | 19.60% |
| Difficulty walking | HP:0002355 | 6MWD <80%predicted,No. (%) | PMID:33220049 | van den Borst | 2020 | Outpatient; mean of 91.0 days post diagnosis | 03/27/21 | 11.10% |
| Difficulty walking | HP:0002355 | 6MWD <80%predicted,No. (%) | PMID:33220049 | van den Borst | 2020 | Inpatient (non-ICU); mean of 91.0 days post discharge | 13/51 | 25.50% |
| Memory impairment | HP:0002354 | Memory loss | PMID:33113469 | Akter | 2020 | Inpatient; mean of 28.0 days post discharge | 141/734 | 19.20% |
| Memory impairment | HP:0002354 | memory impairment (6%) | PMID:33731329 | Cheng | 2021 | Mixed; mean of 63.0 days post diagnosis | 7/113 | 6.20% |

| | | | | | | | | |
|---|---|---|---|---|---|---|---|---|
| Memory impairment | HP:0002354 | Memory loss | PMID:32853602 | Garrigues | 2020 | Inpatient (non-ICU); mean of 110.9 days post discharge | 36/96 | 37.50% |
| Memory impairment | HP:0002354 | Memory loss | PMID:32853602 | Garrigues | 2020 | Inpatient (ICU); mean of 110.9 days post discharge | 05/24/21 | 20.80% |
| Memory impairment | HP:0002354 | Memory impairment | PMID:33825846 | Havervall | 2021 | Outpatient; mean of 120.0 days post diagnosis | 4/323 | 1.20% |
| Memory impairment | HP:0002354 | Memory impairment | PMID:33825846 | Havervall | 2021 | Outpatient; mean of 60.0 days post diagnosis | 5/323 | 1.50% |
| Memory impairment | HP:0002354 | Memory impairment | PMID:33825846 | Havervall | 2021 | Outpatient; mean of 240.0 days post diagnosis | 1/323 | 0.30% |
| Memory impairment | HP:0002354 | Memory problems | PMID:33624010 | Jacobson | 2021 | Inpatient; mean of 119.3 days post discharge | 05/22/21 | 22.70% |
| Memory impairment | HP:0002354 | Memory problems | PMID:33624010 | Jacobson | 2021 | Outpatient; mean of 119.3 days post diagnosis | 15/96 | 15.60% |
| Memory impairment | HP:0002354 | memory loss | PMID:33205450 | Ludvigsson | 2021 | Outpatient; mean of 210.0 days post diagnosis | 03/05/21 | 60.00% |
| Memory impairment | HP:0002354 | Mnesic complaints | PMID:33450302 | Moreno-Perez | 2021 | Mixed; mean of 77.0 days post diagnosis | 42/277 | 15.20% |
| Memory impairment | HP:0002354 | Memory difficulties | PMID:33729425 | Writing Committee | 2021 | Inpatient (ICU); mean of 93.0 days post discharge | 10/62 | 16.10% |
| Memory impairment | HP:0002354 | Memory difficulties | PMID:33729425 | Writing Committee | 2021 | Inpatient (non-ICU); mean of 121.0 days post discharge | 63/354 | 17.80% |
| Cognitive impairment | HP:0100543 | Cognitive disorders | PMID:33120193 | Daher | 2020 | Inpatient (ICU); mean of 56.0 days post discharge | 6/33 | 18.20% |
| Cognitive impairment | HP:0100543 | Five patients had mild and three had moderate cognitive impairment | PMID:33657671 | Darley | 2021 | Mixed; mean of 69.0 days post diagnosis | 8/78 | 10.30% |
| Cognitive impairment | HP:0100543 | Difficulty thinking | PMID:-1 | Davis | 2020 | Mixed; mean of 114.5 days post diagnosis | 2444/3762 | 65.00% |
| Cognitive impairment | HP:0100543 | Cognitive impairment | PMID:33052920 | De Lorenzo | 2020 | Inpatient; mean of 21.5 days post discharge | 36/126 | 28.60% |
| Cognitive impairment | HP:0100543 | Cognitive impairment | PMID:33052920 | De Lorenzo | 2020 | Outpatient; mean of 26.0 days post diagnosis | 11/59 | 18.60% |
| Cognitive impairment | HP:0100543 | Assessment of mild cognitive impairment was more frequent according to MOCA (n = 16 [57.14%]) but not according to the Quality of Life in Neurological Disorders (n = 6 [22%]) score | PMID:32835708 | Ramani | 2021 | Inpatient (ICU); mean of 39.5 days post discharge | 06/28/21 | 21.40% |
| Cognitive impairment | HP:0100543 | Cognitive impairment | PMID:33729425 | Writing Committee | 2021 | Inpatient (ICU); mean of 93.0 days post discharge | 21/50 | 42.00% |
| Cognitive impairment | HP:0100543 | Cognitive impairment (impairment of either MoCA or d2-R score) | PMID:33729425 | Writing Committee | 2021 | Inpatient (non-ICU); mean of 121.0 days post discharge | 40/109 | 36.70% |
| Cognitive impairment | HP:0100543 | Cognitive Failures (CFQ-25 > 31·8) | PMID:33532720 | de Graaf | 2021 | Inpatient (ICU); mean of 42.0 days post discharge | 6/34 | 17.60% |
| Cognitive impairment | HP:0100543 | Cognitive Failures (CFQ-25 > 31·8) | PMID:33532720 | de Graaf | 2021 | Inpatient (non-ICU); mean of 42.0 days post discharge | 07/28/21 | 25.00% |
| Cognitive impairment | HP:0100543 | TICS <34, No. (%) | PMID:33220049 | van den Borst | 2020 | Inpatient (ICU); mean of 91.0 days post discharge | 7/46 | 15.20% |
| Cognitive impairment | HP:0100543 | TICS <34, No. (%) | PMID:33220049 | van den Borst | 2020 | Outpatient; mean of 91.0 days post diagnosis | 03/27/21 | 11.10% |
| Cognitive impairment | HP:0100543 | TICS <34, No. (%) | PMID:33220049 | van den Borst | 2020 | Inpatient (non-ICU); mean of 91.0 days post discharge | 9/51 | 17.60% |
| Ground-glass opacification | HP:0025179 | Chest CT scan Ground-glass | PMID:33676998 | González | 2021 | Inpatient (ICU); mean of 90.0 days post discharge | 34/57 | 59.60% |
| Ground-glass opacification | HP:0025179 | GGO | PMID:33428867 | Huang | 2021 | Inpatient; mean of 186.0 days post discharge | 158/348 | 45.40% |
| Ground-glass opacification | HP:0025179 | five patients showed abnormalities on the HRCT imaging, including two patients with local ground-glass opacification (GGO) and three patients with both local GGO and fibrosis. | PMID:33289374 | Liang | 2020 | Inpatient (non-ICU); mean of 90.0 days post discharge | 05/21/21 | 23.80% |
| Ground-glass opacification | HP:0025179 | GGO and multiple GCOs | PMID:32692945 | Liu | 2020 | Inpatient; mean of 28.0 days post discharge | 17/51 | 33.30% |
| Ground-glass opacification | HP:0025179 | Ground glass opacities | PMID:33303539 | Sonnweber | 2020 | Mixed; mean of 63.0 days post diagnosis | 102/145 | 70.30% |
| Ground-glass opacification | HP:0025179 | Persistent ground-glass opacities | PMID:33729425 | Writing Committee | 2021 | Inpatient (ICU); mean of 93.0 days post discharge | 27/49 | 55.10% |
| Ground-glass opacification | HP:0025179 | Persistent ground-glass opacities | PMID:33729425 | Writing Committee | 2021 | Inpatient (non-ICU); mean of 121.0 days post discharge | 45/121 | 37.20% |
| Ground-glass opacification | HP:0025179 | Ground-glass opacity | PMID:33540129 | Zhang | 2021 | Inpatient; mean of 90.0 days post discharge | 10/263 | 3.80% |
| Ground-glass opacification | HP:0025179 | Ground-glass opacity | PMID:33540129 | Zhang | 2021 | Inpatient; mean of 180.0 days post discharge | 5/96 | 5.20% |
| Ground-glass opacification | HP:0025179 | Ground-glass opacity | PMID:33540129 | Zhang | 2021 | Inpatient; mean of 28.0 days post discharge | 30/428 | 7.00% |
| Ground-glass opacification | HP:0025179 | Pure GGO | PMID:32838236 | Zhao | 2020 | Inpatient; mean of 78.5 days post discharge | 7/55 | 12.70% |
| Ground-glass opacification | HP:0025179 | Ground-glass opacity | PMID:33220049 | van den Borst | 2020 | Inpatient (ICU); mean of 91.0 days post discharge | 34/46 | 73.90% |
| Ground-glass opacification | HP:0025179 | Ground-glass opacity | PMID:33220049 | van den Borst | 2020 | Inpatient (non-ICU); mean of 91.0 days post discharge | 39/51 | 76.50% |
| Posttraumatic stress symptom | HP:0033676 | Results of the IES-R questionnaire were within reference ranges in 136 patients (57.1%), while in 61 patients (25.6%) had mild symptoms, 27 patients (11.3%) had moderate symptoms, and 14 patients (5.9%) had severe symptoms. | PMID:33502487 | Bellan | 2021 | Inpatient; mean of 105.0 days post discharge | 102/238 | 42.90% |
| Posttraumatic stress symptom | HP:0033676 | Posttraumatic stress symptoms were reported in a total of 41 patients (17.2%). | PMID:33502487 | Bellan | 2021 | Inpatient; mean of 105.0 days post discharge | 41/238 | 17.20% |
| Posttraumatic stress symptom | HP:0033676 | PTSD | PMID:33052920 | De Lorenzo | 2020 | Inpatient; mean of 21.5 days post discharge | 18/126 | 14.30% |
| Posttraumatic stress symptom | HP:0033676 | PTSD | PMID:33052920 | De Lorenzo | 2020 | Outpatient; mean of 26.0 days post diagnosis | 23/59 | 39.00% |
| Posttraumatic stress symptom | HP:0033676 | Any PTSD symptoms related to illness | PMID:32729939 | Halpin | 2021 | Inpatient (ICU); mean of 48.0 days post discharge | 15/32 | 46.90% |
| Posttraumatic stress symptom | HP:0033676 | Any PTSD symptoms related to illness | PMID:32729939 | Halpin | 2021 | Inpatient (non-ICU); mean of 48.0 days post discharge | 16/68 | 23.50% |
| Posttraumatic stress symptom | HP:0033676 | Symptoms of PTSD (PCL-5 score) | PMID:33729425 | Writing Committee | 2021 | Inpatient (ICU); mean of 93.0 days post discharge | 5/50 | 10.00% |
| Posttraumatic stress symptom | HP:0033676 | Symptoms of PTSD (PCL-5 score) | PMID:33729425 | Writing Committee | 2021 | Inpatient (non-ICU); mean of 121.0 days post discharge | 19/119 | 16.00% |
| Posttraumatic stress symptom | HP:0033676 | PTSS (PCL-5 ≥ 38) | PMID:33532720 | de Graaf | 2021 | Inpatient (ICU); mean of 42.0 days post discharge | 2/34 | 5.90% |
| Posttraumatic stress symptom | HP:0033676 | PTSS (PCL-5 ≥ 38) | PMID:33532720 | de Graaf | 2021 | Inpatient (non-ICU); mean of 42.0 days post discharge | 3/47 | 6.40% |
| Posttraumatic stress symptom | HP:0033676 | PCL-5 >33, No. (%) | PMID:33220049 | van den Borst | 2020 | Inpatient (ICU); mean of 91.0 days post discharge | 4/46 | 8.70% |
| Posttraumatic stress symptom | HP:0033676 | PCL-5 >33, No. (%) | PMID:33220049 | van den Borst | 2020 | Outpatient; mean of 91.0 days post diagnosis | 02/27/21 | 7.40% |
| Posttraumatic stress symptom | HP:0033676 | PCL-5 >33, No. (%) | PMID:33220049 | van den Borst | 2020 | Inpatient (non-ICU); mean of 91.0 days post discharge | 3/51 | 5.90% |
| Vomiting | HP:0002013 | Emesis | PMID:33120193 | Daher | 2020 | Inpatient (ICU); mean of 56.0 days post discharge | 0/33 | 0.00% |
| Vomiting | HP:0002013 | Nausea/vomiting | PMID:33657671 | Darley | 2021 | Mixed; mean of 69.0 days post diagnosis | 3/78 | 3.80% |
| Vomiting | HP:0002013 | Vomiting | PMID:-1 | Davis | 2020 | Mixed; mean of 114.5 days post diagnosis | 539/3762 | 14.30% |

| | | | | | | | | |
|---|---|---|---|---|---|---|---|---|
| Vomiting | HP:0002013 | Vomiting (1%) | PMID:33257910 | Goertz | 2020 | Mixed; mean of 79.0 days post diagnosis | 21/2113 | 1.00% |
| Vomiting | HP:0002013 | vomiting | PMID:33755344 | Graham | 2021 | Outpatient; mean of 141.0 days post diagnosis | 2/100 | 2.00% |
| Vomiting | HP:0002013 | Diarrhoea or vomiting | PMID:33428867 | Huang | 2021 | Inpatient; mean of 186.0 days post discharge | 80/1655 | 4.80% |
| Vomiting | HP:0002013 | Nausea/vomiting/diarrhea | PMID:33624010 | Jacobson | 2021 | Inpatient; mean of 119.3 days post discharge | 0/22 | 0.00% |
| Vomiting | HP:0002013 | Nausea/vomiting/Diarrhea | PMID:33624010 | Jacobson | 2021 | Outpatient; mean of 119.3 days post diagnosis | 8/96 | 8.30% |
| Vomiting | HP:0002013 | vomiting | PMID:33205450 | Ludvigsson | 2021 | Outpatient; mean of 210.0 days post diagnosis | 02/05/21 | 40.00% |
| Vomiting | HP:0002013 | Diarrhea or vomiting | PMID:33303539 | Sonnweber | 2020 | Mixed; mean of 103.0 days post diagnosis | 13/145 | 9.00% |
| Vomiting | HP:0002013 | Vomiting/nausea | PMID:33273028 | Stavem | 2020 | Outpatient; mean of 117.0 days post diagnosis | 9/451 | 2.00% |
| Vomiting | HP:0002013 | Vomiting | PMID:32730238 | Tenforde | 2020 | Outpatient; mean of 17.5 days post diagnosis | 8/274 | 2.90% |
| Insomnia | HP:0100785 | Can't sleep | PMID:33113469 | Akter | 2020 | Inpatient; mean of 28.0 days post discharge | 65/734 | 8.90% |
| Insomnia | HP:0100785 | Insomnia | PMID:33273026 | Arnold | 2020 | Inpatient (ICU); mean of 84.0 days post discharge | 09/18/21 | 50.00% |
| Insomnia | HP:0100785 | Insomnia | PMID:33273026 | Arnold | 2020 | Inpatient (non-ICU); mean of 84.0 days post discharge | 11/65 | 16.90% |
| Insomnia | HP:0100785 | Insomnia | PMID:33273026 | Arnold | 2020 | Inpatient (non-ICU); mean of 84.0 days post discharge | 06/27/21 | 22.20% |
| Insomnia | HP:0100785 | Insomnia | PMID:-1 | Davis | 2020 | Mixed; mean of 114.5 days post diagnosis | 2582/3762 | 68.60% |
| Insomnia | HP:0100785 | Insomnia | PMID:33052920 | De Lorenzo | 2020 | Inpatient; mean of 21.5 days post discharge | 31/126 | 24.60% |
| Insomnia | HP:0100785 | Insomnia | PMID:33052920 | De Lorenzo | 2020 | Outpatient; mean of 26.0 days post diagnosis | 20/59 | 33.90% |
| Insomnia | HP:0100785 | Insomnia | PMID:33755344 | Graham | 2021 | Outpatient; mean of 141.0 days post diagnosis | 33/100 | 33.00% |
| Insomnia | HP:0100785 | Five of 23 patients who completed the insomnia severity index had moderate to severe insomnia. | PMID:32835708 | Ramani | 2021 | Inpatient (ICU); mean of 39.5 days post discharge | 05/23/21 | 21.70% |
| Insomnia | HP:0100785 | Insomnia | PMID:33413976 | Taboada | 2021 | Inpatient (ICU); mean of 180.0 days post discharge | 28/91 | 30.80% |
| Insomnia | HP:0100785 | Insomnia (ISI score) | PMID:33729425 | Writing Committee | 2021 | Inpatient (ICU); mean of 93.0 days post discharge | 22/50 | 44.00% |
| Insomnia | HP:0100785 | Insomnia (ISI score) | PMID:33729425 | Writing Committee | 2021 | Inpatient (non-ICU); mean of 121.0 days post discharge | 68/118 | 57.60% |
| Reduced forced expiratory volume in one second | HP:0032342 | FEV1 <LLN, n (%) | PMID:33872135 | Abdallah | 2021 | Inpatient; mean of 102.3 days post discharge | 04/25/21 | 16.00% |
| Reduced forced expiratory volume in one second | HP:0032342 | FEV1 <LLN, n (%) | PMID:33872135 | Abdallah | 2021 | Outpatient; mean of 129.8 days post diagnosis | 4/38 | 10.50% |
| Reduced forced expiratory volume in one second | HP:0032342 | FEV1 >80%, n=92/100 | PMID:33662544 | Blanco | 2021 | Inpatient; mean of 104.0 days post discharge | 8/100 | 8.00% |
| Reduced forced expiratory volume in one second | HP:0032342 | FEV1 < lower limit of normal | PMID:33657671 | Darley | 2021 | Mixed; mean of 69.0 days post diagnosis | 5/65 | 7.70% |
| Reduced forced expiratory volume in one second | HP:0032342 | FEV1 <80%, % of predicted | PMID:33428867 | Huang | 2021 | Inpatient; mean of 186.0 days post discharge | 22/349 | 6.30% |
| Reduced forced expiratory volume in one second | HP:0032342 | four (5%) had mild impairment of FEV1 | PMID:33289374 | Liang | 2020 | Inpatient (non-ICU); mean of 90.0 days post discharge | 4/76 | 5.30% |
| Reduced forced expiratory volume in one second | HP:0032342 | Abnormalities were noted in FEV1 % predicted in 6/56 (11%) | PMID:33490928 | Ramani | 2021 | Inpatient; mean of 48.0 days post discharge | 6/56 | 10.70% |
| Reduced forced expiratory volume in one second | HP:0032342 | FVC <80% of predicted normal – no. (%) | PMID:33303539 | Sonnweber | 2020 | Mixed; mean of 63.0 days post diagnosis | 34/126 | 27.00% |
| Reduced forced expiratory volume in one second | HP:0032342 | FEV1 <80% of predicted normal – no. (%) | PMID:33303539 | Sonnweber | 2020 | Mixed; mean of 103.0 days post diagnosis | 30/133 | 22.60% |
| Reduced forced expiratory volume in one second | HP:0032342 | FEV1<LLN, No. (%) | PMID:33220049 | van den Borst | 2020 | Inpatient (ICU); mean of 91.0 days post discharge | 4/46 | 8.70% |
| Reduced forced expiratory volume in one second | HP:0032342 | FEV1<LLN, No. (%) | PMID:33220049 | van den Borst | 2020 | Outpatient; mean of 91.0 days post diagnosis | 02/27/21 | 7.40% |
| Reduced forced expiratory volume in one second | HP:0032342 | FEV1<LLN, No. (%) | PMID:33220049 | van den Borst | 2020 | Inpatient (non-ICU); mean of 91.0 days post discharge | 6/51 | 11.80% |
| Sleep disturbance | HP:0002360 | Disturbance in sound sleep | PMID:33113469 | Akter | 2020 | Inpatient; mean of 28.0 days post discharge | 145/734 | 19.80% |
| Sleep disturbance | HP:0002360 | Sleep disorder | PMID:32853602 | Garrigues | 2020 | Inpatient (non-ICU); mean of 110.9 days post discharge | 29/96 | 30.20% |
| Sleep disturbance | HP:0002360 | Sleep disorder | PMID:32853602 | Garrigues | 2020 | Inpatient (ICU); mean of 110.9 days post discharge | 08/24/21 | 33.30% |
| Sleep disturbance | HP:0002360 | Sleeping disorder | PMID:33825846 | Havervall | 2021 | Outpatient; mean of 120.0 days post diagnosis | 9/323 | 2.80% |
| Sleep disturbance | HP:0002360 | Sleeping disorder | PMID:33825846 | Havervall | 2021 | Outpatient; mean of 60.0 days post diagnosis | 10/323 | 3.10% |
| Sleep disturbance | HP:0002360 | Sleeping disorder | PMID:33825846 | Havervall | 2021 | Outpatient; mean of 240.0 days post diagnosis | 7/323 | 2.20% |
| Sleep disturbance | HP:0002360 | Sleep difficulties | PMID:33428867 | Huang | 2021 | Inpatient; mean of 186.0 days post discharge | 437/1655 | 26.40% |
| Sleep disturbance | HP:0002360 | Poor sleep quality | PMID:33680620 | Iqbal | 2021 | Mixed; mean of 38.1 days post diagnosis | 89/158 | 56.30% |
| Sleep disturbance | HP:0002360 | sleep disorders | PMID:33205450 | Ludvigsson | 2021 | Outpatient; mean of 210.0 days post diagnosis | 02/05/21 | 40.00% |
| Sleep disturbance | HP:0002360 | Sleep disorders | PMID:33303539 | Sonnweber | 2020 | Mixed; mean of 103.0 days post diagnosis | 41/145 | 28.30% |
| Sleep disturbance | HP:0002360 | Somnipathy | PMID:32979574 | Xiong | 2021 | Inpatient; mean of 97.0 days post discharge | 95/538 | 17.70% |
| Rhinorrhea | HP:0031417 | Rhinorrhea | PMID:33120193 | Daher | 2020 | Inpatient (ICU); mean of 56.0 days post discharge | 4/33 | 12.10% |
| Rhinorrhea | HP:0031417 | Rhinorrhea | PMID:33657671 | Darley | 2021 | Mixed; mean of 69.0 days post diagnosis | 5/78 | 6.40% |
| Rhinorrhea | HP:0031417 | Runny nose | PMID:-1 | Davis | 2020 | Mixed; mean of 114.5 days post diagnosis | 1102/3762 | 29.30% |
| Rhinorrhea | HP:0031417 | Runny nose | PMID:33785495 | Dennis | 2021 | Inpatient; mean of 138.0 days post discharge | 13/37 | 35.10% |
| Rhinorrhea | HP:0031417 | Runny nose | PMID:33785495 | Dennis | 2021 | Outpatient; mean of 141.0 days post diagnosis | 55/163 | 33.70% |

| | | | | | | | | |
|---|---|---|---|---|---|---|---|---|
| Rhinorrhea | HP:0031417 | Congestion/rhinorrhea | PMID:33624010 | Jacobson | 2021 | Inpatient; mean of 119.3 days post discharge | 01/22/21 | 4.50% |
| Rhinorrhea | HP:0031417 | Congestion/Rhinorrhea | PMID:33624010 | Jacobson | 2021 | Outpatient; mean of 119.3 days post diagnosis | 7/96 | 7.30% |
| Rhinorrhea | HP:0031417 | Runny nose | PMID:33606031 | Logue | 2021 | Inpatient; mean of 169.0 days post discharge | 0/16 | 0.00% |
| Rhinorrhea | HP:0031417 | Runny nose | PMID:33606031 | Logue | 2021 | Outpatient; mean of 169.0 days post diagnosis | 5/161 | 3.10% |
| Rhinorrhea | HP:0031417 | Rhinorrhea (Figure 1) | PMID:33252665 | Petersen | 2020 | Outpatient; mean of 99.0 days post diagnosis | 16/180 | 8.90% |
| Rhinorrhea | HP:0031417 | Runny nose | PMID:33273028 | Stavem | 2020 | Outpatient; mean of 117.0 days post diagnosis | 18/451 | 4.00% |
| Parageusia | HP:0031249 | Dysgeusia | PMID:32644129 | Carfi | 2020 | Inpatient; mean of 60.3 days post discharge | 14/143 | 9.80% |
| Parageusia | HP:0031249 | Dysgeusia | PMID:33564789 | Chun | 2021 | Mixed; mean of 63.0 days post diagnosis | 10/61 | 16.40% |
| Parageusia | HP:0031249 | Altered sense of taste | PMID:-1 | Davis | 2020 | Mixed; mean of 114.5 days post diagnosis | 943/3762 | 25.10% |
| Parageusia | HP:0031249 | Dysgeusia | PMID:33755344 | Graham | 2021 | Outpatient; mean of 141.0 days post diagnosis | 60/100 | 60.00% |
| Parageusia | HP:0031249 | Taste disorder | PMID:33428867 | Huang | 2021 | Inpatient; mean of 186.0 days post discharge | 120/1655 | 7.30% |
| Parageusia | HP:0031249 | deranged smell and taste | PMID:33205450 | Ludvigsson | 2021 | Outpatient; mean of 210.0 days post diagnosis | 01/05/21 | 20.00% |
| Parageusia | HP:0031249 | Anosmia-dysgeusia | PMID:33450302 | Moreno-Perez | 2021 | Mixed; mean of 77.0 days post diagnosis | 59/277 | 21.30% |
| Parageusia | HP:0031249 | Disgeusia 15% | PMID:33787016 | Prieto | 2021 | Mixed; mean of 53.0 days post diagnosis | 13/85 | 15.30% |
| Parageusia | HP:0031249 | Loss/disturbance of taste | PMID:33273028 | Stavem | 2020 | Outpatient; mean of 117.0 days post diagnosis | 45/451 | 10.00% |
| Parageusia | HP:0031249 | Dysgeusia | PMID:33654513 | Suárez-Robles | 2020 | Inpatient; mean of 90.0 days post discharge | 29/134 | 21.60% |
| Parageusia | HP:0031249 | Anosmia/Dysgeusia | PMID:33461632 | Venturelli | 2021 | Mixed; mean of 81.0 days post diagnosis | 23/515 | 4.50% |
| Abdominal pain | HP:0002027 | Abdominal pain | PMID:33273026 | Arnold | 2020 | Inpatient (ICU); mean of 84.0 days post discharge | 0/18 | 0.00% |
| Abdominal pain | HP:0002027 | Abdominal pain | PMID:33273026 | Arnold | 2020 | Inpatient (non-ICU); mean of 84.0 days post discharge | 1/65 | 1.50% |
| Abdominal pain | HP:0002027 | Abdominal pain | PMID:33273026 | Arnold | 2020 | Inpatient (non-ICU); mean of 84.0 days post discharge | 01/27/21 | 3.70% |
| Abdominal pain | HP:0002027 | Stomach pains | PMID:33120193 | Daher | 2020 | Inpatient (ICU); mean of 56.0 days post discharge | 1/33 | 3.00% |
| Abdominal pain | HP:0002027 | Abdominal pain | PMID:-1 | Davis | 2020 | Mixed; mean of 114.5 days post diagnosis | 1492/3762 | 39.70% |
| Abdominal pain | HP:0002027 | Abdominal pain | PMID:33785495 | Dennis | 2021 | Inpatient; mean of 138.0 days post discharge | 17/37 | 45.90% |
| Abdominal pain | HP:0002027 | Abdominal pain | PMID:33785495 | Dennis | 2021 | Outpatient; mean of 141.0 days post diagnosis | 91/163 | 55.80% |
| Abdominal pain | HP:0002027 | abdominal pain | PMID:33205450 | Ludvigsson | 2021 | Outpatient; mean of 210.0 days post diagnosis | 03/05/21 | 60.00% |
| Abdominal pain | HP:0002027 | Abdominal pain | PMID:33273028 | Stavem | 2020 | Outpatient; mean of 117.0 days post diagnosis | 13/451 | 2.90% |
| Abdominal pain | HP:0002027 | Abdominal pain | PMID:32730238 | Tenforde | 2020 | Outpatient; mean of 17.5 days post diagnosis | 44/274 | 16.10% |
| Vertigo | HP:0002321 | Vertigo | PMID:32644129 | Carfi | 2020 | Inpatient; mean of 60.3 days post discharge | 7/143 | 4.90% |
| Vertigo | HP:0002321 | Dizziness / vertigo / unsteadiness or balance issues | PMID:-1 | Davis | 2020 | Mixed; mean of 114.5 days post diagnosis | 2531/3762 | 67.30% |
| Vertigo | HP:0002321 | Dizziness (27%) | PMID:33257910 | Goertz | 2020 | Mixed; mean of 79.0 days post diagnosis | 571/2113 | 27.00% |
| Vertigo | HP:0002321 | Dizziness | PMID:33755344 | Graham | 2021 | Outpatient; mean of 141.0 days post diagnosis | 47/100 | 47.00% |
| Vertigo | HP:0002321 | Dizziness | PMID:33428867 | Huang | 2021 | Inpatient; mean of 186.0 days post discharge | 101/1655 | 6.10% |
| Vertigo | HP:0002321 | dizziness | PMID:33205450 | Ludvigsson | 2021 | Outpatient; mean of 210.0 days post diagnosis | 04/05/21 | 80.00% |
| Vertigo | HP:0002321 | Vertigo/dizziness/lightheadedness, self-reported | PMID:33682276 | Rass | 2021 | Outpatient; mean of 90.0 days post diagnosis | 2/32 | 6.30% |
| Vertigo | HP:0002321 | Vertigo/dizziness/lightheadedness, self-reported | PMID:33682276 | Rass | 2021 | Inpatient (non-ICU); mean of 90.0 days post discharge | 5/72 | 6.90% |
| Vertigo | HP:0002321 | Vertigo/dizziness/lightheadedness, self-reported | PMID:33682276 | Rass | 2021 | Inpatient (ICU); mean of 90.0 days post discharge | 2/31 | 6.50% |
| Vertigo | HP:0002321 | Dizziness | PMID:32979574 | Xiong | 2021 | Inpatient; mean of 97.0 days post discharge | 14/538 | 2.60% |
| Reduced total lung capacity | HP:0033169 | TLC <LLN, n (%) | PMID:33872135 | Abdallah | 2021 | Inpatient; mean of 102.3 days post discharge | 12/25/21 | 48.00% |
| Reduced total lung capacity | HP:0033169 | TLC <LLN, n (%) | PMID:33872135 | Abdallah | 2021 | Outpatient; mean of 129.8 days post diagnosis | 3/38 | 7.90% |
| Reduced total lung capacity | HP:0033169 | In eight patients (12%), total lung capacity was abnormal (ie, below lower limit of normal) | PMID:33657671 | Darley | 2021 | Mixed; mean of 69.0 days post diagnosis | 8/65 | 12.30% |
| Reduced total lung capacity | HP:0033169 | TLC, <80% | PMID:33676998 | González | 2021 | Inpatient (ICU); mean of 90.0 days post discharge | 23/62 | 37.10% |
| Reduced total lung capacity | HP:0033169 | TLC <80%, % of predicted | PMID:33428867 | Huang | 2021 | Inpatient; mean of 186.0 days post discharge | 56/334 | 16.80% |
| Reduced total lung capacity | HP:0033169 | TLC <80% of predicted normal – no. (%) | PMID:33303539 | Sonnweber | 2020 | Mixed; mean of 63.0 days post diagnosis | 14/126 | 11.10% |
| Reduced total lung capacity | HP:0033169 | TLC <80% of predicted normal – no. (%) | PMID:33303539 | Sonnweber | 2020 | Mixed; mean of 103.0 days post diagnosis | 15/133 | 11.30% |
| Reduced total lung capacity | HP:0033169 | TLC<LLN, No. (%) | PMID:33220049 | van den Borst | 2020 | Inpatient (ICU); mean of 91.0 days post discharge | 7/46 | 15.20% |
| Reduced total lung capacity | HP:0033169 | TLC<LLN, No. (%) | PMID:33220049 | van den Borst | 2020 | Outpatient; mean of 91.0 days post diagnosis | 01/27/21 | 3.70% |
| Reduced total lung capacity | HP:0033169 | TLC<LLN, No. (%) | PMID:33220049 | van den Borst | 2020 | Inpatient (non-ICU); mean of 91.0 days post discharge | 7/51 | 13.70% |
| Occupational disability | HP:0033695 | Sick leave | PMID:33031948 | Carvalho-Schneider | 2021 | Mixed; mean of 60.0 days post diagnosis | 14/130 | 10.80% |
| Occupational disability | HP:0033695 | Sick leave | PMID:33031948 | Carvalho-Schneider | 2021 | Mixed; mean of 30.0 days post diagnosis | 26/150 | 17.30% |
| Occupational disability | HP:0033695 | Of the 69 patients who worked prior to admission, 27 had not yet returned to work at 6 to 12 week review due to their symptoms (27/69=39%) | PMID:33731329 | Cheng | 2021 | Mixed; mean of 63.0 days post diagnosis | 27/69 | 39.10% |
| Occupational disability | HP:0033695 | (not) ablt to return to work 60 d after discharrge | PMID:33175566 | Chopra | 2020 | Inpatient; mean of 60.0 days post discharge | 78/195 | 40.00% |
| Occupational disability | HP:0033695 | Complement of Returned to work/worked before hospitalization | PMID:32853602 | Garrigues | 2020 | Inpatient (non-ICU); mean of 110.9 days post discharge | 10/41 | 24.40% |
| Occupational disability | HP:0033695 | Returned to work/worked before hospitalization | PMID:32853602 | Garrigues | 2020 | Inpatient (ICU); mean of 110.9 days post discharge | 08/15/21 | 53.30% |
| Occupational disability | HP:0033695 | Any Work Impairment due to health | PMID:33624010 | Jacobson | 2021 | Inpatient; mean of 119.3 days post discharge | 07/12/21 | 58.30% |
| Occupational disability | HP:0033695 | Any Work Impairment due to health | PMID:33624010 | Jacobson | 2021 | Outpatient; mean of 119.3 days post diagnosis | 21/96 | 21.90% |
| Occupational disability | HP:0033695 | All 55 patients had returned to their original work. | PMID:32838236 | Zhao | 2020 | Inpatient; mean of 78.5 days post discharge | 0/55 | 0.00% |
| Diminished ability to concentrate | HP:0031987 | Loss of concentration | PMID:33113469 | Akter | 2020 | Inpatient; mean of 28.0 days post discharge | 188/734 | 25.60% |
| Diminished ability to concentrate | HP:0031987 | Poor attention or concentration | PMID:-1 | Davis | 2020 | Mixed; mean of 114.5 days post diagnosis | 2814/3762 | 74.80% |
| Diminished ability to concentrate | HP:0031987 | New or worsened concentration problem | PMID:32729939 | Halpin | 2021 | Inpatient (ICU); mean of 48.0 days post discharge | 11/32 | 34.40% |

| | | | | | | | | |
|---|---|---|---|---|---|---|---|---|
| Diminished ability to concentrate | HP:0031987 | New or worsened concentration problem | PMID:32729939 | Halpin | 2021 | Inpatient (non-ICU); mean of 48.0 days post discharge | 11/68 | 16.20% |
| Diminished ability to concentrate | HP:0031987 | Concentration impairment | PMID:33825846 | Havervall | 2021 | Outpatient; mean of 120.0 days post diagnosis | 6/323 | 1.90% |
| Diminished ability to concentrate | HP:0031987 | Concentration impairment | PMID:33825846 | Havervall | 2021 | Outpatient; mean of 60.0 days post diagnosis | 7/323 | 2.20% |
| Diminished ability to concentrate | HP:0031987 | Concentration impairment | PMID:33825846 | Havervall | 2021 | Outpatient; mean of 240.0 days post diagnosis | 2/323 | 0.60% |
| Diminished ability to concentrate | HP:0031987 | difficulties concentrating | PMID:33205450 | Ludvigsson | 2021 | Outpatient; mean of 210.0 days post diagnosis | 04/05/21 | 80.00% |
| Diminished ability to concentrate | HP:0031987 | Concentration problems | PMID:33729425 | Writing Committee | 2021 | Inpatient (non-ICU); mean of 121.0 days post discharge | 35/351 | 10.00% |
| Alopecia | HP:0001596 | Hairfall | PMID:33113469 | Akter | 2020 | Inpatient; mean of 28.0 days post discharge | 71/734 | 9.70% |
| Alopecia | HP:0001596 | hair-loss (6%) | PMID:33731329 | Cheng | 2021 | Mixed; mean of 63.0 days post diagnosis | 7/113 | 6.20% |
| Alopecia | HP:0001596 | Hair loss | PMID:32853602 | Garrigues | 2020 | Inpatient (non-ICU); mean of 110.9 days post discharge | 18/96 | 18.80% |
| Alopecia | HP:0001596 | Hair loss | PMID:32853602 | Garrigues | 2020 | Inpatient (ICU); mean of 110.9 days post discharge | 06/24/21 | 25.00% |
| Alopecia | HP:0001596 | Hair loss | PMID:33428867 | Huang | 2021 | Inpatient; mean of 186.0 days post discharge | 359/1655 | 21.70% |
| Alopecia | HP:0001596 | Hair fall | PMID:33680620 | Iqbal | 2021 | Mixed; mean of 38.1 days post diagnosis | 63/158 | 39.90% |
| Alopecia | HP:0001596 | Hair loss | PMID:33624010 | Jacobson | 2021 | Inpatient; mean of 119.3 days post discharge | 04/22/21 | 18.20% |
| Alopecia | HP:0001596 | Hair loss | PMID:33624010 | Jacobson | 2021 | Outpatient; mean of 119.3 days post diagnosis | 10/96 | 10.40% |
| Alopecia | HP:0001596 | Alopecia | PMID:32979574 | Xiong | 2021 | Inpatient; mean of 97.0 days post discharge | 154/538 | 28.60% |
| Parenchymal consolidation | HP:0032177 | Chest CT scan Consolidation | PMID:33676998 | González | 2021 | Inpatient (ICU); mean of 90.0 days post discharge | 9/57 | 15.80% |
| Parenchymal consolidation | HP:0032177 | Chest CT/Consolidation | PMID:33428867 | Huang | 2021 | Inpatient; mean of 186.0 days post discharge | 4/161 | 2.50% |
| Parenchymal consolidation | HP:0032177 | Consolidation | PMID:32692945 | Liu | 2020 | Inpatient; mean of 28.0 days post discharge | 1/51 | 2.00% |
| Parenchymal consolidation | HP:0032177 | CT consolidations | PMID:33303539 | Sonnweber | 2020 | Mixed; mean of 63.0 days post diagnosis | 93/145 | 64.10% |
| Parenchymal consolidation | HP:0032177 | CT: consolidations | PMID:33479105 | Trinkmann | 2021 | Mixed; mean of 68.0 days post diagnosis | 12/17/21 | 70.60% |
| Parenchymal consolidation | HP:0032177 | Consolidation (CT) | PMID:33540129 | Zhang | 2021 | Inpatient; mean of 90.0 days post discharge | 47/263 | 17.90% |
| Parenchymal consolidation | HP:0032177 | Consolidation (CT) | PMID:33540129 | Zhang | 2021 | Inpatient; mean of 180.0 days post discharge | 24/96 | 25.00% |
| Parenchymal consolidation | HP:0032177 | Consolidation (CT) | PMID:33540129 | Zhang | 2021 | Inpatient; mean of 28.0 days post discharge | 48/428 | 11.20% |
| Pain | HP:0012531 | Pain/discomfort | PMID:33113469 | Akter | 2020 | Inpatient; mean of 28.0 days post discharge | 233/734 | 31.70% |
| Pain | HP:0012531 | Pain other than chest | PMID:33755344 | Graham | 2021 | Outpatient; mean of 141.0 days post diagnosis | 43/100 | 43.00% |
| Pain | HP:0012531 | Worsened pain/discomfort | PMID:32729939 | Halpin | 2021 | Inpatient (ICU); mean of 48.0 days post discharge | 9/32 | 28.10% |
| Pain | HP:0012531 | Worsened pain/discomfort | PMID:32729939 | Halpin | 2021 | Inpatient (non-ICU); mean of 48.0 days post discharge | 10/68 | 14.70% |
| Pain | HP:0012531 | Pain or discomfort | PMID:33428867 | Huang | 2021 | Inpatient; mean of 186.0 days post discharge | 431/1616 | 26.70% |
| Pain | HP:0012531 | After recovery, did you feel pain or discomfort? | PMID:33680620 | Iqbal | 2021 | Mixed; mean of 38.1 days post diagnosis | 117/158 | 74.10% |
| Pain | HP:0012531 | Pain | PMID:33303539 | Sonnweber | 2020 | Mixed; mean of 103.0 days post diagnosis | 35/145 | 24.10% |
| Pain | HP:0012531 | Pain or discomfort: Some pain or discomfort or Extreme pain or discomfort | PMID:33413976 | Taboada | 2021 | Inpatient (ICU); mean of 180.0 days post discharge | 44/91 | 48.40% |
| Reduced forced vital capacity | HP:0032341 | FVC <LLN, n (%) | PMID:33872135 | Abdallah | 2021 | Inpatient; mean of 102.3 days post discharge | 06/25/21 | 24.00% |
| Reduced forced vital capacity | HP:0032341 | FVC <LLN, n (%) | PMID:33872135 | Abdallah | 2021 | Outpatient; mean of 129.8 days post diagnosis | 1/38 | 2.60% |
| Reduced forced vital capacity | HP:0032341 | FVC >80%, n=94/100 | PMID:33662544 | Blanco | 2021 | Inpatient; mean of 104.0 days post discharge | 6/100 | 6.00% |
| Reduced forced vital capacity | HP:0032341 | FVC < lower limit of normal | PMID:33657671 | Darley | 2021 | Mixed; mean of 69.0 days post diagnosis | 3/65 | 4.60% |
| Reduced forced vital capacity | HP:0032341 | FVC <80%, % of predicted | PMID:33428867 | Huang | 2021 | Inpatient; mean of 186.0 days post discharge | 14/349 | 4.00% |
| Reduced forced vital capacity | HP:0032341 | Abnormalities were noted in FVC % predicted in 7/56 (13%). | PMID:33490928 | Ramani | 2021 | Inpatient; mean of 48.0 days post discharge | 7/56 | 12.50% |
| Reduced forced vital capacity | HP:0032341 | FVC <80% of predicted normal – no. (%) | PMID:33303539 | Sonnweber | 2020 | Mixed; mean of 63.0 days post diagnosis | 34/126 | 27.00% |
| Reduced forced vital capacity | HP:0032341 | FVC <80% of predicted normal – no. (%) | PMID:33303539 | Sonnweber | 2020 | Mixed; mean of 103.0 days post diagnosis | 29/133 | 21.80% |
| Chills | HP:0025143 | Fever/chills | PMID:33657671 | Darley | 2021 | Mixed; mean of 69.0 days post diagnosis | 5/78 | 6.40% |
| Chills | HP:0025143 | Chills/flushing/sweats | PMID:-1 | Davis | 2020 | Mixed; mean of 114.5 days post diagnosis | 2124/3762 | 56.50% |
| Chills | HP:0025143 | Chills or shivering | PMID:33606031 | Logue | 2021 | Inpatient; mean of 169.0 days post discharge | 01/16/21 | 6.30% |
| Chills | HP:0025143 | Chills or shivering | PMID:33606031 | Logue | 2021 | Outpatient; mean of 169.0 days post diagnosis | 4/161 | 2.50% |
| Chills | HP:0025143 | Chills | PMID:33273028 | Stavem | 2020 | Outpatient; mean of 117.0 days post diagnosis | 5/451 | 1.10% |
| Chills | HP:0025143 | Chills | PMID:32730238 | Tenforde | 2020 | Outpatient; mean of 17.5 days post diagnosis | 14/274 | 5.10% |
| Chills | HP:0025143 | Chills | PMID:32979574 | Xiong | 2021 | Inpatient; mean of 97.0 days post discharge | 25/538 | 4.60% |
| Oxygen desaturation on exertion | HP:0030874 | Severe desaturation on STS test (1 minute Sit to Stand desaturation test) | PMID:33273026 | Arnold | 2020 | Inpatient (ICU); mean of 84.0 days post discharge | 05/18/21 | 27.80% |
| Oxygen desaturation on exertion | HP:0030874 | Severe desaturation on STS test (1 minute Sit to Stand desaturation test) | PMID:33273026 | Arnold | 2020 | Inpatient (non-ICU); mean of 84.0 days post discharge | 10/65 | 15.40% |
| Oxygen desaturation on exertion | HP:0030874 | Severe desaturation on STS test (1 minute Sit to Stand desaturation test) | PMID:33273026 | Arnold | 2020 | Inpatient (non-ICU); mean of 84.0 days post discharge | 0/27 | 0.00% |
| Oxygen desaturation on exertion | HP:0030874 | Four (7%) patients desaturated at the end of the test. | PMID:33490928 | Ramani | 2021 | Inpatient; mean of 48.0 days post discharge | 4/57 | 7.00% |
| Oxygen desaturation on exertion | HP:0030874 | Desaturation >=4% upon6MWT, No. (%) | PMID:33220049 | van den Borst | 2020 | Inpatient (ICU); mean of 91.0 days post discharge | 8/46 | 17.40% |
| Oxygen desaturation on exertion | HP:0030874 | Desaturation >=4% upon6MWT, No. (%) | PMID:33220049 | van den Borst | 2020 | Outpatient; mean of 91.0 days post diagnosis | 01/27/21 | 3.70% |
| Oxygen desaturation on exertion | HP:0030874 | Desaturation >=4% upon6MWT, No. (%) | PMID:33220049 | van den Borst | 2020 | Inpatient (non-ICU); mean of 91.0 days post discharge | 11/51 | 21.60% |
| Reduced FEV1/FVC ratio | HP:0030877 | FEV1/FVC < LLN, n (%) | PMID:33872135 | Abdallah | 2021 | Inpatient; mean of 102.3 days post discharge | 01/25/21 | 4.00% |
| Reduced FEV1/FVC ratio | HP:0030877 | FEV1/FVC < LLN, n (%) | PMID:33872135 | Abdallah | 2021 | Outpatient; mean of 129.8 days post diagnosis | 6/38 | 15.80% |
| Reduced FEV1/FVC ratio | HP:0030877 | FEV1/FVC <70% | PMID:33428867 | Huang | 2021 | Inpatient; mean of 186.0 days post discharge | 22/349 | 6.30% |
| Reduced FEV1/FVC ratio | HP:0030877 | 17 (22%) had mild impairment of FEV1/FVC | PMID:33289374 | Liang | 2020 | Inpatient (non-ICU); mean of 90.0 days post discharge | 17/76 | 22.40% |
| Reduced FEV1/FVC ratio | HP:0030877 | Obstruction was considered as FEV1/FVC <0.7 | PMID:33450302 | Moreno-Perez | 2021 | Mixed; mean of 77.0 days post diagnosis | 34/269 | 12.60% |

| | | | | | | | | |
|---|---|---|---|---|---|---|---|---|
| Reduced FEV1/FVC ratio | HP:0030877 | FEV1:FVC <70% – no. (%) | PMID:33303539 | Sonnweber | 2020 | Mixed; mean of 63.0 days post diagnosis | 5/126 | 4.00% |
| Reduced FEV1/FVC ratio | HP:0030877 | FEV1:FVC <70% – no. (%) | PMID:33303539 | Sonnweber | 2020 | Mixed; mean of 103.0 days post diagnosis | 11/133 | 8.30% |
| Dysphagia | HP:0002015 | Lump in throat/difficulty swallowing | PMID:-1 | Davis | 2020 | Mixed; mean of 114.5 days post diagnosis | 1222/3762 | 32.50% |
| Dysphagia | HP:0002015 | Dysphagia | PMID:33755344 | Graham | 2021 | Outpatient; mean of 141.0 days post diagnosis | 1/100 | 1.00% |
| Dysphagia | HP:0002015 | Swallow problem | PMID:32729939 | Halpin | 2021 | Inpatient (ICU); mean of 48.0 days post discharge | 4/32 | 12.50% |
| Dysphagia | HP:0002015 | Swallow problem | PMID:32729939 | Halpin | 2021 | Inpatient (non-ICU); mean of 48.0 days post discharge | 4/68 | 5.90% |
| Dysphagia | HP:0002015 | Dysphagia | PMID:33682276 | Rass | 2021 | Outpatient; mean of 90.0 days post diagnosis | 0/32 | 0.00% |
| Dysphagia | HP:0002015 | Dysphagia | PMID:33682276 | Rass | 2021 | Inpatient (non-ICU); mean of 90.0 days post discharge | 0/72 | 0.00% |
| Dysphagia | HP:0002015 | Dysphagia | PMID:33682276 | Rass | 2021 | Inpatient (ICU); mean of 90.0 days post discharge | 0/31 | 0.00% |
| Impairment of activities of daily living | HP:0031058 | Unable to wash or dress myself and Some problem in washing or dressing | PMID:33113469 | Akter | 2020 | Inpatient; mean of 28.0 days post discharge | 70/734 | 9.50% |
| Impairment of activities of daily living | HP:0031058 | 29 showing both an SPPB score ≤10 and 1-MSTST repetitions lower than expectancy | PMID:33565741 | Baricich | 2021 | Inpatient; mean of 124.7 days post discharge | 29/204 | 14.20% |
| Impairment of activities of daily living | HP:0031058 | After recovery, were you able to carry out your usual activities? (business, office work, study, housework or leisure activities) | PMID:33680620 | Iqbal | 2021 | Mixed; mean of 38.1 days post diagnosis | 125/158 | 79.10% |
| Impairment of activities of daily living | HP:0031058 | Any Activity impairment due to health | PMID:33624010 | Jacobson | 2021 | Inpatient; mean of 119.3 days post discharge | 14/19 | 73.70% |
| Impairment of activities of daily living | HP:0031058 | Any Activity impairment due to health | PMID:33624010 | Jacobson | 2021 | Outpatient; mean of 119.3 days post diagnosis | 40/87 | 46.00% |
| Impairment of activities of daily living | HP:0031058 | All but one patient (96%) performed activities of daily life without difficulty. | PMID:32835708 | Ramani | 2021 | Inpatient (ICU); mean of 39.5 days post discharge | 01/28/21 | 3.60% |
| Impairment of activities of daily living | HP:0031058 | Usual activities, some problems or unable to perform | PMID:33413976 | Taboada | 2021 | Inpatient (ICU); mean of 180.0 days post discharge | 34/91 | 37.40% |
| Skin rash | HP:0000988 | Skin rashes | PMID:-1 | Davis | 2020 | Mixed; mean of 114.5 days post diagnosis | 1045/3762 | 27.80% |
| Skin rash | HP:0000988 | Skin rash | PMID:33428867 | Huang | 2021 | Inpatient; mean of 186.0 days post discharge | 47/1655 | 2.80% |
| Skin rash | HP:0000988 | Rash | PMID:33606031 | Logue | 2021 | Inpatient; mean of 169.0 days post discharge | 0/16 | 0.00% |
| Skin rash | HP:0000988 | Rash | PMID:33606031 | Logue | 2021 | Outpatient; mean of 169.0 days post diagnosis | 3/161 | 1.90% |
| Skin rash | HP:0000988 | skin rashes | PMID:33205450 | Ludvigsson | 2021 | Outpatient; mean of 210.0 days post diagnosis | 03/05/21 | 60.00% |
| Skin rash | HP:0000988 | Rashes (Figure 1) | PMID:33252665 | Petersen | 2020 | Outpatient; mean of 99.0 days post diagnosis | 5/180 | 2.80% |
| Skin rash | HP:0000988 | Skin rash | PMID:33273028 | Stavem | 2020 | Outpatient; mean of 117.0 days post diagnosis | 9/451 | 2.00% |
| Blurred vision | HP:0000622 | Blurred vision | PMID:-1 | Davis | 2020 | Mixed; mean of 114.5 days post diagnosis | 1343/3762 | 35.70% |
| Blurred vision | HP:0000622 | Blurred vision | PMID:33755344 | Graham | 2021 | Outpatient; mean of 141.0 days post diagnosis | 30/100 | 30.00% |
| Blurred vision | HP:0000622 | Blurred vision | PMID:33680620 | Iqbal | 2021 | Mixed; mean of 38.1 days post diagnosis | 30/158 | 19.00% |
| Blurred vision | HP:0000622 | Blurring vision, selfreported | PMID:33682276 | Rass | 2021 | Outpatient; mean of 90.0 days post diagnosis | 1/32 | 3.10% |
| Blurred vision | HP:0000622 | Blurring vision, selfreported | PMID:33682276 | Rass | 2021 | Inpatient (non-ICU); mean of 90.0 days post discharge | 7/72 | 9.70% |
| Blurred vision | HP:0000622 | Blurring vision, selfreported | PMID:33682276 | Rass | 2021 | Inpatient (ICU); mean of 90.0 days post discharge | 01/31/21 | 3.20% |
| Blurred vision | HP:0000622 | Vision disturbance/blurring | PMID:33273028 | Stavem | 2020 | Outpatient; mean of 117.0 days post diagnosis | 18/451 | 4.00% |
| Increased sputum production | HP:0033709 | Sputum production | PMID:32644129 | Carfi | 2020 | Inpatient; mean of 60.3 days post discharge | 12/143 | 8.40% |
| Increased sputum production | HP:0033709 | mucus (18%) | PMID:33257910 | Goertz | 2020 | Mixed; mean of 79.0 days post diagnosis | 254/2113 | 12.00% |
| Increased sputum production | HP:0033709 | Phlegm | PMID:33306721 | Jacobs | 2020 | Inpatient (non-ICU); mean of 35.0 days post discharge | 27/183 | 14.80% |
| Increased sputum production | HP:0033709 | Sputum | PMID:33289374 | Liang | 2020 | Inpatient (non-ICU); mean of 90.0 days post discharge | 33/76 | 43.40% |
| Increased sputum production | HP:0033709 | Sputum | PMID:32692945 | Liu | 2020 | Inpatient; mean of 28.0 days post discharge | 2/51 | 3.90% |
| Increased sputum production | HP:0033709 | Sputum | PMID:33654513 | Suárez-Robles | 2020 | Inpatient; mean of 90.0 days post discharge | 7/134 | 5.20% |
| Increased sputum production | HP:0033709 | Sputum | PMID:32979574 | Xiong | 2021 | Inpatient; mean of 97.0 days post discharge | 16/538 | 3.00% |
| Chest tightness | HP:0031352 | Shortness of breath/chest tightness/wheezing | PMID:33175566 | Chopra | 2020 | Inpatient; mean of 60.0 days post discharge | 81/488 | 16.60% |
| Chest tightness | HP:0031352 | chest tightness (14) | PMID:33657671 | Darley | 2021 | Mixed; mean of 69.0 days post diagnosis | 14/78 | 17.90% |
| Chest tightness | HP:0031352 | Tightness of Chest | PMID:-1 | Davis | 2020 | Mixed; mean of 114.5 days post diagnosis | 2813/3762 | 74.80% |
| Chest tightness | HP:0031352 | Chest tightness (44%) | PMID:33257910 | Goertz | 2020 | Mixed; mean of 79.0 days post diagnosis | 930/2113 | 44.00% |
| Chest tightness | HP:0031352 | Chest tightness on exertion | PMID:33289374 | Liang | 2020 | Inpatient (non-ICU); mean of 90.0 days post discharge | 47/76 | 61.80% |
| Chest tightness | HP:0031352 | Chest tightness (Figure 1) | PMID:33252665 | Petersen | 2020 | Outpatient; mean of 99.0 days post diagnosis | 11/180 | 6.10% |
| Chest tightness | HP:0031352 | Chest distress | PMID:32979574 | Xiong | 2021 | Inpatient; mean of 97.0 days post discharge | 76/538 | 14.10% |
| Elevated circulating D-dimer concentration | HP:0033106 | D-dimer abnormal | PMID:33755344 | Graham | 2021 | Outpatient; mean of 141.0 days post diagnosis | 08/27/21 | 29.60% |
| Elevated circulating D-dimer concentration | HP:0033106 | 30.1% (of 176) had persistently elevated d-dimer concentration | PMID:33172844 | Mandal | 2020 | Inpatient; mean of 54.0 days post discharge | 53/176 | 30.10% |
| Elevated circulating D-dimer concentration | HP:0033106 | D-dimers > 0.5 mg/mL | PMID:33450302 | Moreno-Perez | 2021 | Mixed; mean of 77.0 days post diagnosis | 68/273 | 24.90% |
| Elevated circulating D-dimer concentration | HP:0033106 | D-dimer, NT-proBNP, and serum ferritin were still elevated in 27%, 23%, and 17% of COVID-19 patients at second follow-up, respectively | PMID:33303539 | Sonnweber | 2020 | Mixed; mean of 103.0 days post diagnosis | 36/135 | 26.70% |
| Elevated circulating D-dimer concentration | HP:0033106 | elevated D-dimer levels were observed in 38 (25.3%) patients at time of follow-up | PMID:33587810 | Townsend | 2021 | Mixed; mean of 80.5 days post diagnosis | 38/150 | 25.30% |
| Elevated circulating D-dimer concentration | HP:0033106 | D-dimer ⩾2000 | PMID:33461632 | Venturelli | 2021 | Mixed; mean of 81.0 days post diagnosis | 39/727 | 5.40% |
| Weight loss | HP:0001824 | Weight loss ≥5% | PMID:33031948 | Carvalho-Schneider | 2021 | Mixed; mean of 60.0 days post diagnosis | 15/130 | 11.50% |

| | | | | | | | | |
|---|---|---|---|---|---|---|---|---|
| Weight loss | HP:0001824 | Weight loss ≥5% | PMID:33031948 | Carvalho-Schneider | 2021 | Mixed; mean of 30.0 days post diagnosis | 13/150 | 8.70% |
| Weight loss | HP:0001824 | Sudden loss of BW (3%) | PMID:33257910 | Goertz | 2020 | Mixed; mean of 79.0 days post diagnosis | 63/2113 | 3.00% |
| Weight loss | HP:0001824 | Loss of weight | PMID:33654513 | Suárez-Robles | 2020 | Inpatient; mean of 90.0 days post discharge | 50/134 | 37.30% |
| Weight loss | HP:0001824 | Weight loss >5% baseline weight | PMID:33729425 | Writing Committee | 2021 | Inpatient (ICU); mean of 93.0 days post discharge | 1/61 | 1.60% |
| Weight loss | HP:0001824 | Weight loss >5% baseline weight | PMID:33729425 | Writing Committee | 2021 | Inpatient (non-ICU); mean of 121.0 days post discharge | 30/281 | 10.70% |
| Confusion | HP:0001289 | Altered consciousness/confusion | PMID:33657671 | Darley | 2021 | Mixed; mean of 69.0 days post diagnosis | 0/78 | 0.00% |
| Confusion | HP:0001289 | Acute (sudden) confusion/disorientation | PMID:-1 | Davis | 2020 | Mixed; mean of 114.5 days post diagnosis | 691/3762 | 18.40% |
| Confusion | HP:0001289 | Confusion | PMID:33306721 | Jacobs | 2020 | Inpatient (non-ICU); mean of 35.0 days post discharge | 16/183 | 8.70% |
| Confusion | HP:0001289 | Confusion/changed consciousness | PMID:33273028 | Stavem | 2020 | Outpatient; mean of 117.0 days post diagnosis | 9/451 | 2.00% |
| Confusion | HP:0001289 | Confusion | PMID:32730238 | Tenforde | 2020 | Outpatient; mean of 17.5 days post diagnosis | 55/274 | 20.10% |
| Confusion | HP:0001289 | Confusion | PMID:33461632 | Venturelli | 2021 | Mixed; mean of 81.0 days post diagnosis | 23/515 | 4.50% |
| Seizure | HP:0001250 | Seizures (confirmed) | PMID:-1 | Davis | 2020 | Mixed; mean of 114.5 days post diagnosis | 22/3762 | 0.60% |
| Seizure | HP:0001250 | Seizure | PMID:33755344 | Graham | 2021 | Outpatient; mean of 141.0 days post diagnosis | 1/100 | 1.00% |
| Seizure | HP:0001250 | Seizures | PMID:33682276 | Rass | 2021 | Outpatient; mean of 90.0 days post diagnosis | 0/32 | 0.00% |
| Seizure | HP:0001250 | Seizures | PMID:33682276 | Rass | 2021 | Inpatient (non-ICU); mean of 90.0 days post discharge | 0/72 | 0.00% |
| Seizure | HP:0001250 | Seizures | PMID:33682276 | Rass | 2021 | Inpatient (ICU); mean of 90.0 days post discharge | 0/31 | 0.00% |
| Seizure | HP:0001250 | Seizures/cramps | PMID:33273028 | Stavem | 2020 | Outpatient; mean of 117.0 days post diagnosis | 9/451 | 2.00% |
| Pulmonary fibrosis | HP:0002206 | Chest CT scan Fibrotic | PMID:33676998 | González | 2021 | Inpatient (ICU); mean of 90.0 days post discharge | 12/57 | 21.10% |
| Pulmonary fibrosis | HP:0002206 | Pulmonary fibrosis | PMID:33680620 | Iqbal | 2021 | Mixed; mean of 38.1 days post diagnosis | 6/158 | 3.80% |
| Pulmonary fibrosis | HP:0002206 | Lung fibrotic lesions | PMID:33729425 | Writing Committee | 2021 | Inpatient (ICU); mean of 93.0 days post discharge | 18/49 | 36.70% |
| Pulmonary fibrosis | HP:0002206 | Lung fibrotic lesions | PMID:33729425 | Writing Committee | 2021 | Inpatient (non-ICU); mean of 121.0 days post discharge | 15/121 | 12.40% |
| Pulmonary fibrosis | HP:0002206 | residual CT abnormality: fibrosis | PMID:33220049 | van den Borst | 2020 | Inpatient (ICU); mean of 91.0 days post discharge | 14/46 | 30.40% |
| Pulmonary fibrosis | HP:0002206 | residual CT abnormality: fibrosis | PMID:33220049 | van den Borst | 2020 | Inpatient (non-ICU); mean of 91.0 days post discharge | 8/51 | 15.70% |
| Paresthesia | HP:0003401 | Tingling/prickling/pins and needles sensation | PMID:-1 | Davis | 2020 | Mixed; mean of 114.5 days post diagnosis | 1852/3762 | 49.20% |
| Paresthesia | HP:0003401 | Numbness/tingling | PMID:33755344 | Graham | 2021 | Outpatient; mean of 141.0 days post diagnosis | 60/100 | 60.00% |
| Paresthesia | HP:0003401 | Numbness/tingling/burning, selfreported | PMID:33682276 | Rass | 2021 | Inpatient (non-ICU); mean of 90.0 days post discharge | 11/72 | 15.30% |
| Paresthesia | HP:0003401 | Numbness/tingling/burning, selfreported | PMID:33682276 | Rass | 2021 | Inpatient (ICU); mean of 90.0 days post discharge | 10/31/21 | 32.30% |
| Paresthesia | HP:0003401 | Paresthesia | PMID:33729425 | Writing Committee | 2021 | Inpatient (ICU); mean of 93.0 days post discharge | 11/62 | 17.70% |
| Paresthesia | HP:0003401 | Paresthesia | PMID:33729425 | Writing Committee | 2021 | Inpatient (non-ICU); mean of 121.0 days post discharge | 40/359 | 11.10% |
| Asthenia | HP:0025406 | Flulike symptoms (Myalgia, headache and/or asthenia) | PMID:33031948 | Carvalho-Schneider | 2021 | Mixed; mean of 60.0 days post diagnosis | 28/130 | 21.50% |
| Asthenia | HP:0025406 | Flulike symptoms (Myalgia, headache and/or asthenia) | PMID:33031948 | Carvalho-Schneider | 2021 | Mixed; mean of 30.0 days post diagnosis | 54/150 | 36.00% |
| Asthenia | HP:0025406 | Weakness | PMID:-1 | Davis | 2020 | Mixed; mean of 114.5 days post diagnosis | 1675/3762 | 44.50% |
| Asthenia | HP:0025406 | asthenia (30.5%), | PMID:33521308 | Rosales-Castillo | 2021 | Inpatient; mean of 50.8 days post discharge | 36/118 | 30.50% |
| Asthenia | HP:0025406 | Asthenia | PMID:33413976 | Taboada | 2021 | Inpatient (ICU); mean of 180.0 days post discharge | 34/91 | 37.40% |
| Asthenia | HP:0025406 | Asthenia | PMID:33461632 | Venturelli | 2021 | Mixed; mean of 81.0 days post diagnosis | 186/515 | 36.10% |
| Exertional dyspnea | HP:0002875 | Exertional breathlessness | PMID:33872135 | Abdallah | 2021 | Inpatient; mean of 102.3 days post discharge | 17/25 | 68.00% |
| Exertional dyspnea | HP:0002875 | Exertional breathlessness | PMID:33872135 | Abdallah | 2021 | Outpatient; mean of 129.8 days post diagnosis | 31/38 | 81.60% |
| Exertional dyspnea | HP:0002875 | Breathlessness walking up stairs | PMID:33175566 | Chopra | 2020 | Inpatient; mean of 60.0 days post discharge | 112/488 | 23.00% |
| Exertional dyspnea | HP:0002875 | Dyspnea on exertion | PMID:33413976 | Taboada | 2021 | Inpatient (ICU); mean of 180.0 days post discharge | 52/91 | 57.10% |
| Exertional dyspnea | HP:0002875 | Postactivity polypnoea | PMID:32979574 | Xiong | 2021 | Inpatient; mean of 97.0 days post discharge | 115/538 | 21.40% |
| Exertional dyspnea | HP:0002875 | exertional dyspnea (14.55%), | PMID:32838236 | Zhao | 2020 | Inpatient; mean of 78.5 days post discharge | 8/55 | 14.50% |
| Poor appetite | HP:0004396 | Lack of appetite | PMID:32644129 | Carfi | 2020 | Inpatient; mean of 60.3 days post discharge | 11/143 | 7.70% |
| Poor appetite | HP:0004396 | Loss of Appetite | PMID:-1 | Davis | 2020 | Mixed; mean of 114.5 days post diagnosis | 1942/3762 | 51.60% |
| Poor appetite | HP:0004396 | Appetite problem severity 2 or more | PMID:32729939 | Halpin | 2021 | Inpatient (non-ICU); mean of 48.0 days post discharge | 6/68 | 8.80% |
| Poor appetite | HP:0004396 | Decreased appetite | PMID:33428867 | Huang | 2021 | Inpatient; mean of 186.0 days post discharge | 138/1655 | 8.30% |
| Poor appetite | HP:0004396 | poor appetite | PMID:33205450 | Ludvigsson | 2021 | Outpatient; mean of 210.0 days post diagnosis | 01/05/21 | 20.00% |
| Poor appetite | HP:0004396 | Loss of appetite | PMID:33654513 | Suárez-Robles | 2020 | Inpatient; mean of 90.0 days post discharge | 36/134 | 26.90% |
| Reduced residual volume | HP:0033753 | RV <LLN, n (%) | PMID:33872135 | Abdallah | 2021 | Inpatient; mean of 102.3 days post discharge | 08/25/21 | 32.00% |
| Reduced residual volume | HP:0033753 | RV <LLN, n (%) | PMID:33872135 | Abdallah | 2021 | Outpatient; mean of 129.8 days post diagnosis | 4/38 | 10.50% |
| Reduced residual volume | HP:0033753 | RV <80%, % of predicted | PMID:33428867 | Huang | 2021 | Inpatient; mean of 186.0 days post discharge | 87/333 | 26.10% |
| Reduced residual volume | HP:0033753 | RV<LLN, No. (%) | PMID:33220049 | van den Borst | 2020 | Inpatient (ICU); mean of 91.0 days post discharge | 3/46 | 6.50% |
| Reduced residual volume | HP:0033753 | RV<LLN, No. (%) | PMID:33220049 | van den Borst | 2020 | Outpatient; mean of 91.0 days post diagnosis | 02/27/21 | 7.40% |
| Reduced residual volume | HP:0033753 | RV<LLN, No. (%) | PMID:33220049 | van den Borst | 2020 | Inpatient (non-ICU); mean of 91.0 days post discharge | 4/51 | 7.80% |
| Restrictive ventilatory defect | HP:0002091 | Restrictive pattern spirometry | PMID:33273026 | Arnold | 2020 | Inpatient (ICU); mean of 84.0 days post discharge | 03/18/21 | 16.70% |
| Restrictive ventilatory defect | HP:0002091 | Restrictive pattern spirometry | PMID:33273026 | Arnold | 2020 | Inpatient (non-ICU); mean of 84.0 days post discharge | 8/65 | 12.30% |
| Restrictive ventilatory defect | HP:0002091 | Restrictive pattern spirometry | PMID:33273026 | Arnold | 2020 | Inpatient (non-ICU); mean of 84.0 days post discharge | 0/27 | 0.00% |
| Restrictive ventilatory defect | HP:0002091 | Restriction | PMID:33450302 | Moreno-Perez | 2021 | Mixed; mean of 77.0 days post diagnosis | 10/269 | 3.70% |
| Restrictive ventilatory defect | HP:0002091 | Spirometry Interpretation: Restrictive pattern | PMID:33865161 | Ordinola Navarro | 2021 | Mixed; mean of 40.0 days post diagnosis | 20/115 | 17.40% |
| Restrictive ventilatory defect | HP:0002091 | five (19.23%) had restriction,one (3.85%) had mixed obstruction and restriction | PMID:32835708 | Ramani | 2021 | Inpatient (ICU); mean of 39.5 days post discharge | 06/28/21 | 21.40% |
| Elevated circulating C-reactive protein concentration | HP:0011227 | C-reactive protein abnormal | PMID:33755344 | Graham | 2021 | Outpatient; mean of 141.0 days post diagnosis | 10/52 | 19.20% |

| | | | | | | | | |
|---|---|---|---|---|---|---|---|---|
| Elevated circulating C-reactive protein concentration | HP:0011227 | 9.5% (of 332) had persistently elevated d-dimer and CRP concentration | PMID:33172844 | Mandal | 2020 | Inpatient; mean of 54.0 days post discharge | 32/332 | 9.60% |
| Elevated circulating C-reactive protein concentration | HP:0011227 | C-reactive protein > 0.5 mg/dL | PMID:33450302 | Moreno-Perez | 2021 | Mixed; mean of 77.0 days post diagnosis | 32/276 | 11.60% |
| Elevated circulating C-reactive protein concentration | HP:0011227 | mild elevations in inflammatory markers such as CRP (12%) | PMID:33303539 | Sonnweber | 2020 | Mixed; mean of 103.0 days post diagnosis | 17/145 | 11.70% |
| Elevated circulating C-reactive protein concentration | HP:0011227 | Mild increases in CRP were only present in 17/150 | PMID:33587810 | Townsend | 2021 | Mixed; mean of 80.5 days post diagnosis | 17/150 | 11.30% |
| Elevated circulating C-reactive protein concentration | HP:0011227 | C-reactive protein >1.0 | PMID:33461632 | Venturelli | 2021 | Mixed; mean of 81.0 days post diagnosis | 52/727 | 7.20% |
| Ear pain | HP:0030766 | Ear pain | PMID:-1 | Davis | 2020 | Mixed; mean of 114.5 days post diagnosis | 971/3762 | 25.80% |
| Ear pain | HP:0030766 | Ear pain (8%) | PMID:33257910 | Goertz | 2020 | Mixed; mean of 79.0 days post diagnosis | 169/2113 | 8.00% |
| Ear pain | HP:0030766 | Ear pain | PMID:33606031 | Logue | 2021 | Inpatient; mean of 169.0 days post discharge | 0/16 | 0.00% |
| Ear pain | HP:0030766 | Ear pain | PMID:33606031 | Logue | 2021 | Outpatient; mean of 169.0 days post diagnosis | 4/161 | 2.50% |
| Ear pain | HP:0030766 | Ear pain | PMID:33273028 | Stavem | 2020 | Outpatient; mean of 117.0 days post diagnosis | 9/451 | 2.00% |
| Bronchiectasis | HP:0002110 | Chest CT Scan Bronchiectasis | PMID:33676998 | González | 2021 | Inpatient (ICU); mean of 90.0 days post discharge | 41/57 | 71.90% |
| Bronchiectasis | HP:0002110 | Bronchiectasis | PMID:32692945 | Liu | 2020 | Inpatient; mean of 28.0 days post discharge | 2/51 | 3.90% |
| Bronchiectasis | HP:0002110 | Bronchial dilation | PMID:33303539 | Sonnweber | 2020 | Mixed; mean of 63.0 days post diagnosis | 46/145 | 31.70% |
| Bronchiectasis | HP:0002110 | Bronchi(ol)ectasis | PMID:33220049 | van den Borst | 2020 | Inpatient (ICU); mean of 91.0 days post discharge | 22/46 | 47.80% |
| Bronchiectasis | HP:0002110 | Bronchi(ol)ectasis | PMID:33220049 | van den Borst | 2020 | Inpatient (non-ICU); mean of 91.0 days post discharge | 29/51 | 56.90% |
| Productive cough | HP:0031245 | Cough with mucus production | PMID:-1 | Davis | 2020 | Mixed; mean of 114.5 days post diagnosis | 1062/3762 | 28.20% |
| Productive cough | HP:0031245 | Wet cough | PMID:33676998 | González | 2021 | Inpatient (ICU); mean of 90.0 days post discharge | 11/62 | 17.70% |
| Productive cough | HP:0031245 | Cough with expectoration (Figure 1) | PMID:33252665 | Petersen | 2020 | Outpatient; mean of 99.0 days post diagnosis | 11/180 | 6.10% |
| Productive cough | HP:0031245 | Productive cough | PMID:33273028 | Stavem | 2020 | Outpatient; mean of 117.0 days post diagnosis | 18/451 | 4.00% |
| Productive cough | HP:0031245 | cough and sputum (1.81%) | PMID:32838236 | Zhao | 2020 | Inpatient; mean of 78.5 days post discharge | 1/55 | 1.80% |
| Reticular pattern on pulmonary HRCT | HP:0025390 | Chest CT Scan Reticular | PMID:33676998 | González | 2021 | Inpatient (ICU); mean of 90.0 days post discharge | 28/57 | 49.10% |
| Reticular pattern on pulmonary HRCT | HP:0025390 | Chest CT/Reticular pattern | PMID:33428867 | Huang | 2021 | Inpatient; mean of 186.0 days post discharge | 2/253 | 0.80% |
| Reticular pattern on pulmonary HRCT | HP:0025390 | Reticular pattern | PMID:32692945 | Liu | 2020 | Inpatient; mean of 28.0 days post discharge | 0/51 | 0.00% |
| Reticular pattern on pulmonary HRCT | HP:0025390 | CT reticulation | PMID:33303539 | Sonnweber | 2020 | Mixed; mean of 63.0 days post diagnosis | 84/145 | 57.90% |
| Reticular pattern on pulmonary HRCT | HP:0025390 | CT: reticulations | PMID:33479105 | Trinkmann | 2021 | Mixed; mean of 68.0 days post diagnosis | 02/17/21 | 11.80% |
| Stroke | HP:0001297 | Stroke | PMID:33680620 | Iqbal | 2021 | Mixed; mean of 38.1 days post diagnosis | 1/158 | 0.60% |
| Stroke | HP:0001297 | Stroke with clinical symptoms | PMID:33682276 | Rass | 2021 | Outpatient; mean of 90.0 days post diagnosis | 0/32 | 0.00% |
| Stroke | HP:0001297 | Stroke with clinical symptoms | PMID:33682276 | Rass | 2021 | Inpatient (non-ICU); mean of 90.0 days post discharge | 1/72 | 1.40% |
| Stroke | HP:0001297 | Stroke with clinicalsymptoms | PMID:33682276 | Rass | 2021 | Inpatient (ICU); mean of 90.0 days post discharge | 0/31 | 0.00% |
| Short term memory impairment | HP:0033687 | Short-term memory loss (memory that lasts ~30 seconds, i.e. remembering a phone number before writing it down, or forgetting you’re in the middle of a task) | PMID:-1 | Davis | 2020 | Mixed; mean of 114.5 days post diagnosis | 2438/3762 | 64.80% |
| Short term memory impairment | HP:0033687 | Short-term memory deficit | PMID:33755344 | Graham | 2021 | Outpatient; mean of 141.0 days post diagnosis | 32/100 | 32.00% |
| Short term memory impairment | HP:0033687 | New or worsened short-term memory problem | PMID:32729939 | Halpin | 2021 | Inpatient (ICU); mean of 48.0 days post discharge | 6/32 | 18.80% |
| Short term memory impairment | HP:0033687 | New or worsened short-term memory problem | PMID:32729939 | Halpin | 2021 | Inpatient (non-ICU); mean of 48.0 days post discharge | 12/68 | 17.60% |
| Stiff neck | HP:0025258 | Stiff neck | PMID:-1 | Davis | 2020 | Mixed; mean of 114.5 days post diagnosis | 1471/3762 | 39.10% |
| Stiff neck | HP:0025258 | Neck stiffness | PMID:33682276 | Rass | 2021 | Outpatient; mean of 90.0 days post diagnosis | 0/32 | 0.00% |
| Stiff neck | HP:0025258 | Neck stiffness | PMID:33682276 | Rass | 2021 | Inpatient (non-ICU); mean of 90.0 days post discharge | 0/72 | 0.00% |
| Stiff neck | HP:0025258 | Neck stiffness | PMID:33682276 | Rass | 2021 | Inpatient (ICU); mean of 90.0 days post discharge | 0/31 | 0.00% |
| Dysphonia | HP:0001618 | Changes in the voice | PMID:-1 | Davis | 2020 | Mixed; mean of 114.5 days post diagnosis | 1012/3762 | 26.90% |
| Dysphonia | HP:0001618 | Voice change | PMID:32729939 | Halpin | 2021 | Inpatient (ICU); mean of 48.0 days post discharge | 8/32 | 25.00% |
| Dysphonia | HP:0001618 | Voice change | PMID:32729939 | Halpin | 2021 | Inpatient (non-ICU); mean of 48.0 days post discharge | 12/68 | 17.60% |
| Dysphonia | HP:0001618 | Dysphonia | PMID:33654513 | Suárez-Robles | 2020 | Inpatient; mean of 90.0 days post discharge | 11/134 | 8.20% |
| Ataxia | HP:0001251 | Ataxia | PMID:33755344 | Graham | 2021 | Outpatient; mean of 141.0 days post diagnosis | 1/100 | 1.00% |
| Ataxia | HP:0001251 | Cerebellar ataxia | PMID:33682276 | Rass | 2021 | Outpatient; mean of 90.0 days post diagnosis | 0/32 | 0.00% |
| Ataxia | HP:0001251 | Cerebellar ataxia | PMID:33682276 | Rass | 2021 | Inpatient (non-ICU); mean of 90.0 days post discharge | 0/72 | 0.00% |
| Ataxia | HP:0001251 | Cerebellar ataxia | PMID:33682276 | Rass | 2021 | Inpatient (ICU); mean of 90.0 days post discharge | 0/31 | 0.00% |
| Red eye | HP:0025337 | Red eyes | PMID:32644129 | Carfi | 2020 | Inpatient; mean of 60.3 days post discharge | 14/143 | 9.80% |
| Red eye | HP:0025337 | Red eyes | PMID:33564789 | Chun | 2021 | Mixed; mean of 63.0 days post diagnosis | 0/61 | 0.00% |
| Red eye | HP:0025337 | Bloodshot eyes | PMID:-1 | Davis | 2020 | Mixed; mean of 114.5 days post diagnosis | 579/3762 | 15.40% |
| Red eye | HP:0025337 | Eye irritation | PMID:33306721 | Jacobs | 2020 | Inpatient (non-ICU); mean of 35.0 days post discharge | 15/183 | 8.20% |
| Dysarthria | HP:0001260 | Dysarthria | PMID:33755344 | Graham | 2021 | Outpatient; mean of 141.0 days post diagnosis | 2/100 | 2.00% |
| Dysarthria | HP:0001260 | Dysarthria | PMID:33682276 | Rass | 2021 | Outpatient; mean of 90.0 days post diagnosis | 1/32 | 3.10% |
| Dysarthria | HP:0001260 | Dysarthria | PMID:33682276 | Rass | 2021 | Inpatient (non-ICU); mean of 90.0 days post discharge | 1/72 | 1.40% |
| Dysarthria | HP:0001260 | Dysarthria | PMID:33682276 | Rass | 2021 | Inpatient (ICU); mean of 90.0 days post discharge | 01/31/21 | 3.20% |
| Hypertension | HP:0000822 | Abnormally high blood pressure | PMID:-1 | Davis | 2020 | Mixed; mean of 114.5 days post diagnosis | 755/3762 | 20.10% |
| Hypertension | HP:0000822 | Uncontrolled BP requiring therapeutic change | PMID:33052920 | De Lorenzo | 2020 | Inpatient; mean of 21.5 days post discharge | 26/126 | 20.60% |

| | | | | | | | | |
|---|---|---|---|---|---|---|---|---|
| Hypertension | HP:0000822 | Uncontrolled BP requiring therapeutic change | PMID:33052920 | De Lorenzo | 2020 | Outpatient; mean of 26.0 days post diagnosis | 14/59 | 23.70% |
| Hypertension | HP:0000822 | Newly diagnosed hypertension | PMID:32979574 | Xiong | 2021 | Inpatient; mean of 97.0 days post discharge | 7/538 | 1.30% |
| Muscle weakness | HP:0001324 | muscle weakness | PMID:33205450 | Ludvigsson | 2021 | Outpatient; mean of 210.0 days post diagnosis | 04/05/21 | 80.00% |
| Muscle weakness | HP:0001324 | Paresis | PMID:33682276 | Rass | 2021 | Outpatient; mean of 90.0 days post diagnosis | 0/32 | 0.00% |
| Muscle weakness | HP:0001324 | Paresis | PMID:33682276 | Rass | 2021 | Inpatient (non-ICU); mean of 90.0 days post discharge | 3/72 | 4.20% |
| Muscle weakness | HP:0001324 | Paresis | PMID:33682276 | Rass | 2021 | Inpatient (ICU); mean of 90.0 days post discharge | 4/31 | 12.90% |
| Hypoesthesia | HP:0033748 | Numbness/loss of sensation | PMID:-1 | Davis | 2020 | Mixed; mean of 114.5 days post diagnosis | 1332/3762 | 35.40% |
| Hypoesthesia | HP:0033748 | numbness | PMID:33205450 | Ludvigsson | 2021 | Outpatient; mean of 210.0 days post diagnosis | 01/05/21 | 20.00% |
| Hypoesthesia | HP:0033748 | Decreased/disturbed sensibility | PMID:33682276 | Rass | 2021 | Inpatient (non-ICU); mean of 90.0 days post discharge | 9/72 | 12.50% |
| Hypoesthesia | HP:0033748 | Decreased/disturbed sensibility | PMID:33682276 | Rass | 2021 | Inpatient (ICU); mean of 90.0 days post discharge | 08/31/21 | 25.80% |
| Abnormal exteroceptive sensation | HP:0033747 | Sensory dysfunction | PMID:33755344 | Graham | 2021 | Outpatient; mean of 141.0 days post diagnosis | 8/53 | 15.10% |
| Abnormal exteroceptive sensation | HP:0033747 | Decreased/disturbed sensibility | PMID:33682276 | Rass | 2021 | Outpatient; mean of 90.0 days post diagnosis | 3/32 | 9.40% |
| Abnormal exteroceptive sensation | HP:0033747 | Decreased/disturbed sensibility | PMID:33682276 | Rass | 2021 | Inpatient (non-ICU); mean of 90.0 days post discharge | 9/72 | 12.50% |
| Abnormal exteroceptive sensation | HP:0033747 | Decreased/disturbed sensibility | PMID:33682276 | Rass | 2021 | Inpatient (ICU); mean of 90.0 days post discharge | 08/31/21 | 25.80% |
| Hypogeusia | HP:0000224 | Hypogeusia, self-reported | PMID:33682276 | Rass | 2021 | Outpatient; mean of 90.0 days post diagnosis | 7/32 | 21.90% |
| Hypogeusia | HP:0000224 | Hypogeusia, self-reported | PMID:33682276 | Rass | 2021 | Inpatient (non-ICU); mean of 90.0 days post discharge | 11/72 | 15.30% |
| Hypogeusia | HP:0000224 | Hypogeusia, self-reported | PMID:33682276 | Rass | 2021 | Inpatient (ICU); mean of 90.0 days post discharge | 0/31 | 0.00% |
| Hypogeusia | HP:0000224 | 2 female patients still experienced a decrease sense of taste during follow-up period | PMID:32838236 | Zhao | 2020 | Inpatient; mean of 78.5 days post discharge | 2/55 | 3.60% |
| Wheezing | HP:0030828 | Shortness of breath/chest tightness/wheezing | PMID:33175566 | Chopra | 2020 | Inpatient; mean of 60.0 days post discharge | 81/488 | 16.60% |
| Wheezing | HP:0030828 | Wheezing | PMID:33785495 | Dennis | 2021 | Inpatient; mean of 138.0 days post discharge | 23/37 | 62.20% |
| Wheezing | HP:0030828 | Wheezing | PMID:33785495 | Dennis | 2021 | Outpatient; mean of 141.0 days post diagnosis | 75/163 | 46.00% |
| Wheezing | HP:0030828 | Wheeze | PMID:33273028 | Stavem | 2020 | Outpatient; mean of 117.0 days post diagnosis | 18/451 | 4.00% |
| Nonproductive cough | HP:0031246 | Dry cough | PMID:-1 | Davis | 2020 | Mixed; mean of 114.5 days post diagnosis | 2491/3762 | 66.20% |
| Nonproductive cough | HP:0031246 | Dry cough | PMID:33676998 | González | 2021 | Inpatient (ICU); mean of 90.0 days post discharge | 10/62 | 16.10% |
| Nonproductive cough | HP:0031246 | Dry cough (Figure 1) | PMID:33252665 | Petersen | 2020 | Outpatient; mean of 99.0 days post diagnosis | 7/180 | 3.90% |
| Nonproductive cough | HP:0031246 | Dry cough | PMID:33273028 | Stavem | 2020 | Outpatient; mean of 117.0 days post diagnosis | 27/451 | 6.00% |
| Encephalopathy | HP:0001298 | Mild encephalopathy | PMID:33682276 | Rass | 2021 | Outpatient; mean of 90.0 days post diagnosis | 0/32 | 0.00% |
| Encephalopathy | HP:0001298 | Mild encephalopathy | PMID:33682276 | Rass | 2021 | Inpatient (non-ICU); mean of 90.0 days post discharge | 1/72 | 1.40% |
| Encephalopathy | HP:0001298 | Mild encephalopathy | PMID:33682276 | Rass | 2021 | Inpatient (ICU); mean of 90.0 days post discharge | 01/31/21 | 3.20% |
| Pleural thickening | HP:0031944 | Pleural incrassation | PMID:33540129 | Zhang | 2021 | Inpatient; mean of 90.0 days post discharge | 28/263 | 10.60% |
| Pleural thickening | HP:0031944 | Pleural incrassation | PMID:33540129 | Zhang | 2021 | Inpatient; mean of 180.0 days post discharge | 9/96 | 9.40% |
| Pleural thickening | HP:0031944 | Pleural incrassation | PMID:33540129 | Zhang | 2021 | Inpatient; mean of 28.0 days post discharge | 35/428 | 8.20% |
| Orthostatic hypotension | HP:0001278 | Orthostatic hypotension | PMID:33682276 | Rass | 2021 | Outpatient; mean of 90.0 days post diagnosis | 0/32 | 0.00% |
| Orthostatic hypotension | HP:0001278 | Orthostatic hypotension | PMID:33682276 | Rass | 2021 | Inpatient (non-ICU); mean of 90.0 days post discharge | 0/72 | 0.00% |
| Orthostatic hypotension | HP:0001278 | Orthostatic hypotension | PMID:33682276 | Rass | 2021 | Inpatient (ICU); mean of 90.0 days post discharge | 01/31/21 | 3.20% |
| Interlobular septal thickening | HP:0030879 | Chest CT scan Interlobular septal thickening | PMID:33676998 | González | 2021 | Inpatient (ICU); mean of 90.0 days post discharge | 46/57 | 80.70% |
| Interlobular septal thickening | HP:0030879 | Chest CT/Interlobular septal thickening | PMID:33428867 | Huang | 2021 | Inpatient; mean of 186.0 days post discharge | 3/256 | 1.20% |
| Interlobular septal thickening | HP:0030879 | Interlobular septal thickening | PMID:32692945 | Liu | 2020 | Inpatient; mean of 28.0 days post discharge | 18/51 | 35.30% |
| Bradyphrenia | HP:0031843 | Slowed thoughts | PMID:-1 | Davis | 2020 | Mixed; mean of 114.5 days post diagnosis | 1572/3762 | 41.80% |
| Bradyphrenia | HP:0031843 | Mental slowness | PMID:33729425 | Writing Committee | 2021 | Inpatient (ICU); mean of 93.0 days post discharge | 4/62 | 6.50% |
| Bradyphrenia | HP:0031843 | Mental slowness | PMID:33729425 | Writing Committee | 2021 | Inpatient (non-ICU); mean of 121.0 days post discharge | 38/353 | 10.80% |
| Hyperhidrosis | HP:0000975 | Sweats | PMID:33606031 | Logue | 2021 | Inpatient; mean of 169.0 days post discharge | 01/16/21 | 6.30% |
| Hyperhidrosis | HP:0000975 | Sweats | PMID:33606031 | Logue | 2021 | Outpatient; mean of 169.0 days post diagnosis | 4/161 | 2.50% |
| Hyperhidrosis | HP:0000975 | Sweating | PMID:32979574 | Xiong | 2021 | Inpatient; mean of 97.0 days post discharge | 127/538 | 23.60% |
| Gait disturbance | HP:0001288 | Gait dysfunction | PMID:33755344 | Graham | 2021 | Outpatient; mean of 141.0 days post diagnosis | 5/100 | 5.00% |
| Gait disturbance | HP:0001288 | Gait abnormality | PMID:33682276 | Rass | 2021 | Inpatient (non-ICU); mean of 90.0 days post discharge | 2/72 | 2.80% |
| Gait disturbance | HP:0001288 | Gait abnormality | PMID:33682276 | Rass | 2021 | Inpatient (ICU); mean of 90.0 days post discharge | 4/31 | 12.90% |
| Syncope | HP:0001279 | Fainting | PMID:-1 | Davis | 2020 | Mixed; mean of 114.5 days post diagnosis | 486/3762 | 12.90% |
| Syncope | HP:0001279 | No patient reported a recent syncope. | PMID:32730619 | Puntmann | 2020 | Mixed; mean of 71.0 days post diagnosis | 0/100 | 0.00% |
| Syncope | HP:0001279 | Syncope | PMID:33461632 | Venturelli | 2021 | Mixed; mean of 81.0 days post diagnosis | 1/515 | 0.20% |
| Frontal release signs | HP:0000743 | Positive frontal release signs | PMID:33682276 | Rass | 2021 | Outpatient; mean of 90.0 days post diagnosis | 3/32 | 9.40% |
| Frontal release signs | HP:0000743 | Positive frontal release signs | PMID:33682276 | Rass | 2021 | Inpatient (non-ICU); mean of 90.0 days post discharge | 13/72 | 18.10% |
| Frontal release signs | HP:0000743 | Positive frontal release signs | PMID:33682276 | Rass | 2021 | Inpatient (ICU); mean of 90.0 days post discharge | 4/31 | 12.90% |
| Hyposmia | HP:0004409 | Hyposmia, self-reported | PMID:33682276 | Rass | 2021 | Outpatient; mean of 90.0 days post diagnosis | 10/32 | 31.30% |
| Hyposmia | HP:0004409 | Hyposmia, self-reported | PMID:33682276 | Rass | 2021 | Inpatient (non-ICU); mean of 90.0 days post discharge | 11/72 | 15.30% |
| Hyposmia | HP:0004409 | Hyposmia, self-reported | PMID:33682276 | Rass | 2021 | Inpatient (ICU); mean of 90.0 days post discharge | 0/31 | 0.00% |
| Urinary incontinence | HP:0000020 | Bladder control issues | PMID:-1 | Davis | 2020 | Mixed; mean of 114.5 days post diagnosis | 532/3762 | 14.10% |
| Urinary incontinence | HP:0000020 | New bladder control problem | PMID:32729939 | Halpin | 2021 | Inpatient (ICU); mean of 48.0 days post discharge | 4/32 | 12.50% |
| Urinary incontinence | HP:0000020 | New bladder control problem | PMID:32729939 | Halpin | 2021 | Inpatient (non-ICU); mean of 48.0 days post discharge | 6/68 | 8.80% |
| Hearing impairment | HP:0000365 | Ear and Hearing Symptoms - Hearing loss | PMID:-1 | Davis | 2020 | Mixed; mean of 114.5 days post diagnosis | 326/3762 | 8.70% |
| Hearing impairment | HP:0000365 | Decreased hearing | PMID:33755344 | Graham | 2021 | Outpatient; mean of 141.0 days post diagnosis | 2/100 | 2.00% |

| | | | | | | | | |
|---|---|---|---|---|---|---|---|---|
| Hearing impairment | HP:0000365 | Hearing loss, self-reported | PMID:32735466 | Munro | 2020 | Inpatient; mean of 56.0 days post discharge | 8/121 | 6.60% |
| Tinnitus | HP:0000360 | Tinnitus | PMID:-1 | Davis | 2020 | Mixed; mean of 114.5 days post diagnosis | 1280/3762 | 34.00% |
| Tinnitus | HP:0000360 | Tinnitus | PMID:33755344 | Graham | 2021 | Outpatient; mean of 141.0 days post diagnosis | 29/100 | 29.00% |
| Tinnitus | HP:0000360 | Tinnitus, self-reported | PMID:32735466 | Munro | 2020 | Inpatient; mean of 56.0 days post discharge | 8/121 | 6.60% |
| Tremor | HP:0001337 | Tremors | PMID:-1 | Davis | 2020 | Mixed; mean of 114.5 days post diagnosis | 1511/3762 | 40.20% |
| Tremor | HP:0001337 | Tremors | PMID:33682276 | Rass | 2021 | Inpatient (non-ICU); mean of 90.0 days post discharge | 9/72 | 12.50% |
| Tremor | HP:0001337 | Tremors | PMID:33682276 | Rass | 2021 | Inpatient (ICU); mean of 90.0 days post discharge | 4/31 | 12.90% |
| Brain fog | HP:0033630 | Cognitive Dysfunction, overall (Brain fog) | PMID:-1 | Davis | 2020 | Mixed; mean of 114.5 days post diagnosis | 3203/3762 | 85.10% |
| Brain fog | HP:0033630 | Brain fog | PMID:33755344 | Graham | 2021 | Outpatient; mean of 141.0 days post diagnosis | 85/100 | 85.00% |
| Brain fog | HP:0033630 | Brain fog | PMID:33680620 | Iqbal | 2021 | Mixed; mean of 38.1 days post diagnosis | 30/158 | 19.00% |
| Decreased maximal oxygen uptake | HP:0033760 | VO2 %predicted <85% | PMID:33872135 | Abdallah | 2021 | Inpatient; mean of 102.3 days post discharge | 20/25 | 80.00% |
| Decreased maximal oxygen uptake | HP:0033760 | VO2 %predicted <85% | PMID:33872135 | Abdallah | 2021 | Outpatient; mean of 129.8 days post diagnosis | 21/38 | 55.30% |
| Decreased maximal oxygen uptake | HP:0033760 | VO2 peak, % of predicted VO2 max < 80% | PMID:33490928 | Ramani | 2021 | Inpatient; mean of 48.0 days post discharge | 28/51 | 54.90% |
| Bradykinesia | HP:0002067 | Bradykinesia | PMID:33682276 | Rass | 2021 | Outpatient; mean of 90.0 days post diagnosis | 0/32 | 0.00% |
| Bradykinesia | HP:0002067 | Bradykinesia | PMID:33682276 | Rass | 2021 | Inpatient (non-ICU); mean of 90.0 days post discharge | 3/72 | 4.20% |
| Bradykinesia | HP:0002067 | Bradykinesia | PMID:33682276 | Rass | 2021 | Inpatient (ICU); mean of 90.0 days post discharge | 4/31 | 12.90% |
| Reduced ejection fraction | HP:0012664 | Left ventricular ejection fraction (%) Impaired (≤51%) | PMID:33785495 | Dennis | 2021 | Inpatient; mean of 138.0 days post discharge | 4/37 | 10.80% |
| Reduced ejection fraction | HP:0012664 | Left ventricular ejection fraction, impaired | PMID:33785495 | Dennis | 2021 | Outpatient; mean of 141.0 days post diagnosis | 7/162 | 4.30% |
| Reduced ejection fraction | HP:0012664 | reduced left ventricular ejection fraction | PMID:33303539 | Sonnweber | 2020 | Mixed; mean of 63.0 days post diagnosis | 4/145 | 2.80% |
| Suicidal ideation | HP:0031589 | Suicidality | PMID:-1 | Davis | 2020 | Mixed; mean of 114.5 days post diagnosis | 436/3762 | 11.60% |
| Suicidal ideation | HP:0031589 | Thoughts of self-harm | PMID:32729939 | Halpin | 2021 | Inpatient (ICU); mean of 48.0 days post discharge | 1/32 | 3.10% |
| Suicidal ideation | HP:0031589 | Thoughts of self-harm | PMID:32729939 | Halpin | 2021 | Inpatient (non-ICU); mean of 48.0 days post discharge | 1/68 | 1.50% |
| Abdominal symptom | HP:0011458 | Digestive disorders | PMID:33031948 | Carvalho-Schneider | 2021 | Mixed; mean of 60.0 days post diagnosis | 15/130 | 11.50% |
| Abdominal symptom | HP:0011458 | Digestive disorders | PMID:33031948 | Carvalho-Schneider | 2021 | Mixed; mean of 30.0 days post diagnosis | 26/150 | 17.30% |
| Abdominal symptom | HP:0011458 | Lower gastro-intestinal symptoms | PMID:33461632 | Venturelli | 2021 | Mixed; mean of 81.0 days post diagnosis | 7/515 | 1.40% |
| Lymphopenia | HP:0001888 | 7.3% of 247 patients had persisting lymphopaenia | PMID:33172844 | Mandal | 2020 | Inpatient; mean of 54.0 days post discharge | 18/247 | 7.30% |
| Lymphopenia | HP:0001888 | Lymphocytes, <1500 per mm3 | PMID:33450302 | Moreno-Perez | 2021 | Mixed; mean of 77.0 days post diagnosis | 55/277 | 19.90% |
| Lymphopenia | HP:0001888 | Lymphocytes <1000 | PMID:33461632 | Venturelli | 2021 | Mixed; mean of 81.0 days post diagnosis | 28/727 | 3.90% |
| Tachycardia | HP:0001649 | Tachycardia | PMID:-1 | Davis | 2020 | Mixed; mean of 114.5 days post diagnosis | 2308/3762 | 61.40% |
| Tachycardia | HP:0001649 | Increased resting heart rate (28%) | PMID:33257910 | Goertz | 2020 | Mixed; mean of 79.0 days post diagnosis | 591/2113 | 28.00% |
| Tachycardia | HP:0001649 | Resting heart rate increase | PMID:32979574 | Xiong | 2021 | Inpatient; mean of 97.0 days post discharge | 60/538 | 11.20% |
| Parkinsonism | HP:0001300 | Parkinsonism | PMID:33682276 | Rass | 2021 | Outpatient; mean of 90.0 days post diagnosis | 0/32 | 0.00% |
| Parkinsonism | HP:0001300 | Parkinsonism | PMID:33682276 | Rass | 2021 | Inpatient (non-ICU); mean of 90.0 days post discharge | 0/72 | 0.00% |
| Parkinsonism | HP:0001300 | Parkinsonism | PMID:33682276 | Rass | 2021 | Inpatient (ICU); mean of 90.0 days post discharge | 01/31/21 | 3.20% |
| Increased circulating ferritin concentration | HP:0003281 | ferritin abnormal | PMID:33755344 | Graham | 2021 | Outpatient; mean of 141.0 days post diagnosis | 02/11/21 | 18.20% |
| Increased circulating ferritin concentration | HP:0003281 | Ferritin > 150 mg/L | PMID:33450302 | Moreno-Perez | 2021 | Mixed; mean of 77.0 days post diagnosis | 112/276 | 40.60% |
| Increased circulating ferritin concentration | HP:0003281 | D-dimer, NT-proBNP, and serum ferritin were still elevated in 27%, 23%, and 17% of COVID-19 patients at second follow-up, respectively | PMID:33303539 | Sonnweber | 2020 | Mixed; mean of 103.0 days post diagnosis | 23/135 | 17.00% |
| Pulmonary bulla | HP:0032446 | Emphysema/Pulmonary bulla | PMID:33540129 | Zhang | 2021 | Inpatient; mean of 90.0 days post discharge | 12/263 | 4.60% |
| Pulmonary bulla | HP:0032446 | Emphysema/Pulmonary bulla | PMID:33540129 | Zhang | 2021 | Inpatient; mean of 180.0 days post discharge | 8/96 | 8.30% |
| Pulmonary bulla | HP:0032446 | Emphysema/Pulmonary bulla | PMID:33540129 | Zhang | 2021 | Inpatient; mean of 28.0 days post discharge | 19/428 | 4.40% |
| Diminished health-related quality of life | HP:0033665 | EQ-5D-5L Decreased by at least 0.05 (MCID;Minimal clinically important difference as validated in respiratory disease) | PMID:32729939 | Halpin | 2021 | Inpatient (ICU); mean of 48.0 days post discharge | 22/32 | 68.80% |
| Diminished health-related quality of life | HP:0033665 | EQ-5D-5L Decreased by at least 0.05 (MCID;Minimal clinically important difference as validated in respiratory disease) | PMID:32729939 | Halpin | 2021 | Inpatient (non-ICU); mean of 48.0 days post discharge | 31/68 | 45.60% |
| Diminished health-related quality of life | HP:0033665 | The most frequently impaired outcomes were quality of life (51%) | PMID:33008936 | Wong | 2020 | Inpatient; mean of 91.0 days post discharge | 40/78 | 51.30% |
| Anorexia | HP:0002039 | Anorexia (Figure 1) | PMID:33252665 | Petersen | 2020 | Outpatient; mean of 99.0 days post diagnosis | 4/180 | 2.20% |
| Anorexia | HP:0002039 | Anorexia | PMID:33729425 | Writing Committee | 2021 | Inpatient (ICU); mean of 93.0 days post discharge | 9/66 | 13.60% |
| Anorexia | HP:0002039 | Anorexia | PMID:33729425 | Writing Committee | 2021 | Inpatient (non-ICU); mean of 121.0 days post discharge | 25/370 | 6.80% |
| Frailty | HP:0033675 | Frail | PMID:33220049 | van den Borst | 2020 | Inpatient (ICU); mean of 91.0 days post discharge | 4/46 | 8.70% |
| Frailty | HP:0033675 | Frail | PMID:33220049 | van den Borst | 2020 | Outpatient; mean of 91.0 days post diagnosis | 01/27/21 | 3.70% |
| Frailty | HP:0033675 | Frail | PMID:33220049 | van den Borst | 2020 | Inpatient (non-ICU); mean of 91.0 days post discharge | 6/51 | 11.80% |
| Shivering | HP:0025144 | Chills or shivering | PMID:33606031 | Logue | 2021 | Inpatient; mean of 169.0 days post discharge | 01/16/21 | 6.30% |
| Shivering | HP:0025144 | Chills or shivering | PMID:33606031 | Logue | 2021 | Outpatient; mean of 169.0 days post diagnosis | 4/161 | 2.50% |
| Abnormal reflex | HP:0031826 | Abnormal reflex status | PMID:33682276 | Rass | 2021 | Inpatient (non-ICU); mean of 90.0 days post discharge | 15/72 | 20.80% |
| Abnormal reflex | HP:0031826 | Abnormal reflex status | PMID:33682276 | Rass | 2021 | Inpatient (ICU); mean of 90.0 days post discharge | 12/31/21 | 38.70% |
| Aphasia | HP:0002381 | Aphasia | PMID:33755344 | Graham | 2021 | Outpatient; mean of 141.0 days post diagnosis | 1/100 | 1.00% |
| Aphasia | HP:0002381 | Aphasia | PMID:33682276 | Rass | 2021 | Outpatient; mean of 90.0 days post diagnosis | 0/32 | 0.00% |
| Elevated circulating aspartate aminotransferase concentration | HP:0031956 | Aspartate transaminase (IU/L), Above reference range | PMID:33393696 | Garg | 2021 | Outpatient; mean of 47.0 days post diagnosis | 01/19/21 | 5.30% |

| | | | | | | | | |
|---|---|---|---|---|---|---|---|---|
| Elevated circulating aspartate aminotransferase concentration | HP:0031956 | AST >126 IU/L (>3xULN) | PMID:33490928 | Ramani | 2021 | Inpatient; mean of 48.0 days post discharge | 0/55 | 0.00% |
| Babinski sign | HP:0003487 | Babinski sign | PMID:33682276 | Rass | 2021 | Inpatient (non-ICU); mean of 90.0 days post discharge | 1/72 | 1.40% |
| Babinski sign | HP:0003487 | Babinski sign | PMID:33682276 | Rass | 2021 | Inpatient (ICU); mean of 90.0 days post discharge | 01/31/21 | 3.20% |
| Pseudo-chilblain | HP:0033696 | COVID toes (discoloration, swelling, painful, or blistering toes) | PMID:-1 | Davis | 2020 | Mixed; mean of 114.5 days post diagnosis | 490/3762 | 13.00% |
| Pseudo-chilblain | HP:0033696 | Red spots on toes/feet (2%) | PMID:33257910 | Goertz | 2020 | Mixed; mean of 79.0 days post diagnosis | 42/2113 | 2.00% |
| Flushing | HP:0031284 | Hot flushes (13%) | PMID:33257910 | Goertz | 2020 | Mixed; mean of 79.0 days post diagnosis | 274/2113 | 13.00% |
| Flushing | HP:0031284 | Discontinuous flushing | PMID:32979574 | Xiong | 2021 | Inpatient; mean of 97.0 days post discharge | 26/538 | 4.80% |
| Pancreatitis | HP:0001733 | Pancreatic inflammation (T1 in ms) Impaired ≥803 ms | PMID:33785495 | Dennis | 2021 | Inpatient; mean of 138.0 days post discharge | 11/33 | 33.30% |
| Pancreatitis | HP:0001733 | Pancreatic inflammation (T1 in ms) | PMID:33785495 | Dennis | 2021 | Outpatient; mean of 141.0 days post diagnosis | 17/163 | 10.40% |
| Hepatic steatosis | HP:0001397 | Liver fat Impaired ≥4.8% | PMID:33785495 | Dennis | 2021 | Inpatient; mean of 138.0 days post discharge | 13/37 | 35.10% |
| Hepatic steatosis | HP:0001397 | Liver fat Impaired ≥4.8% | PMID:33785495 | Dennis | 2021 | Outpatient; mean of 141.0 days post diagnosis | 29/163 | 17.80% |
| Polyneuropathy | HP:0001271 | poly-neuropathy | PMID:33682276 | Rass | 2021 | Outpatient; mean of 90.0 days post diagnosis | 1/32 | 3.10% |
| Polyneuropathy | HP:0001271 | poly-neuropathy | PMID:33682276 | Rass | 2021 | Inpatient (ICU); mean of 90.0 days post discharge | 03/31/21 | 9.70% |
| Hepatomegaly | HP:0002240 | Liver volume Impaired ≥1935 mL | PMID:33785495 | Dennis | 2021 | Inpatient; mean of 138.0 days post discharge | 12/37 | 32.40% |
| Hepatomegaly | HP:0002240 | Liver volume Impaired ≥1935 mL | PMID:33785495 | Dennis | 2021 | Outpatient; mean of 141.0 days post diagnosis | 9/163 | 5.50% |
| Elevated circulating alanine aminotransferase concentration | HP:0031964 | Alanine transaminase (IU/L), Above reference range | PMID:33393696 | Garg | 2021 | Outpatient; mean of 47.0 days post diagnosis | 05/19/21 | 26.30% |
| Elevated circulating alanine aminotransferase concentration | HP:0031964 | ALT >135 IU/L (>3xULN) | PMID:33490928 | Ramani | 2021 | Inpatient; mean of 48.0 days post discharge | 1/58 | 1.70% |
| Irritability | HP:0000737 | Irritability | PMID:-1 | Davis | 2020 | Mixed; mean of 114.5 days post diagnosis | 1924/3762 | 51.10% |
| Irritability | HP:0000737 | Irritability | PMID:33654513 | Suárez-Robles | 2020 | Inpatient; mean of 90.0 days post discharge | 5/134 | 3.70% |
| Short attention span | HP:0000736 | Attention disorder | PMID:32853602 | Garrigues | 2020 | Inpatient (non-ICU); mean of 110.9 days post discharge | 28/96 | 29.20% |
| Short attention span | HP:0000736 | Attention disorder | PMID:32853602 | Garrigues | 2020 | Inpatient (ICU); mean of 110.9 days post discharge | 04/24/21 | 16.70% |
| Increased circulating NT-proBNP concentration | HP:0031185 | Pro-BNP increased | PMID:33450302 | Moreno-Perez | 2021 | Mixed; mean of 77.0 days post diagnosis | 11/276 | 4.00% |
| Increased circulating NT-proBNP concentration | HP:0031185 | D-dimer, NT-proBNP, and serum ferritin were still elevated in 27%, 23%, and 17% of COVID-19 patients at second follow-up, respectively | PMID:33303539 | Sonnweber | 2020 | Mixed; mean of 103.0 days post diagnosis | 31/135 | 23.00% |
| Conjunctivitis | HP:0000509 | Pink eye (conjunctivitis) | PMID:-1 | Davis | 2020 | Mixed; mean of 114.5 days post diagnosis | 560/3762 | 14.90% |
| Conjunctivitis | HP:0000509 | Conjunctivitis | PMID:33273028 | Stavem | 2020 | Outpatient; mean of 117.0 days post diagnosis | 13/451 | 2.90% |
| Hypotonia | HP:0001252 | Decreased muscle tone | PMID:33682276 | Rass | 2021 | Inpatient (non-ICU); mean of 90.0 days post discharge | 0/72 | 0.00% |
| Hypotonia | HP:0001252 | Decreased muscle tone | PMID:33682276 | Rass | 2021 | Inpatient (ICU); mean of 90.0 days post discharge | 01/31/21 | 3.20% |
| Abnormal pulmonary thoracic imaging finding | HP:0031983 | Abnormal CXR | PMID:33731329 | Cheng | 2021 | Mixed; mean of 63.0 days post diagnosis | 66/113 | 58.40% |
| Abnormal pulmonary thoracic imaging finding | HP:0031983 | Chest X-rays score ≥2 | PMID:33450302 | Moreno-Perez | 2021 | Mixed; mean of 77.0 days post diagnosis | 51/269 | 19.00% |
| Elevated myocardial native T1 | HP:4000006 | Elevated myocardial native T1 | PMID:32730619 | Puntmann | 2020 | Mixed; mean of 71.0 days post diagnosis | 73/100 | 73.00% |
| Elevated myocardial native T1 | HP:4000006 | Native T1 (basal myocardium) > 1197 ms (>2SD from control mean) | PMID:33490928 | Ramani | 2021 | Inpatient; mean of 48.0 days post discharge | 13/50 | 26.00% |
| Exercise intolerance | HP:0003546 | 50 patients (21.0%) reported that their tolerance to exercise had worsened after COVID-19. | PMID:33502487 | Bellan | 2021 | Inpatient; mean of 105.0 days post discharge | 50/238 | 21.00% |
| Exercise intolerance | HP:0003546 | When directly questioned, 50 patients (21.0%) reported that their tolerance to exercise had worsened after COVID-19 | PMID:33502487 | Bellan | 2021 | Inpatient; mean of 105.0 days post discharge | 50/238 | 21.00% |
| Myocarditis | HP:0012819 | Evidence of myocarditis ≥3 segments with high T1 (≥1229 ms at 3T; ≥1015 ms at 1.5T) | PMID:33785495 | Dennis | 2021 | Inpatient; mean of 138.0 days post discharge | 8/37 | 21.60% |
| Myocarditis | HP:0012819 | Evidence of myocarditis-≥3 segments with high T1 (≥1229 ms at 3T; ≥1015 ms at 1.5T) | PMID:33785495 | Dennis | 2021 | Outpatient; mean of 141.0 days post diagnosis | 30/163 | 18.40% |
| Skeletal muscle atrophy | HP:0003202 | Muscle atrophy | PMID:33682276 | Rass | 2021 | Inpatient (non-ICU); mean of 90.0 days post discharge | 1/72 | 1.40% |
| Skeletal muscle atrophy | HP:0003202 | Muscle atrophy | PMID:33682276 | Rass | 2021 | Inpatient (ICU); mean of 90.0 days post discharge | 07/31/21 | 22.60% |
| Increased circulating interleukin 6 | HP:0030783 | mild elevations in inflammatory markers such as IL-6 (6%) | PMID:33303539 | Sonnweber | 2020 | Mixed; mean of 103.0 days post diagnosis | 9/145 | 6.20% |
| Increased circulating interleukin 6 | HP:0030783 | increased IL-6 levels in 10/150 | PMID:33587810 | Townsend | 2021 | Mixed; mean of 80.5 days post diagnosis | 10/150 | 6.70% |
| Rhinitis | HP:0012384 | Rhinitis | PMID:32644129 | Carfi | 2020 | Inpatient; mean of 60.3 days post discharge | 20/143 | 14.00% |
| Rhinitis | HP:0012384 | rhinitis | PMID:33479105 | Trinkmann | 2021 | Mixed; mean of 68.0 days post diagnosis | 1/246 | 0.40% |
| Spasticity | HP:0001257 | Spasticity | PMID:33682276 | Rass | 2021 | Inpatient (non-ICU); mean of 90.0 days post discharge | 0/72 | 0.00% |
| Spasticity | HP:0001257 | Spasticity | PMID:33682276 | Rass | 2021 | Inpatient (ICU); mean of 90.0 days post discharge | 01/31/21 | 3.20% |
| Hemoptysis | HP:0002105 | Hemoptysis | PMID:33120193 | Daher | 2020 | Inpatient (ICU); mean of 56.0 days post discharge | 0/33 | 0.00% |
| Hemoptysis | HP:0002105 | Coughing up Blood | PMID:-1 | Davis | 2020 | Mixed; mean of 114.5 days post diagnosis | 194/3762 | 5.20% |
| Increased circulating lactate dehydrogenase concentration | HP:0025435 | Lactate dehydrogenase> 250 U/L | PMID:33450302 | Moreno-Perez | 2021 | Mixed; mean of 77.0 days post diagnosis | 27/274 | 9.90% |
| Increased circulating lactate dehydrogenase concentration | HP:0025435 | LDH >256 | PMID:33461632 | Venturelli | 2021 | Mixed; mean of 81.0 days post diagnosis | 124/727 | 17.10% |

| | | | | | | | | |
|---|---|---|---|---|---|---|---|---|
| Hepatitis | HP:0012115 | Liver inflammation (cT1 in ms) Impaired ≥784 ms | PMID:33785495 | Dennis | 2021 | Inpatient; mean of 138.0 days post discharge | 9/37 | 24.30% |
| Hepatitis | HP:0012115 | Liver inflammation (cT1 in ms) | PMID:33785495 | Dennis | 2021 | Outpatient; mean of 141.0 days post diagnosis | 148/163 | 90.80% |
| Dystonia | HP:0001332 | Dystonia | PMID:33682276 | Rass | 2021 | Outpatient; mean of 90.0 days post diagnosis | 0/32 | 0.00% |
| Dystonia | HP:0001332 | Dystonia | PMID:33682276 | Rass | 2021 | Inpatient (non-ICU); mean of 90.0 days post discharge | 0/72 | 0.00% |
| Hyperglycemia | HP:0003074 | High blood sugar (if measured) | PMID:-1 | Davis | 2020 | Mixed; mean of 114.5 days post diagnosis | 164/3762 | 4.40% |
| Hyperglycemia | HP:0003074 | Fasting blood sugar (mg/dl), Above reference range | PMID:33393696 | Garg | 2021 | Outpatient; mean of 47.0 days post diagnosis | 04/19/21 | 21.10% |
| Decreased glomerular filtration rate | HP:0012213 | eGFR <90 mL/min per 1.73 m2 | PMID:33428867 | Huang | 2021 | Inpatient; mean of 186.0 days post discharge | 487/1393 | 35.00% |
| Decreased glomerular filtration rate | HP:0012213 | eGFR <60 | PMID:33490928 | Ramani | 2021 | Inpatient; mean of 48.0 days post discharge | 3/58 | 5.20% |
| Sneeze | HP:0025095 | Sneezing | PMID:-1 | Davis | 2020 | Mixed; mean of 114.5 days post diagnosis | 989/3762 | 26.30% |
| Sneeze | HP:0025095 | Sneezing (12%) | PMID:33257910 | Goertz | 2020 | Mixed; mean of 79.0 days post diagnosis | 254/2113 | 12.00% |
| Low-grade fever | HP:0011134 | Elevated temperature (98.8-100.4 F) | PMID:-1 | Davis | 2020 | Mixed; mean of 114.5 days post diagnosis | 2188/3762 | 58.20% |
| Low-grade fever | HP:0011134 | Low grade fever | PMID:33428867 | Huang | 2021 | Inpatient; mean of 186.0 days post discharge | 2/1655 | 0.10% |
| Pericardial effusion | HP:0001698 | Pericardial effusion | PMID:32730619 | Puntmann | 2020 | Mixed; mean of 71.0 days post diagnosis | 20/100 | 20.00% |
| Pericardial effusion | HP:0001698 | pericardial effusion | PMID:33303539 | Sonnweber | 2020 | Mixed; mean of 63.0 days post diagnosis | 8/145 | 5.50% |
| Tachypnea | HP:0002789 | Tachypnoea | PMID:33052920 | De Lorenzo | 2020 | Inpatient; mean of 21.5 days post discharge | 33/126 | 26.20% |
| Tachypnea | HP:0002789 | Tachypnoea | PMID:33052920 | De Lorenzo | 2020 | Outpatient; mean of 26.0 days post diagnosis | 8/59 | 13.60% |
| Angina pectoris | HP:0001681 | Angina pectoris | PMID:33120193 | Daher | 2020 | Inpatient (ICU); mean of 56.0 days post discharge | 6/33 | 18.20% |
| Angina pectoris | HP:0001681 | No patient reported typical angina symptoms | PMID:32730619 | Puntmann | 2020 | Mixed; mean of 71.0 days post diagnosis | 0/100 | 0.00% |
| Malnutrition | HP:0004395 | Malnutrition | PMID:33052920 | De Lorenzo | 2020 | Inpatient; mean of 21.5 days post discharge | 5/126 | 4.00% |
| Malnutrition | HP:0004395 | Malnutrition | PMID:33052920 | De Lorenzo | 2020 | Outpatient; mean of 26.0 days post diagnosis | 5/59 | 8.50% |
| Increased left ventricular end-diastolic volume | HP:0033755 | Left ventricular end diastolic volume (mL) >264 mL in Men; >206 mL in Women | PMID:33785495 | Dennis | 2021 | Inpatient; mean of 138.0 days post discharge | 4/37 | 10.80% |
| Increased left ventricular end-diastolic volume | HP:0033755 | Left ventricular end diastolic volume (mL). >264 mL in Men; >206 mL in Women | PMID:33785495 | Dennis | 2021 | Outpatient; mean of 141.0 days post diagnosis | 4/163 | 2.50% |
| Pancreatic steatosis | HP:0033757 | Pancreatic fat Impaired ≥4.6% | PMID:33785495 | Dennis | 2021 | Inpatient; mean of 138.0 days post discharge | 21/37 | 56.80% |
| Pancreatic steatosis | HP:0033757 | Pancreatic fat Impaired ≥4.6% | PMID:33785495 | Dennis | 2021 | Outpatient; mean of 141.0 days post diagnosis | 53/163 | 32.50% |
| Elevated circulating alkaline phosphatase concentration | HP:0003155 | Alkaline phosphatase, Above reference range | PMID:33393696 | Garg | 2021 | Outpatient; mean of 47.0 days post diagnosis | 05/19/21 | 26.30% |
| Elevated circulating alkaline phosphatase concentration | HP:0003155 | Alk Phos >260 IU/L (>2xULN) | PMID:33490928 | Ramani | 2021 | Inpatient; mean of 48.0 days post discharge | 0/58 | 0.00% |
| Rigidity | HP:0002063 | Rigidity | PMID:33682276 | Rass | 2021 | Inpatient (non-ICU); mean of 90.0 days post discharge | 0/72 | 0.00% |
| Rigidity | HP:0002063 | Rigidity | PMID:33682276 | Rass | 2021 | Inpatient (ICU); mean of 90.0 days post discharge | 03/31/21 | 9.70% |
| Visual loss | HP:0000572 | Vision loss | PMID:33564789 | Chun | 2021 | Mixed; mean of 63.0 days post diagnosis | 2/61 | 3.30% |
| Visual loss | HP:0000572 | Visual loss | PMID:33450302 | Moreno-Perez | 2021 | Mixed; mean of 77.0 days post diagnosis | 15/277 | 5.40% |
| Decreased RV/TLC ratio | HP:0033773 | RV /TLC <LLN, n (%) | PMID:33872135 | Abdallah | 2021 | Inpatient; mean of 102.3 days post discharge | 07/25/21 | 28.00% |
| Decreased RV/TLC ratio | HP:0033773 | RV /TLC <LLN, n (%) | PMID:33872135 | Abdallah | 2021 | Outpatient; mean of 129.8 days post diagnosis | 13/38 | 34.20% |
| Hypoxemia | HP:0012418 | pO2 <75 mmHg – no. (%) | PMID:33303539 | Sonnweber | 2020 | Mixed; mean of 63.0 days post diagnosis | 40/126 | 31.70% |
| Hypoxemia | HP:0012418 | pO2 <75 mmHg – no. (%) | PMID:33303539 | Sonnweber | 2020 | Mixed; mean of 103.0 days post diagnosis | 45/133 | 33.80% |
| Diminished physical functioning | HP:0033666 | Overall 66 patients of the study (32%) had at least one test indicating physical impairment | PMID:33565741 | Baricich | 2021 | Inpatient; mean of 124.7 days post discharge | 66/204 | 32.40% |
| Diminished physical functioning | HP:0033666 | With regard to physical function, 53 patients (22.3%) were found to have limited mobility based on SPPB test results. All other patients underwent a 2-minute walk test, which revealed a subtler impairment in 75 patients (31.5%). By this method, we identified 128 patients (53.8%) with some degree of functional impairment | PMID:33502487 | Bellan | 2021 | Inpatient; mean of 105.0 days post discharge | 128/238 | 53.80% |
| Nasal congestion | HP:0001742 | Nasal congestion | PMID:33564789 | Chun | 2021 | Mixed; mean of 63.0 days post diagnosis | 6/61 | 9.80% |
| Nasal congestion | HP:0001742 | Congestion | PMID:32730238 | Tenforde | 2020 | Outpatient; mean of 17.5 days post diagnosis | 82/274 | 29.90% |
| Splenomegaly | HP:0001744 | Splenic volume (mL) Impaired ≥350 mL | PMID:33785495 | Dennis | 2021 | Inpatient; mean of 138.0 days post discharge | 4/37 | 10.80% |
| Splenomegaly | HP:0001744 | Splenic volume (mL) Impaired ≥350 mL | PMID:33785495 | Dennis | 2021 | Outpatient; mean of 141.0 days post diagnosis | 3/163 | 1.80% |
| Night sweats | HP:0030166 | Night sweats | PMID:-1 | Davis | 2020 | Mixed; mean of 114.5 days post diagnosis | 1535/3762 | 40.80% |
| Night sweats | HP:0030166 | Night sweat | PMID:33303539 | Sonnweber | 2020 | Mixed; mean of 103.0 days post diagnosis | 35/145 | 24.10% |
| Bowel incontinence | HP:0002607 | New bowel control problem | PMID:32729939 | Halpin | 2021 | Inpatient (ICU); mean of 48.0 days post discharge | 1/32 | 3.10% |
| Bowel incontinence | HP:0002607 | New bowel control problem | PMID:32729939 | Halpin | 2021 | Inpatient (non-ICU); mean of 48.0 days post discharge | 2/68 | 2.90% |
| Thrombocytopenia | HP:0001873 | Platelet count (×100,000/µl) below reference range | PMID:33393696 | Garg | 2021 | Outpatient; mean of 47.0 days post diagnosis | 04/19/21 | 21.10% |
| Thrombocytopenia | HP:0001873 | convalescent patients had no evidence of hypofibrinogenemia or thrombocytopenia during recovery. | PMID:33587810 | Townsend | 2021 | Mixed; mean of 80.5 days post diagnosis | 0/150 | 0.00% |
| Elevated circulating creatinine concentration | HP:0003259 | Creatinine > 133 umol/L | PMID:33490928 | Ramani | 2021 | Inpatient; mean of 48.0 days post discharge | 1/58 | 1.70% |
| Elevated circulating creatinine concentration | HP:0003259 | Creatinine >1.1 (females),Creatinine >1.3 (males) | PMID:33461632 | Venturelli | 2021 | Mixed; mean of 81.0 days post diagnosis | 54/727 | 7.40% |

| | | | | | | | | |
|---|---|---|---|---|---|---|---|---|
| Elevated circulating creatine kinase concentration | HP:0003236 | CK > 170 U/L | PMID:33450302 | Moreno-Perez | 2021 | Mixed; mean of 77.0 days post diagnosis | 34/276 | 12.30% |
| Postexertional malaise | HP:0030973 | Post-Exertional Malaise | PMID:-1 | Davis | 2020 | Mixed; mean of 114.5 days post diagnosis | 3350/3762 | 89.00% |
| Gaze-evoked nystagmus | HP:0000640 | gaze-evoked nystagmus | PMID:33755344 | Graham | 2021 | Outpatient; mean of 141.0 days post diagnosis | 2/100 | 2.00% |
| Panic attack | HP:0025269 | Panic attack | PMID:33113469 | Akter | 2020 | Inpatient; mean of 28.0 days post discharge | 98/734 | 13.40% |
| Somatic sensory dysfunction | HP:0003474 | Numbness/loss of sensation | PMID:-1 | Davis | 2020 | Mixed; mean of 114.5 days post diagnosis | 1332/3762 | 35.40% |
| Renal insufficiency | HP:0000083 | Renal failure | PMID:33680620 | Iqbal | 2021 | Mixed; mean of 38.1 days post diagnosis | 2/158 | 1.30% |
| Gastroesophageal reflux | HP:0002020 | Lower Esophagus Burning / gastroesophageal reflux / acid reflux | PMID:-1 | Davis | 2020 | Mixed; mean of 114.5 days post diagnosis | 1317/3762 | 35.00% |
| Long term memory impairment | HP:0033688 | Long-term memory loss (long-term memory can be anything from remembering yesterday, forgetting you've done a task, forgetting recently learned information, or forgetting your third-grade experience) | PMID:-1 | Davis | 2020 | Mixed; mean of 114.5 days post diagnosis | 1359/3762 | 36.10% |
| Anterograde memory impairment | HP:0033689 | Inability to make new memories | PMID:-1 | Davis | 2020 | Mixed; mean of 114.5 days post diagnosis | 275/3762 | 7.30% |
| Procedural memory loss | HP:0033691 | Forgetting how to do routine tasks (tying your shoe laces, washing your hands) | PMID:-1 | Davis | 2020 | Mixed; mean of 114.5 days post diagnosis | 453/3762 | 12.00% |
| Phantosmia | HP:0033693 | Phantom smells (imagining/hallucinating smells - smelling things that aren't there) | PMID:-1 | Davis | 2020 | Mixed; mean of 114.5 days post diagnosis | 872/3762 | 23.20% |
| Tactile hallucination | HP:0033694 | Tactile (touch) Hallucinations | PMID:-1 | Davis | 2020 | Mixed; mean of 114.5 days post diagnosis | 116/3762 | 3.10% |
| Constipation | HP:0002019 | Constipation | PMID:-1 | Davis | 2020 | Mixed; mean of 114.5 days post diagnosis | 930/3762 | 24.70% |
| Diplopia | HP:0000651 | Double vision | PMID:-1 | Davis | 2020 | Mixed; mean of 114.5 days post diagnosis | 260/3762 | 6.90% |
| Anti-thyroid peroxidase antibody positivity | HP:0025379 | Thyroid peroxidase Ab >60 | PMID:33461632 | Venturelli | 2021 | Mixed; mean of 81.0 days post diagnosis | 115/727 | 15.80% |
| Abnormality of movement | HP:0100022 | Movement disorder | PMID:33755344 | Graham | 2021 | Outpatient; mean of 141.0 days post diagnosis | 2/100 | 2.00% |
| Male sexual dysfunction | HP:0040307 | Sexual dysfunction - cis men/trans men | PMID:-1 | Davis | 2020 | Mixed; mean of 114.5 days post diagnosis | 105/718 | 14.60% |
| Hypophosphatemia | HP:0002148 | Phosphate, serum (mg/dl) below reference range | PMID:33393696 | Garg | 2021 | Outpatient; mean of 47.0 days post diagnosis | 01/19/21 | 5.30% |
| Hyperkinetic movements | HP:0002487 | Muscle spasms | PMID:-1 | Davis | 2020 | Mixed; mean of 114.5 days post diagnosis | 1222/3762 | 32.50% |
| Visual hallucinations | HP:0002367 | Visual (seeing) Hallucinations | PMID:-1 | Davis | 2020 | Mixed; mean of 114.5 days post diagnosis | 391/3762 | 10.40% |
| Neuralgia | HP:0033345 | Neuralgia (nerve pain) | PMID:-1 | Davis | 2020 | Mixed; mean of 114.5 days post diagnosis | 1177/3762 | 31.30% |
| Euphoria | HP:0031844 | Euphoria (a feeling or state of intense excitement and happiness) | PMID:-1 | Davis | 2020 | Mixed; mean of 114.5 days post diagnosis | 188/3762 | 5.00% |
| Hallucinations | HP:0000738 | Hallucinations, other | PMID:-1 | Davis | 2020 | Mixed; mean of 114.5 days post diagnosis | 87/3762 | 2.30% |
| Irregular menstruation | HP:0000858 | Abnormally irregular periods | PMID:-1 | Davis | 2020 | Mixed; mean of 114.5 days post diagnosis | 474/1792 | 26.50% |
| Decreased circulating calcifediol concentration | HP:0012053 | 25 hydroxy vitamin D, serum (ng/ml), Below reference range | PMID:33393696 | Garg | 2021 | Outpatient; mean of 47.0 days post diagnosis | 09/19/21 | 47.40% |
| Blindness | HP:0000618 | Total loss of vision | PMID:-1 | Davis | 2020 | Mixed; mean of 114.5 days post diagnosis | 37/3762 | 1.00% |
| Photophobia | HP:0000613 | Sensitivity to light | PMID:-1 | Davis | 2020 | Mixed; mean of 114.5 days post diagnosis | 1159/3762 | 30.80% |
| Hypoglycemia | HP:0001943 | Low blood sugar (if measured) | PMID:-1 | Davis | 2020 | Mixed; mean of 114.5 days post diagnosis | 65/3762 | 1.70% |
| Impaired ability to bathe oneself | HP:0031059 | Problems with washing/dressing self | PMID:33680620 | Iqbal | 2021 | Mixed; mean of 38.1 days post diagnosis | 48/158 | 30.40% |
| Sleep apnea | HP:0010535 | Sleep apnea | PMID:-1 | Davis | 2020 | Mixed; mean of 114.5 days post diagnosis | 267/3762 | 7.10% |
| Impulsivity | HP:0100710 | Impulsivity and Disinhibition | PMID:-1 | Davis | 2020 | Mixed; mean of 114.5 days post diagnosis | 451/3762 | 12.00% |
| Vitreous floaters | HP:0100832 | Floaters | PMID:-1 | Davis | 2020 | Mixed; mean of 114.5 days post diagnosis | 755/3762 | 20.10% |
| Impaired ability to dress oneself | HP:0031060 | Problems with washing/dressing self | PMID:33680620 | Iqbal | 2021 | Mixed; mean of 38.1 days post diagnosis | 48/158 | 30.40% |
| Polydipsia | HP:0001959 | Extreme thirst | PMID:-1 | Davis | 2020 | Mixed; mean of 114.5 days post diagnosis | 1346/3762 | 35.80% |
| Hyperacusis | HP:0010780 | Sensitivity to noise | PMID:-1 | Davis | 2020 | Mixed; mean of 114.5 days post diagnosis | 1305/3762 | 34.70% |
| Elevated circulating thyroid-stimulating hormone concentration | HP:0002925 | Thyroid stimulating hormone (TSH, IU/L), Above reference range | PMID:33393696 | Garg | 2021 | Outpatient; mean of 47.0 days post diagnosis | 02/19/21 | 10.50% |
| Recurrent fever | HP:0001954 | Intermittent fever | PMID:33680620 | Iqbal | 2021 | Mixed; mean of 38.1 days post diagnosis | 54/158 | 34.20% |
| Pruritus | HP:0000989 | Itchy, other | PMID:-1 | Davis | 2020 | Mixed; mean of 114.5 days post diagnosis | 192/3762 | 5.10% |
| Delusions | HP:0000746 | Delusions | PMID:-1 | Davis | 2020 | Mixed; mean of 114.5 days post diagnosis | 112/3762 | 3.00% |
| Apathy | HP:0000741 | Apathy (lack of feeling, emotion, interest, or concern) | PMID:-1 | Davis | 2020 | Mixed; mean of 114.5 days post diagnosis | 1473/3762 | 39.20% |
| Attention deficit hyperactivity disorder | HP:0007018 | Attention deficit | PMID:33755344 | Graham | 2021 | Outpatient; mean of 141.0 days post diagnosis | 27/100 | 27.00% |
| Pleuritis | HP:0002102 | Pain/burning feeling in lungs (24%) | PMID:33257910 | Goertz | 2020 | Mixed; mean of 79.0 days post diagnosis | 507/2113 | 24.00% |
| Increased heart rate variability | HP:0031862 | Self-reported rapid variations of heart rate (HR) (24) | PMID:33755344 | Graham | 2021 | Outpatient; mean of 141.0 days post diagnosis | 24/100 | 24.00% |
| Dermatographic urticaria | HP:0011971 | Dermatographia (writing on your skin causes red lines where you scratched) | PMID:-1 | Davis | 2020 | Mixed; mean of 114.5 days post diagnosis | 288/3762 | 7.70% |
| Agnosia | HP:0010524 | Agnosia (failure to recognize or identify objects despite intact sensory functioning) | PMID:-1 | Davis | 2020 | Mixed; mean of 114.5 days post diagnosis | 345/3762 | 9.20% |
| Pericardial late gadolinium enhancement | HP:4000005 | Pericardial late gadolinium enhancement | PMID:32730619 | Puntmann | 2020 | Mixed; mean of 71.0 days post diagnosis | 22/100 | 22.00% |
| Fragile nails | HP:0001808 | Brittle/discolored nail | PMID:-1 | Davis | 2020 | Mixed; mean of 114.5 days post diagnosis | 358/3762 | 9.50% |
| Elevated myocardial native T2 | HP:4000003 | Elevated myocardial native T2 | PMID:32730619 | Puntmann | 2020 | Mixed; mean of 71.0 days post diagnosis | 60/100 | 60.00% |
| Myocardial late gadolinium enhancement | HP:4000004 | Myocardial late gadolinium enhancement | PMID:32730619 | Puntmann | 2020 | Mixed; mean of 71.0 days post diagnosis | 32/100 | 32.00% |
| Aggressive behavior | HP:0000718 | Aggression | PMID:-1 | Davis | 2020 | Mixed; mean of 114.5 days post diagnosis | 280/3762 | 7.40% |
| Emotional lability | HP:0000712 | Mood Lability | PMID:-1 | Davis | 2020 | Mixed; mean of 114.5 days post diagnosis | 1743/3762 | 46.30% |

| | | | | | | | | |
|---|---|---|---|---|---|---|---|---|
| Temperature instability | HP:0005968 | Temperature lability | PMID:-1 | Davis | 2020 | Mixed; mean of 114.5 days post diagnosis | 1539/3762 | 40.90% |
| Gastroparesis | HP:0002578 | gastroparesis | PMID:33755344 | Graham | 2021 | Outpatient; mean of 141.0 days post diagnosis | 2/100 | 2.00% |
| Arthritis | HP:0001369 | Arthritis | PMID:33654513 | Suárez-Robles | 2020 | Inpatient; mean of 90.0 days post discharge | 33/134 | 24.60% |
| Elevated erythrocyte sedimentation rate | HP:0003565 | Erythrocyte sedimentation rate abnormal | PMID:33755344 | Graham | 2021 | Outpatient; mean of 141.0 days post diagnosis | 8/47 | 17.00% |
| Anomic aphasia | HP:0030784 | Difficulty finding the right words while speaking/writing | PMID:-1 | Davis | 2020 | Mixed; mean of 114.5 days post diagnosis | 1743/3762 | 46.30% |
| Gastric ulcer | HP:0002592 | Ulcer | PMID:33306721 | Jacobs | 2020 | Inpatient (non-ICU); mean of 35.0 days post discharge | 2/183 | 1.10% |
| Edema | HP:0000969 | Limb oedema | PMID:32979574 | Xiong | 2021 | Inpatient; mean of 97.0 days post discharge | 14/538 | 2.60% |
| Petechiae | HP:0000967 | Petechiae (tiny purple, red, or brown spots on the skin, usually on arms, legs, stomach, buttocks, and occasionally inside mouth or on eyelids) | PMID:-1 | Davis | 2020 | Mixed; mean of 114.5 days post diagnosis | 671/3762 | 17.80% |
| Hypocalcemia | HP:0002901 | Calcium, serum (mg/dl) below reference range | PMID:33393696 | Garg | 2021 | Outpatient; mean of 47.0 days post diagnosis | 05/19/21 | 26.30% |
| Tearfulness | HP:0033705 | Tearfulness | PMID:-1 | Davis | 2020 | Mixed; mean of 114.5 days post diagnosis | 1599/3762 | 42.50% |
| Slurred speech | HP:0001350 | Slurring words/speech | PMID:-1 | Davis | 2020 | Mixed; mean of 114.5 days post diagnosis | 594/3762 | 15.80% |
| Subpleural curvilinear line | HP:0033702 | Subpleural lines | PMID:32692945 | Liu | 2020 | Inpatient; mean of 28.0 days post discharge | 4/51 | 7.80% |
| Diabetes mellitus | HP:0000819 | Diabetes | PMID:33680620 | Iqbal | 2021 | Mixed; mean of 38.1 days post diagnosis | 1/158 | 0.60% |
| Limb pain | HP:0009763 | limb pain | PMID:33479105 | Trinkmann | 2021 | Mixed; mean of 68.0 days post diagnosis | 2/246 | 0.80% |
| Facial paralysis | HP:0007209 | Facial paralysis (please indicate where onface was paralyzed) | PMID:-1 | Davis | 2020 | Mixed; mean of 114.5 days post diagnosis | 127/3762 | 3.40% |
| Testicular pain | HP:0033839 | Pain in testicles | PMID:-1 | Davis | 2020 | Mixed; mean of 114.5 days post diagnosis | 80/777 | 10.30% |
| Dysphoria | HP:0033838 | Dysphoria | PMID:32979574 | Xiong | 2021 | Inpatient; mean of 97.0 days post discharge | 9/538 | 1.70% |
| Anti-thyroglobulin antibody positivity | HP:0032069 | Anti-Thyroglobulin Ab >60 | PMID:33461632 | Venturelli | 2021 | Mixed; mean of 81.0 days post diagnosis | 61/727 | 8.40% |
| Rest dyspnea | HP:0033710 | Nonmotor polypnoea | PMID:32979574 | Xiong | 2021 | Inpatient; mean of 97.0 days post discharge | 25/538 | 4.60% |
| Elevated circulating soluble CD25 concentration | HP:0033833 | increased sCD25 in only 6/150 | PMID:33587810 | Townsend | 2021 | Mixed; mean of 80.5 days post diagnosis | 6/150 | 4.00% |
| Pulmonary interstitial thickening | HP:0033711 | Interstitial thickening | PMID:32838236 | Zhao | 2020 | Inpatient; mean of 78.5 days post discharge | 15/55 | 27.30% |
| Malaise | HP:0033834 | General malaise | PMID:33654513 | Suárez-Robles | 2020 | Inpatient; mean of 90.0 days post discharge | 25/134 | 18.70% |
| Pulmonary embolism | HP:0002204 | Pulmonary embolism | PMID:33479105 | Trinkmann | 2021 | Mixed; mean of 68.0 days post diagnosis | 02/17/21 | 11.80% |
| Receptive aphasia | HP:0033848 | Difficulty processing/understanding what others say | PMID:-1 | Davis | 2020 | Mixed; mean of 114.5 days post diagnosis | 894/3762 | 23.80% |
| Phantageusia | HP:0033847 | Phantom taste (imagining/hallucinating tastes- tasting things when there's nothing in your mouth) | PMID:-1 | Davis | 2020 | Mixed; mean of 114.5 days post diagnosis | 339/3762 | 9.00% |
| Bilingual aphasia | HP:0033849 | Changes to non-primary (second/third)language skills | PMID:-1 | Davis | 2020 | Mixed; mean of 114.5 days post diagnosis | 191/662 | 28.90% |
| Solid pulmonary nodule | HP:0033609 | Chest CT scan Solid nodule | PMID:33676998 | González | 2021 | Inpatient (ICU); mean of 90.0 days post discharge | 22/57 | 38.60% |
| Postmenopausal bleeding | HP:0033840 | Post-Menopausal bleeding/spotting | PMID:-1 | Davis | 2020 | Mixed; mean of 114.5 days post diagnosis | 63/2061 | 3.10% |
| Body ache | HP:0033047 | Body aches | PMID:32730238 | Tenforde | 2020 | Outpatient; mean of 17.5 days post diagnosis | 47/274 | 17.20% |
| Early satiety | HP:0033842 | Feeling full quickly when eating | PMID:-1 | Davis | 2020 | Mixed; mean of 114.5 days post diagnosis | 1158/3762 | 30.80% |
| Ocular pruritus | HP:0033841 | Itchy eyes | PMID:-1 | Davis | 2020 | Mixed; mean of 114.5 days post diagnosis | 909/3762 | 24.20% |
| Tachyphrenia | HP:0033844 | Thoughts moving too quickly | PMID:-1 | Davis | 2020 | Mixed; mean of 114.5 days post diagnosis | 570/3762 | 15.20% |
| Sense of impending doom | HP:0033845 | | PMID:-1 | Davis | 2020 | Mixed; mean of 114.5 days post diagnosis | 1269/3762 | 33.70% |
| Impaired executive functioning | HP:0033051 | Difficulty with executive functioning (planning, organizing, figuring out the sequence of actions, abstracting) | PMID:-1 | Davis | 2020 | Mixed; mean of 114.5 days post diagnosis | 2166/3762 | 57.60% |
| Ocular pain | HP:0200026 | Eye pressure or pain | PMID:-1 | Davis | 2020 | Mixed; mean of 114.5 days post diagnosis | 992/3762 | 26.40% |
| Menorrhagia | HP:0000132 | Abnormally heavy periods/clotting | PMID:-1 | Davis | 2020 | Mixed; mean of 114.5 days post diagnosis | 355/1762 | 20.10% |
| Coldness | HP:0033850 | Coldness | PMID:-1 | Davis | 2020 | Mixed; mean of 114.5 days post diagnosis | 1261/3762 | 33.50% |
| Subsolid pulmonary nodule | HP:0033610 | Chest CT scan Nonsolid nodule | PMID:33676998 | González | 2021 | Inpatient (ICU); mean of 90.0 days post discharge | 2/57 | 3.50% |
| Auditory hallucinations | HP:0008765 | Auditory (hearing) Hallucinations | PMID:-1 | Davis | 2020 | Mixed; mean of 114.5 days post diagnosis | 244/3762 | 6.50% |
| Expressive aphasia | HP:0002427 | Difficulting speaking in complete sentences | PMID:-1 | Davis | 2020 | Mixed; mean of 114.5 days post diagnosis | 835/3762 | 22.20% |
| Hand muscle weakness | HP:0030237 | Focal motor deficit Hand weakness lasting days to weeks—right-sided (3), left-sided (1). | PMID:33755344 | Graham | 2021 | Outpatient; mean of 141.0 days post diagnosis | 4/100 | 4.00% |
| Pulsatile tinnitus | HP:0008629 | Tinnitus | PMID:33680620 | Iqbal | 2021 | Mixed; mean of 38.1 days post diagnosis | 30/158 | 19.00% |
| Dysmetria | HP:0001310 | Dysmetria | PMID:33682276 | Rass | 2021 | Inpatient (non-ICU); mean of 90.0 days post discharge | 2/72 | 2.80% |
| Intrascapular pain | HP:0033746 | Pain between shoulder blades (33%) | PMID:33257910 | Goertz | 2020 | Mixed; mean of 79.0 days post diagnosis | 697/2113 | 33.00% |
| Restless legs | HP:0012452 | Restless leg syndrome | PMID:-1 | Davis | 2020 | Mixed; mean of 114.5 days post diagnosis | 668/3762 | 17.80% |
| Reduced functional residual capacity | HP:0033750 | FRC <80%, % of predicted | PMID:33428867 | Huang | 2021 | Inpatient; mean of 186.0 days post discharge | 27/332 | 8.10% |
| Xerostomia | HP:0000217 | Sicca syndrome | PMID:32644129 | Carfi | 2020 | Inpatient; mean of 60.3 days post discharge | 21/143 | 14.70% |
| Venous thrombosis | HP:0004936 | Blood clots (Thrombosis) | PMID:-1 | Davis | 2020 | Mixed; mean of 114.5 days post diagnosis | 131/3762 | 3.50% |
| Bone pain | HP:0002653 | Bone ache or burning | PMID:-1 | Davis | 2020 | Mixed; mean of 114.5 days post diagnosis | 910/3762 | 24.20% |
| Increased circulating procalcitonin concentration | HP:0032308 | mild elevations in inflammatory markers such as PCT (9%) w | PMID:33303539 | Sonnweber | 2020 | Mixed; mean of 103.0 days post diagnosis | 13/145 | 9.00% |
| Hypofibrinogenemia | HP:0011900 | convalescent patients had no evidence of hypofibrinogenemia or thrombocytopenia during recovery. | PMID:33587810 | Townsend | 2021 | Mixed; mean of 80.5 days post diagnosis | 0/150 | 0.00% |
| Unilateral facial palsy | HP:0012799 | facial droop | PMID:33755344 | Graham | 2021 | Outpatient; mean of 141.0 days post diagnosis | 1/100 | 1.00% |
| Female sexual dysfunction | HP:0030014 | Sexual dysfunction - cis women/trans women | PMID:-1 | Davis | 2020 | Mixed; mean of 114.5 days post diagnosis | 237/2965 | 8.00% |
| Muscle spasm | HP:0003394 | Muscle spasms | PMID:-1 | Davis | 2020 | Mixed; mean of 114.5 days post diagnosis | 1222/3762 | 32.50% |

| | | | | | | | | |
|---|---|---|---|---|---|---|---|---|
| Keratoconjunctivitis sicca | HP:0001097 | Dry eyes | PMID:-1 | Davis | 2020 | Mixed; mean of 114.5 days post diagnosis | 1077/3762 | 28.60% |
| Centrilobular ground-glass opacification on pulmonary HRCT | HP:0025180 | ground glass opacities | PMID:33479105 | Trinkmann | 2021 | Mixed; mean of 68.0 days post diagnosis | 11/17/21 | 64.70% |
| Phonophobia | HP:0002183 | Sensitivity to noise | PMID:-1 | Davis | 2020 | Mixed; mean of 114.5 days post diagnosis | 1305/3762 | 34.70% |
| Hyperesthesia | HP:0100963 | Hyperanaesthesia | PMID:33205450 | Ludvigsson | 2021 | Outpatient; mean of 210.0 days post diagnosis | 02/05/21 | 40.00% |
| Anaphylactic shock | HP:0100845 | New/unexpected anaphylaxis reaction | PMID:-1 | Davis | 2020 | Mixed; mean of 114.5 days post diagnosis | 153/3762 | 4.10% |
| Pleuritic chest pain | HP:0033771 | Pleuritic pain | PMID:33865161 | Ordinola Navarro | 2021 | Mixed; mean of 40.0 days post diagnosis | 14/115 | 12.20% |
| Hypotension | HP:0002615 | Abnormally low blood pressure | PMID:-1 | Davis | 2020 | Mixed; mean of 114.5 days post diagnosis | 443/3762 | 11.80% |
| Bradycardia | HP:0001662 | Bradycardia (low heart rate, <60 beats per minute) | PMID:-1 | Davis | 2020 | Mixed; mean of 114.5 days post diagnosis | 658/3762 | 17.50% |
| Migraine | HP:0002076 | Migraines | PMID:-1 | Davis | 2020 | Mixed; mean of 114.5 days post diagnosis | 872/3762 | 23.20% |
| Sleep onset insomnia | HP:0031354 | Difficulty falling asleep | PMID:-1 | Davis | 2020 | Mixed; mean of 114.5 days post diagnosis | 1489/3762 | 39.60% |
| Maintenance insomnia | HP:0031355 | Waking up several times during the night | PMID:-1 | Davis | 2020 | Mixed; mean of 114.5 days post diagnosis | 1791/3762 | 47.60% |
| Elevated gamma-glutamyltransferase level | HP:0030948 | GGT >80 IU/L (>2xULN) | PMID:33490928 | Ramani | 2021 | Inpatient; mean of 48.0 days post discharge | 6/54 | 11.10% |
| Terminal insomnia | HP:0031356 | Waking up early in the morning | PMID:-1 | Davis | 2020 | Mixed; mean of 114.5 days post diagnosis | 936/3762 | 24.90% |
| Crazy-paving pattern | HP:0033659 | Crazy paving | PMID:32838236 | Zhao | 2020 | Inpatient; mean of 78.5 days post discharge | 3/55 | 5.50% |
| Scaling skin | HP:0040189 | Peeling skin | PMID:-1 | Davis | 2020 | Mixed; mean of 114.5 days post diagnosis | 488/3762 | 13.00% |
| Peripheral visual field loss | HP:0007994 | Tunnel vision | PMID:-1 | Davis | 2020 | Mixed; mean of 114.5 days post diagnosis | 125/3762 | 3.30% |
| Rhonchi | HP:0030831 | Rattling of breath | PMID:-1 | Davis | 2020 | Mixed; mean of 114.5 days post diagnosis | 641/3762 | 17.00% |
| Hypothermia | HP:0002045 | Low temperature | PMID:-1 | Davis | 2020 | Mixed; mean of 114.5 days post diagnosis | 776/3762 | 20.60% |
| Heat intolerance | HP:0002046 | Heat intolerance | PMID:-1 | Davis | 2020 | Mixed; mean of 114.5 days post diagnosis | 1024/3762 | 27.20% |
| Antinuclear antibody positivity | HP:0003493 | Antinuclear antibody ≥ 1:160 | PMID:33755344 | Graham | 2021 | Outpatient; mean of 141.0 days post diagnosis | 11/33 | 33.30% |
| Increased circulating troponin T concentration | HP:0410174 | Troponin T, > 14 ng/L | PMID:33450302 | Moreno-Perez | 2021 | Mixed; mean of 77.0 days post diagnosis | 40/275 | 14.50% |
| Increased circulating troponin I concentration | HP:0410173 | High-sensitivity troponin T values were detectable (3 pg/mL or greater) in 71 patients recently recovered from COVID-19 (71%) and significantly elevated (13.9 pg/mL or greater) in 5 (5%). | PMID:32730619 | Puntmann | 2020 | Mixed; mean of 71.0 days post diagnosis | 5/100 | 5.00% |
| Atelectasis | HP:0100750 | Chest CT Scan Atelectasis | PMID:33676998 | González | 2021 | Inpatient (ICU); mean of 90.0 days post discharge | 14/57 | 24.60% |
| Diminished mental health | HP:0033667 | Emotionally affected at least mildly by health conditions | PMID:33175566 | Chopra | 2020 | Inpatient; mean of 60.0 days post discharge | 238/488 | 48.80% |
| Lymphadenopathy | HP:0002716 | Enlarged lymph nodes | PMID:33273028 | Stavem | 2020 | Outpatient; mean of 117.0 days post diagnosis | 5/451 | 1.10% |
| Airway obstruction | HP:0006536 | four (15.38%) had obstruction,- one (3.85%) had mixed obstruction and restriction | PMID:32835708 | Ramani | 2021 | Inpatient (ICU); mean of 39.5 days post discharge | 05/28/21 | 17.90% |
| Mania | HP:0100754 | Mania (abnormally elevated/excited mood, decreased need for sleep, occasionally with delusions) | PMID:-1 | Davis | 2020 | Mixed; mean of 114.5 days post diagnosis | 96/3762 | 2.60% |
