## Supplemental File 3 for "Characterizing Long COVID: Deep Phenotype of a Complex Condition"

neuropsychiatric-speech-language

| HPO Term | Definition | Synonyms | Plain-language label | Plain-language definition |
|---|---|---|---|---|
| Anomic aphasia (HP:0030784) | An inability to name people and objects that are correctly perceived. The individual is able to describe the object in question, but cannot provide the name. | Amnesic aphasia; Amnestic aphasia; Anomia; Nominal aphasia | Word-finding difficulty | Trouble naming objects (inability to find the right word) when speaking and writing. |
| Aphasia (HP:0002381) | An acquired language impairment of some or all of the abilities to produce or comprehend speech and to read or write. | Losing words; Loss of words; Difficulty finding words | Aphasia | Loss of ability to understand or express speech. |
| Bilingual aphasia (HP:0033849) | The term bilingual aphasia is used to refer to aphasia in persons who speak two or more languages. When a multilingual speaker has aphasia following a stroke, the languages spoken premorbidly may show comparable or differential patterns of impairment. Differential patterns may manifest as greater impairment in one language compared to | - | Changes to second language skills | Changes to non-primary (second/third) language skills such as difficulty to find the right word or to understand others, whereby the changes are more pronounced than with the primary language. |

| | | | | |
|---|---|---|---|---|
| | another, or as differences in the characteristics of aphasia. Clinical reports of bilingual aphasia show dissociations in the processing of the language learned first (L1) and and second (L2), with one language more impaired than the other. Other cases show a pattern of differential recovery where L2 is recovered only after L1. Another pattern is alternating antagonism; i.e., patients access one language in spontaneous speech and inhibit the other language for alternating periods. This term should be used for a type of aphasia in a person who speaks multiple languages in which the impairment is different for different languages. | | | |
| Expressive aphasia (HP:0002427) | Impairment of expressive language and relative preservation of receptive | Broca's aphasia; Motor aphasia; Loss of expressive speech | Non-fluent aphasia | Difficulty speaking or writing complete sentences with correct grammar. |

| | | | | |
|---|---|---|---|---|
| | language abilities. That is, the patient understands language (speech, writing) but cannot express it. | | | |
| Receptive aphasia (HP:0033848) | A type of aphasia that is characterized by impaired language comprehension. | Wernicke aphasia. | Fluent aphasia | Reduced ability to understand spoken or written language. |
| Slurred speech (HP:0001350) | Abnormal coordination of muscles involved in speech. | - | Slurred speech | Slurring words/speech. |

HEENT-ENT

| HPO Term | Definition | Synonyms | Plain-language label | Plain-language definition |
|---|---|---|---|---|
| Dysphonia (HP:0001618) | An impairment in the ability to produce voice sounds. | Inability to produce voice sounds; Voice change | Voice change | A change in the sound or quality of the voice. |
| Nasal congestion (HP:0001742) | Reduced ability to pass air through the nasal cavity often leading to mouth breathing. | Nasal blockage; Nasal obstruction; Blockage of nose; Stuffy nose; Congestion of nose; Obstruction of nose | Stuffy nose | Stuffy nose. |
| Pharyngalgia (HP:0033050) | An unpleasant sensation characterized by physical discomfort (such as pricking, throbbing, or aching) and perceived to originate in the throat. | Pharyngodynia; Throat discomfort; Throat pain; Sore throat | Sore throat | Sore throat. |
| Rhinitis (HP:0012384) | Inflammation of the nasal mucosa with nasal congestion. | Nasal inflammation | Runny nose | Runny nose. |

neuropsychiatric-cognitive-dysfunction

| HPO Term | Definition | Synonyms | Plain-language label | Plain-language definition |
|---|---|---|---|---|
| Agnosia (HP:0010524) | Inability to recognize objects not because of sensory deficit but because of the inability to combine components of sensory impressions into a complete pattern. Thus, agnosia is a neurological condition which results in an inability to know, to name, to identify, and to extract meaning from visual, auditory, or tactile impressions. | - | Agnosia | Failure to recognize or identify objects despite intact sensory functioning. |
| Bradykinesia (HP:0002067) | Bradykinesia literally means slow movement, and is used clinically to denote a slowness in the execution of movement (in contrast to hypokinesia, which is used to refer to slowness in the initiation of movement). | Slowness of movements; Slow movements | Bradykinesia | Abnormally slow movements |
| Bradyphrenia (HP:0031843) | Abnormal slowness of thought processes. | Slowed thinking; Slowness of thought; Mental slowness; Slowed thoughts | Mental slowness | Slowness of thought. |
| Cognitive impairment (HP:0100543) | Abnormal cognition with deficits in | Cognitive abnormality; Cognitive defects; | Difficulty thinking | Reduced ability to think clearly or process |

| | | | | |
|---|---|---|---|---|
| | thinking, reasoning, or remembering. | Intellectual impairment; Cognitive deficits; Abnormality of cognition; An individual with cognitive impairment may have trouble remembering, learning new things, concentrating, or making decisions. | | information with troubles remembering, learning new things, concentrating, or making decisions. |
| Confusion (HP:0001289) | Lack of clarity and coherence of thought, perception, understanding, or action. | Easily confused; Disorientation; Mental disorientation | Confusion | Inability to think clearly and quickly. |
| Diminished ability to concentrate (HP:0031987) | Being unable to focus one's attention or mental effort on a particular object or activity. | Lack of concentration; Concentration problems; Poor concentration | Poor concentration | Being unable to focus one's attention or mental effort on a particular object or activity. |
| Encephalopathy (HP:0001298) | Encephalopathy is a term that means brain disease, damage, or malfunction. In general, encephalopathy is manifested by an altered mental state. | - | Brain disease | A general term that means brain disease, damage, or malfunction. |
| Tachyphrenia (HP:0033844) | The sensation that thoughts are moving too quickly. | Racing thoughts | Racing thoughts | The sensation that thoughts are moving too quickly. |

skin-findings

| HPO Term | Definition | Synonyms | Plain-language label | Plain-language definition |
|---|---|---|---|---|
| Alopecia (HP:0001596) | A noncongenital process of hair loss, which may progress to partial or complete baldness. | Hair loss | Hair loss | Hair loss. |
| Dermatographic urticaria (HP:0011971) | An exaggerated whealing tendency when the skin is stroked, that is, formation of red, itchy bumps and lines on the skin as a result of pressure on the skin (for instance, stroking the skin with a pen or tongue depressor). | Dermatographism; Skin writing; Dermographism | Skin writing | A condition in which any scratch or pressure applied to the skin results in an exaggerated, rash-like reaction. |
| Flushing (HP:0031284) | Recurrent episodes of redness of the skin together with a sensation of warmth or burning of the affected areas of skin. | - | Flushing | Involuntary, temporary reddening of the skin of the face or neck. |
| Fragile nails (HP:0001808) | Nails that easily break. | Brittle nails | Brittle nails | Nails that easily break. |
| Hyperhidrosis (HP:0000975) | Abnormal excessive perspiration (sweating) despite the lack of appropriate stimuli like hot and humid weather. | Diaphoresis; Sweating; Excessive sweating; Sweating profusely; Sweating, increased; Increased sweating; Profuse sweating | Excessive sweating | Excessive sweating even when weather is not hot |
| Petechiae (HP:0000967) | Petechiae are pinpoint-sized | - | Tiny purple spots on skin | Pinpoint-sized reddish or |

| | | | | |
|---|---|---|---|---|
| | reddish/purple spots, resembling a rash, that appear just under the skin or a mucous membrane when capillaries have ruptured and some superficial bleeding into the skin has happened. This term refers to an abnormally increased susceptibility to developing petechiae. | | | purplish spots containing blood that appear in the skin or mucous membrane as a result of localized bleeding. |
| Pruritus (HP:0000989) | Pruritus is an itch or a sensation that makes a person want to scratch. This term refers to an abnormally increased disposition to experience pruritus. | Skin itching; Itching; Itchy skin; pruritis | Itchy skin | Itchy skin. |
| Pseudo-chilblain (HP:0033696) | Acral areas of erythema with vesicles or pustules. The lesions resemble chilblains and have purpuric areas, affecting hands and feet. | COVID toe; Chilblain-like lesion | COVID toe | COVID toe (swelling, purple or reddish patches, blisters) |
| Scaling skin (HP:0040189) | Refers to the loss of the outer layer of the epidermis in large, scale-like flakes. | peeling skin; flaking skin; Scaly skin; Desquamation | Peeling skin | Peeling skin |
| Skin rash (HP:0000988) | A red eruption of the skin. | Rash | Rash | Rash. |

pulmonary-imaging

| HPO Term | Definition | Synonyms | Plain-language label | Plain-language definition |
|---|---|---|---|---|
| Abnormal pulmonary thoracic imaging finding (HP:0031983) | This term groups terms representing abnormal findings derived from chest X-ray investigation of the lung. In general, lung abnormalities can manifest as opacities (areas of increased density) or as regions with decreased density. | Abnormal chest radiograph finding (lung) | Abnormal lung radiology result | Abnormal results of chest X-ray, computed tomography (CT), or magnetic resonance imaging (MRI) of the lung. |
| Atelectasis (HP:0100750) | Collapse of part of a lung associated with absence of inflation (air) of that part. | Pulmonary atelectasis; Partial or complete collapse of part or entire lung | Atelectasis | Collapse of part of a lung. |
| Bronchiectasis (HP:0002110) | Persistent abnormal dilatation of the bronchi owing to localized and irreversible destruction and widening of the large airways. | Permanent enlargement of the airways of the lungs | Bronchiectasis | Permanent enlargement of the airways of the lungs. |
| Centrilobular ground-glass opacification on pulmonary HRCT (HP:0025180) | A hazy area of increased attenuation in centrilobular areas of the lung with preserved bronchial and vascular markings seen on a computer tomography scan. Centrilobular refers to a location that is | Centrilobular groundglass opacity; Centrilobular groundglass opacification | Centrilobular ground-glass opacification | Areas of haziness (moderately increased whiteness) in the centers of lung lobules (parts of the lung). This feature can be observed by computed tomography of the lung. |

| | | | | |
|---|---|---|---|---|
| | central within secondary pulmonary lobules. | | | |
| Crazy-paving pattern (HP:0033659) | This pattern appears as thickened interlobular septa and intralobular lines superimposed on a background of ground-glass opacity, resembling irregularly shaped paving stones. The crazy-paving pattern is often sharply demarcated from more normal lung and may have a geographic outline. It was originally reported in patients with alveolar proteinosis and is also encountered in other diffuse lung diseases that affect both the interstitial and airspace compartments, such as lipoid pneumonia. | - | Crazy paving | An abnormal finding seen on computer tomography of the lung that is said to resemble paving stones. |
| Interlobular septal thickening (HP:0030879) | Presence of thickening of the interlobular septa of the lungs as seen on a CT scan. | Short lines (pumonary CT finding); Peripheral lines (pulmonary CT finding); Septal thickening (pulmonary CT finding); Septal | Interlobular septal thickening | an abnormal finding on lung computed tomography (CT) whereby the normally invisible separations between subunits of the lung |

| | | | | |
|---|---|---|---|---|
| | | lines (pulmonary CT finding); Interlobular lines (pulmonary CT finding); Interlobular septal thickening on pulmonary HRCT | | (lobules) appear as thickened lines. |
| Parenchymal consolidation (HP:0032177) | Consolidation refers to an exudate or other product of disease that replaces alveolar air, rendering the lung solid (as in infective pneumonia). | - | Consolidation | An area of complete whiteness in a chest X-ray or computed tomography (CT scan) that is typically observed when the air sacs in the lungs are filled with fluid, pus, or other material rather than air. |
| Pleural thickening (HP:0031944) | An increase in the thickness of the pleura, generally related to scarring of the pleural tissue. | Pleural incrassation | Pleural thickening | An increase in the thickness of the lining of the lungs (pleura), which can occur because of scarring. |
| Pulmonary bulla (HP:0032446) | Pulmonary bullae are rounded focal regions of emphysema with a thin wall which measure more than 1 cm in diameter. They are often subpleural in location and are typically larger in the apices. In some cases, bullae can be very large and result in compression of adjacent lung tissue. A giant | Pulmonary bullae | Lung bulla | A relatively large cavity within the lung that can be seen as a dark bubble on chest X-ray or computed tomography. |

| | | | | |
|---|---|---|---|---|
| | bulla is arbitrarily defined as one that occupies at least one third of the volume of a hemithorax. When large, bullae can simulate pneumothorax. The most common cause is paraseptal emphysema but bullae may also be seen in association with centrilobular emphysema. | | | |
| Pulmonary fibrosis (HP:0002206) | Replacement of normal lung tissues by fibroblasts and collagen. | - | Lung scarring | Replacement of normal lung tissue by scar tissue. |
| Pulmonary interstitial thickening (HP:0033711) | Pathological thickening of the pulmonary interstitium visualized radiographically and divided into interlobular and intralobular septal thickening. | - | Pulmonary interstitial thickening | An abnormal finding in chest X-ray or computed tomography (CT) that appears as patchy whitish areas. |
| Reticular pattern on pulmonary HRCT (HP:0025390) | On pulmonary high-resolution computed tomography, reticular pattern is characterised by innumerable interlacing shadows suggesting a mesh. | - | Reticular pattern | An abnormal finding on lung computed tomography (CT) that resembles a network of shadows on the lung. |
| Solid pulmonary nodule | A type of pulmonary nodule | - | Solid lung nodule | A small round or oval spot with a |

| | | | | |
|---|---|---|---|---|
| (HP:0033609) | whose density obscures the underlying parenchyma and thus has a "solid" appearance. | | | solid in the lung with a completely opaque (white) appearance in the lung seen on chest X-ray, CT, or MRI. |
| Subsolid pulmonary nodule (HP:0033610) | Pulmonary subsolid nodules (SSNs) refer to pulmonary nodules with pure ground-glass nodules and part-solid ground-glass nodules. A ground-glass nodule (GGN) is the morphologic description of a pulmonary nodule category on thin-section chest computed tomography (CT). During the past decade, the natural history, management strategy and long-term prognosis in the case of GGNs have attracted attention. | - | Subsolid lung nodule | A small round or oval spot with a solid in the lung with a partially opaque (white)/partially transparent appearance in the lung seen on chest X-ray, CT, or MRI. |
| Subpleural curvilinear line (HP:0033702) | This finding is a thin curvilinear opacity, 1-3 mm in thickness, lying less than 1 cm from and parallel to the pleural surface. It corresponds to atelectasis of normal lung if seen in the dependent posteroinferior | - | Subpleural curvilinear line | An abnormal chest computed tomography (CT) finding that appears as a curvy white line near the surface of the lung (i.e., near the pleura). |

| | |
|---|---|
| | portion of lung of a patient in the supine position and is subsequently shown to disappear on CT sections acquired with the patient prone. It may also be encountered in patients with pulmonary edema or fibrosis (other signs are usually present). |

reproductive-genitourinary-endocrine-metabolism

| HPO Term | Definition | Synonyms | Plain-language label | Plain-language definition |
|---|---|---|---|---|
| Decreased glomerular filtration rate (HP:0012213) | An abnormal reduction in the volume of fluid filtered out of plasma through glomerular capillary walls into Bowman's capsules per unit of time. | Decreased GFR; Reduced creatinine clearance; Impaired renal creatinine clearance | Decreased GFR | A reduction in the filter function of the kidneys (i.e., in the function of the glomeruli, the tiny filters of the kidney that normally remove waste from the blood). |
| Diabetes mellitus (HP:0000819) | A group of abnormalities characterized by hyperglycemia and glucose intolerance. | - | Diabetes | A disease characterized by high blood sugar. |
| Edema (HP:0000969) | An abnormal accumulation of fluid beneath the skin, or in one or more cavities of the body. | Dropsy; Oedema; Hydrops; Water retention; Fluid retention | Water retention | Swelling of a body part related to leakage of fluid from small blood vessels into tissues. |
| Female sexual dysfunction (HP:0030014) | A problem occurring during any phase of the female sexual response cycle that prevents the individual from experiencing satisfaction from the sexual activity | - | Female sexual dysfunction | Problems with sexual response, desire, orgasm, or pain. |
| Fever (HP:0001945) | Body temperature elevated above the normal range. | Pyrexia; Hyperthermia | Fever | Body temperature above the normal range. |
| Heat intolerance (HP:0002046) | The inability to maintain a comfortable body temperature in warm or hot weather. | Intolerance to heat and fevers | Heat intolerance | The inability to maintain a comfortable body temperature in warm or hot weather. |
| Hypothermia (HP:0002045) | Reduced body temperature due to failed | Abnormally low body temperature | Low body temperature | Abnormally low body temperature, |

| | | | | |
|---|---|---|---|---|
| | thermoregulation. | | | defined as 35.0 degrees C (95.0 degrees F) or less. |
| Irregular menstruation (HP:0000858) | Abnormally high variation in the amount of time between periods. | Menstrual irregularity; Irregular periods; Menstrual irregularities; Irregular menses | Irregular menstruation | Increased variation in the amount of time between periods. |
| Low-grade fever (HP:0011134) | Mild fever that does not exceed 38.5 degrees centigrade. | Mild fever | Mild fever | Mild fever that does not exceed 38.5 degrees centigrade. |
| Male sexual dysfunction (HP:0040307) | A problem occurring during any phase of the male sexual response cycle that prevents the individual from experiencing satisfaction from the sexual activity | - | Male sexual dysfunction | Problems with sexual response, desire, orgasm, or pain. |
| Menorrhagia (HP:0000132) | Prolonged and excessive menses at regular intervals in excess of 80 mL or lasting longer than 7 days. | Abnormally heavy periods; Hypermenorrhea; Abnormally heavy bleeding during menstruation | Abnormally heavy periods | Abnormally heavy bleeding during menstruation. |
| Pancreatitis (HP:0001733) | The presence of inflammation in the pancreas. | Pancreatic inflammation | Pancreatitis | Inflammation of the pancreas |
| Postmenopausal bleeding (HP:0033840) | Vaginal bleeding that occurs after 1 year of amenorrhea in a woman who is not receiving hormone therapy. | - | Postmenopausal bleeding | Bleeding in a woman who has underwent menopause. |
| Recurrent fever (HP:0001954) | Periodic (episodic or recurrent) bouts of fever. | Increased body temperature, episodic; Episodic fever; Hyperthermia, episodic; Intermittent fever | Periodic fever | Periodic (episodic or recurrent) bouts of fever. |

| | | | | |
|---|---|---|---|---|
| Renal insufficiency (HP:0000083) | A reduction in the level of performance of the kidneys in areas of function comprising the concentration of urine, removal of wastes, the maintenance of electrolyte balance, homeostasis of blood pressure, and calcium metabolism. | Renal failure; Renal failure in adulthood | Renal failure | Reduced function of the kidney. |
| Temperature instability (HP:0005968) | Disordered thermoregulation characterized by an impaired ability to maintain a balance between heat production and heat loss, with resulting instability of body temperature. | Body temperature instability | Body temperature instability | Instability of body temperature. |
| Testicular pain (HP:0033839) | An unpleasant sensation characterized by physical discomfort (such as pricking, throbbing, or aching) localized to one or both testes. | Pain in testicles | Pain in testicles | Pain in one or both testicles. |
| Urinary incontinence (HP:0000020) | Loss of the ability to control the urinary bladder leading to involuntary urination. | Bladder incontinence; Loss of bladder control | Urinary incontinence | Not able to control urine/pee; Urine/pee leak. |

pulmonary-finding

| HPO Term | Definition | Synonyms | Plain-language label | Plain-language definition |
|---|---|---|---|---|
| Airway obstruction (HP:0006536) | Obstruction of conducting airways of the lung. | Obstructive lung disease; Pulmonary obstruction | Airway obstruction | Reduction of the ability of the airways of the lung to transport air. |
| Decreased DLCO (HP:0045051) | Reduced ability of the lungs to transfer gas from inspired air to the bloodstream as measured by the diffusing capacity of the lungs for carbon monoxide (DLCO) test. | Decreased diffusing capacity | Decreased diffusing capacity in the lungs | Reduced ability of the lungs to transfer gas from inhaled air to the blood. |
| Decreased maximal oxygen uptake (HP:0033760) | Maximum oxygen uptake (VO2max) is defined as the highest rate of oxygen uptake and utilization by the body during intense, maximal exercise, whereby further increases in work rate do not bring on additional rises in VO2 (i.e. plateau). VO2Max is typically measured with a treadmill anad ergometer and the participant exercises with increasing levels of intensity. VO2Max is the point at which oxygen uptake no longer increases despite an increase in | - | Decreased maximal oxygen uptake | A reduced maximum rate of oxygen uptake by the lungs. |

| | | | | |
|---|---|---|---|---|
| | workload. | | | |
| Decreased RV/TLC ratio (HP:0033773) | An abnormally low ratio of residual volume (RV) to total lung capacity (TLC) on pulmonary function testing. RV is the amount of air remaining after maximal expiration and TLC is the total amount of air in the lungs at full inspiration. These volumes cannot be determined by spirometry, but can be measured by inert gas dilution, nitrogen washout, and body plethysmography. | - | Decreased RV/TLC ratio | Reduced ratio of the amount of air left in lungs after breathing out as compared to the total amount of air in the lungs after maximal inhalation. |
| Ground-glass opacification (HP:0025179) | On chest radiographs, ground-glass opacity appears as an area of hazy increased lung opacity, usually extensive, within which margins of pulmonary vessels may be indistinct. On CT scans, it appears as hazy increased opacity of lung, with preservation of bronchial and vascular margins. It is caused by partial filling of airspaces, interstitial thickening (due to | Ground-glass opacification on pulmonary HRCT | Ground-glass opacification | Areas of haziness (moderately increased whiteness) in the lung as seen on chest X-ray, CT, or MRI. |

| | | | | |
|---|---|---|---|---|
| | fluid, cells, and/or fibrosis), partial collapse of alveoli, increased capillary blood volume, or a combination of these, the common factor being the partial displacement of air. Ground-glass opacity is less opaque than consolidation, in which bronchovascular margins are obscured. | | | |
| Hypoxemia (HP:0012418) | An abnormally low level of blood oxygen. | Low blood oxygen level; Hypoxia | Low blood oxygen | An abnormally low level of oxygen in the blood. |
| Oxygen desaturation on exertion (HP:0030874) | Oxygen saturation less than 95% on exertion or arterial partial pressure of oxygen falling by more than 1kPa. | O2 desaturation on exertion | Oxygen desaturation on exertion | A drop in oxygen level that occurs in response to physical activity. |
| Reduced FEV1/FVC ratio (HP:0030877) | Abnormally low FEV1/FVC (FEV1 - forced expiratory volume in 1 second; FVC forced vital capacity). | Obstructive deficit on pulmonary function test; Obstructive deficit on pulmonary function testing | Reduced FEV1/FVC ratio | A reduced ratio of the amount of air a patient can breathe out in one second as compared to the total amount of air the patient can breathe out. |
| Reduced forced expiratory volume in one second (HP:0032342) | | An abnormal reduction in the amount of air a person can forcefully expel in one second. | Reduced FEV1 | A reduction in the amount of air a patient can forcefully breathe out in one second. |
| Reduced forced vital capacity (HP:0032341) | An abnormal reduction in the amount of air a | Reduced FVC; Decreased forced vital capacity | Reduced FVC | A reduction in the amount of air a patient can |

| | | | | |
|---|---|---|---|---|
| | person can expel following maximal inspiration. | | | forcefully breathe out. |
| Reduced functional residual capacity (HP:0033750) | An abnormal reduction in the volume remaining in the lungs after a normal, passive exhalation. | - | Reduced FRC | A reduction in the amount of air left in lungs after a normal expiration. |
| Reduced residual volume (HP:0033753) | Abnormal decrease in the amount of air remaining in a person's lungs after full exhalation. | - | Reduced RV | A reduction in the amount of air left in lungs after complete expiration. |
| Reduced total lung capacity (HP:0033169) | | Reduced TLC; Abnormally reduced volume of air in the lungs upon the maximum effort of inspiration. | Reduced TLC | A reduction in the amount of air a patient can inhale with maximum effort. |
| Restrictive ventilatory defect (HP:0002091) | A functional defect characterized by reduced total lung capacity (TLC) not associated with abnormalities of expiratory airflow or airway resistance. Spirometrically, a restrictive defect is defined as FEV1 (forced expiratory volume in 1 second) and FVC (forced vital capacity) less than 80 per cent. Restrictive lung disease may be caused by alterations in lung parenchyma or | Restrictive respiratory disease; Restrictive respiratory insufficiency; Spirometric restriction; Restrictive lung disease; Restrictive deficit on pulmonary function tests; Restrictive deficit on pulmonary function testing; Stiff lung or chest wall causing decreased lung volume; Restrictive respiratory syndrome | Restrictive lung disease | A restrictive lung disease, meaning that the lung can not expand as much as a normal lung does, which leads to problems with air flow. |

| | | | | |
|---|---|---|---|---|
| | because of a disease of the pleura, chest wall, or neuromuscular apparatus. | | | |
| Pleuritis (HP:0002102) | Inflammation of the pleura. | Pleurisy; Inflammation of tissues lining lungs and chest | Pleuritis | Inflammation of the lining of the lungs and chest cavity (pleura). |
| Pulmonary embolism (HP:0002204) | An embolus (that is, an abnormal particle circulating in the blood) located in the pulmonary artery and thereby blocking blood circulation to the lung. Usually the embolus is a blood clot that has developed in an extremity (for instance, a deep venous thrombosis), detached, and traveled through the circulation before becoming trapped in the pulmonary artery. | Blood clot in artery of lung | Lung embolism | Blood clot in artery of lung. |

immunology-autoimmunity

| HPO Term | Definition | Synonyms | Plain-language label | Plain-language definition |
|---|---|---|---|---|
| Anaphylactic shock (HP:0100845) | An acute hypersensitivity reaction due to exposure to a previously encountered antigen. | Anaphylaxis | Anaphylaxis | A severe allergic reaction that may result in swelling, hives, lowered blood pressure and in severe cases, shock. |
| Antinuclear antibody positivity (HP:0003493) | The presence of autoantibodies in the serum that react against nuclei or nuclear components. | Antinuclear antibody positive; Serum antinuclear antibody; Antinuclear antibodies; Elevated antinuclear antibody | Antinuclear antibody | An autoantibody is a kind of antibody that is directed against a substance made by one's own body (usually, antibodies are directed against external agents such as bacteria). This term refers to autoantibodies directed against components of the nucleus of the cell. |
| Anti-thyroid peroxidase antibody positivity (HP:0025379) | The presence of autoantibodies (immunoglobulins ) in the serum that react against thyroid peroxidase. | - | Anti-thyroid peroxidase antibody | An autoantibody is a kind of antibody that is directed against a substance made by one's own body (usually, antibodies are directed against external agents such as bacteria). This term refers to an autoantibody directed against the protein thyroid peroxidase. |
| Anti-thyroglobulin antibody positivity (HP:0032069) | The presence of autoantibodies (immunoglobulins | - | Anti-thyroglobulin antibody | An autoantibody is a kind of antibody that is |

| | | | | |
|---|---|---|---|---|
| | ) in the serum that react to thyroglobulin. | | | directed against a substance made by one’s own body (usually, antibodies are directed against external agents such as bacteria). This term refers to an autoantibody directed against the protein thyroglobulin. |
| Lymphadenopathy (HP:0002716) | Enlargment (swelling) of a lymph node. | Swollen lymph nodes; Lymph node hyperplasia | Swollen lymph nodes | Swollen lymph nodes. |
| Lymphopenia (HP:0001888) | A reduced number of lymphocytes in the blood. | Absolute lymphocyte count decrease; Low lymphocyte number; Decreased blood lymphocyte number; Lymphocytopenia | Low lymphocyte count | A decreased number of lymphocytes (a type of white blood cell) in the blood. |

neuropsychiatric-headache

| HPO Term | Definition | Synonyms | Plain-language label | Plain-language definition |
|---|---|---|---|---|
| Headache (HP:0002315) | Cephalgia, or pain sensed in various parts of the head, not confined to the area of distribution of any nerve. | Headaches | Headache | A feeling of pain in the head. |
| Migraine (HP:0002076) | Migraine is a chronic neurological disorder characterized by episodic attacks of headache and associated symptoms. | Migraine headache; Intermittent migraine headaches; Migraine headaches | Migraine | Migraine headache. |

HEENT-ear

| HPO Term | Definition | Synonyms | Plain-language label | Plain-language definition |
|---|---|---|---|---|
| Ear pain (HP:0030766) | Pain in the ear can be a consequence of otologic disease (primary or otogenic otalgia), or can arise from pathologic processes and structures other than the ear (secondary or referred otalgia). | Otalgia; Pain in the ear | Ear pain | Pain in the ear. |
| Hearing impairment (HP:0000365) | A decreased magnitude of the sensory perception of sound. | Hearing loss; Hearing defect; Hypoacusis; Hypacusis; Deafness | Hearing loss | Decreased ability to hear sounds. |
| Hyperacusis (HP:0010780) | Over-sensitivity to certain frequency ranges of sound. | Loudness intolerance; Sensitivity to noise | Loudness intolerance | Oversensitivity to sound. |
| Pulsatile tinnitus (HP:0008629) | Pulsatile tinnitus is generally classified a kind of objective tinnitus, meaning that it is not only audible to the patient but also to the examiner on auscultation of the auditory canal and/or of surrounding structures with use of an auscultation tube or stethoscope. Usually, pulsatile tinnitus is heard as a lower pitched thumping or booming, a | - | Rhythmic ringing in the ears | Rhythmic thumping or whooshing sound heard in the ear that is caused by a patient’s heartbeat or pulse. |

| | | | | |
|---|---|---|---|---|
| | rougher blowing sound which is coincidental with respiration, or as a clicking, higher pitched rhythmic sensation. | | | |
| Tinnitus (HP:0000360) | Tinnitus is an auditory perception that can be described as the experience of sound, in the ear or in the head, in the absence of external acoustic stimulation. | Ringing in ears; Ringing in the ears | Ringing in the ears | Hearing ringing or buzzing in the ear that is not caused by external sound. |
| Vertigo (HP:0002321) | An abnormal sensation of spinning while the body is actually stationary. | Dizziness; Dizzy spell | Dizziness | The feeling of spinning when the body is still. |

neuropsychiatric-smell-taste

| HPO Term | Definition | Synonyms | Plain-language label | Plain-language definition |
|---|---|---|---|---|
| Ageusia (HP:0041051) | A rare condition that is characterized by a complete loss of taste function of the tongue. | Absent sense of taste; Impaired taste sensation; Lost taste | Loss of taste | Complete loss of the sense of taste. |
| Anosmia (HP:0000458) | An inability to perceive odors. This is a general term describing inability to smell arising in any part of the process of smelling from absorption of odorants into the nasal mucous overlying the olfactory epithelium, diffusion to the cilia, binding to olfactory receptor sites, generation of action potentials in olfactory neurons, and perception of a smell. | Loss of smell; Lost smell | Loss of smell | Complete loss of the sense of smell. |
| Hypogeusia (HP:0000224) | A decreased ability to perceive flavor. | Decreased taste sensation; Decreased taste | Decreased sense of taste. | Decreased ability to taste. |
| Hyposmia (HP:0004409) | A decreased sensitivity to odorants (that is, a decreased ability to perceive odors). | Decreased smell sensation; Sense of smell impaired | Decreased sense of smell. | Decreased ability to smell. |
| Parageusia (HP:0031249) | A distortion of the sense of taste, often characterized by the sensation of a metallic taste. | Altered sense of taste; Dysgeusia; Metallic taste in mouth; Metallic taste | Altered sense of taste | Altered sense of taste. For example, an affected individual may have the sensation of |

| | | | | |
|---|---|---|---|---|
| | | | | metallic taste. |
| Phantageusia (HP:0033847) | A form of altered taste sensation in which the affected person perceives a taste, usually an unpleasant one, in the absence of a corresponding stimulus in the environment. | Phantom taste | Phantom taste | Imagining/halluci nating tastes - tasting things when there's nothing in your mouth. |
| Phantosmia (HP:0033693) | Perception of an odor in the absence of any stimuli in the surrounding environment that could emit the odor. | Olfactory hallucination; Phantom odour; Phantom smell; Phantom odor | Phantosmia | Imagining/halluci nating smells that aren't there. |

General-symptom

| HPO Term | Definition | Synonyms | Plain-language label | Plain-language definition |
|---|---|---|---|---|
| Arthritis (HP:0001369) | Inflammation of a joint. | Joint inflammation | Joint inflammation | Inflammation of a joint. |
| Asthenia (HP:0025406) | A state characterized by a feeling of weakness and loss of strength leading to a generalized weakness of the body. | Weakness; Lack of energy and strength | Weakness | Generalized weakness and lack of strength, with a sense of exhaustion which occurs before any effort. |
| Chest tightness (HP:0031352) | An unpleasant sensation of tightness or pressure in the chest. | Tightness in chest; Tightness of chest; Chest distress | Chest tightness | An unpleasant feeling of tightness or pressure in the chest. |
| Chills (HP:0025143) | A sudden sensation of feeling cold. | - | Chills | A sudden sensation of feeling cold. |
| Coldness (HP:0033850) | Relative coldness of a body part to palpitation, often acccompanied by feelings of coldness. | Cold skin temperature; Cool skin temperature; Cool skin; Coolness to palpation | Coldness | Coldness (cool skin) of s body part such as the hands or feet. |
| Difficulty walking (HP:0002355) | Reduced ability to walk (ambulate). | Difficulty in walking; Walking disability | Difficulty walking | Difficulty walking. |
| Diminished health-related quality of life (HP:0033665) | A reduction in an individual's subjective assessment of his or her sense of well-being and ability to perform social roles. | - | Diminished health-related quality of life | A reduced sense of well-being and ability to perform one's usual social roles. |
| Diminished mental health (HP:0033667) | A reduction in the subjective feeling of mental well being. | Reduced mental health; Mental impairment | Diminished mental health | A reduction in the subjective feeling of mental well being. |
| Diminished physical functioning (HP:0033666) | A reduction in the ability to perform activities of daily living as | Diminished physical health; Decline in physical functional health | Diminished physical functioning | A reduction in the ability to perform activities of daily living as |

| | | | | |
|---|---|---|---|---|
| | compared to previous abilities because of functional deficits due to illness. The 36-item Short Form (SF-36) health survey questionnaire is one of many methods used to measure patients' perceptions of diminished physical functioning. | | | compared to previous abilities because of functional deficits due to illness. |
| Exercise intolerance (HP:0003546) | A functional motor deficit where individuals whose responses to the challenges of exercise fail to achieve levels considered normal for their age and gender. | Poor exercise tolerance; Low exercise endurance; Inability to exercise; Decreased ability to exercise | Exercise intolerance | Inability or decreased ability to perform physical exercise at the normally expected level or duration. |
| Fatigue (HP:0012378) | A subjective feeling of tiredness characterized by a lack of energy and motivation. | Tired; Tiredness | Fatigue | Feeling tired, having a lack of energy and no desire do so anything. |
| Frailty (HP:0033675) | A clinically recognizable state of increased vulnerability resulting from a decline in reserve and function across multiple physiologic systems such that the ability to cope with everyday or acute stressors is compromised. | - | Frailty | A state of having delicate health and lack of robustness, characterized by symptoms such as i) low grip strength; (ii) low energy; (iii) slowed waking speed; (iv) low physical activity; and (v) unintentional weight loss. |

| | | | | |
|---|---|---|---|---|
| Impaired ability to bathe oneself (HP:0031059) | This term applies to an individual who requires help to bathe more than one part of the body, get in or out of the tub or shower, or who requires total bathing. | - | Impaired ability to bathe oneself | Affected person cannot bathe without help |
| Impaired ability to dress oneself (HP:0031060) | This applies to an individual who needs help with dressing or needs to be completely dressed. | - | Impaired ability to dress oneself | Affected person cannot dress without help |
| Impairment of activities of daily living (HP:0031058) | Difficulty in performing one or more activities normally performed every day, such as eating, bathing, dressing, grooming, work, homemaking, and leisure. | - | Impairment of activities of daily living | Difficulty in performing one or more activities normally performed every day, such as eating, bathing, dressing, grooming, work, homemaking, and leisure. |
| Malaise (HP:0033834) | A feeling of general discomfort, weakness, or lack of health. | - | Malaise | A feeling of general discomfort, weakness, or lack of health. |
| Night sweats (HP:0030166) | Occurrence of excessive sweating during sleep. | Nocturnal hyperhidrosis | Night sweats | Excessive sweating during sleep. |
| Occupational disability (HP:0033695) | This is a general term that denotes a reduced ability to perform the work that one performed prior to an illness, and may be related to pain, cognitive dysfunction, fatigue or other | - | Occupational disability | Inability or reduced ability to perform the work that one performed prior to an illness. |

| | | | | |
|---|---|---|---|---|
| | physical disabilities. | | | |
| Postexertional malaise (HP:0030973) | A subjective feeling of tiredness characterized by a lack of energy and motivation and that is induced by exertion or exercise. | Postexertional fatigue; Exercise-induced fatigue; Exercise-induced malaise | Postexertional malaise | A feeling of tiredness and fatigue that occurs following physical activity or exercise. |
| Shivering (HP:0025144) | Involuntary contraction or twitching of the muscles. | Shuddering | Shivering | Shaking slightly and uncontrollably, usually as a result of being cold. |
| Stiff neck (HP:0025258) | A sensation of tightness in the neck when attempting to move it, especially after a period of inactivity. Neck stiffness often involves soreness and difficulty moving the neck, especially when trying to turn the head to the side. | Neck stiffness | Stiff neck | A sensation of tightness or stiffness in the neck when attempting to move it. |
| Weight loss (HP:0001824) | Reduction of total body weight. | Loss of weight | Weight loss | Reduction of body weight. |
| Xerostomia (HP:0000217) | Dryness of the mouth due to salivary gland dysfunction. | Dry mouth; Decreased salivary flow; Reduced salivation; Dry mouth syndrome | Dry mouth | Dry mouth. |

cardiovascular-finding

| HPO Term | Definition | Synonyms | Plain-language label | Plain-language definition |
|---|---|---|---|---|
| Bradycardia (HP:0001662) | A slower than normal heart rate (in adults, slower than 60 beats per minute). | Brachycardia; Slow heartbeats | Bradycardia | Slow heart rate. |
| Elevated myocardial native T1 (HP:4000006) | Increased duration of myocardial T1 time without gadolinium contrast. T1 mapping consists of quantifying the T1 relaxation time of a tissue by using analytical expressions of image-based signal intensities. A fundamental principle of MR imaging is that the signal intensity of pixels is based on the relaxation of hydrogen nuclei protons in a static magnetic field. The T1 relaxation times between two tissues vary substantially. Edema, fat infiltration, and fibrosis also cause differences in T1 relaxivity. | Prolonged myocardial native T1 | Elevated myocardial native T1 | This is an abnormal finding of magnetic resonance imaging (MRI) indicating an abnormality in the heart muscle. |
| Elevated myocardial native T2 (HP:4000003) | Increased duration of myocardial T2 time without gadolinium contrast. Elevated T2, which can | - | Elevated myocardial native T2 | This is an abnormal finding of magnetic resonance imaging (MRI) indicating an abnormality in the |

| | | | | |
|---|---|---|---|---|
| | detect myocardial edema. | | | heart muscle. |
| Hypertension (HP:0000822) | The presence of chronic increased pressure in the systemic arterial system. | Arterial hypertension; Systemic hypertension; High blood pressure | High blood pressure | High blood pressure. |
| Hypotension (HP:0002615) | Low Blood Pressure, vascular hypotension. | Arterial hypotension; Low blood pressure | Low blood pressure | Low blood pressure. |
| Increased circulating troponin T concentration (HP:0410174) | An increased concentration of tropnin T in the blood, which is a cardiac regulatory protein that controls the calcium mediated interaction between actin and myosin. Raised cardiac troponin concentrations are now accepted as the standard biochemical marker for the diagnosis of myocardial infarction. | Increased troponin T level in blood | High troponin T level | An increased level of troponin T in the blood. Elevated troponin T levels are a marker of damage to the heart muscle. |
| Increased circulating troponin I concentration (HP:0410173) | An increased concentration of tropnin I in the blood, which is a cardiac regulatory protein that controls the calcium mediated interaction between actin and myosin. Raised cardiac troponin concentrations are now accepted | Increased troponin I level in blood | High troponin I level | An increased level of troponin I in the blood. Elevated troponin I levels are a marker of damage to the heart muscle. |

| | | | | |
|---|---|---|---|---|
| | as the standard biochemical marker for the diagnosis of myocardial infarction. | | | |
| Increased heart rate variability (HP:0031862) | Increased variation of beat-to-beat intervals of the heart that occurs in conjunction with the respiratory cycle. | - | Increased heart rate variability | Reduced consistency of time lapse between heartbeats. |
| Increased left ventricular end-diastolic volume (HP:0033755) | Abnormally high volume of blood in the left ventricle at the end of diastole (just before systole). | - | Increased left ventricular end-diastolic volume | Abnormally high volume of blood in the left ventricle when the heart muscle is relaxed. |
| Myocardial late gadolinium enhancement (HP:4000004) | Areas of high signal intensity in magnetic resonance imaging of the heart appearing 10 to 15 minutes after injection of the intercellular contrast agent gadolinium. | Delayed myocardial gadolinium enhancement | Myocardial late gadolinium enhancement | This is an abnormal finding of magnetic resonance imaging (MRI) indicating an abnormality in the heart muscle. |
| Myocarditis (HP:0012819) | Inflammation of the myocardium. | Inflammation of heart muscle | Myocarditis | Inflammation of the heart muscle. |
| Pericardial effusion (HP:0001698) | Accumulation of fluid within the pericardium. | Fluid around heart; Pericardial effusions | Pericardial effusion | Buildup of extra fluid underneath the membrane that covers the heart (pericardium). |
| Pericardial late gadolinium enhancement (HP:4000005) | Areas of high signal intensity in magnetic resonance imaging of the pericardium appearing around | - | Pericardial late gadolinium enhancement | This is an abnormal finding of magnetic resonance imaging (MRI) indicating an abnormality in the |

| | | | | |
|---|---|---|---|---|
| | 10 minutes after injection of the intercellular contrast agent gadolinium. | | | membrane that covers the heart (pericardium). |
| Reduced ejection fraction (HP:0012664) | A diminution of the volumetric fraction of blood pumped out of the ventricle with each cardiac cycle. | - | Reduced ejection fraction | A reduction in the amount of blood the left ventricle of the heart pumps out with each contraction. |
| Tachycardia (HP:0001649) | A rapid heartrate that exceeds the range of the normal resting heartrate for age. | Elevated heart rate; Rapid heart beat; Increased heart rate; Fast heart rate; Racing heart; Heart racing | Tachycardia | A rapid heart rate |
| Venous thrombosis (HP:0004936) | Formation of a blood clot (thrombus) inside a vein, causing the obstruction of blood flow. | Blood clot in vein | Venous thrombosis | Blood clot in vein. |

neuropsychiatric-emotion-mood

| HPO Term | Definition | Synonyms | Plain-language label | Plain-language definition |
|---|---|---|---|---|
| Aggressive behavior (HP:0000718) | Aggressive behavior can denote verbal aggression, physical aggression against objects, physical aggression against people, and may also include aggression towards oneself. | Aggressiveness; physical aggression; Aggressive behaviour; Aggression | Aggressive behavior | Verbal or physical aggression. |
| Depression (HP:0000716) | Frequent feelings of being down, miserable, and/or hopeless; difficulty recovering from such moods; pessimism about the future; pervasive shame; feeling of inferior self-worth; thoughts of suicide and suicidal behavior. | Depressive disorder; Depressivity | Depression | Frequent feelings of being down, miserable, and/or hopeless; difficulty recovering from such moods; pessimism about the future; pervasive shame; feeling of inferior self-worth; thoughts of suicide and suicidal behavior. |
| Dysphoria (HP:0033838) | A state of feeling very unhappy, uneasy, or dissatisfied. | - | Dysphoria | A state of feeling very unhappy, uneasy, or dissatisfied. |
| Emotional lability (HP:0000712) | Unstable emotional experiences and frequent mood changes; emotions that are easily aroused, intense, and/or out of proportion to events and circumstances. | Mood lability; Mood alterations; Mood changes; Emotional instability | Mood changes | Rapid, intense changes in mood. |
| Euphoria (HP:0031844) | A sense of intense joy or happiness | - | Euphoria | A sense of intense joy or happiness |

| | | | | |
|---|---|---|---|---|
| | that is beyond what would be expected under the given circumstances. | | | that is beyond what would be expected under the given circumstances. |
| Mania (HP:0100754) | A state of abnormally elevated or irritable mood, arousal, and or energy levels. | Manic | Mania | A condition of intense moods, extreme excitement and euphoria. |
| Sense of impending doom (HP:0033845) | A feeling that something life-threatening or tragic is about to occur. | Sense of doom | Sense of doom | A feeling that something life-threatening or tragic is about to occur. |
| Suicidal ideation (HP:0031589) | Frequent thinking about or preoccupation with killing oneself. | Suicidality | Suicidal ideation | Thinking about or planning suicide. |
| Tearfulness (HP:0033705) | A feeling of sadness characterized by crying episodes that can come on suddenly and are not under usual social control. | - | Tearfulness | A feeling of sadness with episodes of uncontrolable crying. |

neuropsychiatric-memory

| HPO Term | Definition | Synonyms | Plain-language label | Plain-language definition |
|---|---|---|---|---|
| Anterograde memory impairment (HP:0033689) | The impaired ability to establish new long-term memories. | Anterograde amnesia | Anterograde memory impairment | Loss of ability to create new memories. |
| Long term memory impairment (HP:0033688) | A deficit in the ability to retrieve information from long-term memory, which can be defined as a seemingly unlimited capacity to store memories can last years and relate to the performance of actions or skills (i.e., procedural memories, knowing how) and memories of facts, rules, concepts, and events (i.e., declarative memories, knowing that). | Long term memory loss | Long term memory loss | Long term memory loss |
| Memory impairment (HP:0002354) | An impairment of memory as manifested by a reduced ability to remember things such as dates and names, and increased forgetfulness. | Memory loss; Memory problems; Poor memory; Forgetfulness | Memory loss | Reduced ability to remember things. |
| Procedural memory loss (HP:0033691) | A reduction in the ability to retrieve information about how to perform activities, such as how to ride a bike or drive a car, how to perform | Impaired procedural memory; Procedural memory deficit | Procedural memory loss | Forgetting how to do routine tasks. |

| | | | | |
|---|---|---|---|---|
| | activities of daily living, or how to play a musical instrument. | | | |
| Short term memory impairment (HP:0033687) | A deficit in the retention of pieces of information (memory chunks) for a relatively short time (usually up to 30 seconds). | Short term memory loss | Short term memory loss | Difficulty remembering pieces of information that one has recently been exposed to. For instance, affected individuals may ask the same question repeatedly or forget where they just put the keys. |

General-pain

| HPO Term | Definition | Synonyms | Plain-language label | Plain-language definition |
|---|---|---|---|---|
| Arthralgia (HP:0002829) | Joint pain. | Joint pains; Arthralgias; Arthritic pain; Joint pain | Joint Pain | Joint Pain |
| Body ache (HP:0033047) | Body ache is a complaint that is often used to denote vague symptoms of mild fatigue, lethargy, or dull aches. We will define it here to mean a dull and poorly localizable pain that is described by the affected individual to affect multiple joints or body parts or even the entire body. | - | Body ache | A sharp or dull ache and pain in most or all of the body. |
| Bone pain (HP:0002653) | An unpleasant sensation characterized by physical discomfort (such as pricking, throbbing, or aching) localized to bone. | - | Bone pain | Pain in a bone. |
| Chest pain (HP:0100749) | An unpleasant sensation characterized by physical discomfort (such as pricking, throbbing, or aching) localized to the chest. | Chest discomfort; Thoracic pain | Chest pain | Aching, burning or sharp pain in the chest. |
| Intrascapular pain (HP:0033746) | An unpleasant sensation characterized by physical | Pain between shoulder blades | Pain between shoulder blades | Pain between shoulder blades. |

| | | | | |
|---|---|---|---|---|
| | discomfort (such as pricking, throbbing, or aching) localized to the area between the shoulder blades. | | | |
| Limb pain (HP:0009763) | Chronic pain in the limbs with no clear focal etiology. | Pain in extremities | Limb pain | Pain in a leg or arm. |
| Myalgia (HP:0003326) | Pain in muscle. | Myalgias; Muscle ache; Muscle pain | Muscle pain | Muscle pain. |
| Neuralgia (HP:0033345) | Pain (An unpleasant sensory and emotional experience) along the course of a nerve. | - | Neuralgia | Intense pain along the course of a nerve, Typically, the pain of neuralgia comes and goes. |
| Pain (HP:0012531) | An unpleasant sensory and emotional experience associated with actual or potential tissue damage, or described in terms of such damage. | - | Pain | Physical discomfort. |

gi-findings

| HPO Term | Definition | Synonyms | Plain-language label | Plain-language definition |
|---|---|---|---|---|
| Gastroesophageal reflux (HP:0002020) | A condition in which the stomach contents leak backwards from the stomach into the esophagus through the lower esophageal sphincter. | Gastro-oesophageal reflux; Gastroesophageal reflux disease; Acid reflux disease; GERD; Acid reflux; Heartburn | Gastroesophageal reflux disease | Backwash of the acidic fluid of the stomach into the esophagus (the tube that connects the stomach and the mouth). |
| Gastroparesis (HP:0002578) | Decreased strength of the muscle layer of stomach, which leads to a decreased ability to empty the contents of the stomach despite the absence of obstruction. | Delayed gastric emptying | Weakened stomach muscle | Decreased strength of the muscle layer of stomach. |
| Gastric ulcer (HP:0002592) | An ulcer, that is, an erosion of an area of the gastric mucous membrane. | Stomach ulcer | Stomach ulcer | A sore on the lining of the stomach |
| Hepatic steatosis (HP:0001397) | Steatosis is a term used to denote lipid accumulation within hepatocytes. | Steatosis; Liver steatosis; Fatty infiltration of liver; Fatty liver | Fatty liver | Accumulation of fat in the liver. |
| Hepatomegaly (HP:0002240) | Abnormally increased size of the liver. | Enlarged liver | Enlarged liver | Enlarged liver. |
| Hepatitis (HP:0012115) | Inflammation of the liver. | Liver inflammation | Liver inflammation | Liver inflammation |
| Malnutrition (HP:0004395) | A deficiency in the intake of energy and nutrients. | - | Malnutrition | A deficiency in the intake of energy and nutrients. |
| Pancreatic steatosis (HP:0033757) | Fat infiltration in the pancreas. | Pancreatic lipomatosis; Non-alcoholic fatty pancreatic disease; | Fatty pancreas | Accumulation of fat in the pancreas. |

| | | | | |
|---|---|---|---|---|
| | | Pancreatic fatty infiltration; Pancreatic fatty replacement; Fatty pancreas; Pancreatic lipomatous pseudohypertrophy | | |
| Splenomegaly (HP:0001744) | Abnormal increased size of the spleen. | Increased spleen size | Enlarged spleen | Enlarged spleen. |

Lab

| HPO Term | Definition | Synonyms | Plain-language label | Plain-language definition |
|---|---|---|---|---|
| Decreased circulating calcifediol concentration (HP:0012053) | A reduced concentration of calcifediol in the blood. Calcifediol is also known as calcidiol, 25-hydroxycholecalciferol and 25-Hydroxyvitamin D3. | Low serum 25-hydroxycholecalciferol; Decreased 25-hydroxyvitamin D3; Low serum calcidiol; Low serum calcifediol | Low 25-hydroxyvitamin D3 level | A reduced concentration of calcifediol in the blood. Calcifediol is a form of vitamin D. |
| Elevated circulating alkaline phosphatase concentration (HP:0003155) | Abnormally increased serum levels of alkaline phosphatase activity. | Increased serum alkaline phosphatase; Increased alkaline phosphatase; Elevated alkaline phosphatase; High serum alkaline phosphatase; Hyperphosphatasia; Elevated ALP; Greatly elevated alkaline phosphatase; Hyperphosphatasemia | High alkaline phosphatase level | An increased level of alkaline phosphatase in the blood. High alkaline phosphatase levels can be found due to many different causes including liver damage or bone disorders. |
| Elevated circulating alanine aminotransferase concentration (HP:0031964) | An abnormally high concentration in the circulation of alanine aminotransferase (ALT). | Elevated serum ALT; Alanine aminotransferase increased; Elevated serum alanine aminotransferase; Elevated serum glutamic-pyruvic transaminase | High ALT level | An increased level of alanine aminotransferase (ALT) in the blood. High ALT levels may indicate damage or disease in the liver. |
| Elevated circulating aspartate aminotransferase concentration (HP:0031956) | An abnormally high concentration in the circulation of aspartate aminotransferase (AST). | Elevated serum glutamic oxaloacetic transaminase; Elevated serum AST; Aspartate aminotransferase increased; | High AST level | An increased level of aspartate aminotransferase (AST) in the blood. High AST levels may indicate damage or disease in the |

| | | | | |
|---|---|---|---|---|
| | | Elevated serum aspartate aminotransferase | | liver. |
| Elevated circulating creatinine concentration (HP:0003259) | An increased amount of creatinine in the blood. | High blood creatinine level; Elevated serum creatinine; Increased creatinine; Increased serum creatinine; Elevated creatinine | High creatinine level | An increased level of creatinine in the blood. High creatinine levels can indicate impaired kidney function or kidney disease. |
| Elevated circulating creatine kinase concentration (HP:0003236) | An elevation of the level of the enzyme creatine kinase (also known as creatine phosphokinase, CPK; EC 2.7.3.2) in the blood. CPK levels can be elevated in a number of clinical disorders such as myocardial infarction, rhabdomyolysis, and muscular dystrophy. | Elevated serum CPK; Increased CPK; Increased serum creatine phosphokinase; Elevated serum creatine phosphokinase; Increased creatine phosphokinase; High serum creatine kinase; Increased serum CK; Increased creatine kinase; Elevated creatine kinase; Increased serum creatine kinase; Elevated circulating creatine phosphokinase; Elevated serum creatine kinase; Elevated blood creatine phosphokinase | High CK level | An increased level of creatine kinase (CK) in the blood. Increased CK levels may indicate damage or disease of the skeletal muscles, heart, or brain. |
| Elevated circulating C-reactive protein concentration (HP:0011227) | An abnormal elevation of the C-reactive protein level in the blood circulation. | Elevated CRP; Elevated C-reactive protein level | High CRP level | An increased level of C-reactive protein (CRP) in the blood. CRP is a marker for inflammation. |
| Elevated | An increased | Elevated fibrin | High D-dimer level | An increased level |

| | | | | |
|---|---|---|---|---|
| circulating D-dimer concentration (HP:0033106) | concentration of D-dimers, a marker of fibrin degradation, in the blood circulation. | degradation fragment concentration; Elevated D-dimers | | of D-dimers in the blood. High D-dimers may be observed in patients with abnormal clotting of blood within blood vessels. |
| Elevated circulating soluble CD25 concentration (HP:0033833) | Increased concentration of the interleukin-2 receptor alpha-chain (CD25) in the blood circulation. CD25 is shed upon immune activation. Increased levels of soluble CD25, therefore, are an indication of an on-going immune response. | Elevated circulating interleukin-2 receptor alpha-chain | High sCD25 level | In increased level of sCD25 in the blood, which can be a sign of ongoing inflammation |
| Elevated circulating thyroid-stimulating hormone concentration (HP:0002925) | Increased concentration of thyroid-stimulating hormone (TSH) in the blood circulation. | Elevated thyroid stimulating hormone levels; Increased thyroid-stimulating hormone level; Increased thyroid-stimulating hormone; Increased serum thyroid-stimulating hormone; Increased thyrotropin level; TSH excess; Elevated thyroid stimulating hormone; Thyroid-stimulating hormone excess; High TSH | High TSH level | An increased level of thyroid-stimulating hormone (TSH) in the blood. High TSH levels can be observed in a number of diseases affecting the thyroid. |

| | | | | |
|---|---|---|---|---|
| Elevated erythrocyte sedimentation rate (HP:0003565) | An increased erythrocyte sedimentation rate (ESR). The ESR is a test that measures the distance that erythrocytes have fallen after one hour in a vertical column of anticoagulated blood under the influence of gravity. The ESR is a nonspecific finding. An elevation may indicate inflammation or may be caused by any condition that elevates fibrinogen. | Raised erythrocyte sedimentation rate; Increased erythrocyte sedimentation rate; Elevated ESR; Elevated sedimentation rate; High erythrocyte sedimentation rate; High ESR | Elevated ESR | An increased erythrocyte sedimentation rate (ESR). High ESR values may indicate inflammation. |
| Elevated gamma-glutamyltransferase level (HP:0030948) | Increased level of the enzyme gamma-glutamyltransferase (GGT). GGT is mainly present in kidney, liver, and pancreatic cells, but small amounts are present in other tissues. | Elevated serum GGT | High GGT level | An increased level of gamma-glutamyltransferase (GGT) in the blood. High GGT levels can indicate liver or bile duct disease. |
| Hypocalcemia (HP:0002901) | An abnormally decreased calcium concentration in the blood. | Low blood calcium levels; Hypocalcaemia | Low calcium level | A low level of calcium in the blood. |
| Hypofibrinogenemia (HP:0011900) | Decreased concentration of fibrinogen in the blood. | Low fibrinogen level; Low fibrinogen activity | Low fibrinogen level | A low level of fibronogen in the blood. |
| Hyperglycemia (HP:0003074) | An increased concentration of glucose in the | High blood glucose; High blood sugar | High blood sugar | Increased blood sugar level. |

| | | | | |
|---|---|---|---|---|
| | blood. | | | |
| Hypoglycemia (HP:0001943) | A decreased concentration of glucose in the blood. | Hypoglycaemia; Low blood sugar | Low blood sugar | Decreased blood sugar level. |
| Hypophosphatemia (HP:0002148) | An abnormally decreased phosphate concentration in the blood. | Low blood phosphate level; Hypophosphataemia | Low phosphate level | A low level of phosphate in the blood. |
| Increased circulating ferritin concentration (HP:0003281) | Increased concentration of ferritin in the blood circulation. | High ferritin level; Increased serum ferritin level; Elevated serum ferritin; Increased ferritin; Hyperferritinemia; Increased plasma ferritin; Hyperferritinaemia | High ferritin level | An increased level of ferritin in the blood. Ferritin is a ubiquitous intracellular protein that stores iron. Ferritin levels measured in serum usually have a direct correlation with the total amount of iron stored in the body, but ferritin levels may also be increased with inflammation. |
| Increased circulating interleukin 6 (HP:0030783) | An increased concentration of interleukin-6 in the circulation. | Increased serum IL-6; Increased serum interleukin-6 | High IL6 level | An elevated level of interleukin-6 (IL6) in the bloodstream. High IL6 levels may indicate ongoing inflammation. |
| Increased circulating lactate dehydrogenase concentration (HP:0025435) | An elevated level of the enzyme lactate dehydrogenase in the blood circulation. | Increased lactate dehydrogenase level | High LDH leve | An increased level of lactate dehydrogenase (LDH) in the blood. LDH is a nonspecific marker of tissue damage and can rise with problems in the blood, heart, |

| | | | | |
|---|---|---|---|---|
| | | | | kidneys, brain, or lungs. |
| Increased circulating NT-proBNP concentration (HP:0031185) | An elevated level of circulating N-terminal part of the prohormone of B-type natriuretic peptide (BNP). | Increased NT-proBNP level | High NT-proBNP level | An increased level of NT-proBNP in the blood. High NT-proBNP levels may indicate disease or damage to the heart. |
| Increased circulating procalcitonin concentration (HP:0032308) | An elevated concentration of procalcitonin in the blood circulation. | Increased circulating procalcitonin level | High procalcitonin level | An increased level of procalcitonin in the blood. High procalcitonin levels may be found in conditions including bacterial infections and sepsis. |
| Thrombocytopenia (HP:0001873) | A reduction in the number of circulating thrombocytes. | Low platelet count | Low platelet count | A decreased number of platelets in the blood. |

cardiovascular-symptom

| HPO Term | Definition | Synonyms | Plain-language label | Plain-language definition |
|---|---|---|---|---|
| Angina pectoris (HP:0001681) | Paroxysmal chest pain that occurs with exertion or stress and is related to myocardial ischemia. | - | Angina | Chest pain or discomfort caused by lack of adequate blood supply to the heart |
| Palpitations (HP:0001962) | A sensation that the heart is pounding or racing, which is a non-specific sign but may be a manifestation of arrhythmia. | Missed heart beat; Skipped heart beat; Heart palpitations | Palpitations | The feeling of a racing, pounding, and/or irregular heartbeat. |
| Stroke (HP:0001297) | Sudden impairment of blood flow to a part of the brain due to occlusion or rupture of an artery to the brain. | Cerebrovascular accidents; Cerebrovascular accident; Cerebral vascular events | Stroke | The blockage of a blood vessel by a clot or burst which prevents oxygen from reaching the brain. |
| Syncope (HP:0001279) | Syncope refers to a generalized weakness of muscles with loss of postural tone, inability to stand upright, and loss of consciousness. Once the patient is in a horizontal position, blood flow to the brain is no longer hindered by gravitation and consciousness is regained. Unconsciousness usually lasts for seconds to minutes. | Fainting spell | Fainting | Fainting. |

| | |
|---|---|
| | Headache and drowsiness (which usually follow seizures) do not follow a syncopal attack. Syncope results from a sudden impairment of brain metabolism usually due to a reduction in cerebral blood flow. |

neuropsychiatric-behavioral

| HPO Term | Definition | Synonyms | Plain-language label | Plain-language definition |
|---|---|---|---|---|
| Anxiety (HP:0000739) | Intense feelings of nervousness, tenseness, or panic, often in reaction to interpersonal stresses; worry about the negative effects of past unpleasant experiences and future negative possibilities; feeling fearful, apprehensive, or threatened by uncertainty; fears of falling apart or losing control. | Anxiety disease; Anxiousness; Excessive, persistent worry and fear | Anxiety | Intense feelings of nervousness, tenseness, or panic, often in reaction to interpersonal stresses; worry about the negative effects of past unpleasant experiences and future negative possibilities; feeling fearful, apprehensive, or threatened by uncertainty; fears of falling apart or losing control. |
| Apathy (HP:0000741) | | Lack of feeling, emotion, interest | Apathy | A lack of any feelings, emotions, or interest. |
| Attention deficit hyperactivity disorder (HP:0007018) | Attention deficit hyperactivity disorder (ADHD) manifests at age 2-3 years or by first grade at the latest. The main symptoms are distractibility, impulsivity, hyperactivity, and often trouble organizing tasks and projects, difficulty going to sleep, and social problems from being aggressive, loud, or impatient. | Attention deficit disorder; Childhood attention deficit/hyperactivity disorder; Attention deficit; Attention deficits; Attention deficit-hyperactivity disorder | ADHD | A state of distractibility, impulsivity, hyperactivity, and often trouble organizing tasks and projects, difficulty going to sleep, and social problems from being aggressive, loud, or impatient. |
| Auditory | The false | Hallucinations of | Auditory | Imagining/halluci |

| | | | | |
|---|---|---|---|---|
| hallucinations (HP:0008765) | perception of sound. | sound; Hearing sounds | hallucinations | nating sounds that are not there. |
| Brain fog (HP:0033630) | Brain fog is a type of transient cognitive dysfunction that comprises a constellation of symptoms that impair intellectual functioning to a level that interferes with daily activities, commonly including forgetfulness, mental slowness, difficulty thinking or focusing, a perceived slowing of mental processing speed, inability to find the right words, a sensation that the mind went blank or is "cloudy". Brain fog tends to recur and may be triggered by factors such as physical fatigue, lack of sleep, and prolonged standing or may appear to occur spontaneously. | Mental clouding; Mental fatigue; Mental fog | Brain fog | A temporary feeling of mental fatigue and difficulty thinking. |
| Delusions (HP:0000746) | A false belief that is held despite evidence to the contrary. | - | Delusions | Beliefs that are wrong and is held despite evidence to the contrary. |
| Hallucinations (HP:0000738) | Perceptions in a conscious and awake state in the absence of external stimuli | Hallucination; Sensory hallucination | Hallucinations | Imagining/halluci nating things that are not there. |

| | | | | |
|---|---|---|---|---|
| | which have qualities of real perception, in that they are vivid, substantial, and located in external objective space. | | | |
| Impaired executive functioning (HP:0033051) | A disturbance of executive functioning, which is broadly defined as the set of abilities that allow for the planning, executing, monitoring, and self-correcting of goal-directed behavior while inhibiting task-irrelevant behavior. At least some degree of executive skill is needed to complete most cognitive tasks, and deficits in executive abilities are central to many clinical conditions, including fronto-temporal dementia. | - | Impaired executive functioning | Difficulty with planning, organizing, figuring out the sequence of actions, and abstracting tasks. |
| Impulsivity (HP:0100710) | Acting on the spur of the moment in response to immediate stimuli; acting on a momentary basis without a plan or consideration of outcomes; difficulty establishing or | Impulsive | Impulsivity | A tendency to act on a momentary basis without a plan or consideration of outcomes. |

| | | | | |
|---|---|---|---|---|
| | following plans; a sense of urgency and self-harming behavior under emotional distress. | | | |
| Irritability (HP:0000737) | A proneness to anger, i.e., a condition of being easily bothered or annoyed. | Irritable | Irritability | Being easily bothered or annoyed. |
| Panic attack (HP:0025269) | A sudden episode of intense fear in a situation in which there is no danger or apparent cause. | - | Panic attack | A sudden episode of intense fear in a situation in which there is no danger or apparent cause. |
| Phonophobia (HP:0002183) | An abnormally heightened sensitivity to loud sounds. | Fear of loud sounds | Phonophobia | An abnormally heightened sensitivity to loud sounds. |
| Polydipsia (HP:0001959) | Excessive thirst manifested by excessive fluid intake. | Extreme thirst | Polydipsia | A feeling of extreme thirstiness and excessive fluid intake. |
| Posttraumatic stress symptom (HP:0033676) | A behavioral or psychological symptom that typically occurs following exposure to one or more traumatic events. Posttraumatic stress disorder (PTSD) symptoms include intrusive recollections (re-experiencing the trauma in flashbacks, memories or nightmares); avoidant and numbing symptoms | PTSD | Posttraumatic stress symptom | Physical and emotional reactions to a past traumatic event including problems such as trouble concentrating, irritability, angry outbursts, or aggressive behavior. |

| | | | | |
|---|---|---|---|---|
| | (including diminished emotions and avoidance of situations that are reminders of the traumatic event); and hyperarousal (including increased irritability, exaggerated startle reactions or difficulty sleeping or concentrating). | | | |
| Short attention span (HP:0000736) | Reduced attention span characterized by distractibility and impulsivity but not necessarily satisfying the diagnostic criteria for attention deficit hyperactivity disorder. | Poor attention span; Problem paying attention; Easily distracted | Short attention span | Reduced attention span |
| Tactile hallucination (HP:0033694) | The false perception of tactile sensory input that creates a hallucinatory sensation of physical contact with an imaginary object. | Tactile hallucinations | Tactile hallucination | Imagining/hallucinating feeling things that are not there. |
| Visual hallucinations (HP:0002367) | Visual perceptions that are not elicited by a corresponding stimulus from the outside world. | - | Visual hallucinations | Imagining/hallucinating seeing things that are not there. |

pulmonary-symptom

| HPO Term | Definition | Synonyms | Plain-language label | Plain-language definition |
|---|---|---|---|---|
| Cough (HP:0012735) | A sudden, audible expulsion of air from the lungs through a partially closed glottis, preceded by inhalation. | Coughing | Cough | Cough. |
| Dyspnea (HP:0002094) | Difficult or labored breathing. Dyspnea is a subjective feeling only the patient can rate, e.g., on a Borg scale. | Shortness of breath; Abnormal breathing; Panting; Difficult to breathe; Dyspnoea; Difficulty breathing; Breathing difficulty; Trouble breathing | Shortness of breath | An uncomfortable feeling of shortness of breath that has been described as air hunger. |
| Exertional dyspnea (HP:0002875) | Perceived difficulty to breathe that occurs with exercise or exertion and improves with rest. | Exertional dyspnoea; Exertional breathlessness; Shortness of breathing upon physical activity | Shortness of breathing upon physical activity | An uncomfortable feeling of shortness of breath that occurs following physical activity. |
| Hemoptysis (HP:0002105) | Coughing up (expectoration) of blood or blood-streaked sputum from the larynx, trachea, bronchi, or lungs. | Coughing up blood; Haemoptysis; Coughing up blood or blood-stained mucus | Hemoptysis | Coughing up blood. |
| Increased sputum production (HP:0033709) | An increase in the amount of airway mucus. This feature may be characterized by frequent or excessive throat clearing (exhalation through tightly constricted | Increased phlegm | Increased phlegm | An increase in the amount of airway mucus (sputum (phlegm) with the need to clear one’s throat more often than normal. |

| | | | | |
|---|---|---|---|---|
| | laryngopharyngeal tissues accompanied by vibration of the palatoglossal arch and the vocal folds serving to clear mucus from the airway). | | | |
| Nonproductive cough (HP:0031246) | A cough that does not produce phlegm or mucus. | Dry cough; Dry coughing | Dry cough | A cough that does not produce phlegm or mucus. |
| Pleuritic chest pain (HP:0033771) | Pleuritic chest pain is characterized by sudden and intense sharp, stabbing, or burning pain in the chest when inhaling and exhaling. | - | Pleuritic chest pain | Pain related to inflammation of the lining of the lungs (pleura), usually an intense sharp, stabbing, or burning pain in the chest when inhaling and exhaling. |
| Productive cough (HP:0031245) | A cough that produces phlegm or mucus. | Cough with mucus production; Wet cough | Wet cough | A cough that produces phlegm or mucus. |
| Rest dyspnea (HP:0033710) | A perception of shortness of breath that occurs independently of exertion. | Shortness of breath at rest; Breathlessness at rest; Dyspnea at rest; Dyspnoea at rest | Shortness of breath at rest | An uncomfortable feeling of shortness of breath that occurs while at rest. |
| Rhinorrhea (HP:0031417) | Increased discharge of mucus from the nose. | Runny Nose; Nasal Discharge | Runny nose | Runny nose. |
| Rhonchi (HP:0030831) | Abnormal breath sounds characterized by low-pitched, snoring or rattle-like sounds. | - | Rattle-like breath sounds | Abnormal breath sounds comparable to wheezing but with a low-pitched, rattle-like sound. |
| Sneeze (HP:0025095) | A sudden violent, spasmodic, audible expiration of breath through the nose and mouth. | - | Sneeze | Sneezing. |

| | | | | |
|---|---|---|---|---|
| Tachypnea (HP:0002789) | Very rapid breathing. | Polypnea; Increased respiratory rate or depth of breathing | Rapid breathing | Very rapid breathing. |
| Wheezing (HP:0030828) | A high-pitched whistling sound associated with labored breathing. | - | Wheezing | A high-pitched whistling sound associated with labored breathing. |

neuropsychiatric-finding

| HPO Term | Definition | Synonyms | Plain-language label | Plain-language definition |
| --- | --- | --- | --- | --- |
| Abnormal exteroceptive sensation (HP:0033747) | A type of somatic sensory dysfunction characterized by abnormality of superficial sensation that is mediated by receptors in skin and mucous membranes. | - | Sensory impairment. | A general term for sensory impairment such as being inable to perceive touch, pain, or vibration. |
| Abnormal reflex (HP:0031826) | Any anomaly of a reflex, i.e., of an automatic response mediated by the nervous system (a reflex does not need the intervention of conscious thought to occur). | - | Abnormal reflex | Abnormality of a reflex such as the knee jerk (a sudden involuntary reflex kick caused by tapping the knee with a reflex hammer). |
| Abnormality of movement (HP:0100022) | An abnormality of movement with a neurological basis characterized by changes in coordination and speed of voluntary movements. | Unusual movement; Movement disorder | Abnormal movements | Abnormal movements |
| Ataxia (HP:0001251) | Cerebellar ataxia refers to ataxia due to dysfunction of the cerebellum. This causes a variety of elementary neurological deficits including asynergy (lack of coordination between muscles, limbs and joints), | Cerebellar ataxia | Ataxia | A lack of coordination of voluntary movements that can cause slurred speech, stumbling, falling, and lack of coordination. |

| | | | | |
|---|---|---|---|---|
| | dysmetria (lack of ability to judge distances that can lead to under- or overshoot in grasping movements), and dysdiadochokinesia (inability to perform rapid movements requiring antagonizing muscle groups to be switched on and off repeatedly). | | | |
| Babinski sign (HP:0003487) | Upturning of the big toe (and sometimes fanning of the other toes) in response to stimulation of the sole of the foot. If the Babinski sign is present it can indicate damage to the corticospinal tract. | Extensor plantar responses; Extensor plantar reflexes; Extensor plantar response; Positive Babinski sign | Babinski sign | An abnormal neurological reflex whereby the big toe turns upward if the sole of the foot is stroked. |
| Dysarthria (HP:0001260) | Dysarthric speech is a general description referring to a neurological speech disorder characterized by poor articulation. Depending on the involved neurological structures, dysarthria may be further classified as spastic, flaccid, ataxic, hyperkinetic and | Difficulty articulating speech; Dysarthric speech | Dysarthria | A speech disorder characterized by slurred or slow speech that is difficult to understand. |

| | | | | |
|---|---|---|---|---|
| | hypokinetic, or mixed. | | | |
| Dysmetria (HP:0001310) | A type of ataxia characterized by the inability to carry out movements with the correct range and motion across the plane of more than one joint related to incorrect estimation of the distances required for targeted movements. | Lack of coordination of movement; Abnormal finger chase test; Abnormal finger-nose-finger test | Dysmetria | Inability to perform a movement towards an object at a precise distance. |
| Dysphagia (HP:0002015) | Difficulty in swallowing. | Deglutition disorder; Swallowing difficulties; Poor swallowing; Difficulty swallowing; Swallowing difficulty | Dysphagia | Difficulty swallowing |
| Dystonia (HP:0001332) | An abnormally increased muscular tone that causes fixed abnormal postures. There is a slow, intermittent twisting motion that leads to exaggerated turning and posture of the extremities and trunk. | Dystonic disease; Dystonic movements | Dystonia | Abnormally increased muscular tension that causes fixed abnormal postures. |
| Facial paralysis (HP:0007209) | Complete loss of ability to move facial muscles innervated by the facial nerve (i.e., the seventh | Facial paresis | Weakness of face muscles | An inability to move the muscles on one or both sides of the face. |

| | | | | |
|---|---|---|---|---|
| | cranial nerve). | | | |
| Frontal release signs (HP:0000743) | Primitive reflexes traditionally held to be a sign of disorders that affect the frontal lobes. | Frontal release reflexes | Frontal release signs | Signs of neurological damage that include uncontrollable grasping, sucking, pursing of lips. |
| Gait disturbance (HP:0001288) | The term gait disturbance can refer to any disruption of the ability to walk. In general, this can refer to neurological diseases but also fractures or other sources of pain that is triggered upon walking. However, in the current context gait disturbance refers to difficulty walking on the basis of a neurological or muscular disease. | Abnormal gait; Gait abnormalities; Impaired gait; Gait disturbances; Gait difficulties; Abnormal walk | Gait disturbance | Difficulty walking |
| Hand muscle weakness (HP:0030237) | Reduced strength of the musculature of the hand. | - | Hand weakness | Reduced strength of the hand muscles. |
| Hyperesthesia (HP:0100963) | Increased sensitivity to stimulation, excluding the special senses, which may refer to various modes of cutaneous sensibility including touch and thermal sensation without pain, as well as to pain. | Hyperaesthesia | Hyperesthesia | Increased sensitivity to touch |

| | | | | |
|---|---|---|---|---|
| Hyperkinetic movements (HP:0002487) | Motor hyperactivity with excessive movement of muscles of the body as a whole. | Muscle spasms; Hyperkinesia; Hyperkinesis | Hyperkinetic movements | Excessive movement of muscles. |
| Hypoesthesia (HP:0033748) | Decreased ability to perceive touch. | Numbness; Hypoaesthesia | Numbness | Decreased ability to perceive touch (numbness). |
| Hypotonia (HP:0001252) | Hypotonia is an abnormally low muscle tone (the amount of tension or resistance to movement in a muscle). Even when relaxed, muscles have a continuous and passive partial contraction which provides some resistance to passive stretching. Hypotonia thus manifests as diminished resistance to passive stretching. Hypotonia is not the same as muscle weakness, although the two conditions can co-exist. | Central hypotonia; Peripheral hypotonia; Low or weak muscle tone; Low muscle tone; Muscle hypotonia; Muscular hypotonia | Hypotonia | An abnormally low amount of tension (resistance to being moved) in muscles. |
| Muscle spasm (HP:0003394) | Sudden and involuntary contractions of one or more muscles. | Muscle cramps | Muscle cramp | Sudden and involuntary contractions of one or more muscles. |
| Muscle weakness (HP:0001324) | Reduced strength of muscles. | Muscular weakness | Muscle weakness | Reduced strength of muscles. |
| Orthostatic hypotension (HP:0001278) | A form of hypotension characterized by a sudden fall in blood pressure | Decrease in blood pressure upon standing up; Postural hypotension | Orthostatic hypotension | Decrease in blood pressure upon standing up. |

| | | | | |
|---|---|---|---|---|
| | that occurs when a person assumes a standing position. | | | |
| Paresthesia (HP:0003401) | Abnormal sensations such as tingling, pricking, or numbness of the skin with no apparent physical cause. | Pins and needles feeling; Tingling; Paresthesias | Tingling | Abnormal sensations such as tingling, pricking, or numbness of the skin with no apparent physical cause. |
| Parkinsonism (HP:0001300) | Characteristic neurologic anomaly resulting from degeneration of dopamine-generating cells in the substantia nigra, a region of the midbrain, characterized clinically by shaking, rigidity, slowness of movement and difficulty with walking and gait. | Parkinsonian disease | Parkinsonism | Manifestations that resemble symptoms of Parkinson disease such as shaking, rigidity, slowness of movement and difficulty with walking and gait. |
| Polyneuropathy (HP:0001271) | A generalized disorder of peripheral nerves. | Peripheral nerve disease | Peripheral nerve disease | Peripheral nerve disease |
| Rigidity (HP:0002063) | Continuous involuntary sustained muscle contraction. When an affected muscle is passively stretched, the degree of resistance remains constant regardless of the rate at which the muscle is stretched. This feature helps to | Muscle rigidity | Rigidity | The inability of the muscles to relax normally due to involuntary and sustained contraction. |

| | | | | |
|---|---|---|---|---|
| | distinguish rigidity from muscle spasticity. | | | |
| Seizure (HP:0001250) | A seizure is an intermittent abnormality of nervous system physiology characterised by a transient occurrence of signs and/or symptoms due to abnormal excessive or synchronous neuronal activity in the brain. | Seizures; Epilepsy; Epileptic seizure | Seizure | Convulsion (uncontrollable jerking movements of the arms and legs) or change in behavior often accompanied by altered consciousness. Individuals who have two or more seizures without an identifiable cause are generally considered to have epilepsy. |
| Skeletal muscle atrophy (HP:0003202) | The presence of skeletal muscular atrophy (which is also known as amyotrophy). | Neurogenic muscle atrophy, especially in the lower limbs; Muscle wasting; Amyotrophy involving the extremities; Muscle hypotrophy; Neurogenic muscle atrophy; Muscle degeneration; Amyotrophy; Muscle atrophy; Neurogenic muscular atrophy; Muscular atrophy; Muscle atrophy, neurogenic | Skeletal muscle atrophy | Wasting (atrophy) of muscles. |
| Somatic sensory dysfunction (HP:0003474) | An abnormality of the primary sensation that is mediated by peripheral nerves | Sensory impairment | General sensory impairment | An impairment of sensation with reduced ability to sense pain, temperature, |

| | | | | |
|---|---|---|---|---|
| | (pain, temperature, touch, vibration, joint position). The word hypoesthesia (or hypesthesia) refers to a reduction in cutaneous sensation to a specific type of testing. | | | touch, vibration, and/or joint position. |
| Spasticity (HP:0001257) | A motor disorder characterized by a velocity-dependent increase in tonic stretch reflexes with increased muscle tone, exaggerated (hyperexcitable) tendon reflexes. | Muscular spasticity; Involuntary muscle stiffness, contraction, or spasm; Muscle spasticity | Spasticity | A condition in which muscles remain contract and do not relax normally, which prevents normal, smooth movement. |
| Tremor (HP:0001337) | An unintentional, oscillating to-and-fro muscle movement about a joint axis. | Tremors | Tremor | Uncontrollable shaking movements of a body part. |
| Unilateral facial palsy (HP:0012799) | One-sided weakness of the muscles of facial expression and eye closure. | Unilateral facial muscle weakness; Unilateral facial weakness; Paralysis of one side of the face; Unilateral facial paralysis; Weakness of one side of the face; Facial droop; Unilateral facial muscle paralysis | One-sided facial palsy | One-sided weakness of the muscles of the face. |

neuropsychiatric-sleep

| HPO Term | Definition | Synonyms | Plain-language label | Plain-language definition |
|---|---|---|---|---|
| Insomnia (HP:0100785) | Persistent difficulty initiating or maintaining sleep. | Fragmented sleep; Difficulty staying or falling asleep | Insomnia | Difficulty falling and/or staying asleep. |
| Maintenance insomnia (HP:0031355) | Abnormal difficulty in staying asleep. Affected individuals tend to wake up at night and have difficulty returning to sleep. | Waking up several times during the night | Maintenance insomnia | Difficulty sleeping through the night. |
| Restless legs (HP:0012452) | A feeling of uneasiness and restlessness in the legs after going to bed (sometimes causing insomnia). | Willis-Ekbom disease; Restless legs syndrome; Wittmaack-Ekbom syndrome | Restless legs | A feeling of uneasiness and restlessness in the legs after going to bed (sometimes causing insomnia). |
| Sleep apnea (HP:0010535) | An intermittent cessation of airflow at the mouth and nose during sleep. Apneas of at least 10 seconds are considered important, but persons with sleep apnea may have apneas of 20 seconds to up to 2 or 3 minutes. Patients may have up to 15 events per hour of sleep. | Pauses in breathing while sleeping; Sleep apnoea | Sleep apnea | Pauses in breathing while sleeping. |
| Sleep disturbance (HP:0002360) | An abnormality of sleep including such phenomena as 1) insomnia/hypersomnia, 2) non-restorative sleep, 3) sleep schedule disorder, 4) | Sleep disturbances; Sleep dysfunction; Difficulty sleeping; Trouble sleeping | Sleep disturbance | A general term for sleep problem such as 1) insomnia/hypersomnia, 2) non-restorative sleep, 3) sleep schedule disorder, 4) excessive daytime |

| | | | | |
|---|---|---|---|---|
| | excessive daytime somnolence, 5) sleep apnea, and 6) restlessness. | | | somnolence, 5) sleep apnea, and 6) restlessness. |
| Sleep onset insomnia (HP:0031354) | Difficulty initiating sleep, that is, increased sleep onset latency. | Difficulty falling asleep | Sleep onset insomnia | Difficulty falling asleep. |
| Terminal insomnia (HP:0031356) | A type of insomnia characterized by waking up (too) early in the morning. | Late insomnia | Terminal insomnia | Consistently waking up earlier than desired and unable to fall back asleep. |

gi-symptoms

| HPO Term | Definition | Synonyms | Plain-language label | Plain-language definition |
| --- | --- | --- | --- | --- |
| Abdominal pain (HP:0002027) | An unpleasant sensation characterized by physical discomfort (such as pricking, throbbing, or aching) and perceived to originate in the abdomen. | Gastro pain; Pain in stomach; Stomach pain; Abdominal discomfort; Upset stomach; Gastrointestinal pain | Abdominal pain | Pain in the belly. |
| Abdominal symptom (HP:0011458) | A subjective manifestation of disease localized to the abdomen. | - | Gut symptom | A general term for symptoms that are related to the abdomen (gut, belly). |
| Anorexia (HP:0002039) | A lack or loss of appetite for food (as a medical condition). | - | Anorexia | A lack or loss of appetite for food. |
| Bowel incontinence (HP:0002607) | Involuntary fecal soiling in adults and children who have usually already been toilet trained. | Anal incontinence; Loss of bowel control; Faecal incontinence; Fecal incontinence | Bowel incontinence | Loss of control over bowel movements with involuntary leakage of stool (feces) from the rectum. |
| Constipation (HP:0002019) | Infrequent or difficult evacuation of feces. | Dyschezia; Costiveness | Constipation | Infrequent and difficult bowel movements. |
| Diarrhea (HP:0002014) | Abnormally increased frequency of loose or watery bowel movements. | Watery stool; Diarrhoea | Diarrhea | Increased frequency of loose or watery bowel movements. |
| Early satiety (HP:0033842) | The condition of being unable to eat a full meal because of a feeling of fullness (satiety), or or feeling very full | Feeling full quickly when eating; Not able to finish a normal-sized meal | Feeling full quickly when eating | Feeling full quickly when eating, not being able to eat a normal-sized meal. |

| | | | | |
|---|---|---|---|---|
| | after eating only a small amount of food. | | | |
| Nausea (HP:0002018) | A sensation of unease in the stomach together with an urge to vomit. | - | Nausea | An urge to vomit. |
| Vomiting (HP:0002013) | Forceful ejection of the contents of the stomach through the mouth by means of a series of involuntary spasmic contractions. | Throwing up; Emesis | Vomiting | Throwing up. |
| Poor appetite (HP:0004396) | A reduced desire to eat. | No appetite; Decreased appetite; Loss of appetite | Poor appetite | A reduced desire to eat. |

HEENT-eye

| HPO Term | Definition | Synonyms | Plain-language label | Plain-language definition |
|---|---|---|---|---|
| Blindness (HP:0000618) | Blindness is the condition of lacking visual perception defined as visual perception below 3/60 and/or a visual field of no greater than 10 degrees in radius around central fixation. | Legal blindness; Total vision loss | Blindness | The complete loss of vision. |
| Blurred vision (HP:0000622) | Lack of sharpness of vision resulting in the inability to see fine detail. | - | Blurred vision | Vision that is blurry or lacks clarity/sharpness. |
| Conjunctivitis (HP:0000509) | Inflammation of the conjunctiva. | Pink eye; Conjunctivitis, recurrent | Conjunctivitis | Inflammation or infection of the outer lining of the eyeball resulting in “pink eye” appearance. |
| Diplopia (HP:0000651) | Diplopia is a condition in which a single object is perceived as two images, it is also known as double vision. | Double vision | Double vision | Double vision. |
| Gaze-evoked nystagmus (HP:0000640) | Nystagmus made apparent by looking to the right or to the left. | - | Nystagmus triggered by gaze | Repetitive, involuntary, to-and-fro oscillation of the eyes that is triggered by looking in a certain direction. |
| Keratoconjunctivitis sicca (HP:0001097) | Dryness of the eye related to deficiency of the tear film components (aqueous, mucin, or lipid), lid surface | Keratitis sicca; Dry eye syndrome; Xerophthalmia; Dry eyes | Keratoconjunctivitis sicca (a form of dry eyes) | Dryness of the eye due to inadequate tears, which may cause feelings of irritation, itching, burning, or the sensation of a foreign body in |

| | | | | |
|---|---|---|---|---|
| | abnormalities, or epithelial abnormalities. Keratoconjunctivitis sicca often results in a scratchy or sandy sensation (foreign body sensation) in the eyes, and may also be associated with itching, inability to produce tears, photosensitivity, redness, pain, and difficulty in moving the eyelids. | | | the eye. |
| Ocular pain (HP:0200026) | An unpleasant sensation characterized by physical discomfort (such as pricking, throbbing, or aching) localized to the eye. | Eye pain | Eye pain | Physical discomfort or pain felt in the eye. |
| Ocular pruritus (HP:0033841) | Pruritus is an itch or a sensation that makes a person want to scratch. This term refers to an abnormally increased sensation of itching in the region of the eye. | Ocular itch; Itchy eyes | Itchy eyes | Itchiness in or around the eyes. |
| Peripheral visual field loss (HP:0007994) | Loss of peripheral vision with retention of central vision, resulting in a constricted circular tunnel-like field of vision. | Loss of peripheral vision; Tunnel vision; Kalnienk vision | Tunnel vision | Loss of peripheral vision with retention of central vision, resulting in a constricted circular tunnel-like field of vision. |

| | | | | |
|---|---|---|---|---|
| Photophobia (HP:0000613) | Excessive sensitivity to light with the sensation of discomfort or pain in the eyes due to exposure to bright light. | Photodysphoria; Light hypersensitivity; Extreme sensitivity of the eyes to light | Photophobia | Excessive sensitivity to light. |
| Red eye (HP:0025337) | A reddish appearance over the white part (sclera) of the eye ranging from a few enlarged blood vessels appearing as wiggly lines over the sclera to a bright red color completely covering to sclera. | Red eyes | Red eye | Redness of the eye related to inflammation. |
| Visual loss (HP:0000572) | Loss of visual acuity (implying that vision was better at a certain time point in life). Otherwise the term reduced visual acuity should be used (or a subclass of that). | Vision loss; Loss of vision | Vision loss | Vision loss. |
| Vitreous floaters (HP:0100832) | Deposits of various size, shape, consistency, refractive index, and motility within the eye's vitreous humour, which is normally transparent. | Eye floaters; Flitting flies; Spots in front of eyes; Mouches volantes; Vitreous condensations; Myodesopsia; Vitreous opacities; Vitreous debris; Vitreous veils; Myodeopsia | Eye floaters | Eye floaters are spots that appear to float in the field of vision and typically drift about rather than staying in the same place. |
